# Immunogenic Amino Acid Motifs and Linear Epitopes of COVID-19 mRNA Vaccines

**DOI:** 10.1101/2021.05.25.21257427

**Authors:** Adam V Wisnewski, Carrie A Redlich, Kathy Kamath, Queenie-Ann Abad, Richard F Smith, Louis Fazen, Romero Santiago, Jian Liu, Julian Campillo Luna, Brian Martinez, Elizabeth Baum-Jones, Rebecca Waitz, Winston A Haynes, John C Shon

## Abstract

Reverse vaccinology is an evolving approach for improving vaccine effectiveness and minimizing adverse responses by limiting immunizations to critical epitopes. Towards this goal, we sought to identify immunogenic amino acid motifs and linear epitopes of the SARS-CoV-2 spike protein that elicit IgG in COVID-19 mRNA vaccine recipients. Paired pre/post vaccination samples from N=20 healthy adults, and post-vaccine samples from an additional N=13 individuals were used to immunoprecipitate IgG targets expressed by a bacterial display random peptide library, and preferentially recognized peptides were mapped to the spike primary sequence. The data identify several distinct amino acid motifs recognized by vaccine-induced IgG, a subset of those targeted by IgG from natural infection, which may mimic 3-dimensional conformation (mimotopes). Dominant linear epitopes were identified in the C-terminal domains of the S1 and S2 subunits (aa 558-569, 627-638, and 1148-1159) which have been previously associated with SARS-CoV-2 neutralization in vitro and demonstrate identity to bat coronavirus and SARS-CoV, but limited homology to non-pathogenic human coronavirus. The identified COVID-19 mRNA vaccine epitopes should be considered in the context of variants, immune escape and vaccine and therapy design moving forward.

## INTRODUCTION

The humoral immune response to vaccination is thought to be an important component of protective immunity (1, 2). The first mRNA vaccines for COVID-19 have been shown to elicit antibodies with significant neutralizing capacity in vitro and protect against severe disease in vivo (3-6). The SARS-CoV-2 spike protein epitopes that induce specific IgG in COVID-19 mRNA vaccine recipients remain incompletely characterized and could form the basis of future “epitope vaccines” for SARS-CoV-2 (7-9).

Despite limited knowledge of immunogenic SARS-CoV-2 spike epitopes in vaccinated individuals, immunodominant epitopes in naturally infected COVID-19 patients have been identified based on recognition by serum IgG (10-18). The receptor binding domain (RBD) is an important region that contains conformational epitopes encoded by non-contiguous regions of the spike protein (19). Dominant linear epitopes are located in the CTDs of S1 and S2, the S1/S2 cleavage site, and the fusion peptide region of S2 (10-16). Importantly, dominant linear epitopes and specific conformational epitopes have been shown to mediate viral neutralization in vitro (10, 13, 16, 20).

The present study mapped COVID-19 mRNA vaccine epitopes using Serum Epitope Repertoire Analysis (SERA), a technique based on a high throughput random bacterial peptide display technology (12). SERA’s unbiased, whole proteome approach to epitope discovery is ideal for mapping the targets of humoral responses to vaccination. The primary focus herein is on de novo responses of naïve individuals, whose immune system’s specificity was not influenced by prior SARS-CoV-2 exposure. Pre vs. post vaccine serum samples were available for most participants to help ensure immune specificity was a result of vaccination, not pre-existing cross-reactivity. Similarities and differences in epitopes induced by mRNA vaccine vs. natural COVID-19, and their association with viral neutralization are discussed, along with preliminary findings from vaccinated individuals that previously had COVID-19.

## RESULTS AND DISCUSSION

### Identification of amino acid motifs recognized by vaccine-induced serum IgG

Amino acid motifs preferentially recognized by serum IgG from individuals after COVID-19 mRNA vaccination were readily identified by SERA analysis as previously described (12). The relative increase in amino acid motif-specific IgG vs. pre-pandemic controls is shown in Figure 1 for (A) N=20 subjects without prior COVID-19 before Pfizer-BioNTech immunization, (B) N=20 subjects without prior COVID-19 after immunization with the Pfizer-BioNTech COVID-19 vaccine, (C) N=8 subjects without prior COVID-19 after immunization with Moderna vaccine, and (D) 5 subjects with prior COVID-19 immunized with either Pfizer-BioNTech (N=2) or Moderna (N=3) mRNA vaccines. Additional information on the study subjects and amino acid motifs are provided in supplemental materials (Tables S1 and S2).

**Figure 1.**
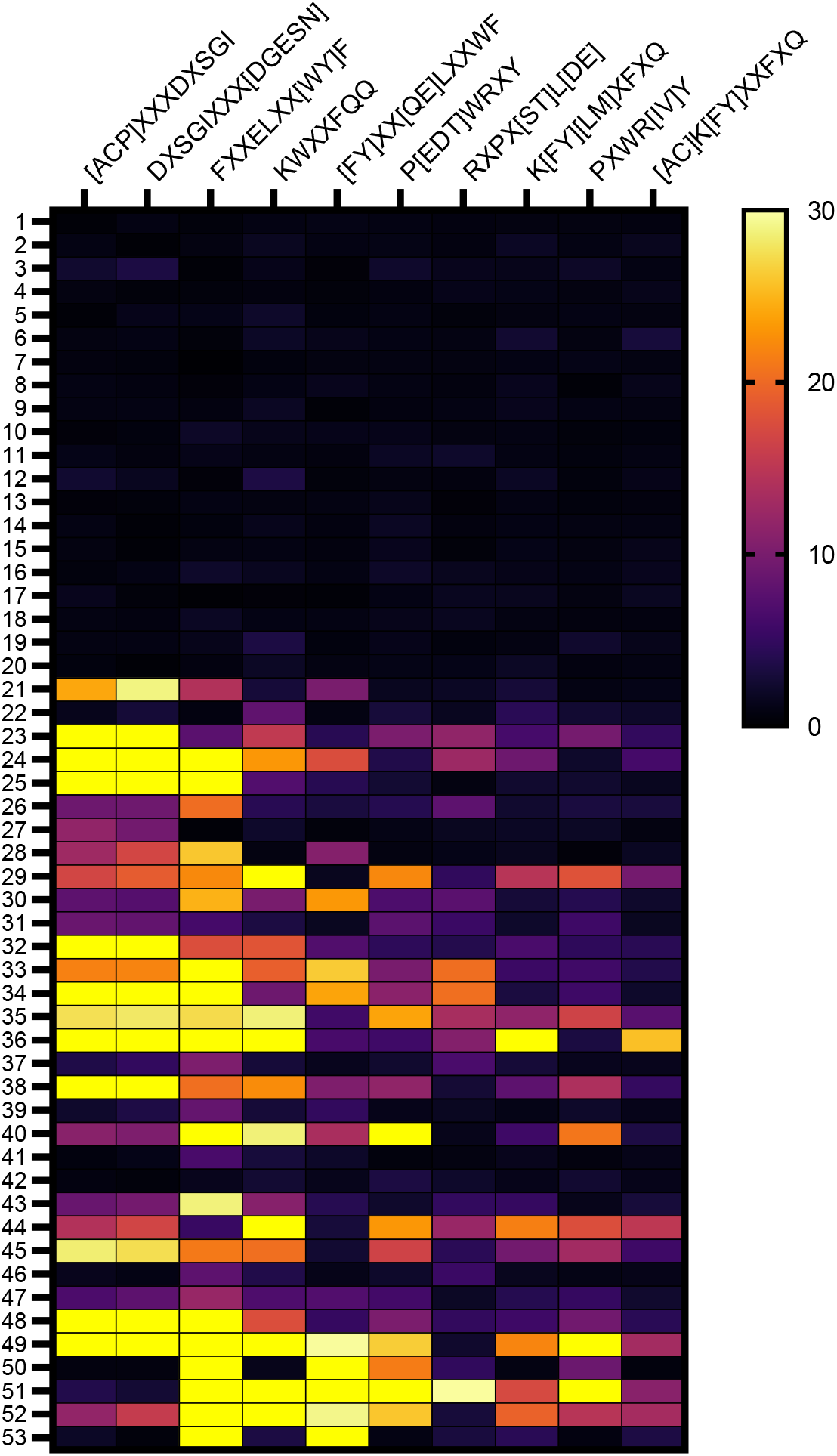
Heat map identifying amino acid motifs preferentially recognized by IgG from vaccinated subjects. Amino acid motifs were identified among the peptides immunoprecipitated by IgG from vaccine recipients using IMUNE algorithm and the level of enrichment (fold increase) in serum of individual subjects relative to pre-pandemic control subjects is depicted in the heat map. Samples 1-20 are pre-vaccine and samples 21-40 are the same subjects after Pfizer-BioNTech COVID-19 mRNA vaccination. Subjects 41-48 are from subjects that received Moderna vaccine. Samples 49-53 are vaccinated subjects that previously had COVID-19.

Some preferentially recognized motifs share overlapping sequence and their IgG recognition show excellent correlation (*r*_s_ >0.9) with each other (see supplemental materials Figure S1). Preferentially recognized motifs without overlapping amino acids also showed significant (*p* < 0.001), but more limited correlation with one another; the most substantial between K[FY][LM]XFXQ and P[EDT]WRXY had an *r*_s_ of 0.66 (supplemental materials Figure S2). There was significant (*p* < 0.044) but weak to moderate (*r*_s_ = 0.35-0.46) correlation of ELISA OD against spike antigen with several amino acid motifs, but no significant correlation with gender (see Supplemental Material Correlation Matrix 1). A moderate inverse relationship between age and recognition of several amino acid motifs were observed, the most significant (*p* < 0.012) for overlapping amino acid motifs KWXXFQQ and K[FY][LM]XFXQ (*r*_s_ = -0.44 see supplemental materials Figure S2. The preferentially recognized amino acid motifs do not map to the primary sequence of SARS-CoV-2 proteome but may mimic 3-dimensional epitopes (mimotopes) formed by non-continuous regions of the spike antigen and are a subset of those preferentially recognized by IgG from hospitalized COVID-19 patients as previously reported (12, 21).

### Identification of linear SARS-CoV-2 spike epitopes

Linear epitopes of COVID-19 mRNA vaccines were identified by analyzing the specificity of IgG from available pre/post vaccine serum samples, using a protein-based immunome wide association study (PIWAS) approach as previously described (12, 22). The linear epitope (LE)-1 with the highest PIWAS values was also the most commonly recognized by IgG from >2/3 of the vaccinated subjects (Figure 2). LE-1 (aa 558-569) is located in the C-terminal domain (CTD) of the spike S1 subunit and contains the core amino acids present in dominant linear epitopes of COVID-19 patients (S14P5, S1-93, S1-55, S556-570), independently described by several other research teams, shown in Figure 3 (10, 11, 14, 16, 18). Previously described epitopes containing the LE-1 core sequence FLPFQQ mediate neutralization in vitro based on studies with epitope-affinity purified/depleted COVID-19 patient serum, and through competitive inhibition assays with clinical isolates (10, 16). We have previously reported a correlation of LE-1 specific IgG in COVID-19 patients with serum neutralizing titers in vitro (13). In the present vaccinated population, we observed a significant (*p* < 0.0004) association of LE-1 recognition with total spike IgG (ELISA OD) and several different amino acid motifs, with strongest *r*_s_ values (>0.6) against overlapping motifs FXXELXX[WY]F and [FY]XX[QE]LXXWF, which contain portions of the FLPFQQ core of LE-1 (see supplemental materials Figure S3).

**Figure 2.**
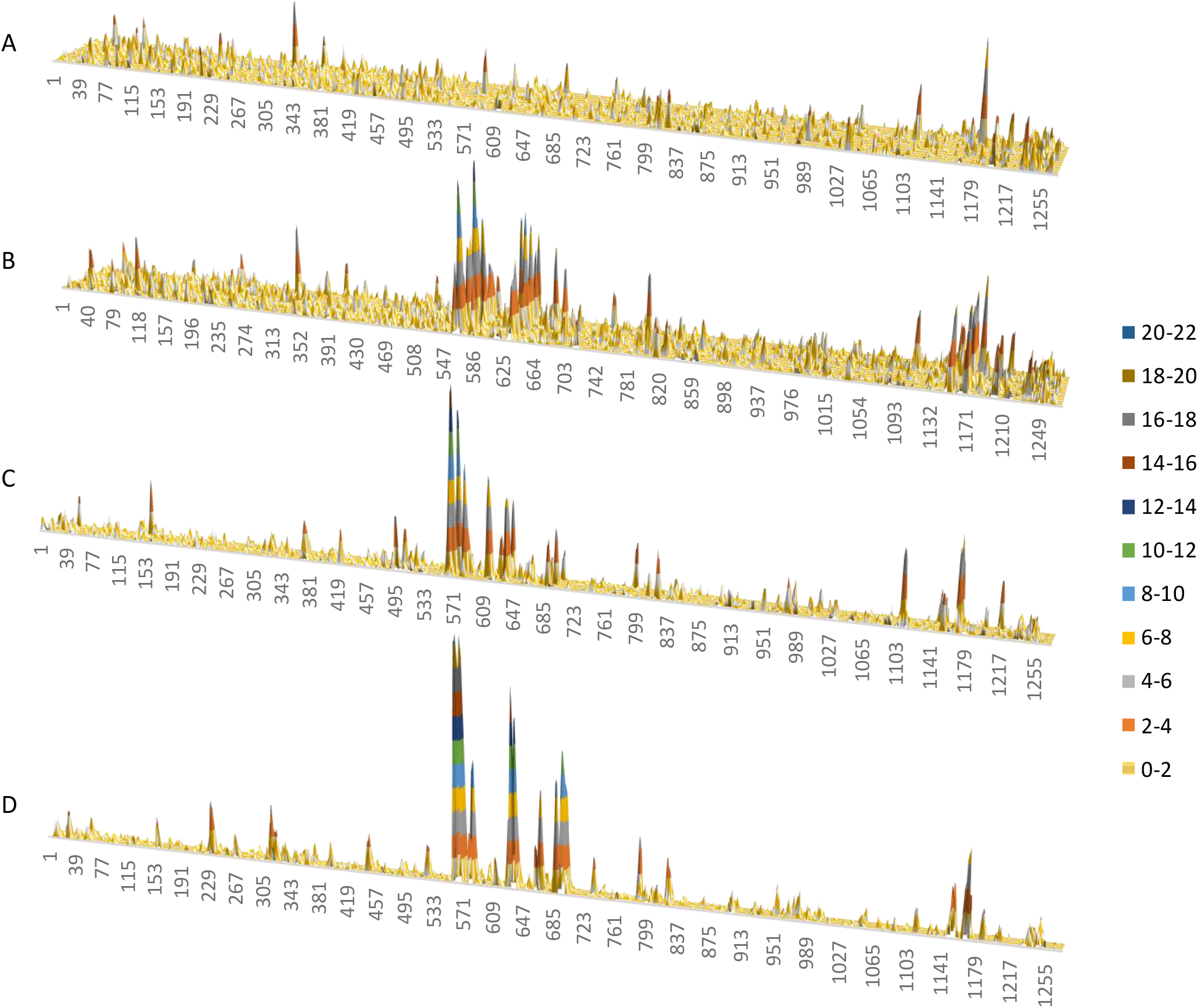
Linear epitope mapping of spike protein in mRNA vaccine recipients. The relative specificty for linear epitopes or PIWAS score (Y-axes, color coded key to right) are graphed across the spike primary sequence (amino acid numbers on X-axis) for paired samples from subjects# 1-20 in panel A (pre-vaccine) and panel B (post-vaccine) that received Pfizer-BioNTech vaccine. Panel C shows data from subjects # 21-28, post Moderna vaccine and panel D shows vaccinated subjects# 29-33 that previously had COVID-19

**Figure 3.**
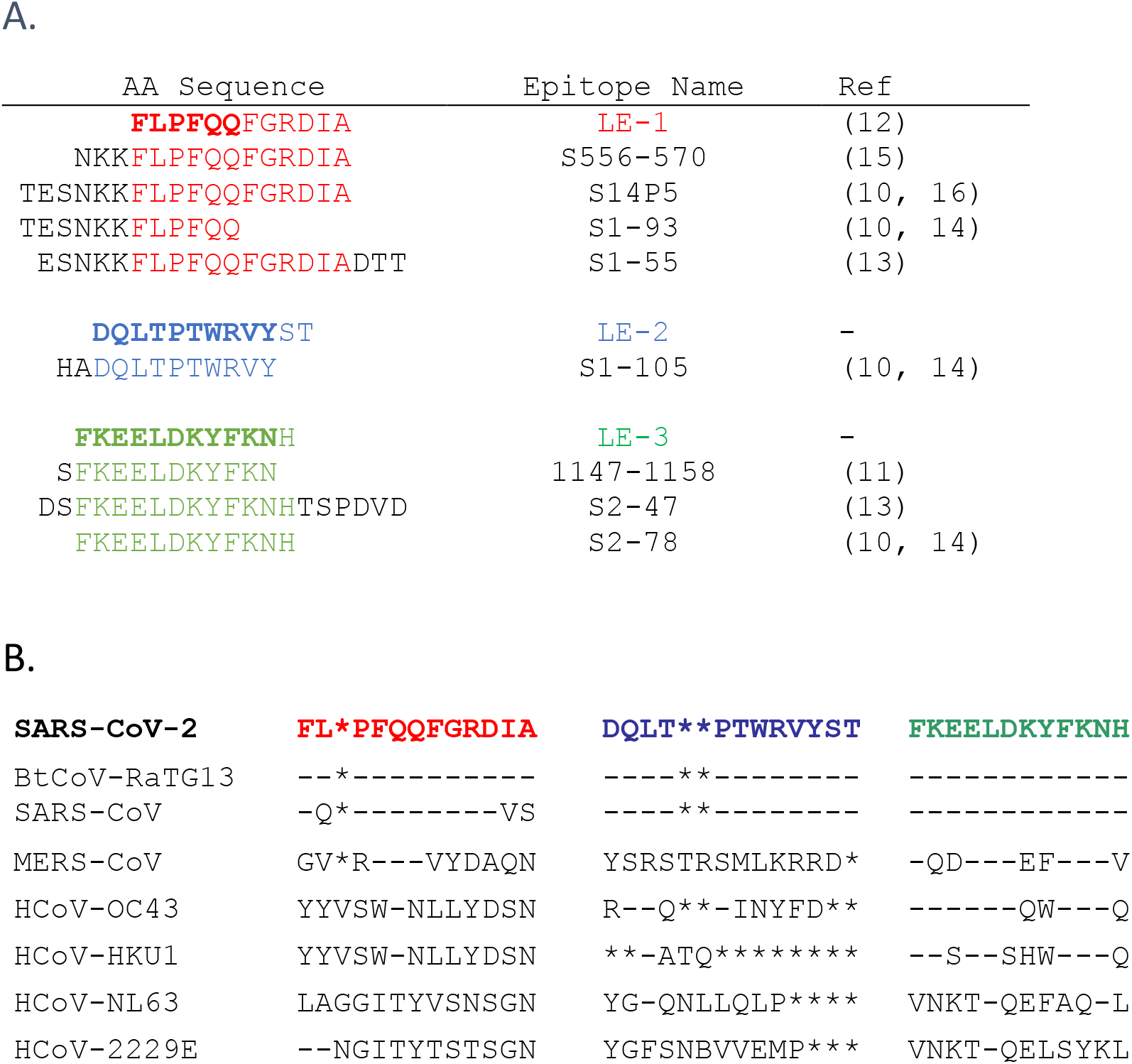
Alignment of dominant linear vaccine epitopes with those induced by infection and in other coronaviruses. Alignment of vaccine epitopes with those previously described in natural SARS-CoV-2 infection (A). Alignment of vaccine epitopes with homologous regions in other coronaviruses (B). * are spaces included to maximize alignment.

The linear epitope (LE-2) with the second highest average PIWAS score was recognized by IgG from 2/3 of the vaccinated subjects (Fig. 2). LE-2 (aa 627-638) is also located in the CTD of S1 and largely overlaps (10/12 amino acids) epitope S1-105, previously defined by Yi et al. (see Figure 3), based on recognition by serum from naturally infected subjects (13). S1-105 mediates a significant portion of COVID-19 patients’ serum neutralizing capacity against a SARS-CoV-2 clinical isolate in limited testing (13). Recognition of LE-2 (PIWAS score) was significantly correlated with that of each of the 10 amino acid motifs defined in Figure 1 and previously published (12), but not age, gender or total spike IgG.

A third linear epitope (LE-3) in the S2 CTD, near the 2^nd^ heptad repeat region (HD2), is shared by a majority of vaccinated individuals. LE-3 (aa 1148-1159) contains the core amino acids present in dominant linear epitopes S2-78, S2-47, and 1147-1158 independently described by three other research teams in COVID-19 patients and shown in Figure 3 (10, 11, 13, 14). These epitopes mediate neutralization in vitro, have been experimentally observed in SARS-CoV (20), and predicted bioinformatically for SARS-CoV-2 (23-25). Recognition of LE-3 (PIWAS) was significantly (*p* < 0.0003) associated with that of overlapping motifs P[EDT]WRXY and PXWR[IV]Y (r > 0.58), but not age, sex or total IgG. An adjacent linear epitope, LE-4, spanning amino acids 1164-1173 is also recognized by IgG from more than ½ of the vaccinated subjects (Figures 2 and 4).

**Figure 4.**
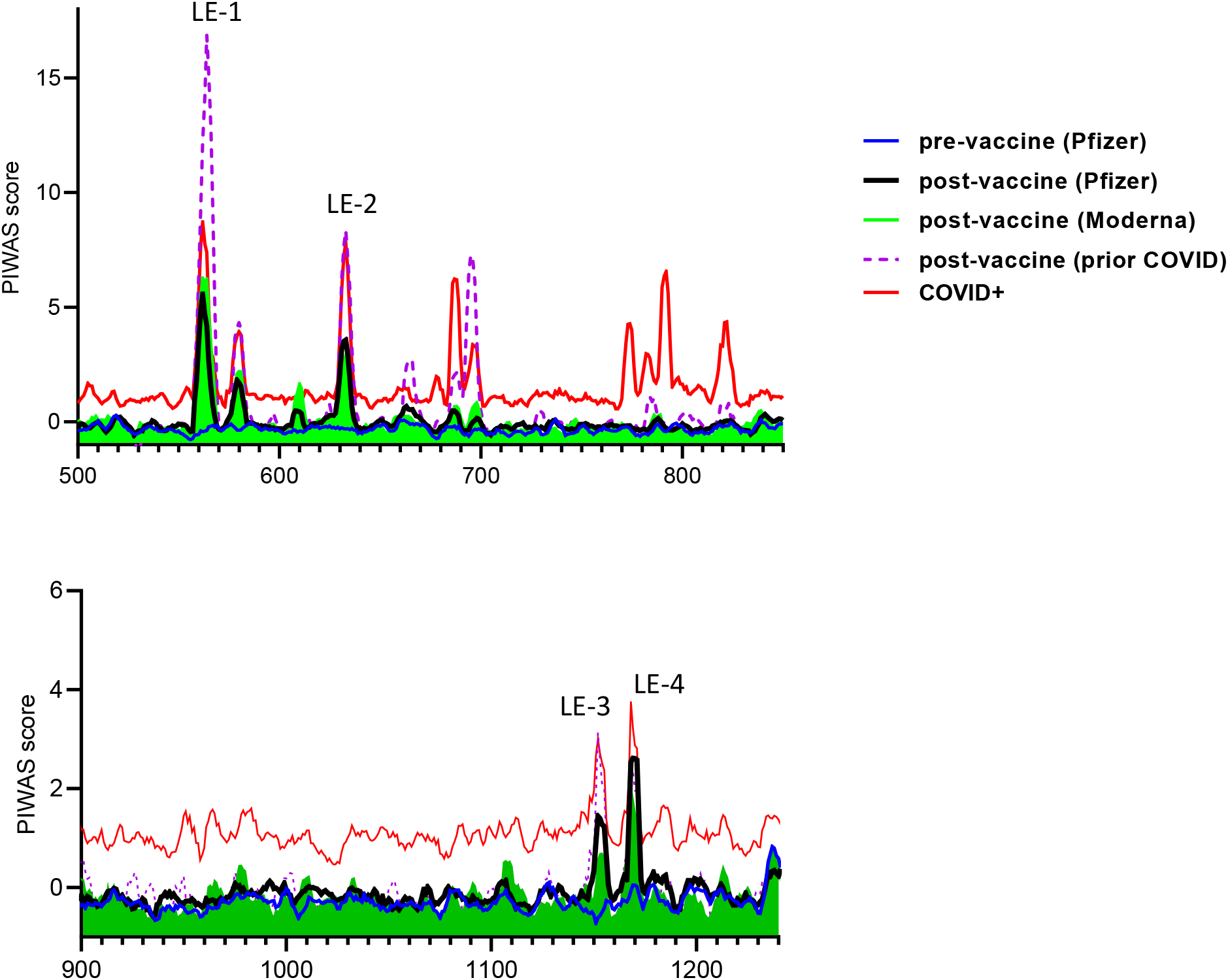
Dominant linear epitopes in COVID-19 mRNA vaccine recipients. The relative IgG binding (average PIWAS, Y-axes) to different linear epitopes (spike amino acid number, X-axis) is highlighted for the C-terminal domains of S1 (A) and S2 (B). Data are from N=20 Pfizer-BioNTech, N=8 Moderna vaccine recipients, and N=5 prior COVID-19 patients that received either Pfizer or Moderna vaccine as labeled in the key in upper right. The red line corresponds to the 95% quantile PIWAS score for a prior cohort of COVID+ subjects (12). Note the highest PIWAS scores for LE-1 and LE-2 as labeled and limited vaccine induced IgG towards fusion peptide region, aa 788-806 (40).

Additional linear spike epitopes recognized by IgG from multiple post mRNA vaccinated subjects include 12-mers centered around spike amino acids 580, 610, 665, and surrounding the S1/S2cleavage loci ∼ 685-700 (Figure 4). A complete list of the PIWAS scores for all 12-mer linear epitopes of the spike protein for each study subjects is provided as supplementary material (Table S3).

### Uniqueness of linear epitopes

Severe outcomes of SARS-CoV-2 infections are thought in part to arise from the novelty of the virus and the naïve immune status of most humans. The dominant vaccine linear epitopes LE-1, LE-2, and LE-3 show identity or near identity to bat coronavirus and SARS-CoV, but not non-pathogenic human coronaviruses (Figure 3). Further homology searches of the linear spike epitopes identify limited sequence identity outside of organisms to which humans are rarely exposed, such as thermophilic bacteria (supplemental materials List S1). The present data support the hypothesis that humoral immune naivete contributes to SARS-CoV-2’s pathogenicity and identifies specific deficiencies in the normal adult immune repertoire readily corrected by mRNA vaccination.

### IgG responses in mRNA vaccine recipients vs. natural infection

We compared the linear epitope recognition by IgG from the different groups of vaccinated subjects in the present study and our prior studies of COVID-19 patients (12). Similar specificity for dominant linear epitopes were observed in individuals that received mRNA vaccine from Pfizer-BioNTech or Moderna (Figure 4), regardless of prior COVID-19 status. However, vaccinated individuals with prior COVID-19 exhibited significantly (*p <* 0.001) higher PIWAS values for dominant epitopes LE-1, LE-2, and LE-3 than those without prior infection. Vaccine-induced IgG specificity overlaps substantially with that of COVID-19 patients (12) and can occur de novo, or augment naturally occurring responses (see pre/post vaccine examples in supplemental material Figure S4). The most notable difference in specificity between vaccine vs. infection induced IgG, is the lack of specificity for linear epitopes in the fusion peptide region of the spike-S2 domain by vaccine-elicited IgG. Despite the induction of specific IgG to the S2 fusion peptide domain in natural SARS-CoV-2 infection, vaccination does little to boost this response compared to the dominant linear epitopes (Figure 4, and supplemental materials Figure S4).

### RBD epitopes

This study did not identify vaccine epitopes of the spike RBD domain, which is a major target for neutralizing antibodies in natural infection. The findings are consistent with prior reports that did not detect spike RBD linear epitopes and may be due to multiple reasons including limitations in sensitivity and frequency/sample size, as well as the RBD’s complex secondary structure, which is thought to form immunogenic 3-D conformations, or non-continuous epitopes (11, 13, 17, 26). RBD specific antibodies are a minor proportion of total anti-spike IgG and linear RBD epitopes are not prominent IgG epitopes in natural SARS-CoV-2 infection (12, 13, 27). Nonetheless, the RBD is an important target of neutralizing antibodies that develop in vivo during natural infection (28), and possibly a driver for the evolution of mutations given its critical role in the virus life cycle (29, 30). Many of the SARS-CoV-2 variants that have evolved to date contain mutations that affect the RBD and result in decreased neutralization by RBD-specific antibodies (31), but not mAbs that bind outside the receptor-binding motif (32, 33). In this context dominant linear neutralizing epitopes may provide an increased role in immunity and compensate for loss of neutralization activity due to decreased IgG recognition of variant RBDs, and perhaps contribute to current mRNA vaccine efficacy in populations with high prevalence of B.1.1.7 and B.1.351 variants (34). Mutations that result in escape from dominant linear epitopes outside the RBD might also adversely affect host immunity and vaccine efficacy, particularly if compounded by RBD escape variants. It is thus important to be vigilant in surveillance for immune escape from dominant linear epitopes as well as conformational RBD affected epitopes in new variants.

### Summary

Spike protein epitopes of COVID-19 mRNA vaccines were identified through immunomics / reverse vaccinology. Dominant epitopes included specific amino acid motifs that may reflect 3-D conformations and linear epitopes of the spike protein, particularly in the C-terminal domain of S1. COVID-19 mRNA vaccine epitopes share identity with bat coronavirus, SARS-CoV (but not the spike protein of non-pathogenic human coronaviruses), and epitopes that trigger IgG during natural SARS-CoV-2 infection, which have been previously associated with SARS-CoV-2 neutralization (10, 13, 16, 20). The finding of increased IgG response in vaccinated individuals with prior infection (vs. naïve individuals) is consistent with previous reports and highlights the effectiveness of “booster shots” towards robust humoral immunity (35). In summary, we identified epitopes of COVID-19 mRNA vaccines that trigger specific IgG responses previously associated with SARS-CoV-2 neutralizing activity and which may form the basis of future diagnostics, therapeutics, and focused vaccine development (e.g., multivalent peptide).

## MATERIALS AND METHODS

### Blood samples from vaccine recipients

Subjects were working aged adults employed in the health care industry. Blood was obtained by venipuncture in a serum separator tube (BD Vacutainer^®^, Franklin Lakes, NJ) and the serum fraction was separated following centrifugation and stored at -80°C. Four groups of samples were studied; (A) N=20 subjects without prior COVID-19 before immunization, (B) N=20 subjects without prior COVID-19 after immunization with the Pfizer-BioNTech COVID-19 vaccine, (C) N=8 subjects without prior COVID-19 after immunization with Moderna vaccine, and (D) 5 subjects with prior COVID-19 immunized with either Pfizer-BioNTech (N=2) or Moderna (N=3). The investigation was approved by the Yale University Institutional Review Board (IRB) and all study subjects provided written informed consent before participating in the study.

### ELISAs

ELISAs were performed as previously described (36, 37). Triton X-100 and RNase A were added to serum samples at final concentrations of 0.5% and 0.5mg/mL respectively and incubated at room temperature (RT) for 30 minutes before use to reduce risk from any potential virus in serum. 96-well MaxiSorp plates (ThermoFisher Scientific, Waltham, MA) were coated with 50 μL/well of recombinant SARS Cov-2 spike ectodomain or nucleocapsid protein (Sino Biological, Wayne, PA) at a concentration of 1 μg/mL in NaCO_3_ buffer pH 9.6 and incubated overnight at 4°C. ELISA plate blocked for 1h at RT with 200 μL of PBS with 3% milk powder. Serum was diluted 1:100 in PBS with 0.05% Tween20, 1% milk powder and 100 μL was added for 1 hr at RT. Plates were washed three times with PBS-T (PBS with 0.1% Tween-20) and incubated with 50 μL of HRP anti-Human IgG Antibody (Pharmingen, 1:2,000) for 1 h at RT. Plates were developed with 100 μL of TMB Substrate Reagent Set (ThermoFisher) and the reaction was stopped when an internal pooled serum positive control sample reaches an OD of 1.0 at 650 nm, by the addition of 2 N sulfuric acid. Plates were then read at a wavelength of 450 nm with 650 nm reference calibration.

### SERA

A description of the SERA assay has been previously published (9). Briefly, serum was incubated with a fully random 12-mer bacterial display peptide library (1×10^10^ diversity, 10-fold oversampled) at a 1:25 dilution in a 96-well, deep well plate format. Antibody-bound bacterial clones were selected with 50 µL Protein A/G Sera-Mag SpeedBeads (GE Life Sciences, cat#17152104010350) (IgG) final assay dilution 1:100 (ThermoFisher). The selected bacterial pools were resuspended in growth media and incubated at 37°C shaking overnight at 300 RPM to propagate the bacteria. Plasmid purification, PCR amplification of peptide encoding DNA, barcoding with well-specific indices was performed as described. Samples were normalized to a final concentration of 4nM for each pool and run on the Illumina NextSeq500. After SERA screening, we applied two complementary discovery tools, IMUNE and PIWAS, to identify antigens and epitopes involved in the SARS-CoV-2 immune response.

### PIWAS analysis

We applied the previously published PIWAS method (22) to identify antigen and epitope signals against the Uniprot reference SARS-CoV-2 proteome (UP000464024) (38). The PIWAS analysis was run on the IgG SERA data with a single sample per COVID-19 patient versus 497 discovery pre-pandemic controls, and 1500 validation controls used for normalization. Additional parameters include: a smoothing window size of 5 5mers and 5 6mers; z-score normalization of kmer enrichments; maximum peak value; and generation of epitope level tiling data.

### IMUNE-based motif discovery

Peptide motifs representing epitopes or mimotopes of SARS CoV-2 specific antibodies were discovered using the IMUNE algorithm as previously described (39). Peptide patterns identified by IMUNE were clustered using a PAM30 matrix and combined into motifs. A motif was classified positive in a given sample if the enrichment was ≥4 times the standard deviation above the mean of a training control set. Thirty eight motifs were enriched in 406 unique confirmed COVID-19 cases from four separate cohorts of COVID patients compared to 1500 pre-pandemic controls.

### Statistical analysis

GraphPad Prism (v8) and Microsoft Excel for Windows 10 (v16.0.13001.20254) were used for statistical analyses. Statistical differences were analyzed with Mann-Whitney U-test. Nonparametric Kruskal-Wallis test was used to perform multiple comparisons between groups analyzed. Correction for multiple comparison was performed with Dunn’s test. Logistic regression or nonparametric Spearman Spearman’s rank correlation were used to compute associations between pairs of data.

## Supporting information

are provided in supplemental materials

## Data Availability

All data are available in the manuscript and supplemental materials

## AUTHOR CONTRIBUTIONS

AW, CR, QA, RFS, RS, LF, JL, JCL obtained samples and performed ELISAs, WAH, KK, JCS performed bacteriophage peptide immunoprecipitation, SERA analysis and bioinformatics. AW wrote and all authors edited, reviewed and approved the final version of the manuscript.

## ACKNOWLEDGEMENTS

We are grateful to the study participants, without which the research could not have been done. The work was supported by funds from the National Institute of Occupational Safety and Health, 5T03OH008607-15.

## COMPETING INTERESTS

All authors have completed the ICMJE uniform disclosure form at www.icmje.org/coi_disclosure.pdf and declare support from the National Institute of Occupational Safety and Health (5T03OH008607-15); no financial relationships with any organizations that might have an interest in the submitted work in the previous three years; and no other relationships or activities that could appear to have influenced the submitted work.

## SUPPLEMENTAL MATERIALS

Supplemental material with available data on correlation between IgG specificities, PIWAS values, and examples of post-COVID/Post-COVID-vaccine samples.

## Notes

### Competing Interest Statement

The authors have declared no competing interest.

### Author Declarations

The study was reviewed and ethical approval was granted by the Yale University Institutional Review Board (IRB).

