## Supplementary material for "Immunogenic Amino Acid Motifs and Linear Epitopes of COVID-19 mRNA Vaccines": are provided in supplemental materials

| Table of Contents | Page |
|---|---|
| Table S1. Table of study subjects | 2 |
| Table S2. Table of fold enrichment in amino acid motifs | 4 |
| Figure S1. Correlation of enrichment for IgG with specificity for overlapping amino acid motifs | 5 |
| Figure S2. Correlation of enrichment for IgG with specificity for non-overlapping amino acid motifs and age | 6 |
| Correlation Matrix 1. Heat map of correlation between recognition of different amino acid motifs | 7 |
| Figure S3. Correlation of PIWAS score for LE-1 vs. enrichment for amino acid motif [FY]XX[QE]LXXWF | 8 |
| Figure S4. Linear epitope mapping of individual subject's IgG, after COVID/pre-vaccine vs. after COVID/post vaccine | 9 |
| List S1. Peptides with strongest identity to SARS-CoV-2 spike mRNA vaccine epitopes identified though BLAST® analysis | 10 |
| Table S3. PIWAS values for all subjects. | 11 |

Table S1. Table of Study Subjects

| Subject # | Sex | Age Range | COVID Vaccination | Vaccine Manufacturer | COVID Vaccination Initial | COVID Vaccination Booster | Timepoint | SARS-CoV-2 PCR Status | SARS-CoV-2 IgG Status | Spike IgG ELISA OD |
|---|---|---|---|---|---|---|---|---|---|---|
| 1 | Male | 35-40 | | | | | pre-vaccine | N/A | Negative | not detected |
| 2 | Female | 30-35 | | | | | pre-vaccine | N/A | Negative | not detected |
| 3 | Female | 46-50 | | | | | pre-vaccine | N/A | Negative | not detected |
| 4 | Male | 25-30 | | | | | pre-vaccine | N/A | Negative | not detected |
| 5 | Female | 46-50 | | | | | pre-vaccine | N/A | Negative | not detected |
| 6 | Female | 40-45 | | | | | pre-vaccine | N/A | Negative | not detected |
| 7 | Female | 56-60 | | | | | pre-vaccine | N/A | Negative | not detected |
| 8 | Male | 35-40 | | | | | pre-vaccine | N/A | Negative | not detected |
| 9 | Female | 35-40 | | | | | pre-vaccine | N/A | Negative | not detected |
| 10 | Female | 40-45 | | | | | pre-vaccine | N/A | Negative | not detected |
| 11 | Male | 30-35 | | | | | pre-vaccine | N/A | Negative | not detected |
| 12 | Female | 26-30 | | | | | pre-vaccine | N/A | Negative | not detected |
| 13 | Female | 26-30 | | | | | pre-vaccine | N/A | Negative | not detected |
| 14 | Female | 30-35 | | | | | pre-vaccine | N/A | Negative | not detected |
| 15 | Female | 26-30 | | | | | pre-vaccine | N/A | Negative | not detected |
| 16 | Female | 30-35 | | | | | pre-vaccine | N/A | Negative | not detected |
| 17 | Female | 26-30 | | | | | pre-vaccine | N/A | Negative | not detected |
| 18 | Male | 26-30 | | | | | pre-vaccine | N/A | Negative | not detected |
| 19 | Male | 56-60 | | | | | pre-vaccine | N/A | Negative | not detected |
| 20 | Female | 26-30 | | | | | pre-vaccine | N/A | Negative | not detected |
| 1 | Male | 35-40 | yes | Pfizer | yes | yes | D36 Post Vaccine | N/A | Positive | 1.6 |
| 2 | Female | 30-35 | yes | Pfizer | yes | yes | D36 Post Vaccine | N/A | Positive | 1.7 |
| 3 | Female | 46-50 | yes | Pfizer | yes | yes | D36 Post Vaccine | N/A | Positive | 2.0 |
| 4 | Male | 25-30 | yes | Pfizer | yes | yes | D36 Post Vaccine | N/A | Positive | 2.5 |
| 5 | Female | 46-50 | yes | Pfizer | yes | yes | D36 Post Vaccine | N/A | Positive | 2.4 |
| 6 | Female | 40-45 | yes | Pfizer | yes | yes | D36 Post Vaccine | N/A | Positive | 1.3 |
| 7 | Female | 56-60 | yes | Pfizer | yes | yes | D36 Post Vaccine | N/A | Positive | 1.4 |
| 8 | Male | 35-40 | yes | Pfizer | yes | yes | D35 Post Vaccine | N/A | Positive | 1.0 |
| 9 | Female | 35-40 | yes | Pfizer | yes | yes | D34 Post Vaccine | N/A | Positive | 2.0 |
| 10 | Female | 40-45 | yes | Pfizer | yes | yes | D34 Post Vaccine | N/A | Positive | 1.9 |
| 11 | Male | 30-35 | yes | Pfizer | yes | yes | D34 Post Vaccine | N/A | Positive | 1.8 |
| 12 | Female | 26-30 | yes | Pfizer | yes | yes | D34 Post Vaccine | N/A | Positive | 1.3 |
| 13 | Female | 26-30 | yes | Pfizer | yes | yes | D34 Post Vaccine | N/A | Positive | 1.8 |
| 14 | Female | 30-35 | yes | Pfizer | yes | yes | D34 Post Vaccine | N/A | Positive | 2.0 |
| 15 | Female | 26-30 | yes | Pfizer | yes | yes | D34 Post Vaccine | N/A | Positive | 2.4 |
| 16 | Female | 30-35 | yes | Pfizer | yes | yes | D33 Post Vaccine | N/A | Positive | 2.0 |
| 17 | Female | 26-30 | yes | Pfizer | yes | yes | D33 Post Vaccine | N/A | Positive | 1.8 |
| 18 | Male | 26-30 | yes | Pfizer | yes | yes | D31 Post Vaccine | N/A | Positive | 3.0 |
| 19 | Male | 56-60 | yes | Pfizer | yes | yes | D30 Post Vaccine | N/A | Positive | 0.9 |
| 20 | Female | 26-30 | yes | Pfizer | yes | yes | D30 Post Vaccine | N/A | Positive | 2.1 |
| 21 | Male | 50-55 | yes | Moderna | yes | yes | D36 Post Vaccine | N/A | Positive | 2.2 |
| 22 | Female | 56-60 | yes | Moderna | yes | yes | D38 Post Vaccine | N/A | Positive | 2.0 |
| 23 | Male | 30-35 | yes | Moderna | yes | yes | D35 Post Vaccine | N/A | Positve | 2.5 |

| | | | | | | | | | | |
|---|---|---|---|---|---|---|---|---|---|---|
| **24** | Male | 20-25 | yes | Moderna | yes | yes | D35 Post Vaccine | N/A | Positve | 2.4 |
| **25** | Female | 95-100 | yes | Moderna | yes | yes | D35 Post Vaccine | N/A | Positve | 2.7 |
| **26** | Female | 50-55 | yes | Moderna | yes | yes | D35 Post Vaccine | N/A | Positve | 1.9 |
| **27** | Female | 20-25 | yes | Moderna | yes | yes | D35 Post Vaccine | N/A | Positve | 2.4 |
| **28** | Male | 26-30 | yes | Moderna | yes | yes | D35 Post Vaccine | Negative | Positve | 2.5 |
| **29** | Female | 40-45 | yes | Moderna | yes | | D13-Post vaccine | positive | Positve | 1.8 |
| **30** | Female | 40-45 | yes | Pfizer | yes | | D11-Post Vaccine | positive | Positve | >3.5 |
| **31** | Male | 46-50 | yes | Moderna | yes | | D10-Post vaccine | positive | Positve | >3.5 |
| **32** | Male | 30-35 | yes | Moderna | yes | | D14-Post Vaccine | positive | Positve | >3.5 |
| **33** | Female | 60-65 | yes | Pfizer | yes | | D17-Post Vaccine | positive | Positve | >3.5 |
| **32** | Male | 30-35 | no | | | | pre-vaccine | positive | Positve | 1.6 |
| **33** | Female | 60-65 | no | | | | pre-vaccine | positive | Positve | 1.1 |

Table 2. Fold Enrichment in Amino Acid Motif Recognition by Different Subjects

| Subject ID # | [ACP]XXXDXSGI | DXSGIXXX[DGESN] | FXXELXX[WY]F | KWXXFQQ | [FY]XX[QE]LXXWF | P[EDT]WRXY | RXPX[ST]L[DE] | K[FY][LM]XFXQ | PXWR[IV]Y | [AC]K[FY]XXFXQ |
|---|---|---|---|---|---|---|---|---|---|---|
| 1 | 24.12188 | 28.89815 | 14.16188 | 2.90406 | 10.03086 | 1.63869 | 1.8324 | 2.86602 | 0.97399 | 1.14955 |
| 2 | 1.3568 | 2.80499 | 0.79658 | 8.14162 | 0.91802 | 2.94699 | 1.60712 | 4.3079 | 2.53118 | 2.10418 |
| 3 | 45.54796 | 42.14705 | 7.65677 | 15.33523 | 4.19016 | 10.15588 | 11.67805 | 6.29133 | 9.65864 | 4.88007 |
| 4 | 74.64611 | 52.41181 | 35.46319 | 23.11651 | 17.59506 | 3.7325 | 12.62332 | 9.11356 | 2.20935 | 6.17086 |
| 5 | 86.52744 | 75.57166 | 35.07616 | 7.11044 | 4.17209 | 2.62366 | 0.83056 | 2.41959 | 2.41142 | 1.62939 |
| 6 | 9.08279 | 9.20852 | 20.26051 | 4.23238 | 3.19377 | 4.01896 | 7.89146 | 2.39675 | 3.20445 | 3.09941 |
| 7 | 11.775 | 9.4349 | 0.34627 | 2.19791 | 0.53115 | 1.09487 | 1.67598 | 1.86742 | 1.91283 | 0.83012 |
| 8 | 12.75622 | 16.91751 | 26.06295 | 0.85512 | 10.78932 | 0.87751 | 1.168 | 1.58294 | 0.42967 | 1.7407 |
| 9 | 16.87036 | 18.89413 | 22.21202 | 31.01272 | 1.58234 | 22.15339 | 4.71435 | 14.63689 | 18.01009 | 9.6201 |
| 10 | 7.991 | 7.36731 | 24.80848 | 9.95705 | 23.19741 | 6.76044 | 7.69714 | 2.83034 | 4.01064 | 2.25895 |
| 11 | 8.81782 | 8.24756 | 6.1367 | 3.31935 | 1.65641 | 7.76581 | 5.46426 | 2.23179 | 5.67561 | 1.65069 |
| 12 | 33.42265 | 30.07481 | 17.76317 | 18.24522 | 7.05165 | 4.54149 | 3.89708 | 6.58812 | 4.62662 | 4.24812 |
| 13 | 21.65294 | 21.84343 | 58.46848 | 19.18538 | 26.32006 | 9.91419 | 20.31965 | 5.5046 | 5.86914 | 3.792 |
| 14 | 56.01096 | 48.77159 | 47.76832 | 9.07255 | 23.93239 | 11.28625 | 20.43179 | 3.24127 | 5.76119 | 2.20879 |
| 15 | 27.55929 | 28.09445 | 27.27589 | 28.66981 | 5.85507 | 23.93945 | 13.42879 | 11.62889 | 16.50785 | 7.58091 |
| 16 | 69.39203 | 72.35511 | 40.49051 | 82.80327 | 6.35543 | 5.81869 | 10.75896 | 31.03799 | 3.28103 | 25.63274 |
| 17 | 3.64422 | 4.90357 | 10.3055 | 2.92826 | 1.50749 | 2.41707 | 6.58392 | 2.84551 | 1.50774 | 1.42116 |
| 18 | 39.47938 | 30.3566 | 20.42899 | 22.36679 | 10.4027 | 11.71427 | 2.73141 | 7.92746 | 13.83002 | 5.05539 |
| 19 | 2.27761 | 3.50932 | 8.46049 | 2.84196 | 4.86742 | 1.2403 | 1.71339 | 1.13049 | 2.1335 | 1.25074 |
| 20 | 11.09478 | 10.25416 | 42.05175 | 28.66604 | 13.59703 | 34.38442 | 1.30943 | 5.73332 | 20.88919 | 3.42484 |
| 21 | 0.66991 | 1.17311 | 6.45055 | 2.99584 | 2.0799 | 0.64202 | 0.87316 | 1.43845 | 0.73027 | 1.2127 |
| 22 | 0.78098 | 0.56674 | 1.44845 | 2.59875 | 1.38039 | 3.30676 | 2.21652 | 1.26755 | 2.51584 | 1.28278 |
| 23 | 8.76277 | 9.59592 | 28.73647 | 10.94687 | 4.11469 | 2.28089 | 4.93592 | 5.0789 | 1.31525 | 2.97093 |
| 24 | 14.24855 | 16.75375 | 5.25025 | 45.38644 | 2.94386 | 23.11886 | 12.23777 | 21.57018 | 17.69763 | 15.08657 |
| 25 | 28.48567 | 27.45478 | 21.08092 | 20.42211 | 2.53462 | 16.57896 | 4.38707 | 9.4571 | 13.02877 | 5.76048 |
| 26 | 1.45674 | 1.00078 | 7.97533 | 3.71951 | 1.18545 | 2.23491 | 5.45684 | 1.57078 | 1.08261 | 1.1916 |
| 27 | 6.6744 | 7.92231 | 12.1883 | 6.86614 | 7.14451 | 6.0221 | 1.8916 | 4.05087 | 5.06876 | 2.38904 |
| 28 | 33.77127 | 36.42345 | 43.57688 | 17.74377 | 5.07247 | 10.1257 | 4.87788 | 5.65815 | 9.38512 | 4.34097 |
| 29 | 118.4104 | 96.1604 | 61.08806 | 41.86992 | 29.68794 | 26.41247 | 2.33624 | 21.91133 | 71.63942 | 13.09448 |
| 30 | 0.744 | 0.74156 | 77.70677 | 1.32981 | 41.33262 | 21.34096 | 4.75237 | 0.91951 | 8.95235 | 0.61937 |
| 31 | 3.77329 | 2.69862 | 57.97974 | 32.51077 | 40.72458 | 54.41257 | 29.78026 | 17.23821 | 44.7197 | 11.13788 |
| 32 | 11.83184 | 15.71697 | 55.50956 | 41.74956 | 29.17609 | 25.97491 | 2.97936 | 19.4578 | 14.60386 | 13.26798 |
| 33 | 1.95214 | 0.56701 | 46.74683 | 3.37901 | 30.57662 | 0.96797 | 3.16383 | 4.33146 | 0.79088 | 3.41584 |

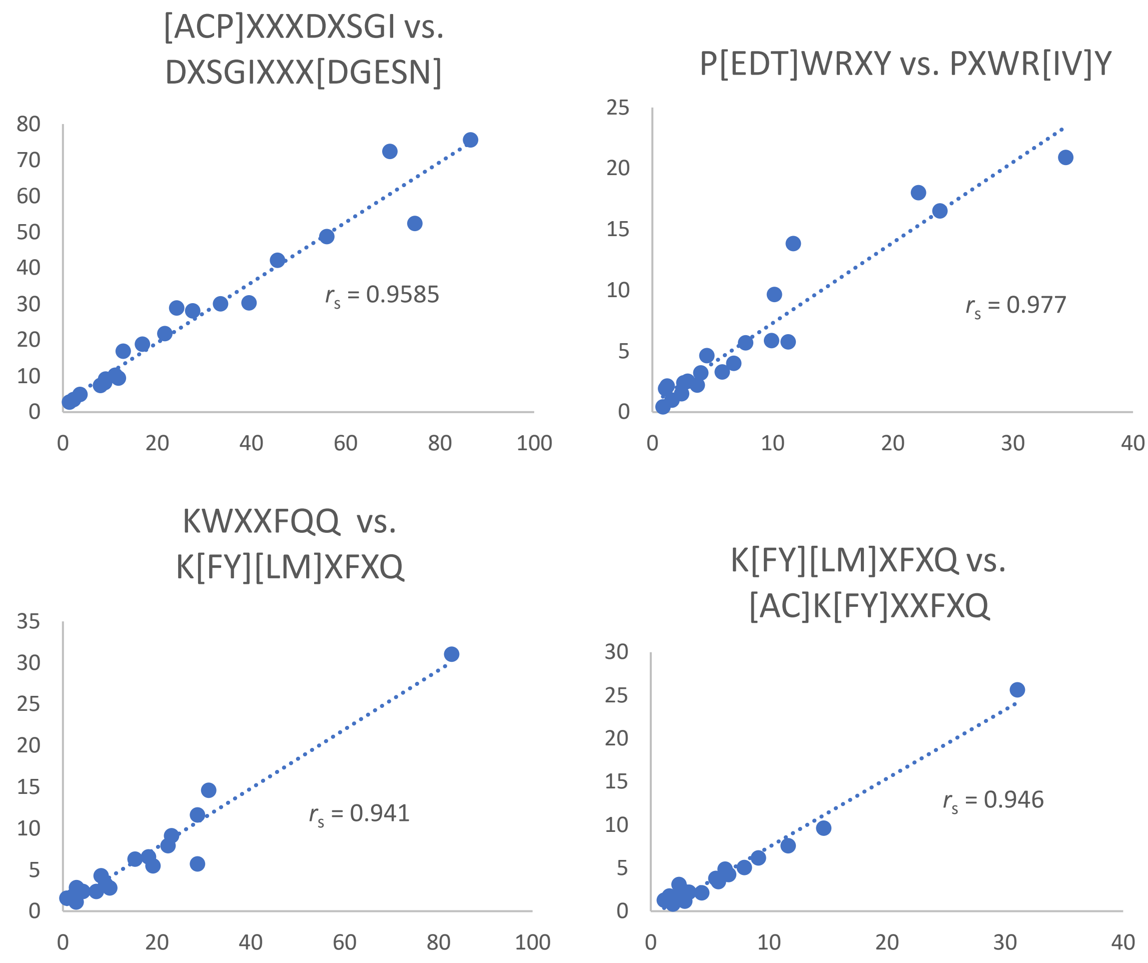

Figure S1. Correlation of IgG recognition of peptide motifs with overlapping amino acids. The fold enrichment in IgG with specificity for the different pairs of amino acid motifs (respective x and Y-axis) are shown along with the Spearman rank correlation ($r_s$) value. Each symbol represents a different COVID-19 mRNA vaccine recipient. Trend lines were fitted by linear regression. Spearman rank correlation ($r_s$) is displayed.

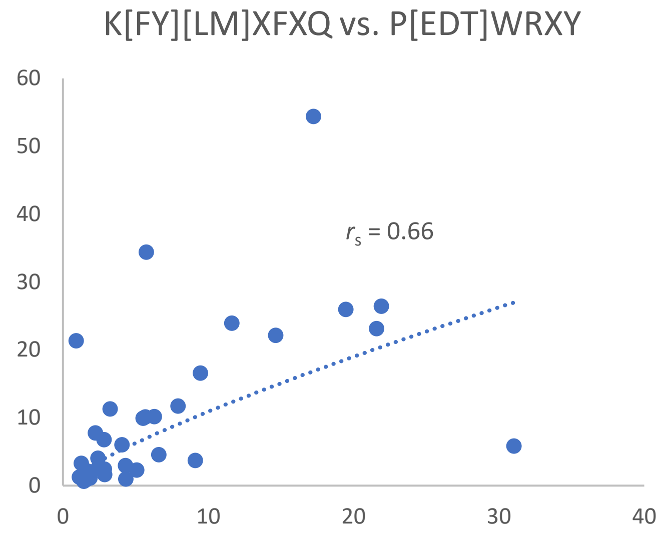

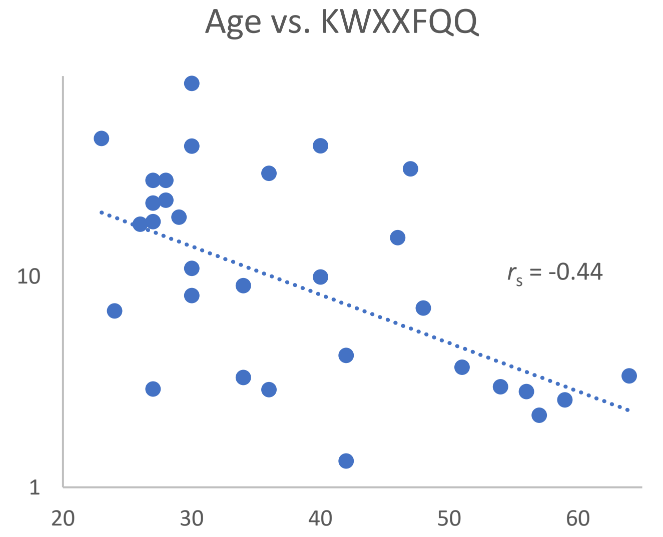

Figure S2. Correlation of amino acid motif recognition and age of vaccinated subjects. Panel A: The fold enrichment in IgG with specificity for the amino acid motifs K[FY][LM]XFXQ vs. P[EDT]WRXY is shown. Each symbol corresponds to a different vaccinated individual. Panel B: The fold enrichment in IgG with specificity for the amino acid motif KWXXFQQ (Y-axis) is plotted against age (X-axis). Trend lines were fitted by linear regression. Spearman rank correlation ($r_s$) is displayed.

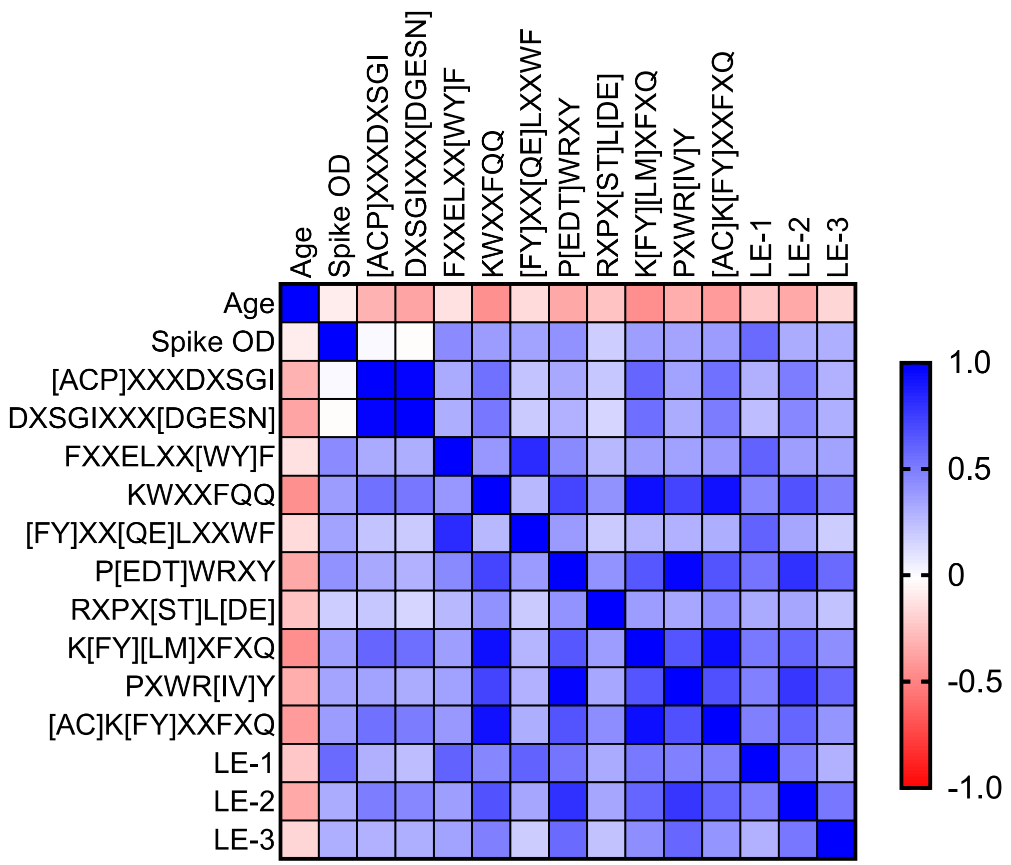

Correlation Matrix 1. The correlation between enrichment in IgG recognition of amino acid motifs and linear epitopes. Spearman rank correlation values are displayed as a heat map by color according to key on right for different epitopes recognized by IgG from COVID-19 mRNA vaccine recipients.

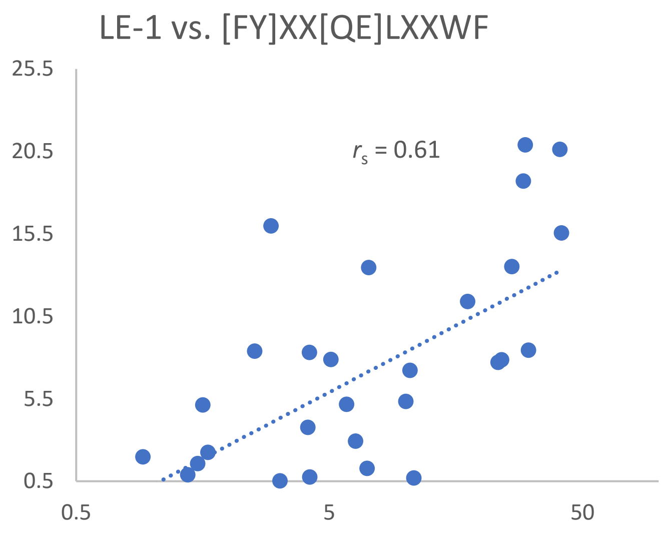

Figure S3. Correlation between IgG recognition of amino acid motif [FY]XX[QE]LXXWF and linear epitope LE-1. The PIWAS score (Y-axis) for IgG recognition of LE-1 for individual subjects was plotted vs. the fold enrichment in IgG (X-axis) with specificity for the amino acid motif [FY]XX[QE]LXXWF in subjects that received mRNA vaccine. Each symbol corresponds to a different vaccinated individual. Trend lines were fitted by linear regression. Spearman rank correlation ($r_s$) is displayed.

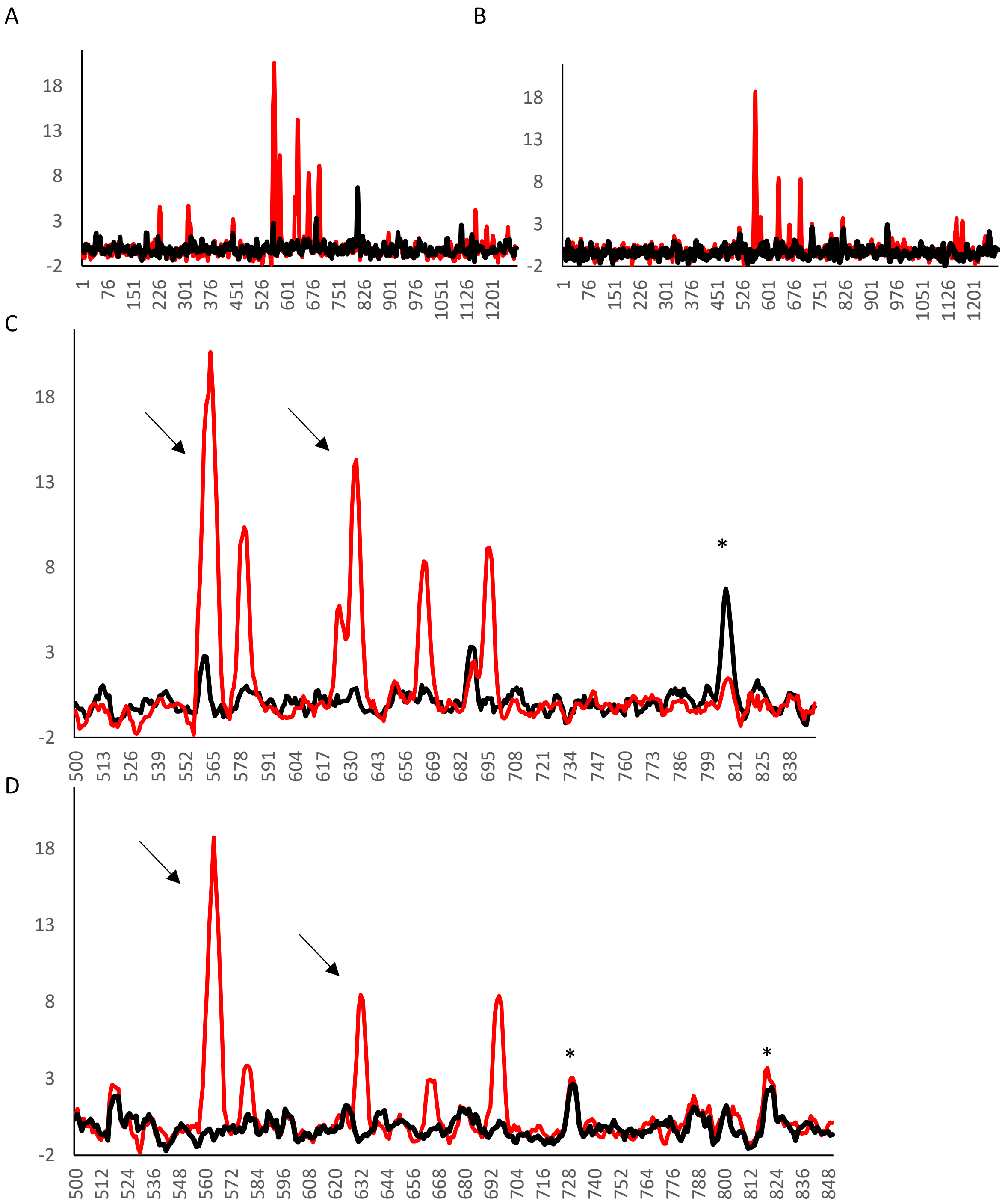

Figure S4. Examples of epitopes that drive IgG responses in mRNA vaccinated subjects with prior COVID-19. The PIWAS scores (Y-axis) are graphed for peptides surrounding different amino acids along the SARS-CoV-2 spike protein (X-axis) that are recognized by IgG after COVID-19/pre-vaccine (black) and after COVID-19 and vaccination (red). Panels A (subject 32) and B (subject 33) show entire spike protein and Panels C and D respectively focus on the region surrounding the S1/S2 junction. Note dominant epitope LE-1 and LE-2 (→) in both subjects and lack of substantial boost to highlighted naturally occurring epitopes (*)

List S1. Peptides with strongest identity to SARS-CoV-2 spike mRNA vaccine epitopes identified though BLAST® analysis (https://blast.ncbi.nlm.nih.gov/Blast.cgi?PAGE=Proteins)

**1) LE-1 Epitope: FLP*FQQFGRDIA***

>translation elongation factor Ts [Candidatus Omnitrophica bacterium]

Sequence ID: MBI1977219.1 Length: 198

```
Query  4   FQQFGRDIA  12
           FQQFGRDIA
Sbjct  92  FQQFGRDIA  100
```

**2) LE-2 Epitope: *DQLTPTWRVY***

>hypothetical protein [Thermoplasmata archaeon]

```
Query  1   DQLTPTWRV  9
           DQLTPTWRV
Sbjct  39  DQLTPTWRV  47
```

**3) LE-3 Epitope: F*KEELDKYFKN***

>MULTISPECIES: universal stress protein [Aquimarina]

Sequence ID: WP_074407094.1 Length: 283

```
Query  2    KEELDKYFKN  11
            KEELDKYFKN
Sbjct  205  KEELDKYFKN  214
```

| | Pre-vaccine samples from N=20 subjects later vaccinated with Pfizer-BioNTech | | | | | | | | | | | | | | | | | | | | Post-vaccine samples from N=20 subjects vaccinated with Pfizer-BioNTech | | | | | | | | | | | | | | | | | | | | Pre-vaccine samples from N=20 subjects later vaccinated with Pfizer-BioNTech | | | | | | | | Post-vaccine samples from N=5 pirio COVID-19 patients post vaccination Moderna recipients | | | | | upper 95% COVID patients (Haynes et al, ) |
|---|---|---|---|---|---|---|---|---|---|---|---|---|---|---|---|---|---|---|---|---|---|---|---|---|---|---|---|---|---|---|---|---|---|---|---|---|---|---|---|---|---|---|---|---|---|---|---|---|---|---|---|---|---|---|
| subject #--> | 1 | 2 | 3 | 4 | 5 | 6 | 7 | 8 | 9 | 10 | 11 | 12 | 13 | 14 | 15 | 16 | 17 | 18 | 19 | 20 | 1 | 2 | 3 | 4 | 5 | 6 | 7 | 8 | 9 | 10 | 11 | 12 | 13 | 14 | 15 | 16 | 17 | 18 | 19 | 20 | 21 | 22 | 23 | 24 | 25 | 26 | 27 | 28 | 29 | 30 | 31 | 32 | 33 | |
| Amino Acid | | | | | | | | | | | | | | | | | | | | | | | | | | | | | | | | | | | | | | | | | | | | | | | | | | | | | | |
| 1 | -0.15892 | -0.20323 | 0 | -0.20121 | -0.10224 | 0.293871 | -0.29944 | -0.10883 | -0.30876 | -0.05256 | -0.32497 | -0.08654 | 0 | 0.005729 | 0.262538 | -0.05138 | -0.15258 | -0.18394 | -0.44557 | -0.24883 | 0.17372 | -0.31253 | -0.20016 | -0.11706 | -0.14139 | 0.189077 | -0.17758 | 0.077367 | -0.30835 | 0 | -0.29597 | -0.29861 | 0.170318 | -0.03527 | -0.21174 | 0 | -0.13363 | -0.06152 | -0.01409 | -0.15452 | -0.27927 | 1.07249 | -0.3261 | -0.09242 | 0.019927 | 0 | -0.22765 | -0.02707 | -0.20676 | -0.11134 | -0.5098 | 0 | -0.11724 | 0.326517 |
| 2 | -0.46057 | -0.38189 | -0.28741 | -0.18167 | -0.04551 | 0.159168 | -0.26898 | -0.30815 | -0.40746 | 0.071569 | -0.55532 | -0.27061 | -0.32926 | -0.03971 | 0.209135 | -0.20159 | -0.13114 | -0.07394 | -0.62985 | 0.148136 | 0.105172 | -0.31253 | -0.31361 | -0.41859 | -0.42738 | 0.275459 | -0.48177 | 0.066945 | -0.41206 | -0.11444 | -0.59815 | -0.36275 | -0.08549 | -0.03309 | -0.1168 | -0.1153 | -0.28272 | -0.29725 | -0.32588 | -0.43424 | -0.19108 | 1.223145 | -0.39256 | -0.40307 | -0.15648 | 0.107963 | -0.22765 | -0.31606 | -0.42319 | -0.30712 | -0.62449 | -0.33351 | -0.23677 | 0.460217 |
| 3 | -0.17148 | -0.03207 | -0.41855 | -0.05123 | -0.10672 | 0.159168 | -0.56305 | -0.47419 | -0.36557 | -0.03261 | -0.63645 | -0.55532 | -0.25584 | -0.03898 | 0.209135 | -0.34466 | -0.05932 | -0.1419 | -0.61382 | 0.195001 | -0.01681 | -0.24923 | -0.0802 | -0.43076 | -0.39518 | 0.275459 | -0.54638 | -0.04394 | -0.45443 | -0.04173 | -0.47151 | -0.12629 | -0.31024 | 0.313669 | -0.14724 | -0.10352 | -0.41979 | -0.60051 | -0.61672 | -0.68483 | -0.00515 | 1.10288 | -0.39256 | -0.34382 | -0.15648 | -0.0552 | -0.40878 | -0.58953 | -0.51794 | -0.30712 | -0.78227 | -0.4001 | -0.53248 | 0.561684 |
| 4 | -0.2409 | -0.02331 | -0.41448 | -0.18745 | -0.10672 | 0.088397 | -0.82279 | -0.74672 | -0.50553 | -0.0939 | -0.92286 | -0.83041 | -0.28797 | -0.03898 | -0.0258 | -0.59901 | -0.05932 | -0.14053 | -0.56224 | -0.20999 | 0.203172 | -0.38049 | -0.00822 | -0.68354 | -0.61186 | 0.070152 | -0.58488 | -0.1126 | -0.5813 | 0.086094 | -0.77347 | 0.095034 | -0.31024 | 0.264046 | -0.25251 | -0.05232 | -0.65619 | -0.60051 | -0.66766 | -0.93571 | 0.038246 | 0.853311 | -0.66631 | -0.48485 | -0.15648 | -0.0552 | -0.57928 | -0.93756 | -0.55324 | -0.30712 | -0.76882 | -0.41271 | -0.81982 | 0.561973 |
| 5 | -0.48238 | 0.217313 | -0.64942 | -0.10568 | 0.203703 | 0.208753 | -0.9308 | -1.02676 | -0.45187 | -0.32726 | -1.37153 | -0.94327 | -0.45628 | -0.19693 | 0.022257 | -0.525 | -0.18737 | -0.35352 | -0.71373 | -0.28153 | 0.203172 | -0.53453 | -0.0706 | -0.69573 | -0.73909 | 0.280331 | -0.88047 | -0.18683 | -0.71013 | -0.02462 | -0.30501 | 0.161449 | -0.31024 | 0.12969 | -0.12587 | 0.01572 | -0.77408 | -0.77386 | -0.85808 | -0.86599 | 0.221676 | 0.597577 | -0.71926 | -0.83221 | -0.10761 | -0.13248 | -0.61252 | -1.06006 | -0.6526 | 0.098692 | -1.0374 | -0.51789 | -0.87494 | 0.745209 |
| 6 | -0.72278 | 0.19777 | -0.53881 | -0.27924 | 0.484992 | -0.08712 | -0.99648 | -0.95904 | -0.102 | -0.17154 | -1.18635 | -0.90627 | -0.67193 | -0.32138 | -0.21767 | -0.45865 | -0.07042 | -0.13372 | -0.23023 | 0.014224 | -0.20376 | -0.29401 | 0.190503 | -0.61315 | -0.97992 | 0.248114 | -0.99837 | -0.3917 | -0.41788 | -0.07817 | 0.026715 | 0.718937 | -0.7197 | -0.22075 | 0.197753 | -0.12721 | -0.79631 | -0.87168 | -0.67086 | -0.83226 | 0.69076 | -0.29417 | -0.73774 | -0.74016 | -0.36623 | -0.14037 | -0.39448 | -0.98511 | -0.52118 | 0.084431 | -0.6468 | -0.29521 | -0.64129 | 0.907273 |
| 7 | -0.11337 | 0.104006 | -0.37428 | 0.811509 | 0.17386 | 0.087716 | -1.18325 | -0.90282 | 0.10986 | -0.49472 | -1.06049 | -0.93738 | -0.38595 | -0.20665 | -0.37273 | -0.46252 | -0.03417 | -0.44406 | -0.41584 | -0.60505 | -0.05025 | -0.35943 | 0.362748 | -0.4325 | -0.58445 | 0.157199 | -0.60142 | -0.49019 | -0.47358 | 0.306798 | 0.117088 | 0.714981 | -0.35065 | -0.40348 | -0.23632 | -0.15682 | -0.62612 | -0.85861 | -0.78136 | -0.77623 | 0.850653 | -0.47569 | -0.80946 | -0.61916 | -0.17256 | -0.07176 | -0.62409 | -0.95551 | -0.7314 | 0.849172 | 0.71422 | 0.020172 | -0.85111 | 1.075244 |
| 8 | -0.46849 | -0.03583 | -0.86969 | 0.438147 | 0.216885 | -0.26706 | -0.98857 | -1.22549 | 0.448264 | -0.5136 | -0.75553 | -0.49114 | -0.41381 | -0.51684 | -0.52774 | 0.078055 | -0.46513 | -0.24958 | -0.55144 | -0.76782 | -0.2493 | 0.266989 | -0.37007 | -0.89698 | -0.60654 | -0.12859 | -0.87176 | -0.57592 | 0.140877 | 0.17934 | -0.15238 | 0.533694 | -0.02249 | -0.47937 | -0.31848 | 0.137309 | -0.54956 | -0.36253 | -0.91763 | -0.56397 | 0.661264 | -0.61859 | -1.08119 | -0.69779 | -0.49845 | 0.277751 | -0.54283 | -0.35234 | -0.65433 | 1.313619 | -0.81248 | 0.11237 | -0.62644 | 1.244681 |
| 9 | -0.35389 | 0.548872 | -1.00124 | 0.693813 | 0.028816 | -0.50092 | -0.67289 | -1.34849 | 0.489945 | -0.3734 | -0.3654 | -0.52907 | -0.1927 | -0.55238 | -0.77461 | 0.65714 | -0.53903 | -0.24153 | -0.96849 | -0.80074 | -0.5679 | 0.974422 | -0.41038 | -0.72275 | -0.679 | -0.10591 | -0.78677 | -0.95118 | 0.647888 | -0.07707 | 0.419286 | 0.163086 | 0.087974 | -0.76687 | -0.28921 | 0.41373 | -0.52626 | -0.36948 | -0.91505 | -0.12008 | 0.621596 | -0.46215 | -0.97565 | -0.76077 | -0.49048 | 0.136123 | -0.33314 | -0.29767 | -0.93477 | 1.237627 | -1.07621 | -0.0286 | -0.38865 | 1.336403 |
| 10 | -0.42062 | 0.344314 | -0.83295 | 0.495548 | -0.44214 | -0.81316 | -0.57514 | -1.04593 | 0.341025 | -0.00744 | 0.002228 | -0.49516 | -0.33547 | -0.03354 | -1.02732 | 0.410616 | -0.3383 | -0.25491 | -0.78594 | -0.88532 | -0.93192 | 1.230336 | -0.40232 | -0.81261 | -0.52121 | -0.34433 | -0.41492 | -1.16707 | 0.952891 | -0.10403 | -0.0263 | 0.035628 | -0.03684 | -0.36217 | -0.44934 | 0.297218 | -0.66338 | -0.23707 | -0.70184 | -0.06605 | 0.384531 | -0.36453 | -0.30701 | -0.12037 | -0.57122 | 0.212819 | -0.35002 | -0.48768 | -1.06861 | 0.904127 | -1.01773 | -0.08427 | -0.40505 | 1.556266 |
| 11 | -0.16246 | 0.288364 | -1.11812 | 0.798302 | -0.89425 | -1.03057 | -0.21681 | -1.09238 | -0.03979 | -0.20241 | -0.00506 | -0.32904 | -0.18624 | 0.260843 | -1.42103 | 0.458107 | -0.25322 | -0.38332 | -0.96841 | -0.82255 | -0.81811 | 0.948141 | -0.9187 | -0.48677 | -0.27971 | -0.45978 | -0.04594 | -1.29909 | 0.898929 | -0.24466 | -0.08242 | -0.24355 | 0.272426 | -0.08757 | -0.86402 | 0.592893 | -0.65181 | -0.21196 | -1.05064 | 0.016132 | 0.242209 | -0.88677 | 0.266876 | -0.17496 | -0.38517 | 0.894443 | -0.12615 | -0.87139 | -1.28606 | 1.638983 | -0.37599 | -0.38675 | 0.205665 | 1.350051 |
| 12–162 | [illegible] | | | | | | | | | | | | | | | | | | | | | | | | | | | | | | | | | | | | | | | | | | | | | | | | | | | | | |

| | | | | | | | | | | | | | | | | |
|---|---|---|---|---|---|---|---|---|---|---|---|---|---|---|---|---|
| 163 | -1.26269 | -0.07211 | -0.51914 | -1.06673 | 0.877132 | -0.23189 | -0.55888 | -0.08247 | 0.815213 | 0.385961 | -0.01105 | -0.14877 | -0.7347 | -0.62888 | 0.771782 | [illegible] |
| 164 | -1.14564 | -0.09472 | -0.45665 | -1.02688 | 1.182404 | -0.27306 | -0.60085 | -0.21462 | 0.216369 | 0.606494 | -0.79241 | -0.03196 | -0.66237 | -0.51452 | 0.849748 | [illegible] |
| 165 | -0.88987 | -0.60371 | -0.20133 | -1.09704 | 1.738722 | -0.03226 | -0.86204 | -0.37178 | -0.21333 | 0.395861 | -0.58104 | -0.27163 | -0.45307 | -0.49166 | 0.76075 | [illegible] |
| 166 | -0.81856 | -0.33786 | -0.26011 | -0.80822 | 1.592212 | -0.04207 | -0.11199 | 0.044047 | -0.60934 | 0.444951 | -0.56786 | 0.210261 | 0.117007 | -0.59926 | 0.576801 | [illegible] |
| 167 | -0.63812 | -0.42554 | -0.26289 | -0.95671 | 1.634223 | -0.17713 | -0.54311 | -0.43946 | -0.96508 | 0.551484 | -0.54555 | 0.127498 | -0.19764 | -0.81134 | 0.3943 | [illegible] |
| 168 | -0.60715 | -0.49608 | -0.24023 | -0.43763 | 1.429709 | -0.14718 | -0.51593 | -0.8651 | -1.02611 | 1.188343 | -0.73659 | 0.350849 | -0.28184 | -1.0796 | 0.427381 | [illegible] |
| 169 | -0.58758 | -0.68318 | -0.27936 | -0.53293 | 0.178344 | -0.08326 | -0.45226 | -0.31657 | -0.12696 | 0.862386 | -0.37412 | 0.436483 | -0.64161 | -0.94733 | 0.910615 | [illegible] |
| 170 | -0.68111 | -0.63743 | -0.21873 | -0.26818 | -0.73696 | -0.04373 | -0.30069 | -0.16285 | 0.170026 | 0.960352 | -0.51761 | 0.624581 | -1.12488 | -0.56487 | 0.607713 | [illegible] |
| 171 | 0.230913 | -0.54794 | -0.09602 | -0.26945 | -1.03088 | -0.09131 | -0.81235 | -0.50861 | -0.077 | 0.324533 | -0.44423 | -0.06578 | -1.31741 | -0.4782 | -0.21112 | [illegible] |
| 172 | 1.011752 | -0.52969 | -0.05996 | 0.269791 | -1.10792 | -0.21137 | -0.79931 | -0.06923 | -0.18812 | -0.1561 | -1.0197 | -0.32032 | -1.38249 | -0.6696 | -0.54916 | [illegible] |
| 173 | 1.228072 | -0.452 | 0.085216 | 0.043962 | -1.01277 | -0.01952 | -0.81577 | 0.228467 | -0.34605 | -0.47504 | -1.03242 | -0.10136 | -1.11703 | -0.38325 | -0.94218 | [illegible] |
| 174 | 1.019929 | -0.54578 | 0.184858 | 0.593402 | -0.65037 | -0.04911 | -0.73923 | -0.33528 | -0.91263 | -0.12183 | -1.39664 | -0.45373 | -0.60461 | -0.56413 | -1.62737 | [illegible] |
| 175 | 0.868116 | -0.14974 | 0.204071 | 0.574056 | -0.64946 | 0.322362 | -0.74616 | -0.68281 | -1.17349 | -0.30671 | -1.88851 | -0.77985 | -0.29364 | -0.73788 | -0.80886 | [illegible] |
| 176 | 0.156409 | 0.032452 | 0.160404 | 0.46315 | -0.67282 | 0.909738 | -1.04382 | -0.24664 | -0.89489 | 0.093043 | -2.15935 | -0.63388 | -0.48822 | -0.69396 | -0.28545 | [illegible] |
| 177 | -0.53509 | -0.00074 | 0.077473 | 0.452369 | -0.69622 | 1.155621 | -0.77449 | 0.251768 | -0.52241 | -0.07886 | -1.60014 | -0.25395 | -0.31586 | -0.37739 | -0.00891 | [illegible] |
| 178 | -0.25553 | -0.12606 | -0.18416 | 0.355942 | -0.83223 | 0.939151 | -0.92641 | 0.124174 | -0.70481 | -0.66275 | -1.40198 | -0.88359 | -0.12015 | -0.37898 | 0.314651 | [illegible] |
| 179 | -0.06125 | -0.03916 | -0.53693 | -0.331 | -1.12989 | 0.981359 | -0.76016 | 0.114913 | -0.96415 | -0.79047 | -1.12049 | -0.88926 | -0.48672 | -0.33764 | 0.490403 | [illegible] |
| 180 | -0.24932 | -0.1263 | -0.98509 | -0.3985 | -1.32267 | 0.369373 | -0.89898 | 0.080386 | -1.09967 | -0.71428 | -0.72957 | -0.3582 | -0.57823 | -0.58109 | -0.36648 | [illegible] |
| 181 | -0.23911 | -0.50914 | -1.22519 | -0.14319 | -1.15121 | -0.50879 | -0.5886 | -0.13483 | -1.22175 | -0.77635 | -0.37007 | 0.013898 | -0.44234 | -1.00146 | -0.2039 | [illegible] |
| 182 | -0.75839 | -0.51973 | -1.28843 | -0.02337 | -1.16352 | -0.48811 | -0.54091 | -0.67629 | -1.35795 | -0.68906 | -0.32515 | 0.011626 | -0.10103 | -1.11078 | -0.18542 | [illegible] |
| 183 | -0.72657 | -0.61208 | -1.24562 | -0.06999 | -0.96518 | -0.45307 | -0.51069 | -0.35944 | -1.46839 | -0.28636 | -0.52257 | 0.136958 | -0.21968 | -1.15713 | -0.15168 | [illegible] |
| 184 | -0.09026 | -0.68146 | -0.84421 | 0.102517 | -0.63677 | -0.50305 | -0.75478 | -0.17542 | -1.20415 | -0.27135 | -0.36863 | 0.541989 | 0.027536 | -1.2773 | -0.42109 | [illegible] |
| 185 | -0.89915 | -0.12317 | -0.56118 | 0.017248 | -0.21192 | -0.26116 | -0.48915 | -0.19028 | -0.73357 | -0.07032 | -0.18203 | 0.049573 | 0.150154 | -0.93901 | -0.27325 | [illegible] |
| 186 | [illegible] | | | | | | | | | | | | | | | |
| 187–329 | [illegible] | | | | | | | | | | | | | | | |

| | | | | | | | | | | | | | | | | | | | | | | | | | | | | | | | | | | | | | | | | | | | | | | | | | | | | | |
|---|---|---|---|---|---|---|---|---|---|---|---|---|---|---|---|---|---|---|---|---|---|---|---|---|---|---|---|---|---|---|---|---|---|---|---|---|---|---|---|---|---|---|---|---|---|---|---|---|---|---|---|---|---|
| 330 | 0.350811 | -0.6315 | -0.74591 | -1.19649 | -0.66174 | -0.45295 | -0.69659 | -0.45248 | -0.27546 | -0.60507 | -0.65273 | -0.04922 | -0.19178 | -1.2474 | -0.39859 | 0.019979 | -1.03417 | -1.13104 | 0.182278 | -0.86608 | 0.260469 | -0.23146 | -0.60629 | -0.80367 | -0.79115 | -0.63997 | -0.19338 | -0.21903 | -0.90535 | 0.046775 | 0.388566 | -0.41699 | 0.594935 | -0.511 | -0.78488 | 0.393034 | -0.9899 | -0.34936 | 0.593997 | -0.36731 | -0.53981 | -0.41164 | -0.56469 | 0.250978 | 0.080959 | -0.20802 | -0.13268 | -0.45843 | 0.083954 | -0.03375 | -0.1052 | 0.030859 | -1.09382 |
| 331 | 0.599516 | -0.92635 | -0.74994 | -1.45195 | -0.69956 | -0.56631 | -0.28147 | -0.54014 | -0.3648 | -0.56431 | -0.82384 | -0.12511 | -0.08308 | -1.02259 | -0.3194 | 0.314224 | -0.79181 | -0.75952 | 0.216216 | -1.05168 | 0.564302 | -0.02354 | -0.89792 | -0.85095 | -0.85591 | -0.69321 | -0.37623 | -0.44554 | -0.73343 | 0.065829 | 0.20047 | -0.53141 | 0.662284 | -0.18852 | -0.42194 | 0.448806 | -0.45054 | 0.07622 | 0.111646 | -0.33396 | -0.31081 | -0.40783 | -0.69923 | 0.086558 | 0.257809 | -0.43411 | -0.25901 | -0.48083 | 0.921177 | 0.270682 | 0.044812 | -0.26437 | -1.01264 |
| 332 | 0.597332 | -0.64212 | -0.32991 | -1.09131 | -0.71361 | -0.26895 | -0.51474 | -0.69839 | -0.29365 | -0.63292 | -0.75177 | 0.328241 | 0.344003 | -0.46337 | -0.36295 | 0.40104 | -1.23678 | -0.65389 | 0.11895 | -0.57757 | 0.480065 | -0.11463 | -0.92326 | -1.01192 | -1.06663 | -0.62276 | -0.41733 | -0.56948 | -0.73059 | 0.271973 | 0.174842 | -0.22119 | 0.692321 | -0.27916 | -0.46164 | 0.217391 | -0.36812 | 0.023559 | -0.05053 | 0.011936 | -0.29525 | -0.58184 | -0.8324 | 0.333053 | 0.28838 | -0.04693 | -0.5172 | -0.19388 | 1.131796 | -0.14606 | 0.242873 | -0.10749 | -0.75938 |
| 333 | 0.483543 | -0.88478 | -0.38561 | -0.67111 | -0.80734 | -0.26944 | -0.59414 | -0.38946 | -0.51782 | -0.25604 | -1.07208 | -0.08395 | 0.33556 | -0.53171 | -0.22619 | 0.585563 | -0.74364 | -0.45762 | -0.28228 | -0.41003 | 0.150889 | -0.24416 | -0.67028 | -0.75278 | -1.06689 | -0.54101 | -0.27979 | -0.51746 | -0.46292 | 0.545352 | -0.18537 | -0.00277 | 0.692321 | -0.25627 | -0.40746 | 0.098513 | -0.42166 | 0.223505 | -0.11927 | -0.0701 | -0.15829 | -0.62921 | -0.89368 | 0.34857 | -0.08356 | -0.16669 | -0.63633 | -0.00238 | 1.675552 | 0.082064 | -0.08302 | -0.0966 | -0.39438 |
| 334 | 0.283367 | -1.05334 | -0.91382 | -0.21108 | -0.53896 | -0.11885 | -0.53423 | -0.5939 | -0.19344 | -0.3061 | -0.97794 | 0.033308 | 0.242303 | -0.25037 | -0.30599 | 0.455141 | -0.63062 | -0.8337 | -0.49938 | -0.16444 | 0.18008 | -0.36056 | -0.80963 | -0.40383 | -0.66132 | -0.45894 | -0.16227 | -0.49712 | -0.12652 | 0.719404 | -0.38252 | -0.24363 | -0.02176 | -0.35305 | -0.68538 | -0.42438 | -0.37702 | 0.063284 | -0.13963 | -0.13199 | -0.37207 | -0.63937 | -0.73027 | -0.10228 | -0.38179 | 0.268833 | -0.90028 | -0.10238 | 1.393162 | -0.41091 | -0.08505 | 0.049529 | -0.56736 |
| 335 | -0.05864 | -0.79075 | -0.47725 | 0.035453 | -0.71836 | 0.139729 | -1.00012 | -0.03665 | -0.00088 | -0.74825 | -0.87581 | -0.14928 | -0.08839 | -0.29751 | -0.12129 | 0.091663 | -0.56312 | -0.57919 | -0.29482 | -0.25164 | -0.27284 | -0.5673 | -0.7629 | 0.131867 | -0.94196 | -0.3966 | -0.44323 | -0.47532 | 0.071672 | 0.04528 | -0.46406 | 0.045458 | -0.1518 | -0.49044 | -0.43241 | -0.2733 | 0.353014 | -0.24288 | -0.15674 | 0.472677 | -0.38852 | -0.55667 | -0.38446 | 0.349031 | -0.01025 | 0.518676 | -0.8395 | 0.178623 | 1.007568 | -0.90412 | -0.45793 | 0.150388 | -0.74584 |
| 336 | -0.82509 | -0.61556 | -0.33937 | -0.10802 | -0.65992 | 0.057783 | -0.86016 | 0.119805 | 0.64188 | -0.69 | -0.62503 | -0.27299 | -0.05108 | -0.24728 | -0.22856 | 0.354985 | -0.87931 | -0.99008 | -0.55917 | -0.25749 | -1.20498 | -0.60439 | -0.92133 | 0.19814 | -0.72562 | 0.031221 | -0.49115 | -0.24265 | 0.069168 | 0.224892 | -0.0056 | -0.2541 | -0.24976 | -0.77812 | -0.76426 | -0.24199 | -0.27561 | -0.532 | -0.56545 | 0.447366 | -0.28029 | -0.07172 | -0.32595 | 0.125855 | -0.27758 | 0.822962 | -1.01295 | 0.403489 | 0.280855 | -1.01531 | -0.55981 | -0.09867 | -0.66362 |
| 337 | -0.8785 | -0.55025 | 0.440205 | -0.17301 | -0.90391 | -0.02737 | -0.73048 | -0.29296 | 0.67552 | -0.32759 | -0.57868 | -0.73388 | -0.07598 | -0.92019 | -0.25606 | 0.218473 | -0.54934 | -0.84624 | -0.39516 | -0.53357 | -1.39465 | -0.6295 | -0.9218 | 0.220764 | -0.69484 | -0.0717 | -0.27043 | -0.38172 | -0.12887 | -0.02218 | 0.097283 | -0.37373 | 0.192978 | -0.76575 | -0.92073 | 0.369018 | 0.186925 | -0.41266 | -0.43471 | 0.158868 | 0.027192 | 0.375757 | -0.12032 | 0.125956 | -0.111 | 0.44142 | -0.91107 | -0.01369 | 1.50549 | -0.82072 | -0.73545 | -0.18809 | -0.38411 |
| 338 | -0.79496 | -0.22725 | 0.753155 | 0.076723 | -0.89723 | -0.34971 | -0.50071 | -0.50624 | 0.884564 | -0.40524 | -0.33537 | -0.46952 | -0.29248 | -0.93826 | -0.16888 | 0.809881 | -0.46202 | -0.99986 | -0.06552 | -0.50991 | -1.20105 | -0.5592 | -0.99527 | 0.463624 | -0.78679 | -0.39748 | -0.23359 | -0.39018 | -0.50629 | 0.208096 | 0.452331 | -0.43349 | 0.254613 | -1.06117 | -0.8148 | 1.169366 | 0.39574 | -0.28737 | -0.1864 | 0.4207 | 0.017531 | 0.732465 | -0.10248 | 0.194476 | 0.254895 | 0.583563 | -0.9025 | -0.4776 | 1.073634 | -0.72306 | -0.68624 | -0.17707 | -0.6382 |
| 339 | -0.76017 | -0.12581 | 1.291718 | -0.01132 | -0.45543 | -0.56004 | -0.44461 | 0.013522 | 0.353941 | -0.04893 | -0.15523 | -0.38978 | -0.67299 | -0.59578 | 0.069417 | 1.07107 | -0.37163 | -0.48163 | 2.11788 | -0.63577 | -1.00127 | -0.26787 | -0.9355 | 0.111188 | -0.44703 | -0.37329 | -0.34631 | -0.46351 | -0.87928 | -0.07861 | 0.431776 | -0.44757 | 0.479191 | -0.5288 | -0.72556 | 1.612925 | 0.377905 | 0.004355 | 1.508497 | 0.491924 | 0.191407 | 0.695993 | -0.00534 | 0.215559 | 0.836496 | 0.48676 | -0.64984 | -0.58173 | 1.287539 | -0.78146 | -0.6662 | 0.493573 | -0.73401 |
| 340 | -0.69818 | -0.28767 | 0.982024 | -0.02412 | 0.27153 | -1.02979 | -0.27368 | -0.27077 | 0.563756 | 0.261259 | -0.34086 | 0.234333 | -0.8961 | -0.60086 | -0.3088 | 0.976884 | -0.70586 | -0.07002 | 2.659319 | -0.73137 | -0.64763 | -0.22461 | -0.7782 | -0.11485 | 0.307009 | -0.37428 | -0.41108 | -0.71827 | -0.74407 | 0.209906 | 0.305064 | -0.49495 | -0.08613 | 0.108323 | -0.6789 | 1.386216 | -0.31313 | -0.032 | 1.752724 | 0.01915 | 0.200167 | 0.631438 | 0.26793 | -0.14193 | 1.27069 | 0.043424 | -0.87203 | -0.81917 | 0.647732 | -0.26785 | -1.19781 | 1.055351 | -0.71469 |
| 341 | 0.310224 | -0.43423 | 0.657306 | 0.466332 | 0.160575 | -1.35328 | -0.41194 | -0.55497 | -0.37576 | -0.03431 | -0.57421 | 0.387177 | -1.54639 | -0.34258 | -0.20154 | 0.43443 | -0.51915 | 0.085356 | 2.865724 | -0.83866 | 0.486279 | -0.23286 | -0.71116 | -0.18149 | 0.494645 | -0.57655 | -0.22057 | -0.45952 | -0.45531 | -0.11687 | 0.046241 | 0.428093 | -0.31915 | 0.310094 | -0.43691 | 1.148388 | -0.07542 | 0.048086 | 2.030567 | 0.283416 | -0.09248 | 0.292867 | -0.05404 | -0.21508 | 1.440379 | -0.11552 | -0.64447 | -0.89238 | 0.300657 | -0.19018 | -0.83641 | 0.935878 | -1.16378 |
| 342 | 0.986709 | -0.55605 | -0.16265 | 0.377382 | 0.111771 | -1.24779 | -0.60843 | -0.37071 | -0.90215 | -0.44245 | -0.66418 | 0.643145 | -1.71852 | 0.122625 | -0.43165 | 0.515647 | -0.81385 | 0.30194 | 2.368844 | -0.61314 | 1.283376 | -0.20776 | -0.54297 | 0.745137 | 0.403435 | -0.03907 | -0.28888 | -0.28438 | -0.23121 | -0.0695 | -0.28523 | 1.068249 | -0.58017 | 0.277655 | -0.05478 | 0.755834 | -0.53795 | -0.28643 | 2.156488 | 0.311525 | -0.187 | -0.15461 | -0.18616 | -0.3678 | 0.771412 | -0.13036 | -0.49467 | -0.91079 | -1.31993 | -0.0657 | -1.00978 | 0.798851 | -1.31066 |
| 343 | 0.900802 | -0.62174 | -0.23652 | -0.0021 | 0.453541 | -1.1244 | -0.83112 | -0.3241 | -0.91001 | -0.50036 | -0.62539 | 0.383254 | -1.50036 | 0.452735 | -0.4757 | -0.26733 | -1.15667 | 0.574924 | 2.063683 | -0.59286 | 1.644978 | -0.04156 | -0.26537 | 0.434667 | 0.461717 | 0.121045 | -0.6234 | -0.41961 | 0.183641 | -0.3674 | -0.42187 | 0.817125 | -0.84012 | 0.367142 | -0.09472 | -0.00991 | -0.66371 | -0.58679 | 1.902023 | 0.113489 | -0.14331 | -0.029 | -0.07845 | -0.43632 | 0.372813 | -0.42243 | -0.41071 | -0.591 | -1.74529 | -0.18749 | -1.13865 | 0.827144 | -1.05656 |
| 344 | 0.753092 | -0.56375 | -0.4015 | -0.10849 | 0.19225 | -0.95874 | -0.81205 | -0.48798 | -0.55579 | -0.35756 | -0.51229 | 0.35695 | -1.56976 | -0.14342 | -0.70908 | -0.52852 | -0.84781 | 0.320481 | 0.144761 | -0.53195 | 1.495734 | -0.09807 | -0.58478 | 0.572759 | 0.198842 | 0.219454 | -0.25383 | -0.34627 | 0.331147 | 0.112851 | -0.2871 | 1.072062 | -0.7339 | 0.519657 | -0.15387 | -0.09327 | -0.57858 | -0.84628 | 0.171748 | 0.238954 | -0.08507 | 0.209948 | -0.17373 | -0.19062 | 0.028284 | -0.34156 | -0.27269 | -0.4928 | -1.57397 | -0.32544 | -1.03946 | 0.169277 | -0.80383 |
| 345 | 0.672573 | -0.49774 | -0.50056 | -0.38386 | -0.48816 | -0.38396 | -0.73537 | -0.22581 | -0.88359 | -0.58988 | -0.4368 | -0.07456 | -1.33714 | 0.009233 | -0.335 | -0.06537 | -0.52111 | 0.078556 | -0.81542 | -0.60284 | 1.357994 | -0.15405 | -0.55473 | 0.248152 | -0.235 | 0.017011 | -0.28013 | 0.050508 | 0.228411 | 0.112851 | -0.22821 | 0.959277 | -0.43134 | 0.204224 | -0.39677 | -0.10419 | -0.25551 | -0.58602 | -0.0399 | 0.275167 | 0 | 0.042858 | -0.24799 | -0.21641 | -0.758 | -0.38254 | -0.10653 | -0.46135 | -1.21226 | -0.72251 | -0.39462 | -0.18446 | -0.4675 |
| 346 | -0.1108 | -0.25207 | -0.39353 | -0.73621 | -0.40646 | -0.03795 | -0.70113 | 0.044549 | -0.66766 | -0.15624 | -0.2936 | -0.63584 | -0.83731 | -0.19327 | -0.47819 | 0.201199 | -0.51383 | 0.078 | -1.02194 | 0.081268 | 0.427093 | -0.12309 | -0.33037 | -0.06604 | -0.4036 | 0.197159 | -0.37904 | -0.16321 | -0.3252 | -0.16986 | 0.102181 | 0.255449 | -0.43286 | -0.05035 | -0.52268 | 0.026364 | -0.2098 | -0.6196 | -0.30718 | 0.036213 | 0 | -0.22029 | 0.108137 | -0.23838 | -0.99339 | -0.38254 | -0.18561 | -0.42858 | -1.09821 | -0.72251 | -0.74828 | 0.194756 | -0.35996 |
| 347 | -0.83113 | 0.352831 | -0.35315 | -0.76241 | -0.12157 | -0.15361 | -0.23253 | -0.26673 | -0.46567 | -0.16833 | -0.18322 | -0.5215 | -0.7903 | -0.10418 | -0.21263 | -0.05048 | -0.31786 | 0.254334 | -0.92008 | -0.12798 | -0.25556 | 0.195656 | -0.34134 | -0.89975 | -0.29811 | 0.09733 | 0.047287 | -0.17606 | -0.40895 | 0.033249 | 0.578759 | -0.18643 | -0.68681 | 0.278271 | -0.26117 | -0.11464 | -0.03099 | -0.24305 | -0.55808 | -0.01486 | -0.11599 | -0.47817 | -0.20213 | -0.24732 | -1.00963 | -0.49647 | -0.37896 | -0.48715 | -0.87008 | -0.89489 | -0.85652 | 0.012522 | 0.230981 |
| 348 | -0.36442 | 0.127331 | -0.35193 | -0.63266 | 0.028479 | -0.14999 | -0.21074 | -0.36648 | -0.36579 | -0.17539 | -0.29533 | -0.23296 | -0.92581 | -0.20306 | -0.23879 | -0.20788 | -0.06236 | 0.177017 | -0.33787 | 0.111903 | -0.77881 | 0.220205 | -0.30561 | -0.83214 | -0.28677 | -0.12595 | 0.074918 | 0.056423 | -0.37177 | 0.282559 | 0.578759 | 0.317117 | -0.66578 | 0.690868 | -0.28606 | -0.13236 | 0.196464 | -0.05628 | -0.08545 | 0.258997 | 0.236875 | -0.89378 | -0.50557 | -0.50664 | -1.00146 | -0.14076 | -0.37896 | -0.77405 | -0.89967 | -0.51804 | -0.54781 | 0.012522 | 0.692843 |
| 349 | 0.154176 | 0.127331 | -0.15323 | -0.52827 | 0.159885 | -0.14999 | -0.56254 | -0.3564 | -0.41946 | -0.31819 | -0.13355 | -0.1705 | -0.58356 | 0.11006 | -0.18404 | -0.12198 | -0.26297 | 0.076186 | -0.20442 | -0.10121 | -0.71223 | 0.319727 | -0.06326 | -0.77994 | -0.12046 | -0.26304 | -0.43017 | 0.056423 | -0.89825 | 0.00481 | 0.600772 | 0.29488 | -0.57771 | 0.188901 | -0.04602 | -0.43949 | 0.196464 | -0.5352 | -0.31111 | 0.249023 | 0.112204 | -1.17204 | -0.46106 | -0.11717 | -1.32977 | -0.36544 | -0.45374 | -0.64879 | -0.21372 | 0.098064 | -0.9442 | -0.262 | 0.69 |
| 350 | -0.08769 | -0.03068 | -0.1911 | -0.43768 | 0.631087 | -0.37525 | -0.63154 | -0.24074 | -0.5457 | -0.214 | -0.11571 | -0.21278 | -1.10761 | -0.08386 | -0.6069 | -0.56825 | -0.54597 | -0.21212 | 0.112281 | -0.08285 | -0.71223 | 0.257549 | 0.038676 | -0.72687 | -0.02665 | -0.05961 | -0.18095 | -0.27326 | -0.98785 | 0.00481 | 0.476481 | 0.023598 | -0.72011 | -0.04628 | 0.114294 | -0.805 | -0.08136 | -0.56698 | -0.58425 | 0.152338 | 0.112243 | -0.96845 | -0.58687 | -0.15501 | -1.25154 | -0.48868 | -0.44332 | -0.47914 | 0.276839 | 0.133588 | -0.1582 | -0.40504 | 0.806185 |
| 351 | -0.12983 | -0.28159 | 0.030278 | 0.499179 | 0.695523 | -0.49459 | -0.52956 | -0.37863 | -0.45151 | -0.32823 | -0.36962 | 0.247728 | -0.6429 | -0.15309 | -0.47591 | -0.37064 | -0.6823 | -0.31496 | 0.293621 | -0.39751 | -0.67617 | 0.115701 | 0.132641 | 0.335207 | -0.09259 | -0.4269 | -0.39267 | -0.27326 | -0.94638 | 0.424183 | 0.282631 | -0.20765 | -0.37234 | 0.109162 | -0.10223 | -0.96185 | -0.05963 | -0.52613 | -0.3241 | 0.217154 | 0.060545 | -0.93122 | -0.42351 | 0.777754 | -0.87581 | -0.76081 | -0.77741 | -0.01538 | 0.491234 | 0.250241 | 0.722543 | -0.82682 | 0.914729 |
| 352 | -0.16913 | -0.62957 | -0.00083 | 0.687037 | 0.695523 | -0.40178 | -0.91144 | -0.18193 | -0.29601 | -0.26528 | -0.37864 | -0.01745 | -0.30326 | -0.4934 | -0.70968 | 0.034042 | -0.83809 | -0.2379 | 0.364634 | -0.06197 | -0.50856 | -0.35946 | 0.126144 | 0.503075 | -0.18736 | -0.54751 | -0.97487 | -0.26042 | -0.86263 | 0.420763 | 0.036504 | -0.25604 | -0.28081 | -0.21207 | -0.60754 | -0.82084 | -0.44396 | -0.48227 | -0.58915 | 0.375658 | 0.00961 | -0.86986 | -0.11325 | 0.539263 | -0.62182 | -0.50666 | -0.66941 | 0.020192 | 0.578317 | 0.631822 | 0.936378 | -0.80815 | 0.737039 |
| 353 | -0.3354 | -0.60176 | -0.00205 | 0.678531 | 0.436258 | -0.08026 | -0.82458 | -0.07691 | -0.3878 | -0.5045 | -0.30448 | -0.2543 | -0.28139 | -0.8764 | -0.93051 | -0.28875 | -0.83809 | -0.27996 | -0.23343 | -0.38759 | -0.49382 | -0.38401 | 0.022724 | 0.209905 | 0.062477 | -0.32423 | -0.82591 | -0.26042 | -0.86263 | 0.171454 | 0.036504 | -0.54677 | -0.10353 | -0.41874 | -0.58265 | -0.85525 | -0.73778 | -0.4589 | -0.95208 | 0.093577 | -0.54362 | -0.88472 | -0.08754 | 0.531293 | -0.78751 | -0.56726 | -0.76672 | 0.011458 | 0.811409 | 0.718743 | 0.691876 | -1.03823 | 0.466402 |
| 354 | -0.56083 | -0.83282 | -0.20075 | 0.379387 | 0.433886 | -0.08026 | -0.67851 | -0.08699 | -0.61048 | -0.36948 | -0.07338 | -0.4262 | -0.22373 | -0.89428 | -0.74493 | -0.48641 | -0.93511 | -0.07971 | -0.70229 | -0.47566 | -0.45222 | -0.30036 | 0.042582 | 0.191188 | -0.23201 | -0.40477 | -1.3765 | -0.26042 | -0.35354 | 0.323396 | -0.14878 | -0.25179 | -0.1916 | -0.64971 | -0.57486 | -0.54812 | -0.83669 | -0.15321 | -0.86823 | 0.103551 | -0.67021 | 0.008181 | -0.15286 | -0.05073 | -0.4597 | -0.20262 | -0.84517 | -0.23948 | -0.17196 | 0.098167 | 0.814443 | -0.76371 | -0.24135 |
| 355 | -0.58985 | -0.67481 | 0.301944 | 0.565777 | 0.167607 | -0.42527 | -0.73474 | -0.26505 | -0.685 | -0.24136 | 0.125732 | -0.49685 | 0.375147 | -0.69642 | -0.63744 | -0.25212 | -0.45309 | -0.03843 | -0.96112 | -0.22432 | -0.45222 | 0.026711 | -0.18844 | 0.222236 | -0.3665 | -0.40477 | -1.44197 | -0.22684 | -0.26394 | 0.079831 | -0.05889 | 0.016576 | -0.03534 | -0.2827 | -1.00097 | -0.37818 | -0.53283 | 0.116752 | -0.7986 | -0.06998 | -0.42336 | -0.28967 | -0.66629 | -0.16245 | -0.3235 | -0.15369 | -1.04293 | -0.53142 | -0.29356 | 0.727836 | -0.26352 | -0.44065 | -0.86595 |
| 356 | -0.88132 | -0.30344 | 0.253465 | -0.53992 | -0.20274 | -0.29982 | -0.94831 | -0.55118 | -0.89791 | -0.17787 | 0.373822 | -0.54892 | -0.09652 | -0.75751 | -0.62524 | -0.6002 | -0.51639 | 0.206003 | -1.13992 | -0.43857 | -0.50784 | 1.199095 | -0.01894 | -0.73945 | -0.71148 | -0.44659 | -1.24527 | -0.47086 | 0.058321 | -0.09162 | -0.15943 | -0.01007 | -0.18214 | -0.43044 | -0.88439 | -0.23235 | -0.53616 | -0.18385 | -1.11541 | -0.13479 | -0.60873 | -0.02727 | -0.66107 | -0.60234 | -0.67173 | 0.052575 | -0.55479 | -1.06934 | -0.3311 | 0.342803 | -0.6588 | -0.53609 | -0.92245 |
| 357 | -0.82441 | -0.38916 | 0.162602 | -0.87788 | 0.059886 | -0.07759 | -1.00732 | -0.75207 | -0.98483 | 0.709893 | 0.377203 | -0.13471 | -0.17902 | -0.66218 | -0.39941 | -0.99629 | -0.10631 | -0.28745 | -1.20284 | -0.39484 | -0.82107 | 1.45328 | 0.049658 | -0.79907 | -0.7152 | -0.49332 | -1.01397 | -0.31345 | 0.085062 | 0.030709 | -0.05214 | 0.482498 | 0.607602 | -0.05274 | -0.92776 | -0.3193 | -0.03683 | -0.30717 | -0.67061 | -0.49867 | -0.8134 | 0.110207 | -0.63011 | -0.57466 | -0.38282 | 0.214466 | -0.46944 | -0.91909 | -0.26855 | 0.069577 | -0.83303 | -0.56084 | -1.50654 |
| 358 | -1.07398 | -0.41538 | 0.284345 | -0.89317 | -0.06109 | -0.20767 | -0.97719 | -0.50411 | -0.70701 | 0.930669 | 0.069974 | 0.011304 | -0.16336 | -0.39956 | -0.18371 | -0.5789 | -0.28218 | -0.10207 | -1.0997 | -0.22 | -0.82852 | 1.252671 | 0.13233 | -0.6153 | -0.56931 | -0.46399 | -0.70366 | 0.267215 | -0.0495 | -0.0034 | -0.42178 | 0.536077 | 0.373003 | -0.43652 | -0.90162 | -0.31152 | -0.30608 | -0.2818 | -0.58879 | -0.38197 | -0.49212 | 0.021583 | -0.29448 | -0.33107 | -0.33971 | 0.403112 | -0.57822 | -0.5394 | -0.29309 | -0.36146 | -0.65253 | -0.46797 | -1.87157 |
| 359 | -1.07398 | -0.39673 | 0.062106 | -0.8528 | -0.43622 | -0.36802 | -0.75708 | -0.63537 | -0.68195 | 0.561081 | -0.30124 | -0.16241 | -0.55477 | -0.39956 | -0.18371 | -0.77558 | -0.2969 | -0.19013 | -1.03345 | -0.20223 | -0.72435 | 1.619921 | 0.298635 | -0.74588 | -0.69417 | -0.26916 | -0.12807 | 0.010244 | -0.27864 | -0.09093 | -0.48445 | 0.200293 | 0.354574 | -0.42149 | -0.95786 | -0.35795 | -0.19001 | -0.23004 | -0.44698 | -0.38197 | -0.27986 | -0.88759 | -0.41089 | -0.17174 | -0.05244 | -0.01193 | -0.20339 | -0.3759 | -0.30939 | -0.13502 | -0.40736 | -0.46797 | -1.25827 |
| 360 | -0.6775 | -0.35068 | -0.00924 | -0.89671 | -0.46816 | 0.152933 | -0.46142 | -0.38022 | -0.6578 | 0.322894 | -0.44809 | 0.147177 | -0.95175 | -0.58919 | 0.118504 | -0.87261 | -0.43107 | -0.28347 | -1.03781 | -0.0887 | -1.01387 | 1.432042 | 0.456431 | -0.45362 | -0.7449 | -0.13759 | -0.14674 | 0.123595 | -0.50977 | 0.292568 | -0.54102 | 0.089255 | 0.295688 | -0.49258 | -0.70318 | -0.50237 | -0.21604 | -0.75913 | -0.49213 | -0.23634 | -0.41953 | -0.72292 | 0.0523 | -0.03378 | -0.23059 | -0.05612 | 0.173409 | 0.034965 | -0.25052 | -0.71381 | -0.1154 | -0.61863 | -0.86602 |
| 361 | -0.70035 | -0.20594 | -0.40394 | -0.52207 | 0.162578 | -0.05882 | -0.21665 | -0.18417 | -0.27249 | 0.14036 | -0.4473 | 0.026384 | -0.9321 | -0.78051 | -0.252 | -0.87989 | -0.24684 | -0.33968 | -0.92574 | 0.271969 | -1.14956 | 0.732511 | -0.01941 | 0.218505 | -0.16579 | 0.050127 | -0.20587 | 0.336661 | -0.81008 | 0.217445 | -0.36673 | 0.149415 | 0.403954 | -0.59226 | -0.51365 | -0.66465 | -0.22416 | -0.57001 | -0.2943 | -0.09918 | -0.43735 | -0.86201 | -0.37244 | -0.21669 | -0.19791 | 0.122165 | 0.056819 | -0.28358 | -0.23746 | -0.42318 | -0.11675 | -0.18612 | -0.46294 |
| 362 | -0.60899 | 0.096625 | 0.574779 | -0.20229 | -0.28941 | -0.15247 | 0.245366 | 0.11786 | -0.38549 | -0.66401 | -0.37236 | -0.32508 | -1.05495 | -0.96255 | -0.47007 | -0.55138 | -0.42842 | 0.115664 | -0.91049 | 0.085458 | -0.63348 | 0.592239 | 0.830341 | 0.311997 | 0.14858 | 0.479661 | -0.42949 | -0.08093 | -0.949 | -0.07198 | -0.45046 | -0.48384 | -0.28932 | -0.92689 | -0.13369 | -0.4948 | -0.52231 | -1.0111 | -0.65668 | 0.273182 | -0.11323 | -0.90165 | -0.18937 | -0.22262 | -0.73888 | -0.09252 | 0.124263 | 0.009673 | -0.03097 | -0.29362 | 0.02579 | 0.222837 | 0.643487 |
| 363 | -0.56418 | 0.050141 | 0.692122 | -0.46297 | -0.10613 | 0.034706 | -0.07555 | -0.14067 | -0.61102 | -0.55916 | -0.1024 | -0.2211 | -1.27029 | -0.96241 | -0.43879 | -0.29285 | -0.17883 | -0.22178 | -1.01536 | 0.155896 | -0.56571 | 0.533392 | 0.81078 | 0.179673 | 0.129183 | 0.199426 | -0.26713 | -0.70028 | -0.90083 | 0.306563 | -0.05674 | -0.60936 | -0.05827 | -0.47237 | -0.34557 | -0.2874 | -0.34531 | -1.24658 | -0.6475 | -0.00524 | -0.1856 | -0.40182 | -0.24579 | -0.35702 | -0.17359 | -0.48152 | 0.197289 | -0.08777 | -0.04323 | 1.561437 | -0.02125 | 0.108959 | 0.705571 |
| 364 | -0.49285 | 0.13768 | 1.339758 | -0.28818 | 0.32264 | 0.21076 | -0.01089 | -0.0017 | -0.36342 | -0.39673 | -0.11069 | -0.04404 | -0.81001 | -1.06181 | -0.06174 | 0.274883 | -0.26589 | -0.324 | -0.31768 | 0.232353 | -0.68431 | 0.136099 | 0.935006 | 0.441728 | 0.093541 | 0.271473 | -0.17824 | -0.3031 | -0.794 | -0.01927 | 0.137853 | -0.63653 | -0.17817 | -0.27141 | -0.25941 | 0.292538 | -0.52012 | -1.20044 | -0.51307 | 0.148627 | 0.122831 | -0.1921 | 0.202063 | -0.36909 | -0.38898 | -0.23124 | -0.40528 | -0.1296 | -0.25939 | 1.351353 | 0.000544 | 0.006918 | 1.448482 |
| 365 | -0.25749 | -0.11306 | 1.163785 | -0.59609 | 0.487797 | 0.381247 | -0.61224 | -0.10312 | 0.061498 | -0.48592 | -0.33884 | -0.3736 | -0.60433 | -0.8514 | -0.22228 | 0.066889 | -0.56133 | -0.10298 | 0.162539 | -0.09127 | 0.041775 | -0.35179 | 0.959276 | 0.156784 | 0.255818 | 0.551603 | -0.33866 | -0.28584 | -0.55583 | 0.118662 | 0.006667 | -0.55166 | -0.206 | -0.61125 | -0.12919 | 0.224781 | -0.54538 | -1.02959 | -0.29458 | 2.349908 | -0.03237 | -0.34206 | 0.386904 | 1.905393 | 1.568366 | 0.566833 | -0.64958 | -0.57087 | -0.47614 | 1.28242 | -0.40801 | -0.17222 | 1.47482 |
| 366 | -0.3571 | -0.41165 | 1.478421 | -1.18892 | -0.0973 | 0.663918 | -0.76104 | -0.41983 | -0.27376 | -0.47952 | -0.2852 | -0.56814 | -0.6852 | -0.54691 | 0.250513 | 0.130351 | -0.58971 | 0.128728 | 0.962247 | -0.54039 | 0.112654 | -0.77734 | 1.618402 | -0.38712 | -0.01776 | 0.715184 | -0.22022 | -0.27433 | -0.35186 | 0.493053 | 0.375584 | -0.53819 | -0.27901 | -0.57336 | 0.07775 | 0.324296 | -0.517 | -0.85865 | 0.336467 | 2.936408 | -0.03725 | -0.40122 | 0.568373 | 2.692426 | 1.949019 | 1.095978 | -0.49185 | -0.16385 | -1.05656 | 1.244768 | -0.75253 | -0.48415 | 1.066917 |
| 367 | -0.39719 | -0.63264 | 0.699449 | -1.31931 | -0.12915 | 0.50427 | -1.12919 | -0.28165 | 0.180238 | -0.27451 | -0.20589 | -0.71992 | -1.17059 | -0.31464 | 0.549403 | -0.45953 | -0.74656 | -0.08264 | 1.588452 | -0.83936 | -0.32874 | -0.70732 | 1.16248 | -0.50087 | -0.07367 | 0.17335 | 0.055155 | -0.033 | 0.184898 | 1.28646 | 0.61762 | -0.28101 | -0.50642 | -0.59885 | 0.170509 | 0.150308 | -0.8681 | -0.74358 | 0.729338 | 2.769679 | -0.07311 | -0.4911 | 0.3327 | 3.049786 | 2.064346 | 1.114808 | -0.63818 | -0.25759 | -1.47328 | 0.833181 | -0.84035 | -0.82744 | 0.351236 |
| 368 | -0.37041 | -0.06625 | 0.408689 | -1.24369 | -0.48 | 0.44717 | -0.71317 | 0.164686 | -0.02201 | -0.24343 | -0.44933 | -0.88438 | -0.5657 | -0.56896 | 0.659891 | -0.82934 | -1.11862 | -0.38383 | 2.110209 | -0.6305 | -0.16913 | -0.16862 | 1.0367 | -0.25914 | -0.48272 | 0.193302 | 0.027181 | 0.664234 | -0.12855 | 1.35729 | 0.323044 | -0.14292 | -0.47761 | -0.91789 | 0.236883 | 0.077887 | -0.49968 | -0.40419 | 0.63564 | 3.002183 | 0.247826 | -0.43688 | 0.012681 | 3.12986 | 1.805192 | 1.240969 | -0.47104 | -0.57889 | -1.7252 | -0.94964 | -1.01962 | -0.78027 | 0.587008 |
| 369 | -0.31337 | -0.01061 | 0.053676 | -1.26409 | -0.55306 | 0.457015 | -0.87685 | 0.088608 | 0.269601 | -0.1291 | -0.69218 | -1.02009 | -0.82568 | -0.58056 | 0.216899 | -1.39856 | -0.91597 | -0.54511 | 1.704635 | -0.31415 | -0.21891 | 0.298349 | 0.629813 | -0.33458 | 0.126543 | -0.10972 | 0.148446 | 0.24158 | -0.34823 | 1.887497 | 0.058419 | -0.52132 | -0.50348 | -0.90389 | 0.24278 | -0.92838 | -0.35826 | -0.47455 | 0.431119 | 2.816928 | 0.279544 | -0.51559 | -0.42971 | 2.735757 | 1.78174 | 0.744964 | 0.013365 | -0.51221 | -1.08597 | -0.5462 | -1.40458 | -0.38253 | -0.05861 |
| 370 | -0.95131 | -0.06789 | -0.21598 | -1.24332 | -0.63389 | 0.46167 | -0.52888 | 0.076227 | 0.338648 | 0.198279 | -0.34685 | -0.98065 | -0.97585 | -0.85328 | 0.113635 | -1.39856 | -0.68538 | -0.41376 | 1.363344 | -0.38712 | -0.93883 | 0.663875 | 0.585458 | -0.211 | 0.490901 | -0.52141 | 0.388517 | 0.337802 | -0.47012 | 1.482288 | 0.113348 | -0.59259 | -0.3739 | -0.58187 | 0.09005 | -0.86072 | -0.37311 | -0.53862 | 0.482266 | 0.746911 | 0.471498 | -0.26895 | -0.79211 | 1.273299 | -0.02079 | -0.22888 | 0.049261 | -0.44755 | -1.20384 | -0.45124 | -0.94063 | -0.3149 | 0.031217 |
| 371 | -0.67624 | -0.2948 | -0.45059 | -0.98674 | -0.49531 | -0.10762 | -0.40165 | 0.495942 | 1.063909 | 0.228074 | -0.63299 | -0.63942 | -1.11919 | -0.80826 | 0.011343 | -1.58101 | -0.54588 | -0.88808 | 0.462282 | 0.053131 | -0.98913 | 0.498274 | 0.246129 | -0.15935 | 0.368901 | -0.73399 | 0.452322 | 0.269423 | 0.030758 | 1.532848 | -0.57793 | -0.34065 | -0.45763 | -0.35018 | -0.1157 | -0.57306 | -0.45556 | -0.65836 | 0.019802 | -0.08628 | 0.922327 | -0.07421 | -0.78649 | 0.313737 | -0.1107 | -1.15379 | 0.065066 | -0.59428 | -0.70786 | -0.32203 | -0.76872 | -0.41033 | -0.08575 |
| 372 | -0.82531 | -0.29084 | -0.53994 | -0.83818 | -0.2741 | -0.26204 | -0.13643 | 0.239137 | 0.63714 | 0.152627 | -1.13465 | -0.49431 | -0.96718 | -1.01744 | -0.3371 | -1.07565 | -0.58331 | -1.07835 | 0.039402 | 0.385154 | -0.99945 | 0.284658 | -0.37324 | -0.09888 | 0.127527 | -0.41634 | 0.155038 | 0.067408 | -0.66464 | 0.906532 | -0.84538 | -0.51399 | -0.16426 | -0.40534 | -0.30124 | -0.50088 | -0.27723 | -0.60819 | -0.27853 | -0.27328 | 1.08601 | 0.114357 | -1.0855 | 0.094293 | -0.12904 | -1.25198 | 0.038561 | -0.91913 | -0.85971 | -0.06074 | -0.79002 | -0.57222 | -0.18074 |
| 373 | -0.5645 | -0.38816 | -0.418 | -0.72385 | -0.00281 | -0.65822 | -0.29804 | -0.2019 | 0.922793 | -0.2972 | -1.05418 | -0.50093 | -1.30099 | -0.6321 | -0.60699 | -0.89102 | 0.110174 | -0.99362 | -0.56682 | -0.08085 | -1.08826 | 0.106746 | -0.13757 | -0.45679 | 0.064348 | -0.34484 | 0.450047 | -0.6752 | -0.22466 | 0.171182 | -0.64366 | -0.80028 | -0.15361 | -0.18484 | -0.43981 | -0.52459 | -0.42262 | -0.82346 | -0.04997 | -0.54995 | 0.897505 | -0.04859 | -0.72147 | 0.002386 | -0.11982 | -1.17013 | -0.00809 | -0.57402 | -0.53784 | 0.123178 | -0.37552 | -0.39599 | -0.50637 |
| 374 | -0.75718 | -0.4532 | -0.615 | -0.944 | -0.03458 | -0.83194 | -0.11358 | -0.4465 | 0.319134 | -0.62264 | -1.05881 | -0.17651 | -1.29414 | -0.61859 | -0.54054 | -0.4882 | 0.22985 | -0.8162 | -0.58906 | -0.17655 | -0.86939 | -0.37582 | -0.33098 | -0.32996 | -0.5632 | -0.04359 | -0.08482 | -0.66711 | 0.134717 | -0.23951 | -0.21443 | -0.62883 | -0.23348 | -0.35114 | -0.62048 | -0.01328 | -0.53195 | -0.98369 | -0.18964 | -0.58265 | 0.690493 | -0.12279 | -0.71777 | 0.170791 | -0.23534 | -0.93888 | -0.11699 | -0.67789 | -0.82547 | -0.08116 | 0.083271 | -1.4049 | -0.61734 |
| 375 | -0.65724 | -0.19122 | -0.13256 | -0.79422 | -0.09071 | -1.33819 | 0.21654 | -0.63109 | 0.189837 | -0.82912 | -1.3578 | -0.09196 | -0.96391 | -0.41142 | -0.58859 | -0.61865 | 0.336281 | -0.91279 | -0.50002 | -0.0238 | -0.58603 | -0.34687 | -0.05044 | -0.47636 | -0.9687 | 0.042063 | -0.17226 | -0.93787 | 0.223409 | -0.3435 | -0.08946 | -0.4689 | -0.31945 | -0.54625 | -0.65919 | 0.098036 | -0.41422 | -0.82311 | -0.52158 | -0.62554 | 0.805484 | 0.010472 | -0.48848 | -0.93575 | -0.4495 | -0.77056 | -0.54745 | -0.70795 | -0.63622 | 0.074861 | -0.26092 | -1.57818 | -0.71753 |
| 376 | -0.65944 | 0.054935 | -0.04968 | -0.62121 | 0.379148 | -1.17729 | 0.208947 | -0.92617 | -0.42851 | -0.76165 | -1.39822 | -0.41394 | -0.76658 | -0.68709 | -0.58053 | -0.60395 | -0.06281 | -0.47695 | -0.62929 | -0.22645 | -0.9771 | 0.174548 | -0.08792 | -0.56243 | -0.85454 | 0.132302 | -0.52549 | -0.85005 | -0.22875 | -0.54832 | 0.064476 | -0.82274 | -0.2374 | -0.86726 | -0.87545 | -0.30632 | -0.63623 | -1.03919 | -0.64811 | -0.60218 | 0.700377 | 0.351657 | -0.48848 | -0.90291 | -0.83132 | -0.58791 | -0.84827 | -0.32534 | -0.38389 | -0.30156 | -0.31967 | -1.39181 | -0.70943 |
| 377 | 0.446403 | 0.055134 | 0.189489 | -0.80508 | 0.198338 | -0.99181 | -0.11585 | -0.68975 | -0.48634 | -0.77087 | -1.24263 | -0.12502 | -0.44972 | -0.46066 | -0.63273 | -0.54955 | -0.07714 | -0.48652 | -0.64266 | -0.11701 | 0.16303 | 0.105266 | 0.302148 | -0.86299 | -0.90925 | 0.110375 | -0.37146 | -0.61469 | 0.248018 | -0.38897 | -0.20924 | -0.53411 | -0.43228 | -0.64149 | -0.68243 | -0.23163 | -0.47083 | -1.07325 | -0.87707 | -0.38982 | 0.34041 | 0.339165 | -0.37185 | -0.79334 | -0.53033 | -0.79907 | -1.04959 | -0.16328 | -0.25645 | -0.34399 | -0.2145 | -1.14794 | -0.61588 |
| 378 | 0.765171 | 0.353048 | -0.1807 | -0.97368 | 0.246385 | -0.53945 | -0.18516 | -0.56945 | -0.79108 | -0.65789 | -1.15011 | -0.27609 | -0.21993 | -0.51996 | -0.72974 | -0.79777 | -0.33052 | -0.46216 | -0.3679 | 0.141569 | 0.462387 | 0.003927 | 0.065484 | -0.39316 | -0.64674 | 0.138827 | -0.57811 | -0.73988 | -0.02577 | -0.11566 | -0.08756 | -0.27776 | -0.13651 | -0.55098 | -0.45241 | -0.22671 | -0.25675 | -0.90265 | -0.96122 | -0.20886 | 0.159434 | 0.362609 | -0.60731 | -0.60298 | -0.37394 | -0.80702 | -1.17316 | -0.07479 | -0.36405 | -0.57754 | -0.34953 | -1.20486 | -0.43489 |
| 379 | 1.15464 | 0.756944 | -0.33143 | -0.43571 | 0.538819 | -0.56792 | -0.30175 | -0.35231 | -0.58211 | -0.55576 | -1.04775 | -0.48359 | -0.27982 | -0.26376 | -0.76564 | -1.0099 | -0.06032 | -0.07804 | -0.43097 | 0.730149 | 0.549136 | 0.318171 | 0.059441 | -0.31882 | -0.08383 | -0.20967 | -0.66083 | -0.53276 | -0.15147 | 0.175261 | -0.54858 | -0.02107 | -0.13946 | -0.17467 | -0.65856 | -0.38174 | [illegible] | [illegible] | -1.0991 | 0.696174 | 0.044064 | 0.252566 | -0.34056 | -0.23582 | -0.29583 | -0.52608 | -1.18126 | 0.26916 | -0.39836 | -0.43854 | -0.22846 | -0.42365 | -0.26308 |
| 380 | 1.640129 | 0.565508 | -0.54671 | -0.46645 | 0.761166 | -0.50361 | -0.6905 | 0.030253 | -0.64379 | -0.19879 | -0.67605 | -0.29094 | -0.49086 | -0.12356 | -0.45308 | -0.84924 | -0.21862 | -0.04531 | -0.74874 | 0.880275 | 0.371133 | 0.196966 | -0.39742 | -0.4346 | 0.045883 | -0.45498 | -0.4098 | -0.53379 | -0.38906 | 0.230311 | -0.49583 | -0.11512 | -0.36149 | 0.01649 | -0.20955 | -0.14685 | [illegible] | [illegible] | -1.02334 | 0.693482 | -0.35566 | -0.03307 | 0.016506 | 0.057791 | 0.145076 | -0.49791 | -0.57289 | 0.569886 | -0.4112 | -0.43603 | 0.133252 | -0.68456 | -0.09925 |
| 381 | 1.525985 | 1.057223 | -0.52744 | -0.42369 | 0.138577 | -0.18078 | -0.61123 | 0.154384 | -0.54127 | -0.35705 | -0.81642 | 0.41609 | -0.54685 | 0.1095 | -0.46115 | -0.71402 | 0.242857 | -0.39943 | -0.10992 | 0.574932 | 0.75843 | 0.151322 | -0.60435 | -0.36349 | -0.05361 | -0.63365 | 0.131928 | -0.56842 | -0.61195 | 0.261562 | -0.68299 | 0.687436 | -0.28413 | 0.390911 | -0.16852 | -0.13408 | [illegible] | [illegible] | -0.48386 | 0.97842 | -0.60785 | -0.57284 | -0.14857 | 0.001693 | 0.542001 | -0.41469 | -0.23708 | 0.286935 | -0.6034 | -0.13376 | 0.106492 | -0.63941 | -0.02928 |
| 382 | 0.893502 | 1.146593 | -0.80234 | -0.26392 | 0.481866 | -0.70371 | -0.2772 | 0.198142 | -0.66326 | -0.18238 | -0.65544 | 0.078188 | -0.71566 | 0.391344 | 0.051923 | -0.81576 | 0.345887 | -0.44899 | -0.34841 | 0.483251 | -0.08314 | 0.456173 | -0.73478 | -0.01565 | 0.020896 | -1.06925 | 0.013906 | -0.6027 | -1.0114 | 0.276768 | -0.44284 | 0.31161 | -0.08926 | 1.100125 | -0.03284 | -0.48379 | [illegible] | [illegible] | -0.2269 | 1.019201 | -0.49597 | -0.37362 | -0.26921 | -0.14112 | 0.286165 | -0.10545 | -0.085 | 0.398081 | -0.36021 | -0.24789 | 0.053464 | -0.49868 | 0.029146 |
| 383 | 0.46989 | 0.737875 | -0.61464 | -0.10181 | 0.576533 | -0.72825 | -0.14281 | -0.13304 | -0.35047 | 0.18475 | -0.71064 | 0.058534 | -0.64716 | 0.963281 | 0.225264 | -0.491 | 0.277684 | -0.27645 | -0.50098 | 0.110238 | -0.56756 | 0.47936 | -1.13285 | -0.16586 | 0.030919 | -0.87595 | -0.09258 | -0.3269 | -0.82149 | 0.846164 | -0.61086 | 0.369135 | -0.49456 | 1.307684 | -0.08252 | -0.70696 | [illegible] | [illegible] | -0.36723 | 1.02514 | -0.38049 | -0.46336 | -0.05323 | -0.16155 | 0.083806 | 0.117474 | -0.06788 | 0.250617 | -0.18292 | 0.146681 | -0.05036 | -0.50633 | 0.048097 |
| 384 | 0.18027 | 0.707576 | -0.61305 | -0.47939 | 0.098576 | -0.73077 | -0.19861 | -0.25888 | -0.4484 | 0.330809 | -1.07607 | -0.14054 | -0.34832 | 0.548111 | 0.090702 | -0.54771 | -0.23552 | -0.16144 | -0.97299 | -0.28301 | -0.87482 | 0.388287 | -0.88389 | -0.46209 | -0.22178 | -0.57089 | 0.037929 | -0.21681 | -0.99005 | 0.610135 | -0.54258 | 0.347927 | 0.048462 | 1.097893 | 0.203764 | -0.87757 | [illegible] | [illegible] | -0.40008 | -0.01877 | -0.6105 | -0.38838 | -0.20491 | -0.32277 | -0.0656 | 0.396548 | -0.16435 | -0.30492 | -0.2693 | -0.08926 | -0.42656 | -0.62051 | -0.61138 |
| 385 | -0.0277 | 0.63383 | -0.60302 | -0.24549 | -0.17314 | -0.95796 | -0.09525 | -0.39466 | -0.68037 | -0.13218 | -1.26381 | -0.61862 | -0.15062 | 0.509772 | -0.2255 | -0.62438 | -0.49925 | -0.20092 | -0.72826 | -0.06445 | -0.88694 | 0.595042 | -0.56116 | -0.41081 | -0.39713 | -0.71625 | -0.32289 | 0.054981 | -1.02525 | 0.508541 | -0.35127 | 0.200779 | 0.202639 | 1.233462 | 0.35806 | -0.83274 | [illegible] | [illegible] | -0.38445 | 0.134071 | -0.42692 | 0.151924 | -0.36675 | -0.63356 | -0.32578 | 0.350976 | -0.3298 | -0.50304 | -0.44248 | -0.45988 | -0.41748 | -0.56311 | -0.57109 |
| 386 | 0.114807 | 0.300284 | -0.347 | -0.28268 | -0.01378 | -0.64026 | -0.50916 | -0.13404 | -0.69953 | -0.01368 | -0.59726 | -1.055 | 0.308661 | 0.672135 | -0.06239 | -0.63943 | -0.68748 | 0.095179 | -1.34294 | 0.150847 | -0.61342 | 0.745706 | -0.42019 | -0.56233 | -0.47321 | -0.52998 | -0.74924 | 0.18561 | -0.92951 | 0.258438 | 0.162871 | -0.52646 | 0.177168 | 1.162896 | 0.686725 | -0.79062 | [illegible] | [illegible] | -0.90055 | -0.09788 | -0.12549 | 0.195116 | -0.20494 | -0.75686 | -0.38408 | 0.442956 | -0.62859 | -0.41275 | -0.26642 | -0.59853 | -0.07364 | -0.3712 | 0.078301 |
| 387 | -0.20299 | 0.328362 | -0.39177 | -0.31776 | -0.28585 | -0.12257 | -0.45964 | -0.6289 | -0.52478 | -0.35147 | -0.56981 | -1.11509 | 0.262546 | 0.400247 | -0.25564 | -0.05087 | -0.44034 | 0.428503 | -0.94265 | -0.08721 | -0.77751 | 0.951384 | -0.66117 | -0.42771 | -0.24327 | -0.04444 | -0.73278 | 0.188918 | -1.05153 | 0.176358 | 0.048602 | -0.48123 | 0.290814 | 0.325265 | 0.165396 | -0.21179 | [illegible] | [illegible] | -1.06826 | -0.17045 | 0.044732 | -0.14394 | 0.092442 | -0.61076 | -0.39392 | 0.237965 | -0.49203 | -0.65082 | -0.37114 | -0.42911 | 0.245831 | -0.45172 | -0.08135 |
| 388 | -0.29352 | 0.182989 | -0.3413 | -0.25792 | -0.38738 | 0.132218 | -0.66272 | -0.37712 | -0.49866 | -0.49218 | 0.239569 | -0.77838 | 0.360897 | -0.23833 | -0.22072 | -0.16489 | -0.55626 | 0.259927 | -0.64498 | 0.278905 | -0.21521 | 1.10611 | -0.00157 | -0.32711 | -0.20565 | 0.238994 | -0.80989 | 0.139294 | -1.0006 | -0.49117 | 0.620294 | -0.89385 | 0.874144 | -0.09899 | -0.10565 | -0.10102 | [illegible] | [illegible] | -1.26294 | -0.12918 | -0.13187 | -0.17609 | 0.155203 | -0.64912 | -0.25735 | -0.20582 | -0.71274 | -0.65082 | -0.47447 | -0.64878 | 0.362772 | -0.32991 | -0.28666 |
| 389 | -0.33122 | 0.101273 | -0.06555 | 0.020182 | -0.27969 | 0.160935 | -0.68698 | -0.47837 | -0.01505 | -0.5498 | 1.052573 | -0.59677 | 0.117398 | 0.017351 | -0.33119 | -0.03534 | 0.003622 | -0.41324 | -0.1396 | -0.24203 | -0.03046 | 1.068739 | -0.07752 | -0.40404 | -0.28806 | -0.04418 | -1.03955 | 0.09645 | -1.00204 | -0.29673 | 1.254239 | -0.83219 | 0.59194 | -0.15773 | 0.045158 | -0.07934 | [illegible] | [illegible] | -0.81071 | 0.196909 | 0.119374 | 0.096701 | 0.142783 | -0.96369 | -0.17267 | -0.47628 | -0.60269 | -0.0246 | -0.29241 | -0.79146 | 0.740366 | -0.19438 | -0.00041 |
| 390 | -0.50023 | 0.151598 | -0.07242 | -0.29545 | -0.19202 | 0.462423 | -0.8198 | -0.78313 | 0.185279 | -0.2373 | 1.225234 | -0.31458 | 0.214704 | -0.05349 | -0.16548 | -0.06791 | 0.046628 | -0.00174 | 0.154144 | -0.71562 | 0.42665 | 0.79758 | -0.12986 | -0.46286 | -0.28836 | 0.291363 | -1.0561 | 0.196994 | -0.96278 | -0.01886 | 1.010227 | -0.65057 | 0.504109 | -0.33228 | -0.1478 | -0.25724 | [illegible] | [illegible] | -0.82773 | 0.154258 | 0.089267 | -0.53742 | -0.2289 | -0.92223 | -0.09448 | -0.23187 | -0.84726 | 0.160536 | -0.17288 | -0.71202 | 0.829049 | 0.459466 | -0.19166 |
| 391 | -0.12212 | 0.211873 | -0.43583 | 0.110429 | -0.37186 | 0.125113 | -0.80797 | -0.58041 | 0.133202 | -0.39082 | 0.985461 | -0.07426 | -0.22416 | 0.094906 | -0.62865 | -0.08393 | -0.1578 | -0.05162 | 0.667461 | -0.50673 | 0.319077 | 0.717062 | 0.012472 | -0.05873 | -0.26055 | 0.004894 | -1.1729 | -0.21365 | -1.0453 | -0.0102 | 0.560331 | -0.35638 | 0.330532 | -0.51017 | -0.53159 | -0.02759 | [illegible] | [illegible] | -1.02447 | 0.214086 | -0.16817 | -0.4547 | -0.4073 | -0.80415 | -0.05129 | -0.18862 | -0.6425 | -0.08222 | -0.50377 | -0.7736 | 0.593197 | 0.084091 | -0.59919 |
| 392 | -0.34022 | -0.25053 | -0.12016 | -0.19865 | -0.40409 | 0.130346 | -1.26402 | -0.12004 | 0.386335 | -0.30209 | 0.804507 | 0.049163 | -0.17619 | 0.096105 | -0.69146 | -0.62031 | -0.42144 | -0.08554 | 0.626578 | -0.50648 | 0.356727 | 0.23497 | 0.714751 | -0.41157 | -0.50305 | -0.14385 | -1.21063 | -0.32251 | -0.65529 | -0.25468 | 0.668195 | -0.16442 | 0.238216 | -0.54416 | -0.33895 | -0.51129 | [illegible] | [illegible] | -0.84858 | 0.46005 | -0.13355 | -0.30744 | -0.45165 | -1.00448 | 0.242831 | -0.04471 | -0.52515 | 0.194843 | -0.56129 | -0.95911 | 0.211863 | 0.031007 | -0.63429 |
| 393 | [illegible] |
| 394 | [illegible] |
| 395 | [illegible] |
| 396 | [illegible] |
| 397 | [illegible] |
| 398 | [illegible] |
| 399 | [illegible] |
| 400 | [illegible] |
| 401 | [illegible] |
| 402 | [illegible] |
| 403 | [illegible] |
| 404 | [illegible] |
| 405 | [illegible] |
| 406 | [illegible] |
| 407 | [illegible] |
| 408 | [illegible] |
| 409 | [illegible] |
| 410 | [illegible] |
| 411 | [illegible] |
| 412 | [illegible] |
| 413 | [illegible] |
| 414 | [illegible] |
| 415 | [illegible] |
| 416 | [illegible] |
| 417 | [illegible] |
| 418 | [illegible] |
| 419 | [illegible] |
| 420 | [illegible] |
| 421 | [illegible] |
| 422 | [illegible] |
| 423 | [illegible] |
| 424 | [illegible] |
| 425 | [illegible] |
| 426 | [illegible] |
| 427 | [illegible] |
| 428 | [illegible] |
| 429 | [illegible] |
| 430 | [illegible] |
| 431 | [illegible] |
| 432 | [illegible] |
| 433 | [illegible] |
| 434 | [illegible] |
| 435 | [illegible] |
| 436 | [illegible] |
| 437 | [illegible] |
| 438 | [illegible] |
| 439 | [illegible] |
| 440 | [illegible] |
| 441 | [illegible] |
| 442 | [illegible] |
| 443 | [illegible] |
| 444 | [illegible] |
| 445 | [illegible] |
| 446 | [illegible] |
| 447 | [illegible] |
| 448 | [illegible] |
| 449 | [illegible] |
| 450 | [illegible] |
| 451 | [illegible] |
| 452 | [illegible] |
| 453 | [illegible] |
| 454 | [illegible] |
| 455 | [illegible] |
| 456 | [illegible] |
| 457 | [illegible] |
| 458 | [illegible] |
| 459 | [illegible] |
| 460 | [illegible] |
| 461 | [illegible] |
| 462 | [illegible] |
| 463 | [illegible] |
| 464 | [illegible] |
| 465 | [illegible] |
| 466 | [illegible] |
| 467 | [illegible] |
| 468 | [illegible] |
| 469 | [illegible] |
| 470 | [illegible] |
| 471 | [illegible] |
| 472 | [illegible] |
| 473 | [illegible] |
| 474 | [illegible] |
| 475 | [illegible] |
| 476 | [illegible] |
| 477 | [illegible] |
| 478 | [illegible] |
| 479 | [illegible] |
| 480 | [illegible] |
| 481 | [illegible] |
| 482 | [illegible] |
| 483 | [illegible] |
| 484 | [illegible] |
| 485 | [illegible] |
| 486 | [illegible] |
| 487 | [illegible] |
| 488 | [illegible] |
| 489 | [illegible] |
| 490 | [illegible] |
| 491 | [illegible] |
| 492 | [illegible] |
| 493 | [illegible] |
| 494 | [illegible] |
| 495 | [illegible] |
| 496 | [illegible] |

| | | | | | | | | | | | | | | | | | | | | |
|---|---|---|---|---|---|---|---|---|---|---|---|---|---|---|---|---|---|---|---|---|
| 497 | -0.89712 | -0.63798 | -0.87471 | 0.096875 | -1.4398 | -0.22093 | 0.244044 | -0.74872 | -0.30809 | 0.027252 | -1.36684 | -0.62617 | -0.24437 | -0.69936 | -0.57587 | -0.24269 | -0.20919 | -0.3427 | 0.503561 | 0.92262 |
| 498 | -0.22195 | -1.27872 | 0.297113 | 0.136473 | -1.40557 | 0.122571 | -0.15563 | -0.38034 | -0.50513 | -0.36342 | -1.55572 | -0.46165 | -0.1429 | -0.50532 | -0.7979 | -0.25161 | 0.547099 | -0.38348 | -0.09109 | -0.35523 |
| 499 | -0.04507 | -1.22344 | 0.531272 | -0.00482 | -1.20771 | -0.01174 | -0.6509 | -0.50894 | -0.40887 | -0.53281 | -1.11364 | -0.10429 | -0.16015 | -0.24349 | -0.53703 | -0.38278 | 0.697802 | -0.19097 | -0.43037 | -0.84126 |
| 500 | -0.12491 | -0.69613 | 0.332545 | -0.39906 | -1.33669 | -0.05738 | -0.76852 | -0.34322 | -0.32067 | -0.60847 | -1.17619 | 0.088078 | -0.70206 | -0.47641 | -0.77011 | -0.17386 | 0.067798 | 0.002078 | -0.20752 | -0.86445 |
| 501 | -0.14414 | -0.53285 | 0.279874 | 0.055983 | -1.36326 | -0.0179 | -0.79951 | -0.49293 | -0.27828 | -0.6604 | -0.97759 | 0.066554 | -0.77696 | -0.4923 | -0.31786 | 0.046211 | -0.04139 | 0.316385 | -0.28832 | -1.06777 |
| 502 | 0.290866 | -0.32213 | 0.830863 | -0.6591 | -1.06521 | -0.25708 | -1.1183 | -0.78713 | -0.29593 | -0.63611 | 0.018394 | -0.21882 | -0.86058 | -0.45353 | -0.4446 | 0.053888 | -0.32928 | 0.221173 | -0.26225 | -1.13787 |
| 503 | -0.27958 | 0.501498 | 0.35655 | -0.56013 | -0.49981 | -0.28754 | -0.94589 | -0.76581 | -0.3178 | -0.65906 | -0.07882 | 0.24387 | -0.96897 | -0.49487 | -0.3091 | 0.222992 | -1.17649 | 0.467259 | 0.011852 | -1.04247 |
| 504 | -0.35113 | 0.535366 | 0.642432 | -0.72488 | -0.45218 | -0.25431 | -0.59133 | -0.60685 | -0.85509 | -0.48284 | -0.32626 | 0.048223 | -1.25906 | -0.8077 | 0.240647 | 0.301513 | -1.60171 | 0.236461 | -0.01729 | -0.84355 |
| 505 | -0.49 | 0.80893 | 0.894137 | -0.34881 | -0.21294 | -0.24803 | -0.3917 | -0.69496 | -1.00182 | -0.48311 | -0.40196 | -0.16455 | -0.84619 | -0.39934 | 0.381897 | -0.04387 | -1.15113 | -0.39449 | -0.6151 | -0.69874 |
| 506 | -0.46191 | 0.896119 | 1.40865 | -0.50725 | 0.541524 | -0.09842 | -0.53387 | -0.62835 | -1.01172 | -0.54709 | -0.51683 | 0.163295 | -0.92486 | -0.38677 | 0.633431 | 0.306455 | -0.79307 | -0.73978 | -0.38197 | -0.65781 |
| 507 | -0.41022 | 0.874092 | 1.811901 | -0.30019 | 1.68913 | -0.07524 | -0.43798 | -0.23237 | -1.01234 | -0.51656 | -1.21733 | 0.399572 | -0.90827 | -0.15927 | 0.747104 | 0.436359 | -0.41861 | -1.05813 | -0.40717 | -0.89002 |
| 508 | -0.3635 | 0.141826 | 1.431733 | -0.53222 | 1.850043 | -0.11665 | -0.36393 | -0.10471 | -1.09315 | -0.58059 | -0.8958 | -0.34085 | -0.54427 | 0.055772 | 0.737698 | 0.409229 | -0.43398 | -1.15465 | -0.27434 | -0.31145 |
| 509 | -0.43183 | -0.03413 | 1.122936 | -0.26642 | 1.709963 | -0.34914 | -0.40115 | -0.35601 | -0.4664 | -0.59094 | -0.82944 | -0.17091 | -0.5213 | 0.074022 | 0.04692 | 1.606748 | 0.006252 | -1.19915 | -0.14696 | -0.31512 |
| 510 | -0.01053 | -0.56855 | 1.224212 | 0.012408 | 1.29865 | -0.24619 | -0.33583 | -0.34905 | -0.46805 | -0.22405 | -0.84306 | 0.229598 | -0.33152 | 0.17208 | 0.284526 | 1.978185 | -0.0705 | -0.66662 | 0.074959 | -0.33787 |
| 511 | -0.11934 | -0.8788 | 0.672588 | -0.16326 | 0.674158 | -0.24386 | -0.13458 | -0.35344 | -0.58427 | -0.11627 | -0.81417 | -0.30169 | -0.23323 | 0.27411 | -0.01375 | 1.084831 | -0.15321 | -0.57187 | 0.020311 | -0.18121 |
| 512 | -0.1574 | -0.72756 | -0.06665 | 0.092982 | -0.13972 | 0.172711 | -0.31921 | -0.3093 | -0.52405 | -0.33306 | -0.40463 | -0.26409 | -0.23458 | 0.179268 | -0.27306 | 0.511331 | -0.30615 | -0.51967 | -0.40026 | 0.091462 |
| 513 | -0.27763 | -0.61268 | -0.39366 | 0.233755 | -0.47007 | 0.233005 | -0.37192 | -0.53593 | -0.15859 | -0.2117 | -0.30866 | 0.137599 | -0.56705 | -0.03233 | -0.36325 | 0.275493 | -0.08002 | -0.29896 | -0.82616 | -0.44235 |
| 514 | -0.04861 | -0.13009 | -0.59556 | 0.054626 | -0.20765 | 0.085876 | -0.78857 | -0.18577 | -0.51413 | 0.062153 | -0.24863 | 0.055564 | -0.44415 | -0.2616 | -0.52276 | -0.68589 | -0.45712 | -0.31516 | -0.9871 | -0.45994 |
| 515 | 0.087997 | 0.112284 | -0.20655 | 0.038654 | -0.03476 | 0.052461 | -0.48849 | -0.0478 | -0.31273 | 0.101638 | 0.232832 | 0.168757 | -0.35086 | -0.43296 | -0.79266 | -0.81968 | 0.106519 | -0.37195 | -0.72544 | -0.36201 |
| 516 | -0.029 | 0.34549 | -0.24024 | 0.014329 | 1.005072 | 0.176244 | -1.2144 | 0.032529 | -0.27951 | 0.131055 | 0.211583 | 1.299256 | -0.25771 | -0.26705 | -0.69634 | -0.49369 | -0.04173 | -0.30112 | -0.56429 | -0.46965 |
| 517 | 0.485342 | 0.571087 | 0.284453 | 0.100877 | 1.363933 | -0.15143 | -1.12029 | -0.14589 | -0.61455 | 0.345054 | -0.11796 | 1.815434 | -0.13071 | 0.374757 | -0.54063 | 0.156165 | -0.18992 | 0.181062 | 0.127597 | -0.39461 |
| 518 | 0.511642 | 1.059845 | 0.655558 | 0.100877 | 1.038874 | -0.43286 | -1.12019 | -0.10906 | -0.70235 | 0.539226 | -0.20176 | 1.968044 | 0.253625 | 0.899811 | 0.016385 | 0.852178 | -0.44198 | -0.20089 | 0.576875 | -0.39724 |
| 519 | -0.37579 | 0.739288 | 0.576338 | 0.190154 | 0.902554 | -0.13721 | -0.59905 | -0.25742 | -0.782 | 0.341403 | -0.0807 | 1.798504 | 0.472082 | 1.098126 | 0.441635 | 0.822563 | -0.29375 | 0.111598 | 0.776828 | -0.28592 |
| 520 | -0.32608 | 0.58882 | 0.052232 | -0.29026 | 0.986502 | 0.072561 | -1.35532 | -0.08864 | -1.1028 | 0.163445 | -0.25254 | 1.329324 | 0.339069 | 0.819913 | 0.579802 | 0.737463 | -0.24849 | 0.511843 | 0.680706 | -0.32367 |
| 521 | -0.08896 | 0.434232 | 0.122423 | 0.189128 | -0.01066 | 0.341732 | -0.89766 | -0.28806 | -1.15305 | 0.170325 | -0.36259 | 0.157956 | 0.30628 | 0.485227 | 0.568741 | 0.666611 | -0.13583 | 0.340193 | 0.513693 | -0.26859 |
| 522 | -0.42965 | 0.03671 | 0.116317 | -0.12289 | -0.37966 | 0.203187 | -0.55676 | -0.39765 | -0.83311 | 0.197749 | -0.0993 | -0.58932 | -0.08006 | -0.05824 | 0.42923 | 0.370699 | 0.16012 | -0.03126 | 0.127202 | -0.19965 |
| 523 | 0.100348 | 0.053255 | -0.02053 | -0.2961 | -0.28438 | 0.406553 | -0.42911 | -0.62449 | -0.61936 | 0.017974 | -0.33407 | -0.95261 | -0.29015 | -0.74541 | -0.31047 | -0.3626 | 0.027798 | -0.11666 | 0.054009 | 0.300797 |
| 524 | 0.581629 | -0.26999 | 0.435906 | -0.18256 | -0.7409 | 0.406553 | -0.6605 | -0.60941 | -0.69842 | 0.068782 | -0.36874 | -1.24268 | -0.30574 | -0.85752 | -0.5378 | -0.51341 | -0.03737 | -0.14407 | 0.364273 | 0.599261 |
| 525 | 0.046278 | -0.22298 | 0.298415 | -0.40061 | -1.02357 | 0.000402 | -0.34845 | -0.95214 | -0.61774 | 0.231367 | -0.32301 | -1.23207 | -0.39224 | -0.68739 | -0.82003 | -0.28888 | -0.68937 | -0.51942 | 0.435405 | 0.274105 |
| 526 | -0.25728 | -0.06839 | 0.154697 | -0.63215 | -0.80407 | -0.664 | -0.18938 | -0.65447 | -0.25528 | 0.278309 | -0.10575 | -1.0067 | -0.97438 | -0.59325 | -0.89566 | -0.12409 | -0.88096 | -0.39969 | 0.173562 | 0.082708 |
| 527 | -0.50161 | 0.021672 | -0.34676 | -1.33159 | -1.06533 | -0.44529 | -0.42412 | -0.64129 | -0.00224 | 0.423366 | -0.11473 | -0.91875 | -0.57173 | -0.52516 | -0.87567 | -0.91972 | -0.80102 | -0.70249 | -0.15171 | -0.0753 |
| 528 | -1.40629 | 0.237649 | -0.5965 | -1.26215 | -1.12861 | -0.67848 | -0.66874 | -0.6351 | -0.06678 | 0.445922 | 0.284067 | -0.89215 | -0.47979 | -0.79942 | -0.28518 | -0.38298 | -0.79722 | -0.11772 | -0.36357 | -0.59626 |
| 529 | -1.31031 | 0.505453 | -1.15398 | -1.47297 | -1.16365 | -1.20896 | -0.63044 | -0.65018 | 0.508624 | 0.204203 | 0.339701 | -0.33845 | -1.04126 | -0.75114 | -0.20311 | 0.026972 | -1.28694 | 0.055202 | -1.17409 | -0.10291 |
| 530 | -1.06724 | 0.451398 | -1.24091 | -1.23489 | -0.66414 | -1.21991 | -0.14269 | -0.4313 | -0.01724 | 0.184546 | -0.03888 | -0.57977 | -0.49275 | -1.04744 | 0.231999 | -0.52423 | -1.18978 | 0.001197 | -1.32328 | 0.243306 |
| 531 | -0.8575 | 0.54948 | -1.23263 | -1.11666 | -0.79674 | -1.06316 | -0.30916 | -0.62167 | -0.13046 | 0.137604 | -0.07426 | -0.51407 | 0.137932 | -1.0781 | 0.135758 | -0.68777 | -0.93942 | -0.01672 | -1.13194 | 0.565985 |
| 532 | -0.83042 | 0.528343 | -0.8965 | -0.24941 | -0.46784 | -1.56258 | -0.37506 | -0.41193 | -0.29443 | -0.0933 | -0.10524 | -0.42837 | 0.07064 | -1.00381 | 0.181635 | -0.0093 | -0.90346 | -0.17786 | -1.06202 | 0.926929 |
| 533 | -0.04991 | -0.1394 | -0.91323 | -0.41529 | -0.14492 | -0.95419 | 0.089352 | -0.0894 | -0.06683 | -0.13682 | 0.281843 | -0.30774 | -0.11698 | -0.65031 | -0.11136 | -0.52097 | -0.88877 | -0.67789 | -0.88249 | 0.898009 |
| 534 | -0.61097 | -0.04977 | -0.62814 | -0.36873 | 0.229821 | -0.77821 | 0.282446 | -0.27387 | 0.094105 | 0.021867 | 0.274527 | -0.66446 | 0.441425 | -0.709 | -0.40231 | -1.00204 | -0.36439 | -0.96084 | -0.59202 | 0.307156 |
| 535 | -0.85777 | 0.802723 | -0.40372 | -0.02011 | -0.04619 | -0.59132 | -0.41851 | -0.00827 | 0.582873 | -0.17686 | 0.509865 | -0.14555 | -0.07555 | -0.13692 | -0.63218 | -0.68173 | -0.5372 | -1.05072 | -0.33413 | 0.171721 |
| 536 | -1.02588 | 0.865015 | -0.29455 | 0.050257 | 0.024802 | -0.62303 | -0.35439 | -0.25522 | 0.615133 | -0.06522 | 0.750885 | 0.135693 | -0.71891 | 0.095857 | -0.25699 | -0.86192 | -0.38929 | -1.30134 | -0.23458 | -0.14721 |
| 537 | -1.22463 | 0.518429 | -0.52385 | -0.30296 | -0.04283 | -0.35501 | -0.27681 | -0.13669 | 0.389857 | 0.019021 | 1.073217 | 0.279207 | -0.71384 | -0.38719 | -0.39728 | -0.88686 | -0.67149 | -0.72563 | -0.07346 | -0.6133 |
| 538 | -1.55468 | 0.454965 | -0.30068 | 0.039412 | -0.33524 | -0.51386 | -0.49182 | -0.23147 | -0.06134 | 0.022007 | 0.410303 | 0.461135 | -0.6095 | -0.3518 | -0.54466 | -0.93594 | -0.63392 | -0.69488 | -0.25622 | -0.35006 |
| 539 | -1.14541 | -0.0814 | -0.30068 | 0.604064 | -0.08688 | -0.05719 | -0.64783 | 0.013916 | -0.49497 | -0.17505 | 0.10768 | 0.317442 | -0.7783 | -0.42445 | -0.33419 | -0.90661 | -0.6135 | -0.35824 | 0.001701 | -0.47975 |
| 540 | -1.1346 | -0.94247 | -0.39367 | 0.019561 | -0.41334 | -0.31151 | -0.79062 | -0.23512 | -0.36812 | 0.161239 | -0.12936 | 0.473128 | -0.90127 | -0.42705 | -0.23522 | -0.83282 | -0.49536 | -0.89621 | -0.11167 | -0.29065 |
| 541 | -0.94557 | -1.25605 | -0.05237 | -0.21821 | 0.019453 | -0.35341 | -0.62887 | 0.059391 | -0.3684 | 0.027815 | -0.91943 | 0.052836 | -0.3821 | -0.38481 | -0.45971 | -0.55516 | -1.04484 | -0.63565 | -0.09258 | -0.2722 |
| 542 | -1.03569 | -0.86033 | -0.00802 | -0.08636 | -0.02119 | -0.12936 | -0.73903 | -0.28742 | -0.59151 | -0.10948 | -0.66249 | 0.248863 | -0.54676 | -0.37496 | -0.18048 | -0.7468 | -0.84234 | -1.00102 | -0.03098 | -0.32611 |
| 543 | -0.9777 | -0.6176 | 0.12065 | -0.10379 | 0.138058 | -0.27655 | -0.88845 | -0.45994 | -0.51106 | 0.020625 | -0.40147 | -0.17369 | -0.61151 | -0.40276 | -0.28421 | -0.4067 | -0.62661 | -1.12595 | 0.148096 | -0.13621 |
| 544 | -0.76133 | -0.07936 | 0.12065 | -0.95326 | -0.24176 | -0.1518 | -0.97963 | -0.83855 | -0.43735 | 0.288553 | -0.43106 | -0.23942 | -0.56573 | -0.12413 | -0.1832 | 0.456506 | -0.47081 | -1.32516 | -0.13516 | -0.02911 |
| 545 | -0.82486 | -0.06373 | 0.213637 | -0.42193 | 0.541192 | 0.122965 | -0.96259 | -0.80291 | -0.44596 | -0.17849 | -0.16797 | -0.7037 | -0.69353 | -0.62255 | -0.20517 | 0.440775 | -0.23938 | -0.62587 | -0.40626 | -0.48871 |
| 546 | -0.94148 | 0.089475 | -0.00077 | -0.33381 | -0.1122 | 0.311273 | -1.23543 | -0.71961 | -0.83588 | -0.33181 | 0.333065 | -0.63408 | -0.56269 | -0.51038 | 0.03721 | 0.336864 | 0.30867 | -0.56597 | -0.4544 | -0.59258 |
| 547 | -0.88555 | -0.02859 | 0.08538 | -0.28025 | 0.04008 | -0.0238 | -0.99567 | -0.79288 | -0.61064 | -0.1929 | -0.53998 | -1.23007 | -0.51216 | -0.17956 | 0.258675 | 0.409386 | 0.207987 | -0.56726 | -0.73459 | -0.33238 |
| 548 | -0.87016 | -0.1708 | 0.229229 | -0.46156 | -0.01635 | -0.87729 | -1.10717 | -0.90166 | -0.84008 | -0.53347 | -0.86401 | -1.07054 | -0.5795 | -0.20171 | 0.594899 | 0.313535 | 0.326589 | 0.043071 | -0.88244 | -0.43615 |
| 549 | -0.90268 | -0.39438 | 0.494415 | -0.12603 | 0.231918 | -1.18572 | -0.77199 | -0.58397 | -0.77705 | -0.77367 | -0.54544 | -1.06086 | -0.43784 | -0.32633 | 0.594899 | -0.61542 | 0.180872 | -0.20969 | -0.93488 | -0.61452 |
| 550 | -0.65807 | -0.46554 | 0.570839 | -0.0834 | -0.21538 | -0.89526 | -0.26765 | -0.58058 | -0.96092 | -0.66549 | -0.85439 | -1.04639 | -0.16305 | -0.14508 | 0.196275 | -0.61542 | 0.077194 | -0.31817 | -0.92252 | -0.17399 |
| 551 | -0.4575 | -0.59223 | 0.560951 | -0.02543 | -0.26132 | -1.2373 | 0.446786 | -0.75162 | -0.82167 | -0.62335 | -0.7832 | -1.13302 | -0.29219 | -0.84679 | -0.03169 | -0.66136 | -0.43245 | -0.62776 | -1.28386 | -0.18402 |
| 552 | -0.33787 | -0.84248 | 0.560951 | -0.16293 | -0.62963 | -0.99128 | 0.642955 | -0.93616 | -0.99058 | -0.62335 | -0.78453 | -0.94187 | -0.50015 | -1.1349 | -0.31845 | -0.40013 | -0.45445 | -0.36984 | -0.91217 | -0.47295 |
| 553 | -0.02228 | -0.63561 | 0.331722 | -0.0369 | -0.87502 | -0.29613 | 0.597386 | -1.13304 | -0.99832 | -0.47722 | -0.8861 | -1.49502 | -0.72502 | -1.00547 | -0.54232 | -0.50048 | -0.78509 | -0.96164 | -1.08523 | -0.4671 |
| 554 | 0.054763 | -0.68663 | 0.066536 | -0.37749 | -1.1271 | -0.39127 | 0.175524 | -1.08546 | -0.9984 | -0.30593 | -0.63795 | -1.09429 | -0.75944 | -1.55536 | -0.51253 | -0.48459 | -0.98766 | -0.91201 | -1.00535 | -0.65261 |
| 555 | -0.17959 | -0.51925 | 0.042602 | -0.66917 | -1.80255 | -0.68172 | -0.31582 | -1.42757 | -0.79162 | -0.09517 | -0.24434 | -1.09885 | -1.22674 | -1.60355 | -0.31539 | -0.45304 | -0.85511 | -0.78657 | -1.05137 | -0.67994 |
| 556 | -0.48185 | -0.60532 | 0.008474 | -1.16554 | -2.14498 | -0.38826 | -0.67649 | -1.09085 | -0.76236 | 0.037771 | -0.65783 | -1.18153 | -1.17397 | -1.3741 | -0.48782 | -0.47736 | -0.61673 | -0.61281 | -0.87409 | -0.86729 |
| 557 | -0.35578 | -0.51192 | -0.01346 | -1.03719 | -1.98211 | -0.32171 | -0.91463 | -0.66379 | -0.36359 | -0.16642 | -0.57832 | -1.32387 | -0.86552 | -1.33951 | -0.66147 | -0.47462 | -0.73602 | -0.61582 | -1.13124 | -0.5179 |
| 558 | -0.62555 | -0.81188 | -0.04097 | -1.33037 | -1.69435 | 0.025322 | -0.58001 | -0.354 | -0.68904 | -0.13357 | -0.47668 | -0.28871 | -0.5108 | -1.57456 | -0.82701 | 0.105559 | -0.36683 | -0.36519 | -0.87747 | -0.18825 |
| 559 | -0.80141 | -0.86995 | -0.09046 | -1.25983 | -1.35752 | 0.128968 | -0.23384 | -0.5269 | -1.012 | -0.36979 | -0.92148 | -0.43333 | -0.27617 | -1.307 | -1.0614 | 0.09086 | -0.29225 | 0.121813 | -0.83846 | -0.11929 |
| 560 | -0.64106 | -1.22467 | -0.20449 | -1.32062 | -0.676 | 0.237814 | -0.10471 | -0.24725 | -0.70971 | -0.62334 | -1.39464 | -0.29182 | 0.135162 | -1.50658 | -1.19648 | -0.01251 | -0.27228 | 0.134934 | -0.62966 | -0.4719 |
| 561 | -0.60396 | -1.08974 | -0.07609 | -1.23026 | -0.05665 | 0.112487 | -0.23552 | -0.26027 | -0.71608 | -0.85052 | -1.05874 | 0.11426 | 0.839982 | -1.55807 | -1.19069 | 0.113594 | 0.013739 | 0.223126 | -0.46484 | 0.064014 |
| 562 | -0.44588 | -1.26636 | -0.2157 | -1.47455 | -0.34582 | -0.16208 | 0.142168 | -0.24796 | -1.09704 | -0.7547 | -1.00997 | 0.666457 | 0.882378 | -1.54602 | -1.40123 | 0.140432 | -0.22771 | -0.05393 | -0.10184 | 0.019689 |
| 563 | -0.52416 | -0.9194 | -0.28222 | -0.80424 | -0.48529 | -0.48555 | -0.04431 | 0.056551 | -0.58198 | -0.92055 | -0.75941 | 0.367808 | 0.577798 | -1.23035 | -1.28461 | -0.0068 | -0.18084 | 0.146299 | -0.16498 | -0.72726 |
| 564 | -0.54812 | -1.1245 | -0.08281 | -0.44103 | -0.72405 | -0.67124 | -0.36887 | -0.20857 | -0.49096 | -0.90452 | -0.74295 | 0.756943 | 0.233871 | -0.92704 | -1.01714 | 0.368066 | -0.09023 | 0.038481 | -0.18327 | -0.5537 |
| 565 | -0.45936 | -0.69919 | -0.15189 | -0.28526 | -0.74697 | -0.35257 | -0.20449 | -0.08078 | -0.68518 | -0.40283 | -0.61305 | 1.164144 | 0.356052 | -0.58793 | -0.6547 | 0.06061 | -0.06643 | 0.354014 | 0.138721 | -0.39496 |
| 566 | -0.00311 | -0.67416 | -0.23628 | 0.092142 | -0.75735 | 0.06297 | -0.30845 | -0.23345 | -0.81166 | 0.081892 | -0.83407 | 1.146421 | -0.03594 | -0.17841 | -0.58912 | 0.004771 | -0.31653 | 0.318589 | 0.170224 | -0.74994 |
| 567 | -0.40629 | -0.34076 | -0.33467 | 0.512039 | -0.13518 | 0.275675 | -0.71488 | -0.47119 | -0.6284 | 0.190259 | -0.69869 | 0.961756 | -0.47431 | -0.2116 | -0.12219 | -0.29228 | 0.130673 | 0.311429 | -0.10253 | -0.58348 |
| 568 | -0.27697 | -0.33315 | -0.28061 | 0.13491 | 0.018352 | 0.097187 | -0.65695 | -0.89492 | -0.3224 | 0.132796 | -0.91516 | 0.640328 | -0.28195 | -0.41637 | 0.08083 | -0.77701 | -0.01673 | 0.045976 | -0.28055 | -0.16618 |
| 569 | -0.12166 | 0.198274 | -0.43051 | 0.041753 | 0.130503 | 0.127719 | -0.4389 | -0.50449 | 0.045766 | 0.140927 | -0.73497 | 0.287919 | -0.13823 | -0.20037 | 0.017944 | -0.89473 | -0.2315 | -0.13008 | -0.09359 | -0.22319 |
| 570 | -0.12509 | -0.16448 | -0.50283 | 0.195826 | 0.197353 | -0.2998 | -0.25621 | -0.5732 | 0.020732 | -0.49213 | -0.78265 | -0.2633 | -0.17349 | -0.45607 | -0.00792 | -0.77273 | -0.50863 | -0.4757 | -0.31156 | -0.12574 |
| 571 | -0.27943 | -0.32169 | -0.50283 | -0.10964 | -0.06805 | -0.75275 | -0.11213 | -0.66658 | 0.300775 | -0.74966 | -0.57384 | -0.48233 | -0.40995 | -0.60642 | 0.126963 | -1.34442 | -0.47853 | -0.39263 | -0.32382 | 0.182596 |
| 572 | 0 | -0.32163 | -0.24291 | -0.55284 | -0.30725 | -0.75275 | -0.09245 | -0.02931 | 0.402689 | -0.71427 | -0.57837 | -1.25905 | -0.11447 | -0.33176 | 0.044227 | -1.35451 | -0.47853 | -0.10842 | -0.30957 | -0.06295 |
| 573 | 0.177646 | -0.12045 | -0.20294 | -0.55284 | -0.73101 | -0.76535 | -0.11294 | 0.280117 | -0.04444 | -0.71059 | -0.29122 | 0.476714 | 0 | -0.08023 | -0.18367 | -1.35365 | -0.45665 | -0.24326 | -0.15533 | -0.16389 |
| 574 | 0.171823 | -0.44405 | -0.20162 | -0.76507 | -0.37177 | -0.66129 | -0.06604 | 0.001723 | -0.20952 | -0.49569 | -0.62758 | 2.675825 | 0.008299 | -0.19331 | -0.35943 | -1.35365 | -0.13845 | -0.43858 | -0.4389 | -0.36983 |
| 575 | 0.171823 | -0.60963 | -0.01625 | -0.4249 | -0.57095 | -1.13529 | -0.41745 | -0.0794 | -0.32889 | 0.123629 | -0.39547 | 3.218506 | -0.09413 | -0.13212 | -0.50236 | -1.12501 | -0.08695 | -0.48306 | -0.08417 | -0.30573 |
| 576 | 0.220667 | -0.46545 | -0.19425 | 0.102053 | -0.67956 | -0.91274 | -0.52965 | -0.12603 | -0.42587 | 0.328443 | -0.39441 | 3.425408 | 0.013556 | -0.18958 | -0.35356 | -0.52419 | -0.18611 | -0.59627 | -0.09572 | -0.32113 |
| 577 | 0.220667 | -1.1357 | -0.25133 | 0.469725 | -0.72293 | -0.97859 | -0.71466 | -0.79487 | -0.51045 | 0.085465 | -0.51119 | 4.139543 | -0.14578 | -0.07287 | -0.47957 | -0.34145 | -0.29595 | -0.79421 | -0.29898 | -0.49286 |
| 578 | -0.23969 | -1.42098 | -0.7427 | 0.441638 | -0.36103 | -0.79122 | -0.70597 | -0.6209 | -0.57184 | 0.336573 | -0.57198 | 2.622352 | -0.04994 | -0.11288 | -0.32369 | -0.10359 | -0.43016 | -0.58973 | -0.26411 | -0.51265 |
| 579 | -0.39321 | -1.35501 | -0.67051 | 0.581535 | -0.96185 | -0.90302 | -0.37639 | -0.65065 | -0.66058 | 0.306272 | -0.46298 | 0.372106 | -0.27693 | -0.48716 | -0.43592 | 0.010101 | -0.49255 | 0.208704 | -0.18603 | -0.41179 |
| 580 | -0.40512 | -1.05961 | -0.34165 | 0.459464 | -0.89413 | -0.50085 | -0.59705 | -0.51409 | -0.42715 | -0.08496 | -0.48096 | -0.05433 | -0.50912 | -0.06232 | -0.29298 | -0.2558 | -0.2787 | 0.214769 | -0.72388 | -0.64203 |
| 581 | -0.32969 | -0.74949 | -0.08033 | 0.208595 | -0.30877 | -0.81433 | -0.36127 | -0.44294 | -0.1326 | -0.33723 | -0.67122 | -0.55296 | -0.57893 | -0.25234 | -0.48602 | -0.3307 | -0.24043 | 0.261181 | -0.38126 | -0.62663 |
| 582 | -0.42065 | 0.053027 | -0.32162 | 0.31757 | -0.06013 | -0.86048 | -0.29101 | 0.151136 | -0.15789 | -0.33156 | -0.42775 | -0.77243 | -0.62371 | -0.54634 | -0.21794 | -0.60327 | -0.21218 | 0.243537 | 0.220926 | -0.39522 |
| 583 | -0.03738 | 0.527884 | 0.03457 | 0.125855 | 0.088101 | -0.84696 | -0.47218 | -0.08421 | -0.00184 | -0.54725 | -0.52017 | -0.74261 | -0.49112 | -0.3916 | -0.04271 | -0.47272 | -0.25936 | 0.048786 | 0.161982 | -0.14181 |
| 584 | 0.043708 | 0.674159 | 0.362049 | -0.05629 | 0.238779 | -1.07073 | -0.23479 | 0.276847 | 0.147634 | -0.8023 | -0.00059 | -0.93721 | -0.27243 | -0.27568 | 0.281038 | -0.60175 | -0.25808 | -0.384 | 0.195696 | -0.15533 |
| 585 | 0.1071 | 0.701463 | 0.352691 | -0.07757 | 0.605164 | -1.07138 | 0.249581 | 0.197455 | 0.048862 | -0.77787 | 0.132712 | -0.76047 | -0.01657 | -0.56519 | 0.02211 | -0.39475 | -0.22369 | -0.29357 | 0.433188 | -0.0775 |
| 586 | 0.009042 | 0.558146 | 0.337719 | 0.149313 | 0.560242 | -1.01923 | 0.018818 | 0.449769 | 0.110482 | -0.48498 | 0.53953 | -0.25583 | -0.28444 | -0.16633 | -0.12123 | -0.53918 | -0.30628 | -0.278 | 0.023844 | -0.2062 |
| 587 | 0.219482 | 0.595994 | 0.325711 | -0.04057 | 0.453614 | -0.8486 | -0.04294 | -0.07649 | 0.096995 | -0.60906 | 0.276198 | -0.0099 | -0.34284 | 0.012921 | -0.06358 | 0.227185 | -0.38788 | -0.40834 | -0.6035 | -0.43636 |
| 588 | 0.150262 | -0.03879 | 0.460889 | -0.025 | 0.214221 | -1.07395 | 0.152942 | -0.12363 | 0.175578 | -0.54973 | 0.085157 | -0.19335 | -0.82139 | -0.22741 | -0.27422 | 0.222975 | -0.43876 | -0.35115 | -0.45609 | -0.53466 |
| 589 | -0.00643 | -0.05382 | 0.059906 | -0.07152 | 0.226681 | -0.39248 | -0.55377 | -0.15705 | -0.35583 | -0.276 | 0.117853 | -0.26782 | -0.75188 | 0.218475 | -0.45149 | 0.169631 | -0.74923 | -0.64788 | -0.35098 | -0.18456 |
| 590 | -0.37247 | -0.07276 | 0.309279 | -0.26833 | 0.22477 | -0.44003 | -0.61929 | -0.18717 | -0.2714 | -0.34026 | 0.057379 | -0.24959 | -0.96751 | 0.237516 | -0.4895 | 0.293677 | -0.9571 | -0.51112 | -0.26762 | -0.11052 |
| 591 | -0.51196 | -0.48081 | 0.403673 | -0.58001 | -0.24924 | -0.40471 | -0.47321 | -0.61997 | -1.08203 | -0.56445 | -0.32696 | -0.25811 | -0.75945 | -0.2351 | -0.7971 | 0.446573 | -0.35977 | -0.59142 | -0.21159 | 0.311206 |
| 592 | -0.61129 | -0.61444 | 1.054068 | -0.8786 | -0.48038 | -0.84722 | -0.24567 | -0.43558 | -0.81347 | -0.35866 | -0.2266 | -0.56965 | -0.63491 | -0.18951 | -0.96731 | -0.21223 | -0.06881 | -0.56584 | 0.308182 | 0.566558 |
| 593 | -0.87177 | -0.46963 | 0.925527 | -0.97269 | -0.59469 | -0.71161 | -0.02032 | -0.27752 | -0.59613 | -0.10316 | -0.13547 | -0.52876 | -0.51302 | 0.227966 | -0.82414 | -0.28171 | 0.104512 | -0.70841 | 0.252555 | 0.481137 |
| 594 | -0.67698 | -0.44016 | 0.861352 | -0.9153 | -0.7423 | -1.08901 | 0.137437 | -0.11574 | -0.13283 | -0.23758 | -0.52369 | 0.045407 | -0.79327 | -0.18095 | -0.9709 | -0.12234 | 0.305239 | -0.91543 | 0.229761 | 0.041215 |
| 595 | -0.42367 | -0.69857 | 0.477502 | -0.61686 | -1.06071 | -1.20037 | 0.221244 | -0.06165 | -0.04298 | -0.04908 | -0.75646 | -0.05577 | -0.51965 | -0.05427 | -1.00888 | -0.15885 | 0.145516 | -0.92117 | -0.40757 | 0.033808 |
| 596 | -0.36659 | -0.60952 | 0.284289 | -0.57838 | -0.88113 | -1.22674 | 0.188452 | -0.01986 | 0.170971 | 0.180702 | -0.3346 | -0.22438 | -0.80579 | 0.238438 | -0.41825 | 0.035176 | -0.45638 | -0.93403 | -0.39373 | -0.08407 |
| 597 | -0.53528 | -0.50728 | -0.2301 | -0.60399 | -0.75504 | -1.13428 | 0.104317 | 0.150132 | -0.14443 | 0.40005 | -0.14834 | 0.313471 | -0.84939 | 0.368814 | -0.41075 | 0.402081 | -0.66921 | -0.97037 | -1.00244 | -0.46249 |
| 598 | -0.40443 | -0.46268 | -0.10289 | -0.1624 | -0.55921 | -0.91954 | 0.177008 | 0.385704 | -0.58812 | 0.079324 | -0.06497 | 0.487095 | -1.63089 | 0.083766 | -1.08458 | 0.788476 | -0.88885 | -0.5091 | -1.09264 | -0.42159 |
| 599 | -0.66779 | -0.39548 | -0.27591 | 0.081209 | -0.40838 | -0.76395 | -0.11544 | 0.016098 | -0.58139 | 0.252812 | -0.15449 | 0.22354 | -1.77677 | 0.428832 | -0.87116 | 0.697786 | -1.08873 | -0.19438 | -1.00318 | -0.21317 |
| 600 | -0.74447 | -0.07932 | -0.8653 | -0.21724 | -0.44823 | -0.82379 | -0.5644 | -0.15911 | -0.86084 | 0.128571 | -0.16654 | 0.324718 | -2.29661 | 0.520093 | -0.76375 | 0.697786 | -0.81637 | 0.397378 | -0.74187 | -0.34919 |
| 601 | -0.7456 | 0.137179 | -0.93583 | -0.17092 | -0.26916 | -1.17524 | -0.76914 | -0.02507 | -0.52335 | -0.07539 | -0.31968 | 0.096049 | -2.68253 | 0.067867 | -0.90344 | -0.02117 | -0.66833 | 1.344143 | -0.80183 | -0.53009 |
| 602 | -0.70911 | -0.10893 | -0.73876 | -0.05214 | -0.49394 | -0.97949 | -0.685 | -0.30151 | -0.59703 | -0.37613 | -0.64394 | -0.58609 | -2.32119 | -0.35143 | -1.1375 | -0.65074 | -0.69225 | 2.324094 | -0.64478 | -0.54249 |
| 603 | -0.43182 | -0.1821 | -0.99135 | -0.8138 | -0.90238 | -1.03997 | -1.0472 | -0.73915 | -0.85292 | -0.31512 | -0.73967 | 0.071616 | -0.83413 | -0.01657 | -0.65729 | -1.11781 | -0.95832 | 2.067963 | -0.74826 | -0.18928 |
| 604 | -0.03278 | -0.13489 | -0.81584 | -0.76214 | -0.74654 | -1.29501 | -0.76903 | -0.39642 | -0.92192 | -0.665 | -1.00378 | 0.324685 | -0.01318 | -0.34438 | -0.45583 | -1.37149 | -0.66528 | 1.521307 | -0.61001 | -0.2399 |
| 605 | -0.24319 | -0.0949 | -0.70272 | -0.75609 | -0.82014 | -1.01658 | -0.62654 | -0.34154 | -1.28565 | -0.57056 | -1.0094 | 0.498037 | 0.523416 | -0.51889 | -0.14061 | -1.39781 | -0.7501 | 1.055523 | -0.31698 | -0.27804 |
| 606 | -0.29451 | -0.1573 | -0.72512 | -0.77931 | -0.95818 | -0.73156 | -0.41153 | -0.31287 | -1.14193 | -0.63401 | -0.80193 | 0.327054 | 1.175803 | -0.39992 | -0.35003 | -1.02443 | -0.85655 | 0.100661 | -0.59562 | -0.51973 |
| 607 | -0.31329 | -0.27911 | -0.54353 | -0.83107 | -0.75219 | -0.56886 | -0.36353 | 0.045845 | -0.96758 | -0.51988 | -0.65687 | 1.063743 | 0.796965 | 0.00551 | -0.40717 | -0.42743 | -0.80718 | -0.47118 | -0.58843 | -0.28881 |
| 608 | -0.77036 | -0.46813 | -0.53874 | -0.09984 | -0.33464 | -0.66358 | -0.25624 | 0.023772 | -0.6933 | -0.29697 | -0.84162 | 0.106254 | 0.031395 | -0.02869 | -0.18601 | -0.39436 | -0.40344 | -0.51893 | -0.3689 | -0.6652 |
| 609 | -1.1103 | -0.34606 | -0.25769 | -0.44394 | 0.374795 | -0.60849 | -0.35315 | -0.50016 | -0.60701 | -0.14707 | -0.55009 | 0.047844 | -0.67447 | -0.29002 | -0.68537 | -0.66224 | -0.77998 | -0.07946 | -0.70998 | -0.83309 |
| 610 | -0.81144 | 0.844138 | -0.37545 | -0.5346 | 0.487428 | -0.66438 | -0.15316 | -0.58138 | -0.1073 | -0.35562 | -0.5528 | -0.13629 | -0.77613 | -0.32227 | -0.77658 | -0.65308 | -0.6922 | -0.50139 | -0.74699 | -0.80308 |
| 611 | -0.20185 | 0.971105 | -0.52281 | -0.63299 | 0.677429 | -0.46492 | -0.27609 | -0.93991 | -0.28638 | -0.51538 | -0.86957 | 0.237249 | -1.02895 | -0.10417 | -0.7933 | -0.76315 | -0.84332 | -0.59001 | -0.45039 | -0.62824 |
| 612 | -0.23483 | 1.193773 | -0.99305 | -0.50118 | 0.623756 | -0.69532 | -0.17242 | -1.14928 | -0.28412 | -0.49418 | -0.73316 | 0.014724 | -0.91791 | -0.45557 | -0.63485 | -1.0874 | -0.54256 | -1.01541 | -0.17495 | -0.73249 |
| 613 | 0.078497 | 1.619457 | -1.088 | -0.7009 | 0.593432 | -0.66414 | -0.47402 | -0.84524 | -0.23059 | -0.29997 | -0.53413 | 0.82364 | -1.0201 | -0.92123 | -0.85991 | -1.22915 | -0.64482 | -0.75613 | -0.33727 | -0.22177 |
| 614 | 0.785451 | 1.623082 | -1.56063 | -0.17155 | -0.23742 | -0.75709 | -0.46184 | -0.78778 | -0.54732 | -0.34349 | -0.67159 | 0.796323 | -0.52106 | -0.55073 | -0.24875 | -0.97729 | -0.08815 | -0.90595 | -0.12732 | -0.13754 |
| 615 | 1.460737 | 0.392889 | -1.01038 | -0.36664 | -0.1512 | -0.63426 | -0.40375 | -0.39208 | -0.59343 | -0.46885 | -0.18966 | 0.932384 | -0.24569 | -0.36184 | -0.24525 | -0.96013 | -0.24957 | -0.89208 | -0.19464 | -0.04114 |
| 616 | 0.947732 | 0.329853 | -0.57934 | -0.35906 | -0.27461 | -0.55059 | -0.2346 | 0.037339 | -0.50891 | -0.61522 | -0.08036 | 0.648251 | -0.13205 | -0.30245 | -0.23887 | -0.74758 | 0.185207 | -0.91913 | -0.02129 | -0.21506 |
| 617 | 0.96386 | 0.355256 | -0.56592 | -0.2437 | -0.35594 | -0.32586 | -0.60849 | -0.07555 | -0.5728 | -0.90138 | -0.10895 | 0.252193 | -0.02425 | -0.01861 | -0.22957 | -0.47818 | -0.49385 | -0.61647 | -0.34823 | -0.19084 |
| 618 | 1.006432 | 0.0457 | -0.39059 | -0.13564 | -0.44533 | -0.35403 | -0.19738 | -0.55171 | -0.22413 | -1.20118 | 0.086035 | -0.44552 | 0.411618 | -0.15112 | 0.053623 | -0.34931 | -0.13162 | -0.88947 | -0.20175 | -0.42045 |
| 619 | 0.817633 | -0.45308 | -0.47121 | -0.48483 | -0.46013 | -0.33704 | 0.3356 | -0.24231 | 0.059885 | -0.9881 | -0.07334 | -0.43589 | 0.128461 | -0.13887 | -0.08017 | 0.491461 | -0.4224 | -0.27855 | -0.36008 | -0.5271 |
| 620 | -0.02283 | -0.46369 | -0.62635 | -0.23331 | -0.6073 | -0.71151 | 0.597917 | -0.37828 | 0.223731 | -0.89507 | -0.07823 | -0.87776 | 0.155954 | -0.43016 | -0.02438 | 0.505413 | -0.34345 | -0.36083 | -0.62423 | -0.57908 |
| 621 | 0.018343 | -0.5366 | -0.91671 | -0.41057 | -0.87125 | -0.50708 | 0.42486 | -0.31048 | 0.004997 | -0.26939 | -0.17514 | -0.69542 | 0.339334 | -0.08026 | 0.272943 | 0.295221 | -0.41688 | -0.14092 | -0.74718 | -0.31964 |
| 622 | -0.15175 | -0.42689 | -0.49871 | -0.33009 | -0.60637 | -0.86074 | 0.647084 | -0.39219 | 0.136145 | 0.083044 | -0.44992 | -0.67741 | 0.319559 | -0.35477 | 0.212927 | 0.27558 | -0.18067 | -0.06961 | -0.49823 | -0.70371 |
| 623 | -0.22544 | -0.28083 | -0.73977 | -0.55799 | -0.42426 | -0.98618 | 0.591949 | -0.0744 | 0.396746 | 0.12631 | -0.83594 | -0.5189 | 0.174132 | -0.06409 | 0.166374 | 0.288459 | -0.46813 | 0.10564 | -0.46496 | -0.33157 |
| 624 | -0.80872 | -0.16195 | -0.35988 | -0.50382 | -0.49334 | -1.40183 | 0.187274 | -0.05738 | 0.315832 | 0.131021 | 0.017876 | -1.01594 | 0.008108 | -0.13681 | 0.06177 | -0.57908 | -0.15686 | 0.097251 | -0.50473 | 0.470345 |
| 625 | -0.56964 | -0.08165 | -0.53513 | 0.32256 | -0.25287 | -1.14246 | -0.07883 | -0.45466 | 0.034115 | -0.03071 | 0.093045 | -1.1078 | -0.22607 | 0.195164 | -0.29699 | -0.49196 | -0.02259 | 0.210146 | -0.31655 | 1.155567 |
| 626 | -0.61829 | -0.24738 | -0.49977 | 0.813117 | -0.19602 | -1.64007 | 0.06312 | -0.64144 | -0.05173 | -0.54799 | 0.097413 | -1.10318 | -0.08159 | -0.32261 | -0.57839 | -0.42628 | -0.12637 | 0.36129 | -0.3027 | 0.812803 |
| 627 | -0.5408 | -0.25786 | -0.84695 | 0.815049 | -0.05388 | -1.57991 | 0.347968 | -0.34949 | -0.07582 | -0.89147 | 0.377724 | -1.47216 | -0.37623 | -0.26104 | -0.78231 | -0.67798 | 0.133753 | 0.758601 | -0.5012 | 1.332008 |
| 628 | -0.54493 | -0.36838 | -0.3808 | 1.084947 | -0.23517 | -1.6498 | 0.136363 | -0.84341 | -0.7656 | -1.16053 | 0.446668 | -0.44262 | -0.52928 | -0.23089 | -0.87157 | -0.84739 | 0.014447 | 0.68662 | -0.06194 | 0.896001 |
| 629 | -0.44799 | -0.14366 | -0.38876 | 0.880664 | -0.06864 | -1.20667 | 0.175716 | -1.03584 | -1.11747 | -1.3318 | -0.13762 | 0.50115 | -0.50908 | -0.04321 | -0.79086 | -0.55093 | -0.04687 | 0.398196 | 0.702452 | 0.511721 |
| 630 | -0.43823 | -0.20267 | -0.04648 | -0.22621 | -0.15619 | -1.21883 | 0.144081 | -1.00778 | -0.97397 | -1.02363 | -0.54954 | 1.300667 | -0.66097 | -0.34635 | -0.48294 | -0.75157 | -0.38438 | 0.244317 | 0.637223 | -0.09649 |
| 631 | -0.44678 | -0.09999 | 0.384455 | -0.45014 | -0.08419 | -0.73664 | 0.092704 | -1.03746 | -0.97182 | -1.00208 | -0.52808 | 1.261407 | -1.16645 | 0.420143 | -0.22088 | -0.73854 | -0.40774 | 0.141412 | 0.660481 | -0.35314 |
| 632 | -0.30204 | -0.37889 | 0.274451 | -1.00168 | -0.36047 | -0.94837 | -0.51399 | -1.32307 | -0.86363 | -0.6586 | -0.72211 | 1.709024 | -1.01976 | 0.691563 | 0.183675 | -0.5585 | -0.6665 | 0.014773 | 0.626595 | -0.26175 |
| 633 | -0.1635 | -0.37224 | 0.040722 | -0.888 | -0.30921 | -0.65849 | -0.13583 | -0.99217 | -0.69983 | -0.78099 | -0.64102 | 0.50046 | -1.17395 | 0.836846 | 0.310469 | -0.31523 | -0.63955 | -0.00065 | 0.019039 | -0.27299 |
| 634 | -0.07264 | -0.69471 | -0.00242 | -0.53773 | -0.31166 | 0.280453 | -0.13953 | -0.96327 | -0.2862 | -0.59541 | -0.3811 | 0.050555 | -1.20939 | 0.612936 | 0.03935 | -0.17683 | -0.99347 | -0.24456 | -0.0818 | -0.18677 |
| 635 | -0.43262 | -0.87628 | -0.29578 | -0.35587 | -0.496 | 0.164883 | -0.18108 | -0.89255 | -0.58031 | -0.39556 | 0.354006 | -0.92947 | -1.28568 | 0.735806 | -0.05424 | -0.41178 | -0.93236 | 0.001867 | 0.165031 | -0.1294 |
| 636 | -0.16626 | -1.02573 | -0.77208 | -0.42636 | -0.47808 | 0.225844 | -0.07801 | -0.99287 | -0.67586 | 0.150197 | 1.003056 | -0.92832 | -1.17891 | 0.120168 | -0.16677 | 0.263145 | -0.8791 | 0.001867 | -0.18671 | 0.337167 |
| 637 | -0.24462 | -0.93697 | -0.48035 | -0.28164 | -0.72938 | 0.33822 | 0.253456 | -1.52583 | -1.08254 | 0.19062 | 1.068643 | -0.92219 | -1.27514 | -0.34528 | -0.21066 | -0.09376 | -0.67876 | -0.58201 | 0.140488 | 0.155879 |
| 638 | -0.38301 | -0.93301 | -0.41831 | -0.59364 | -0.64332 | 0.027949 | 0.086911 | -1.23093 | -1.29243 | 0.604281 | 1.191077 | -0.90061 | -0.38942 | -0.50504 | -0.45209 | -0.16296 | -0.23815 | -1.33839 | -0.07819 | 0.171262 |
| 639 | -0.23034 | -0.77014 | -0.24045 | -0.79618 | -1.23437 | -0.59279 | -0.11119 | -1.06961 | -1.37449 | 0.818098 | 0.855763 | -1.17855 | -0.58286 | -0.52566 | -0.46854 | -0.30081 | -0.14297 | -1.29429 | -0.68179 | -0.35248 |
| 640 | 0.092102 | -0.57107 | -0.17326 | -0.8539 | -1.28109 | -0.15902 | -0.11666 | -0.7011 | -0.98648 | 0.747272 | -0.2558 | -0.49537 | -0.65405 | -0.67032 | -0.45172 | 0.105584 | 0.443399 | -1.60381 | -0.75123 | -0.72411 |
| 641 | -0.00185 | -0.46757 | -0.3093 | -1.07673 | -1.43578 | -0.25224 | -0.06228 | -0.47814 | -0.69949 | 0.259274 | -0.86243 | -0.19616 | -0.82778 | -0.67685 | -0.6709 | -0.33397 | 0.829025 | -1.81118 | -0.70981 | -0.4502 |
| 642 | 0.003764 | -0.39203 | -0.60629 | -0.95688 | -1.22126 | -0.30324 | -0.16323 | 0.164359 | -0.59396 | 0.05741 | -0.96552 | -0.03659 | -0.38895 | -0.68664 | -0.63458 | 0.279751 | 0.859844 | -1.52979 | -1.02229 | -0.25873 |
| 643 | 0.003611 | -0.29212 | -0.74069 | -0.72625 | -1.45051 | -0.11129 | -0.45457 | -0.43533 | -0.54819 | 0.210161 | -1.31412 | 0.154097 | -0.96743 | -0.70231 | -0.50556 | 0.050824 | 0.68 | -0.89765 | -0.41328 | -0.44679 |
| 644 | -0.27122 | -0.38549 | -0.91346 | -0.61159 | -0.44408 | -0.55387 | -0.56823 | -0.53005 | -0.7577 | 0.007162 | -1.01361 | 0.19288 | -0.86153 | -0.6604 | -0.52271 | 0.043819 | 0.811077 | -0.39609 | -0.20174 | -0.18598 |
| 645 | -0.05221 | -0.41368 | -0.90123 | -0.42705 | -0.21877 | -0.73991 | -0.5212 | -0.89037 | -1.08805 | -0.26992 | -0.25538 | 0.148081 | -0.52518 | -0.26535 | -0.4509 | -0.02805 | 0.446347 | -0.11068 | -0.18834 | -0.04719 |
| 646 | -0.42458 | -0.23178 | -0.6251 | -0.32636 | -0.0547 | -0.92935 | -0.89395 | -1.12231 | -1.24642 | -0.13512 | -0.22671 | 0.548184 | -0.31463 | -0.36648 | -0.34554 | -0.04557 | -0.15866 | 0.257513 | -0.17126 | -0.44004 |
| 647 | -0.25629 | -0.23526 | -0.32812 | -0.32828 | 0.011541 | -0.60138 | -1.0014 | -1.01443 | -1.22909 | -0.11584 | 0.127694 | 0.842081 | -0.50927 | 0.004044 | -0.39213 | -0.23073 | -0.38357 | 0.56616 | 0.041522 | -0.67203 |
| 648 | -0.407 | 0.094524 | -0.31395 | -0.44992 | 0.542383 | -0.74769 | -0.68704 | -0.69688 | -0.85948 | -0.35937 | 0.612185 | 0.679568 | -0.47469 | 0.004044 | -0.35673 | -0.10997 | -0.38357 | 0.519344 | -0.1988 | -0.57585 |
| 649 | -0.16245 | 0.32978 | -0.31395 | -0.42805 | 0.129453 | -0.37488 | -0.60693 | -0.59831 | -0.56678 | -0.35879 | 0.539779 | 0.755788 | -0.18023 | -0.08928 | 0.283727 | -0.04758 | -0.38357 | 0.031571 | -0.49909 | -0.78641 |
| 650 | -0.62335 | 0.107452 | -0.14219 | -0.42805 | 0.129453 | -0.7225 | -0.58129 | -0.3662 | -0.30355 | -0.21073 | -0.00849 | 0.420235 | 0.554622 | -0.58233 | 0.383352 | 0.031612 | -0.59793 | -0.03775 | -0.49909 | -0.25477 |
| 651 | -0.67092 | 0.051338 | -0.14219 | 0.028736 | 0.257621 | -0.97253 | -0.28423 | -0.24581 | -0.08337 | -0.08722 | -0.00849 | -0.28339 | 0.870308 | -0.44467 | 0.307026 | 0.031612 | 0.039912 | -0.21548 | -0.24295 | 0.05947 |
| 652 | -0.60302 | -0.21413 | -0.37905 | 0.197104 | 0.012907 | -1.2328 | 0.313019 | -0.32404 | 0.181606 | 0.028048 | -0.49726 | -0.71877 | 0.586384 | -0.49494 | 0.282086 | -0.2249 | 0.234002 | -0.00912 | -0.10503 | 0.27275 |
| 653 | -0.96548 | -0.81906 | 0.221264 | 0.110815 | -0.31835 | -1.19427 | 0.382242 | -0.17379 | 0.015846 | -0.1532 | -0.94537 | -0.26919 | 0.493135 | -0.49494 | 0.373503 | -0.08534 | 0.383583 | 0.080391 | -0.31568 | -0.09651 |
| 654 | -1.38692 | -1.10717 | 0.182355 | -0.21556 | -0.41214 | -1.2659 | 0.489309 | -0.31809 | -0.11018 | -0.31741 | -1.011 | -0.3852 | 0.344526 | -0.49918 | -0.08881 | -0.56175 | 0.314113 | -0.33061 | -0.33571 | 0.348504 |
| 655 | -1.24935 | -0.82081 | -0.18898 | -0.45432 | -0.71072 | -1.13842 | 0.035888 | -0.53248 | 0.259576 | -0.38106 | -0.79682 | -0.0634 | -0.49061 | -0.28142 | -0.22385 | -0.74287 | 0.37537 | -0.6505 | -0.72254 | 0.631911 |
| 656 | -1.09537 | -0.55867 | -0.25448 | -0.60033 | -0.50481 | -1.0479 | -0.21112 | -0.50439 | 0.599597 | -0.40529 | -1.03809 | -0.18089 | -0.43812 | -0.17314 | 0.158062 | -0.46826 | -0.34308 | -0.65613 | -0.89988 | 0.546606 |
| 657 | -1.06015 | -0.25886 | -0.04696 | -0.55724 | -0.34843 | -0.90468 | -0.53338 | -0.37176 | 0.427326 | 0.204588 | -0.8478 | -0.35871 | -0.28154 | -0.29015 | 0.343842 | -0.30872 | -0.56166 | -1.50345 | -0.84766 | 0.414333 |
| 658 | -0.79269 | -0.06063 | -0.37027 | -0.20243 | -0.08599 | -0.60862 | -0.55591 | -0.02806 | 0.18291 | 0.478954 | -0.57753 | -0.69297 | -0.331 | -0.07597 | -0.09753 | -0.47565 | -0.85475 | -1.11157 | -0.6523 | 0.902615 |
| 659 | -0.66437 | 0.23554 | -0.48316 | 0.056886 | 0.30949 | -0.53818 | -0.2248 | 0.471762 | 0.550157 | 1.443704 | -0.73024 | -0.59306 | -0.54379 | -0.16531 | 0.215329 | -0.33139 | -0.79841 | 0.013871 | -0.56545 | 0.917008 |
| 660 | -0.47178 | 0.028336 | -0.41484 | 0.406687 | 0.654187 | -0.00147 | 0.301242 | 0.735535 | 0.131781 | 2.226725 | -0.54493 | -0.28727 | -0.31941 | -0.23795 | 0.067982 | 0.042562 | -0.99269 | 0.553134 | -0.44295 | 0.310278 |
| 661 | -0.38076 | 0.063799 | -0.22798 | 0.195344 | 0.759825 | 0.058668 | 0.641193 | 0.86798 | -0.4544 | 1.869769 | -0.30251 | -0.09257 | -0.4674 | -0.25655 | -0.35634 | -0.1856 | -0.82002 | 0.215355 | -0.2173 | 0.066548 |
| 662 | -0.50222 | 0.42914 | -0.09605 | -0.0506 | 1.165003 | 0.252516 | 0.457807 | 0.74 | -0.38896 | 1.510562 | 0.046853 | 0.006528 | -0.25676 | 0.097776 | -0.77687 | -0.4101 | -0.68702 | 0.540668 | -0.85879 | 0.210543 |
| 663 | -0.38047 | 0.472203 | -0.10521 | -0.51206 | 0.860815 | 0.190322 | 0.339382 | 0.492055 | -0.05247 | 1.27965 | -0.34957 | -0.0238 | -0.14946 | -0.22385 | 0.018769 | -0.62414 | -0.45885 | 0.16466 | -1.11086 | 0.451852 |

| | |
|---|---|
| 497–663 (remaining column groups) | [illegible] |

| | | | | | | | | | | | | | | | | | | | | | | | | | | | | | | | | | | | | | | | | | |
|---|---|---|---|---|---|---|---|---|---|---|---|---|---|---|---|---|---|---|---|---|---|---|---|---|---|---|---|---|---|---|---|---|---|---|---|---|---|---|---|---|---|
| 664 | -0.00581 | 0.414238 | 0.046589 | -0.46687 | 0.446745 | 0.19151 | -0.07042 | -0.03617 | -0.70074 | 0.51157 | -0.12981 | -0.10593 | 0.457981 | -0.17721 | 0.055746 | -0.34533 | -0.58629 | -0.40217 | -1.12008 | 1.274472 | -0.2381 | 0.340516 | 0.95604 | 0.093941 | -0.21717 | -0.31102 | -0.04047 | 0.102284 | -0.42346 | 1.412593 | -0.34427 | 1.702502 | 1.253614 | -0.24008 | 5.817962 | 0.410672 | -1.10025 | 0.201967 | 0.182556 | 1.438728 | [illegible] |
| 665 | 0.054506 | 0.58325 | 0.196959 | -0.4991 | 0.436855 | -0.36159 | -0.35974 | -0.23709 | -0.67567 | -0.43477 | -0.23062 | -0.61654 | 0.641734 | -0.34115 | 0.464465 | -0.73265 | -0.48124 | -0.55221 | -1.12073 | 2.0411 | -0.30628 | 0.141448 | 0.147086 | -0.13071 | 0.304964 | -0.35294 | -0.45176 | 0.170684 | -0.78315 | 0.688191 | -0.38591 | 2.0256 | 1.61652 | -0.22498 | 6.725385 | 0.116039 | -0.73873 | 0.026951 | -0.31119 | 2.335403 | [illegible] |
| 666 | 0.085097 | 0.341758 | 0.075593 | -0.38125 | -0.0821 | -0.05863 | -0.5908 | -0.35948 | -0.37892 | -0.03438 | -0.52236 | -0.53719 | 0.593452 | -0.60486 | 0.751312 | -0.82492 | -0.40926 | -0.1919 | -1.08147 | 2.457687 | -0.28896 | -0.06317 | -0.10992 | 0.451382 | 1.025472 | 0.237521 | -0.48591 | -0.18249 | -0.55146 | 0.397938 | -0.33633 | 2.335231 | 1.170023 | -0.27471 | 6.756213 | -0.00616 | -0.29727 | -0.41299 | -0.51078 | 1.913081 | [illegible] |
| 667 | 0.201772 | -0.1524 | -0.02699 | -0.38551 | -0.39895 | -0.17391 | -0.48363 | -0.48852 | -0.35118 | -0.25816 | -0.77504 | -0.31692 | 0.510169 | -1.03389 | 1.049642 | -0.50345 | -0.50142 | -0.29371 | -0.80496 | 2.591789 | -0.33846 | -0.42522 | -0.13972 | 0.897681 | 0.969315 | -0.2376 | -0.52594 | -0.1766 | -0.51589 | 0.104449 | -0.14337 | 1.677425 | 1.053686 | -0.47529 | 4.592391 | 0.548102 | -0.42175 | -0.01628 | -0.63501 | 1.826596 | [illegible] |
| 668 | 0.112866 | 0.257445 | -0.03762 | -0.31252 | -0.28357 | -0.12401 | -0.43492 | -0.74897 | -0.14576 | -0.0518 | -0.2235 | -0.23946 | 0.392815 | -0.92646 | 0.480069 | -0.15852 | -0.58608 | -0.20821 | -0.51933 | 2.135281 | -0.23297 | -0.58434 | -0.43922 | 1.024897 | 1.193871 | 0.118621 | -0.28973 | -0.35366 | -0.4708 | -0.10811 | -0.25939 | 1.143288 | 1.087495 | 0.394078 | 1.899444 | 0.426364 | -0.48599 | -0.13159 | -0.59047 | 1.749655 | [illegible] |
| 669 | -0.24144 | 0.850013 | -0.03762 | -0.55869 | -0.1134 | -0.12589 | -0.26731 | -0.84449 | 0.214353 | 0.12205 | -0.22492 | -0.19981 | -0.098 | -0.74523 | 0.46663 | -0.39228 | -0.68391 | -0.03732 | -0.40874 | 1.063809 | -0.30244 | -0.23591 | -0.23925 | 0.851975 | 1.122747 | 0.357044 | -0.39822 | -0.24757 | -0.22542 | -0.03741 | -0.04247 | 1.255406 | 0.989638 | 0.40209 | 1.102578 | 0.274325 | -0.15071 | -0.0999 | -0.30897 | 0.482232 | [illegible] |
| 670 | -0.5233 | 0.80066 | 0.021643 | -0.72749 | -0.14963 | 0.242402 | -0.07624 | -0.90948 | -0.02125 | 0.307369 | -0.32821 | -0.31491 | -0.40333 | -0.4116 | 0.322821 | -0.27699 | -0.44159 | -0.11761 | -0.42062 | 0.028802 | -0.1659 | 0.182633 | -0.01096 | 0.996089 | 0.427132 | -0.00652 | -0.29534 | -0.08029 | 0.284998 | -0.11791 | 0.117747 | 0.999427 | 0.616949 | 0.418605 | 0.264277 | 0.031566 | -0.03527 | -0.0999 | -0.13768 | -0.54117 | [illegible] |
| 671 | -0.66822 | 0.974163 | -0.35186 | -0.89588 | 0.039528 | 0.13402 | 0.360989 | -0.86639 | -0.04008 | 0.161967 | -0.13173 | -0.34153 | -0.167 | -0.17445 | 0.191142 | -0.25471 | -0.59583 | 0.15807 | 0.019187 | 0.210594 | 0.080674 | 0.656907 | -0.04386 | -0.09373 | -0.16022 | -0.25859 | 0.033794 | 0.208249 | 0.32494 | 0.196121 | -0.24668 | 0.450311 | 0.329037 | -0.24868 | 0.089187 | 0.120617 | -0.22936 | 0.111637 | 0.028487 | -0.27674 | [illegible] |
| 672 | -1.04058 | 1.473872 | -0.1484 | -0.77357 | 0.065858 | -0.15964 | 0.485255 | -0.96619 | -0.06604 | -0.07803 | -0.37956 | -0.48803 | -0.35602 | 0.017268 | -0.17871 | -0.25024 | -0.68864 | 0.191358 | -0.01629 | -0.08349 | 0.056265 | 0.731852 | -0.22171 | -0.6802 | -0.50758 | 0.292358 | 0.149386 | 0.208249 | 0.061297 | 0.264372 | -0.08216 | -0.10644 | 0.319676 | -0.34653 | -0.08123 | -0.32217 | -0.41929 | -0.61771 | 0.36512 | -0.18568 | [illegible] |
| 673 | -1.06811 | 1.146218 | -0.19368 | -0.74298 | 0.239123 | -0.20922 | 0.464871 | -0.87755 | -0.91041 | 0.013023 | -0.77903 | -0.54705 | -0.24215 | 0.15121 | -0.14681 | -0.22903 | -1.00874 | 0.331961 | -0.1676 | -0.08158 | 0.063203 | 0.72834 | -0.26193 | -0.70052 | -0.80024 | -0.03046 | 0.355253 | 0.146233 | -0.03427 | 0.368927 | -0.05808 | -0.449 | -0.01111 | -1.11706 | -0.205 | -0.12428 | -0.18342 | -0.77775 | 0.093222 | 0.015124 | [illegible] |
| 674 | -1.02593 | 0.331989 | -0.19368 | -0.4968 | 0.166888 | -1.14355 | 0.37778 | -0.89951 | -0.74265 | -0.41079 | -0.8784 | -0.75238 | -0.17289 | 0.183842 | -0.49547 | -0.08424 | -0.76171 | 0.086296 | -0.116 | 0.035199 | -0.07164 | 0.482166 | -0.55278 | -0.74415 | -0.75603 | -0.04726 | 0.656879 | 0.131658 | -0.06016 | 0.389998 | -0.05808 | -0.80339 | -0.22563 | -1.15553 | -0.50644 | -0.15389 | 0.082771 | -0.84449 | -0.27142 | 0.599679 | [illegible] |
| 675 | -0.88928 | 0.211904 | -0.46726 | -0.49931 | 0.102445 | -1.52421 | 0.37778 | -0.683 | -1.11023 | -0.36914 | -0.90582 | -0.75028 | -0.53464 | 0.04833 | -0.63203 | -0.0606 | -0.76171 | -0.0635 | -0.60822 | -0.37816 | -0.25954 | 0.283017 | -0.55278 | -1.0607 | -0.84034 | 0.173598 | 0.588662 | 0.245877 | -0.31926 | 0.143739 | -0.78348 | -0.7785 | -0.29109 | -1.23003 | -0.32262 | 0.088875 | -0.24148 | -0.90688 | -0.43461 | 0.170735 | [illegible] |
| 676 | -0.72539 | 0.246011 | -0.16113 | -0.33092 | -0.02565 | -1.82111 | 0.016791 | -0.6998 | -1.20145 | -0.24354 | -1.35001 | -1.03539 | -0.98368 | -0.26611 | -0.53912 | -0.03704 | -0.58202 | -0.64953 | -1.08371 | -1.02616 | -0.3238 | -0.04032 | -0.55243 | -1.12134 | -1.14871 | 0.201228 | 0.259526 | 0.248991 | -0.45119 | 0.480136 | -0.74 | -0.60215 | -0.63561 | -0.84597 | -0.569 | -0.08303 | -0.45148 | -1.1882 | -0.27727 | -0.26497 | [illegible] |
| 677 | -0.78784 | -0.04626 | -0.53752 | -0.3736 | -0.07331 | -1.73218 | -0.32297 | -0.34724 | -1.16167 | -0.13762 | -0.93294 | -1.01406 | -1.38772 | -0.80837 | -0.23742 | -0.11694 | -0.93929 | -0.70355 | -0.988 | -0.86881 | -0.67118 | 0.507581 | -0.73775 | -1.00165 | -1.39332 | -0.02844 | 0.012598 | 0.482375 | -0.04546 | 0.480136 | -0.85479 | -0.5686 | -0.70337 | -0.75703 | -0.72304 | -0.38244 | -0.25794 | -1.11169 | -0.68855 | -0.33407 | [illegible] |
| 678 | -0.87696 | -0.41284 | -0.83268 | -0.49692 | -0.44422 | -2.11686 | -0.24543 | -0.51924 | -0.45835 | -0.53378 | -0.49515 | -0.98774 | -1.48672 | -0.97945 | -0.2627 | -0.13816 | -0.52578 | -0.75404 | -0.83669 | -1.10908 | -0.74621 | 0.356332 | -0.68269 | -1.29392 | -0.85494 | 0.466648 | -0.32642 | 0.376152 | 0.147833 | 0.342273 | -0.77344 | -0.39034 | -0.66567 | -0.67637 | -0.77114 | -0.9225 | -0.5577 | -1.05499 | -0.52693 | -0.26871 | [illegible] |
| 679 | -0.66075 | -0.36249 | -1.17766 | -0.55519 | -0.74809 | -1.37663 | -0.29212 | -0.3912 | -0.62439 | -0.14364 | 0.014485 | -0.7351 | -1.39768 | -1.29878 | -0.28897 | 0.11206 | -0.7413 | -1.0099 | -1.0805 | -1.36952 | -0.41085 | 0.152315 | -0.46782 | -0.73769 | -0.65167 | 0.572233 | -0.6067 | 0.090328 | -0.02071 | 0.328282 | -0.90679 | -0.03755 | -0.63877 | -0.34343 | -0.5339 | -1.04384 | -0.87961 | -1.11034 | -0.32108 | -0.49644 | [illegible] |
| 680 | -0.66075 | 0.012673 | -1.16224 | -0.46269 | -0.68365 | -1.28869 | 0.199893 | -0.3912 | -0.11963 | 0.419865 | -0.12323 | -0.38464 | -1.03844 | -1.13224 | -0.12222 | 0.220316 | -0.7534 | -0.99119 | -0.31141 | -0.96574 | -0.34954 | 0.776878 | -0.72755 | -0.42114 | -0.5483 | 0.832965 | -0.27472 | -0.12308 | 0.124444 | 0.35717 | -0.10615 | 0.274653 | -0.77277 | 0.151535 | -0.67327 | -0.98208 | -0.81826 | -1.04795 | -0.03498 | -0.21916 | [illegible] |
| 681 | -0.4973 | -0.46393 | -1.36666 | -0.46269 | -0.75842 | -1.02988 | 0.199893 | -0.39693 | -0.06017 | 0.411743 | 0.740414 | -0.09953 | -1.08253 | -0.8096 | -0.32778 | 0.260485 | -0.71994 | -0.83192 | -0.3633 | -0.75541 | -0.24519 | 0.945191 | -0.77676 | -0.28969 | -0.25984 | 1.152958 | -0.58415 | -0.23525 | 0.037376 | -0.3221 | 0.06299 | 0.32356 | -0.29389 | -0.01904 | -0.68832 | -0.93254 | -0.52392 | -0.74203 | -0.35848 | -0.13483 | [illegible] |
| 682 | -0.39876 | -0.53401 | -1.46873 | -0.31046 | -0.73708 | -1.04162 | 0.066593 | -0.12139 | -0.06014 | 0.545825 | 0.618688 | 0.025635 | -0.81833 | -0.26735 | -0.52243 | 0.335912 | -0.09211 | -0.81264 | -0.73683 | -1.05205 | -0.04609 | 0.144891 | -0.5408 | -0.0757 | -0.10342 | 0.848024 | -0.33722 | -0.46864 | -0.26344 | -0.3221 | 0.740059 | 0.403334 | -0.31677 | -0.04373 | -0.75556 | -0.63314 | -0.37271 | -0.46171 | -0.17953 | -0.0198 | [illegible] |
| 683 | -0.03548 | -0.26833 | -1.237 | -0.18204 | -0.78899 | 0.266256 | 0.212302 | 0.148034 | -0.20446 | 0.755259 | 0.367375 | 0.079349 | -0.71501 | -0.19073 | 0.025833 | -0.07873 | -0.25754 | -0.73936 | -0.67976 | -0.8017 | -0.03536 | 0.437529 | -0.5408 | 0.005295 | -0.59265 | 1.089001 | -0.34331 | -0.3004 | -0.37705 | -0.33186 | 0.694292 | 0.312629 | -0.2125 | -0.04373 | -0.25442 | -0.22772 | 0.027008 | -0.18325 | -0.20016 | 0.058715 | [illegible] |
| 684 | -0.04274 | -0.19666 | -1.06136 | -0.12948 | -0.66162 | 0.896723 | -0.23516 | 0.067795 | -0.06093 | 0.399126 | -0.04289 | -0.14408 | -0.93066 | -0.06586 | 0.16524 | -0.46688 | -0.00319 | -0.40873 | -0.68107 | -0.89449 | -0.542 | 0.634808 | -0.7178 | -0.23404 | -0.65678 | 0.819999 | -0.18491 | -0.1361 | 0.788128 | 0.789398 | 0.827642 | 0.089723 | 0.343917 | -0.29226 | -0.34944 | -0.10639 | 0.082722 | 0.146075 | -0.38973 | -0.02219 | [illegible] |
| 685 | -0.08119 | -0.49271 | -0.80321 | -0.23931 | -0.66162 | 1.09534 | 0.010793 | -0.12812 | 1.005458 | -0.34123 | 0.025144 | -0.40701 | -0.31532 | -0.20746 | -0.10018 | -0.90797 | -0.19735 | -0.80002 | -0.79979 | | -0.51811 | -0.13998 | -0.6181 | 0.202848 | -0.4458 | 0.412422 | -0.01618 | -0.1361 | 2.084088 | 3.692936 | 0.742159 | -0.05944 | 1.094228 | -0.67063 | 0.963635 | -0.06194 | 0.217541 | 0.053813 | -0.51562 | 0.180461 | [illegible] |
| 686 | -0.28805 | -0.36304 | -0.57885 | -0.14558 | -0.38411 | 1.246519 | 0.28072 | -0.06226 | 1.159773 | -0.39914 | -0.0149 | -0.39525 | -0.45755 | -0.20746 | -0.19196 | -1.27573 | 0.009215 | -0.31756 | -1.11557 | -0.97125 | -0.47363 | -0.63436 | -0.20464 | 0.508972 | -0.43857 | 0.307996 | 0.307931 | 0.343791 | 2.598625 | 5.552888 | 0.931852 | -0.175 | 0.535628 | -0.65637 | 1.653245 | -0.35117 | 0.296985 | -0.07187 | -0.84809 | 0.372594 | [illegible] |
| 687 | -0.31698 | -0.35047 | -0.17826 | 0.194095 | -0.73855 | 1.396614 | 0.86982 | -0.52005 | 0.79027 | -0.44326 | -0.1144 | -0.30749 | -0.17447 | -0.08753 | 0.032214 | -1.64001 | -0.19096 | -0.25033 | -0.89724 | -0.72612 | -0.3511 | -0.36055 | -0.01233 | 0.757046 | -0.71127 | 0.38903 | 0.017836 | 0.298581 | 2.515693 | 5.573471 | 0.144804 | 0.231009 | 0.519078 | -0.38757 | 1.725266 | -0.47005 | 0.295066 | -0.1585 | -0.70824 | 0.179506 | [illegible] |
| 688 | -0.38687 | -0.54528 | -0.45091 | 0.346504 | -0.38374 | 0.256168 | 0.721167 | -0.73615 | 0.828785 | -0.4784 | -0.04989 | -0.31247 | -0.41363 | -0.20312 | -0.43509 | -1.28594 | -0.06358 | -0.40793 | -0.85532 | -1.11196 | -0.29374 | -0.53133 | -0.16375 | 0.826663 | -0.42139 | -0.19907 | -0.09459 | 0.203559 | 2.303481 | 5.588577 | -0.00213 | 0.287899 | 0.383314 | -0.34168 | 1.192732 | -0.39697 | 0.567077 | -0.53033 | -0.62006 | -0.18698 | [illegible] |
| 689 | -0.51735 | -0.43426 | -0.08063 | 0.532513 | -0.16782 | -0.07894 | 1.060063 | -0.38461 | 0.763201 | -0.07049 | -0.09831 | 0.126753 | -0.06089 | 0.032572 | -0.68672 | -1.04103 | -0.27566 | -0.3039 | -1.13507 | -0.82159 | -0.12285 | -0.60117 | 0.162344 | 0.841238 | -0.58867 | 0.06954 | -0.01309 | 0.261742 | 1.299441 | 4.207493 | -0.06476 | 0.42086 | -0.04149 | -0.47325 | 1.189488 | -0.60785 | 0.423054 | -0.48665 | -0.38294 | -0.53654 | [illegible] |
| 690 | -0.63724 | -0.74074 | -0.4496 | 0.454061 | -0.20279 | -0.23271 | 0.111281 | -0.11132 | -0.62105 | 0.138805 | -0.00035 | 0.159001 | -0.71947 | -0.31816 | -0.5674 | -1.07636 | -0.334 | -0.39792 | -1.21169 | -0.69737 | -0.17412 | -0.4547 | 0.279278 | 0.266426 | -0.83906 | -0.02522 | -0.49415 | 0.078096 | -0.36132 | 1.310703 | -0.559 | 0.272402 | -0.66914 | -0.43034 | -0.12257 | -1.023 | 0.471695 | -0.36248 | -0.36242 | -0.86583 | [illegible] |
| 691 | -0.69498 | -0.88912 | -0.60795 | 0.043596 | -0.53607 | -1.28937 | -0.09969 | -0.43978 | -1.08789 | 0.246431 | -0.47666 | -0.05721 | -0.6102 | -0.34923 | -0.53245 | -0.83611 | -0.45771 | -0.13959 | -0.88633 | -0.59785 | -0.27496 | -0.12864 | -0.17607 | 0.132989 | -0.98467 | -0.5037 | -0.82359 | -0.65612 | -0.49693 | -0.31095 | -0.59688 | 0.024425 | -0.24749 | -0.23069 | -0.86631 | -0.57453 | 0.059802 | -0.24029 | -0.00828 | -0.40114 | [illegible] |
| 692 | -0.42812 | -1.16784 | -0.95035 | -0.28269 | -0.02348 | -1.46235 | -0.63751 | -0.39007 | -0.50094 | 0.29055 | -0.64129 | -0.24887 | -0.40635 | -0.46916 | -0.77407 | -0.74525 | -0.53864 | -0.34238 | -0.89342 | -0.85371 | -0.21025 | -0.42179 | 0.222392 | 0.134618 | -0.50967 | -0.41612 | -0.59757 | -0.61092 | -0.07153 | -0.50409 | -0.53664 | -0.38159 | -0.26994 | -0.31873 | -0.71705 | -0.65796 | -0.16679 | -0.85956 | -0.34616 | -0.43096 | [illegible] |
| 693 | -0.49239 | -0.92047 | -0.48959 | -0.77389 | 0.178925 | -1.19554 | -0.98812 | 0.000178 | -0.21273 | 0.617049 | -0.60598 | -0.2649 | -0.4818 | -0.6589 | -0.84785 | -0.68468 | -0.87377 | -0.1352 | -0.86531 | -0.78934 | -0.45184 | -0.44044 | 0.466845 | 0.095876 | -0.47652 | -0.66278 | -0.61928 | -0.73381 | 0.122503 | -0.22522 | -0.61798 | -0.38143 | 0.24822 | -0.36462 | -0.51379 | -0.71841 | -0.90101 | -0.86184 | -0.41258 | -0.25812 | [illegible] |
| 694 | -0.39782 | -1.03705 | -0.98883 | -0.90493 | -0.15421 | -1.36406 | -0.74164 | -0.05911 | -0.1555 | 0.092732 | -0.76238 | -0.69043 | -0.45792 | -0.87322 | -0.67884 | -0.92483 | -0.75721 | -0.04449 | -0.67419 | -1.20226 | -0.38284 | -0.24011 | 0.325101 | 1.090875 | -0.6141 | -0.75117 | -0.74502 | -0.62195 | 0.133042 | 0.122986 | -0.84115 | -0.6293 | -0.15274 | -0.18707 | -0.64169 | -0.45329 | -0.87904 | -1.00274 | -0.61156 | -0.15098 | [illegible] |
| 695 | -0.50323 | -0.73373 | -0.67282 | -0.67542 | -0.26551 | -1.59857 | -0.69073 | -0.00306 | 0.155652 | 0.134026 | -0.65391 | -0.93811 | -0.30975 | -0.68839 | -0.81378 | -0.74133 | -0.43566 | 0.52773 | -0.40193 | -1.12216 | -0.42578 | -0.40648 | 0.208905 | 1.862495 | -0.51185 | -0.65077 | -0.76782 | -0.22579 | 0.517438 | 0.389743 | -0.82355 | -0.39281 | -0.01456 | 0.691166 | -0.63565 | -0.16769 | -0.8263 | -0.59923 | -0.21503 | 0.187545 | [illegible] |
| 696 | -0.17976 | -0.44603 | -0.76011 | -0.02147 | -0.27492 | -0.84899 | -0.98242 | 0.301479 | 0.457876 | 0.180948 | -0.49143 | -0.83146 | -0.20895 | -0.99321 | -0.81065 | -0.40468 | -0.27075 | 0.491345 | -0.3232 | -1.50682 | -0.53274 | -0.67777 | 0.612005 | 2.191655 | -0.34807 | -0.30674 | -0.48767 | -0.08202 | 0.577536 | 0.088018 | -0.95372 | -0.04032 | -0.08791 | 1.609243 | -0.3399 | 0.279521 | -0.4896 | -0.68509 | -0.3227 | -0.56542 | [illegible] |
| 697 | -0.28913 | -0.31025 | -0.63254 | -0.17375 | -0.68053 | -0.70758 | -0.37582 | 0.457183 | 0.090452 | 0.180948 | -0.41986 | -0.67198 | -0.53659 | -1.29852 | -0.56903 | -0.15914 | 0.203715 | 0.884779 | -0.1851 | -1.1554 | -0.53274 | -0.53707 | -0.07211 | 2.108297 | -0.40598 | -0.39054 | 0.387142 | -0.13253 | 0.133008 | 0.252484 | -0.95372 | 0.238937 | -0.1354 | 1.877033 | -0.3399 | 0.550803 | -0.12371 | -0.03305 | -0.08264 | -0.60294 | [illegible] |
| 698 | -0.44314 | -0.53437 | -0.76052 | 0.381423 | -0.98819 | -0.83766 | -0.16816 | 0.200989 | -0.23571 | -0.24387 | -0.58288 | -0.65251 | -0.50013 | -0.83362 | -0.50856 | -0.06214 | 0.277739 | 0.739379 | -0.31221 | -0.7756 | -0.3062 | -0.68863 | -0.04506 | 2.309009 | -0.4855 | -0.17361 | 0.762226 | 0.557095 | -0.07298 | -0.05143 | -0.99618 | -0.04793 | -0.32899 | 2.270567 | -0.3399 | 0.861742 | 0.109821 | 0.255769 | 0.128355 | -0.68194 | [illegible] |
| 699 | -0.28101 | -0.81034 | -0.70871 | 0.332167 | -0.4836 | -0.76849 | -0.26989 | 0.078223 | -0.32567 | -0.16918 | -0.35856 | -0.43278 | -0.87755 | -0.64648 | -0.68831 | 0.100379 | 0.34457 | 0.544633 | -0.17128 | -0.69321 | -0.21691 | -0.90472 | -0.31438 | 0.964972 | 0.007687 | -0.17361 | 0.536225 | 0.418489 | -0.07611 | -0.14689 | -0.56375 | 0.032278 | -0.20139 | 2.176133 | -0.03065 | 1.01674 | 0.034349 | 0.068974 | 0.311888 | -0.53496 | [illegible] |
| 700 | -0.01728 | -0.95793 | -0.68769 | 0.290936 | -0.33733 | -0.77333 | -0.21036 | -0.14093 | -0.21807 | -0.46928 | -0.28649 | 0.121703 | -1.29732 | -0.28186 | -0.53765 | -0.01353 | -0.035 | -0.22358 | -0.24797 | -0.70273 | -0.21366 | -0.73458 | -0.1551 | 0.079457 | -0.07029 | -0.02624 | 0.553371 | 0.226529 | -0.27951 | -0.43289 | -0.23708 | 0.030241 | -0.32824 | 1.422026 | 0.002358 | 0.84736 | -0.32637 | -0.31017 | -0.00901 | -0.5442 | [illegible] |
| 701 | -0.40641 | -0.95793 | -0.4743 | -0.19296 | -0.31446 | -0.6437 | -0.1344 | -0.13001 | -0.32949 | -0.75429 | -0.30113 | 0.330153 | -1.42308 | -0.06195 | -0.48966 | -0.57315 | -0.1641 | -0.35247 | -0.37657 | -0.25741 | -0.22384 | -0.69612 | -0.18257 | -0.5322 | -0.36147 | -0.34524 | 0.292316 | 0.097603 | -0.45398 | -0.41442 | -0.37305 | -0.29601 | -0.5742 | 0.460618 | -0.25484 | 0.396422 | -0.65144 | -0.22081 | -0.17447 | -0.27751 | [illegible] |
| 702 | -0.18585 | -0.87044 | -0.38507 | -0.16181 | -0.29982 | -0.45274 | -0.44943 | -0.20331 | -0.27677 | -0.75429 | -0.40129 | 0.291389 | -1.2438 | 0.243362 | -0.25328 | -0.54526 | -0.6113 | -0.46825 | -0.23289 | -0.39112 | -0.18296 | -0.84355 | -0.61879 | -1.03582 | -0.33693 | -0.4698 | -0.78213 | 0.132278 | -0.23669 | -0.24216 | -0.42737 | -0.66227 | -0.38771 | -0.02237 | -0.48507 | 0.124237 | -1.02737 | -0.51316 | -0.14961 | -0.01709 | [illegible] |
| 703 | -0.26801 | -0.7934 | -0.5101 | -0.01692 | -0.45629 | -0.52064 | -1.0463 | -0.35689 | -0.28868 | -0.5286 | -0.23827 | -0.06163 | -1.15894 | -0.30383 | -0.37728 | -1.36533 | -0.94384 | -0.71118 | -0.5261 | -0.67058 | -0.48021 | -0.64044 | -0.49222 | -1.09353 | -0.50754 | -0.37115 | -0.96264 | -0.62046 | -0.02077 | -0.24216 | -0.29736 | -0.3754 | -0.29102 | -0.61846 | -0.77679 | -0.19178 | -1.02737 | -0.9978 | -0.39755 | -0.02159 | [illegible] |
| 704 | -0.21751 | -0.32257 | -0.13858 | 0.110217 | -0.31458 | -0.39978 | -1.09327 | -0.35294 | -0.58413 | -0.61054 | -0.11744 | -0.41435 | -1.13853 | -0.31534 | -0.24996 | -1.38552 | -1.09848 | -0.86326 | -0.55119 | -0.53449 | -0.53814 | -0.55231 | -0.18478 | -0.88044 | -0.25353 | -0.51706 | -0.7968 | -0.79333 | -0.27302 | -0.21837 | -0.39689 | -0.50421 | -0.44535 | -0.57071 | -0.5205 | -0.45917 | -0.73594 | -0.76529 | -0.50799 | -0.18915 | [illegible] |
| 705 | 0.020891 | -0.27647 | -0.03194 | -0.38957 | -0.39668 | -0.15249 | -0.9929 | -0.58404 | -0.37068 | -0.28452 | -0.11898 | -0.69093 | -0.86731 | -0.47563 | -0.39419 | -1.14048 | -1.01076 | -0.56295 | -0.46226 | -0.66878 | -0.43098 | -0.69535 | -0.35889 | -0.72856 | -0.31922 | -0.63544 | -1.04866 | -1.08671 | -0.22638 | -0.03502 | -0.36136 | -0.50421 | -0.60732 | -0.82941 | -0.68264 | -0.42469 | -0.4797 | -1.12193 | -0.67635 | -0.25378 | [illegible] |
| 706 | -0.02441 | -0.54101 | 0.155382 | -0.46707 | -0.01862 | -0.27103 | -0.96225 | -0.58404 | -0.55497 | -0.32072 | -0.45886 | -0.969 | -0.63886 | -0.63797 | -0.39448 | -0.95708 | -1.10954 | -0.53257 | -0.20792 | -1.11139 | -0.46527 | -0.69865 | -0.37321 | -0.75273 | -0.08848 | -0.18348 | -0.75295 | -0.84722 | -0.33979 | 0.12373 | -0.08938 | -0.48237 | -0.36136 | -0.93132 | -0.24397 | -0.67424 | -0.21026 | -1.24759 | -0.23459 | -0.23218 | [illegible] |
| 707 | 0.011297 | 0.115441 | 0.121256 | -0.66969 | 0.00735 | -0.22063 | -1.01715 | -0.66602 | -0.45024 | -0.24077 | 0.039283 | -0.87236 | -0.60953 | -0.96901 | -0.70038 | -1.02474 | -0.94593 | -0.77553 | -0.24805 | -1.13458 | -0.55828 | -0.36327 | 0.180056 | -0.66284 | -0.08885 | -0.15217 | -0.71855 | -1.1689 | -0.39265 | -0.17938 | 0.244922 | -0.59627 | -0.11032 | -0.49956 | 0.108813 | -0.38356 | -0.22224 | -1.22707 | 0.04876 | -0.21682 | [illegible] |
| 708 | 0.312844 | 0.488025 | 0.110451 | -1.57026 | 0.120721 | -0.24619 | -0.06194 | -0.77405 | -0.3435 | -0.19936 | -0.35122 | -0.54141 | -0.30748 | -0.70597 | -0.44582 | -0.34155 | -0.4565 | -0.70454 | -0.01526 | -0.92621 | -0.11811 | -0.16958 | -0.28068 | -0.82107 | -0.18294 | -0.1231 | -0.41044 | -0.89809 | -0.76598 | 0.022859 | 0.255919 | -0.70666 | -0.03966 | -0.46947 | 0.289573 | -0.22234 | -0.12787 | -0.72724 | 0.129722 | -0.46596 | [illegible] |
| 709 | -0.0021 | 0.961565 | -0.12382 | -1.52741 | -0.32904 | -0.33884 | -0.46648 | -0.71282 | -0.02596 | 0.107792 | -0.46367 | -0.42347 | -0.32755 | -0.99016 | -0.33739 | -0.26778 | -0.35843 | -0.8399 | 0.202656 | -0.92604 | -0.05771 | 0.47788 | -0.32614 | -0.7921 | -0.84998 | 0.00844 | -0.30603 | -0.62056 | -0.7302 | -0.02681 | 0.208821 | -0.64851 | 0.256427 | -0.31651 | -0.17833 | -0.11683 | -0.0778 | -0.94499 | 0.267909 | -0.50674 | [illegible] |
| 710 | -0.29989 | 1.063625 | -0.42204 | -1.32415 | -0.23912 | -0.5503 | -0.47698 | -0.52926 | -0.51059 | -0.07823 | -0.58546 | -0.31559 | -1.03887 | -1.22498 | -0.39272 | -0.37546 | -0.36622 | -0.86985 | -0.1191 | -0.79175 | -0.10777 | 0.697062 | -0.15994 | -1.07345 | -0.55817 | 0.185552 | -0.1336 | -0.34774 | -0.73819 | -0.02563 | 0.221327 | -0.64851 | 0.229718 | -0.05781 | -0.01619 | 0.092838 | -0.01226 | -0.90088 | 0.509873 | -0.64168 | [illegible] |
| 711 | -0.39395 | 1.41766 | -0.65547 | -1.15898 | -0.35287 | -0.50832 | -0.65572 | -0.70352 | -0.09892 | -0.22596 | -0.50234 | -0.32501 | -1.20763 | -0.81345 | -0.29811 | -0.21921 | -0.58563 | -0.73422 | -0.36095 | -0.5393 | 0.043666 | 1.120544 | -0.46589 | -0.65321 | -0.51954 | -0.11361 | -0.30522 | -0.59266 | -0.95919 | 0.140117 | -0.03336 | -0.57389 | 0.031553 | 0.066705 | -0.23862 | 0.107841 | -0.27727 | -0.68811 | 0.137817 | -0.73881 | [illegible] |
| 712 | -0.82775 | 0.929376 | -0.62135 | -1.10986 | -0.14602 | -0.72934 | -0.43693 | -0.62153 | -0.38273 | -0.11969 | -0.79716 | -0.43847 | -1.18843 | -0.73234 | -0.49072 | -0.01996 | -0.49539 | -0.7811 | -0.44727 | -0.91422 | 0.256225 | 1.025111 | -0.197 | -0.43546 | -0.4858 | -0.1656 | 0.020952 | -0.25514 | -1.08028 | 0.41262 | -0.25442 | -0.22299 | -0.21949 | -0.33744 | -0.36118 | 0.020376 | -0.02674 | -0.69899 | -0.23002 | -0.75417 | [illegible] |
| 713 | -0.79273 | 0.850149 | -0.45928 | -0.42121 | -0.10291 | -0.5058 | -0.85995 | -0.62075 | -0.9422 | -0.38591 | -0.70632 | -0.72805 | -1.65241 | -0.64724 | -0.854 | 0.019917 | -0.53634 | -0.93125 | -0.6674 | -0.88173 | 0.087693 | 0.831425 | 0.186005 | -0.24756 | -0.46559 | -0.26358 | -0.5038 | -0.25809 | -0.68283 | 0.210381 | -0.35325 | -0.11261 | -0.38684 | -0.16498 | -0.25022 | -0.3191 | -0.12111 | -0.8101 | -0.1649 | -0.49316 | [illegible] |
| 714 | -0.65509 | 0.377884 | -0.50049 | -0.79856 | -0.14133 | -0.48674 | -0.36505 | -1.26052 | -0.8019 | -0.23937 | -0.45615 | -0.7122 | -1.48421 | -0.684 | -1.12598 | -0.28487 | -0.52165 | -0.85171 | -0.99852 | -0.78267 | 0.229714 | 0.1488 | 0.025809 | -0.4814 | -0.15067 | -0.53049 | -0.51115 | -0.25809 | -0.52502 | 0.054471 | -0.26452 | 0.045084 | -0.38684 | -0.20093 | -0.37618 | -0.54432 | -0.28934 | -0.59077 | -0.43137 | -0.18668 | [illegible] |
| 715 | -0.74066 | 0.581782 | -0.26407 | -0.68461 | -0.09285 | -0.09556 | -0.65997 | -1.73776 | -0.66947 | -0.39802 | -0.71383 | -0.8382 | -0.58435 | -0.73635 | -0.83632 | -0.08944 | -0.36828 | -0.83205 | -1.0412 | -0.9696 | 1.134221 | 0.184307 | 0.033716 | -0.6171 | -0.46841 | -0.81531 | -0.65262 | -0.87857 | -0.4881 | -0.15162 | -0.57042 | 0.046593 | -0.2896 | -0.27506 | -0.37618 | -0.7575 | -0.27345 | -0.39697 | -0.63007 | -0.24524 | [illegible] |
| 716 | -0.00705 | 0.442693 | -0.02641 | -0.6256 | -0.19663 | 0.223652 | -0.70216 | -1.74158 | -0.8061 | -0.41445 | -0.41393 | -0.61247 | 0.007958 | -0.86956 | -0.96598 | -0.1188 | -0.42072 | -1.16046 | -0.95068 | -0.56794 | 1.667808 | 0.051598 | 0.256018 | -1.0568 | -0.55129 | -1.20069 | -0.48099 | -0.69733 | -0.45516 | -0.46178 | -0.32156 | 0.238532 | 0.036934 | -0.31359 | -0.61323 | -0.7353 | -0.0681 | -0.62462 | -0.36082 | -0.4176 | [illegible] |
| 717 | 0.366469 | 0.25272 | -0.18885 | -0.72216 | -0.13687 | 0.965243 | -0.88957 | -1.44894 | -0.65872 | -0.9047 | -0.5539 | -0.79503 | -0.02949 | -0.54182 | -0.75122 | -0.44339 | -0.42072 | -0.64251 | -0.99672 | -0.30396 | 2.27195 | 0.250485 | -0.03658 | -1.21504 | -0.58202 | -0.45072 | -0.54511 | -0.50242 | 0.023369 | -0.86004 | -0.20413 | 0.063127 | 0.140445 | -0.25691 | -0.78033 | -0.82776 | -0.0681 | -0.09561 | -0.27632 | -0.22975 | [illegible] |
| 718 | 0.20268 | 0.13873 | -0.18815 | -0.82467 | -0.24537 | 0.860859 | -0.78815 | -1.436 | -0.82006 | -0.53276 | -0.07871 | -0.65913 | 0.106476 | -0.83952 | -0.78951 | -0.58129 | -0.10315 | -0.38045 | -0.88939 | -0.63295 | 2.523101 | 0.172739 | -0.16835 | -1.45988 | -0.33753 | -0.39998 | -0.5308 | -0.48708 | -0.11495 | -0.73837 | -0.09801 | 0.051436 | 0.453047 | -0.10163 | -0.92086 | -0.55443 | 0.067933 | 0.046821 | 0.11505 | -0.18186 | [illegible] |
| 719 | 0.362247 | -0.13384 | 0.127757 | -0.61741 | 0.102215 | 0.408974 | -0.73925 | -0.80447 | -0.74513 | -0.72872 | -0.48703 | -0.7541 | -0.13495 | -0.81628 | -0.54593 | -0.58004 | -0.24422 | -0.49198 | -0.72091 | -0.90539 | 2.138103 | 0.132818 | -0.22212 | -1.24886 | -0.69673 | -0.22235 | -0.1142 | -0.73443 | -0.31579 | -0.46831 | -0.00392 | -0.0356 | 0.432919 | -0.09124 | -0.58919 | -0.59875 | 0.015539 | 0.096729 | 0.236651 | -0.09917 | [illegible] |
| 720 | 0.187184 | -0.27066 | -0.12908 | -0.76191 | 0.045917 | -0.00165 | -0.46459 | -0.19399 | -0.96028 | -0.40159 | -0.07834 | -0.39579 | -0.51556 | -0.5267 | -0.89322 | -0.77192 | -0.44916 | -0.50773 | -0.63913 | -0.71846 | 0.952891 | 0.047391 | -0.22212 | -0.99222 | -0.65406 | -0.30482 | 0.321841 | -0.36898 | -0.13856 | -0.09732 | 0.233849 | -0.14422 | 0.273125 | -0.01711 | -0.6459 | -0.66868 | -0.28633 | 0.15918 | 0.36773 | 0.182114 | [illegible] |
| 721 | -0.54403 | -0.22108 | -0.24092 | -1.03726 | 0.219919 | -0.21801 | 0.108322 | -0.00723 | -1.22393 | -0.04423 | -0.68449 | -0.57934 | -0.89383 | -0.512 | -0.60784 | -0.77192 | -0.40912 | -0.16709 | -0.80451 | -1.10013 | 0.164307 | -0.20705 | -0.17681 | -0.6737 | -0.82174 | 0.16564 | 0.396023 | -0.31977 | 0.077411 | 0.438618 | 0.193816 | -0.43396 | 0.118156 | -0.00118 | -0.60431 | -0.44513 | -0.22667 | 0.035499 | 0.642615 | 0.611359 | [illegible] |
| 722 | -0.9208 | -0.43503 | -0.30427 | -0.94639 | 0.101116 | -0.78898 | 0.137792 | -0.29987 | -1.37967 | 0.259804 | -0.23543 | -0.52657 | -1.24707 | -0.86827 | -0.60817 | -0.8467 | -0.61834 | -0.3952 | -0.63202 | -0.9704 | -0.86597 | -0.44236 | -0.03608 | -0.57381 | -0.84375 | -0.35529 | -0.16977 | -0.73837 | -0.42897 | 0.510509 | 0.017474 | -0.40854 | -0.19342 | 0.129724 | -0.33633 | -0.03715 | -0.22667 | 0.058823 | 0.711311 | 0.423504 | [illegible] |
| 723 | -0.66033 | -1.06943 | -0.30427 | -0.67718 | 0.363281 | -0.77657 | 0.060941 | -0.30356 | -0.76341 | 0.060081 | -0.53509 | -0.11542 | -1.37435 | -0.86865 | -0.32293 | -0.70879 | -0.65856 | -0.10707 | -0.4588 | -0.77681 | -0.83872 | -0.86249 | -0.33491 | -0.53047 | -0.84375 | -0.40604 | 0.186084 | -0.74054 | -0.15432 | 0.621183 | 0.132589 | -0.71288 | -0.50602 | -0.02555 | -0.1958 | -0.44984 | -0.4017 | 0.242829 | 0.231141 | 0.363738 | [illegible] |
| 724 | -0.93386 | -0.87809 | -0.76552 | -0.55812 | 0.357769 | -0.34486 | -0.13231 | -0.4702 | -1.19338 | 0.329686 | -0.60448 | 0.004272 | -1.18232 | -0.69214 | -0.6403 | -0.53125 | -0.80518 | 0.180776 | -0.26224 | -0.51605 | -0.31784 | -0.6903 | -0.14091 | -0.27799 | -0.38954 | -0.3024 | -0.39607 | -0.58401 | 0.525458 | 0.284812 | -0.05023 | -0.76557 | -0.49282 | -0.07856 | -0.54667 | -0.14314 | -0.61976 | 0.403736 | 0.027974 | -0.08728 | [illegible] |
| 725 | -0.90715 | -0.73016 | -0.87248 | -0.37751 | -0.05062 | -0.344 | -0.46436 | -0.68638 | -0.60566 | 0.339622 | -0.53627 | -0.55041 | -0.89138 | -0.63125 | -0.43272 | -0.59842 | -0.39505 | -0.04696 | -0.30987 | -0.06354 | -0.38201 | -1.15972 | -0.37147 | -0.10541 | -0.06627 | -0.31424 | -0.6352 | -0.28909 | 0.667484 | -0.02004 | -0.02245 | -0.64423 | -0.32444 | -0.08697 | -0.59319 | 0.079655 | -0.34136 | 0.439399 | -0.55284 | -0.33512 | [illegible] |
| 726 | -0.39485 | -0.82107 | -1.32319 | 0.140076 | -0.37619 | -0.33313 | -0.49942 | -1.0322 | -0.03039 | 0.108026 | -0.18504 | -0.51546 | -1.00091 | -0.88677 | -0.50559 | -0.85259 | -0.23808 | -0.5802 | 0.395419 | 0.819902 | -0.23503 | -1.20986 | -0.51136 | 0.033092 | 0.120203 | -0.1122 | -0.72211 | 0.017731 | 1.187416 | -0.40399 | 0.132251 | -0.67904 | -0.02614 | -0.27707 | -0.73513 | -0.08504 | -0.38503 | 0.195714 | -0.59965 | -0.37501 | [illegible] |
| 727 | -0.32533 | -0.62498 | -1.24255 | 1.237786 | 0.117828 | -0.37857 | -0.45237 | -1.12653 | 0.3335 | 0.396824 | 0.126367 | -0.3125 | -1.33035 | -0.49383 | -0.64751 | -0.6039 | 0.092872 | -0.8195 | -0.0099 | 0.354041 | 0.012168 | -1.31633 | -0.63822 | 0.129407 | 0.388102 | -0.02506 | -0.13864 | 0.520646 | 1.140915 | -0.02867 | 0.465446 | -0.52771 | -0.24104 | -0.14842 | -0.99082 | -0.33808 | -0.37833 | 0.1614 | -0.70158 | -0.38288 | [illegible] |
| 728 | -0.49552 | -0.21522 | -1.17196 | 1.279693 | -0.06822 | -0.49006 | -0.5459 | -0.78007 | 0.375881 | 1.355328 | 0.07529 | -0.78687 | -1.30846 | -0.10636 | -0.76823 | -0.87957 | -0.15485 | -1.35906 | -0.14177 | 0.397102 | -0.18899 | -1.10833 | -0.5107 | 0.225291 | -0.15535 | -0.23395 | -0.39839 | 0.499227 | 0.923366 | 0.179293 | 0.439667 | -0.23436 | -0.4394 | -0.4421 | -1.39219 | -0.12422 | -0.32938 | -0.16083 | -0.77736 | -0.76704 | [illegible] |
| 729 | -0.46745 | -0.29117 | -0.84118 | 1.183104 | -0.35626 | -0.75413 | -0.34992 | -1.25979 | 0.376893 | 1.032421 | -0.27587 | -1.05999 | -1.27975 | 0.072362 | -0.45086 | -0.86842 | 0.159979 | -1.33769 | -0.42119 | 0.314846 | -0.62763 | -1.18278 | -0.39729 | -0.05492 | -0.48165 | -0.50099 | -0.34991 | 0.583985 | 0.080479 | 0.109655 | 0.228094 | -0.03357 | -0.47986 | -0.11957 | -1.49844 | -0.23926 | -0.02428 | -0.4562 | -0.20874 | -0.82386 | [illegible] |
| 730 | -0.51633 | -0.71507 | -0.35525 | 0.734192 | -0.70514 | -0.73801 | -0.35177 | -1.18196 | 0.404047 | 0.881509 | -0.85424 | -1.32981 | -2.18651 | -0.40244 | -0.46655 | -0.57737 | -0.18242 | -1.64535 | -0.5613 | -0.25923 | -0.71223 | -0.73958 | -0.38513 | -0.15536 | -0.52985 | -0.18674 | -0.34769 | 0.520762 | 0.073233 | 0.396532 | 0.346366 | -0.65348 | -0.93089 | 0.354964 | -1.15628 | -0.01226 | -0.01661 | -0.37949 | 0.392945 | -0.51545 | [illegible] |
| 731 | -1.46012 | -0.54354 | 0.026643 | 0.473091 | -0.58563 | -0.68289 | -0.38082 | -1.22268 | 0.135203 | 0.874296 | -0.33587 | -0.94387 | -2.0678 | -0.30829 | -0.39367 | -0.4323 | -0.02224 | -0.71524 | -1.70018 | -0.96127 | -1.00829 | -0.48964 | -0.30287 | -0.28808 | -1.09071 | -0.27762 | -0.50667 | 0.247634 | -0.37236 | 0.452007 | 0.678997 | -0.19237 | -1.14473 | 0.780005 | -0.80447 | 0.141236 | -0.28316 | 0.162358 | -0.29794 | -0.93636 | [illegible] |
| 732 | -1.09537 | -0.42986 | -0.22697 | -0.18411 | -0.87528 | 0.079105 | -0.55965 | -1.26108 | 0.194022 | 0.639793 | -1.0765 | -0.90383 | -1.51779 | -0.48018 | -0.54641 | -0.65029 | -0.38484 | 0.30311 | -1.17411 | -0.41802 | -0.71445 | -0.40857 | -0.24317 | -0.71849 | -1.11633 | 0.05898 | -0.63333 | -0.23335 | -0.23779 | 0.449854 | 0.245387 | -0.33463 | -0.84953 | 0.470347 | -0.82528 | -0.07834 | -0.1612 | -0.04656 | -0.49536 | -1.05122 | [illegible] |
| 733 | -1.30616 | -0.41026 | -0.20673 | -0.48872 | -0.83714 | 0.441867 | -0.2809 | -1.74233 | 0.000687 | -0.61254 | -0.9013 | -0.55118 | -1.35793 | -0.58732 | -0.3304 | -0.376 | -0.58121 | 1.764945 | -0.88728 | -0.61697 | -1.01671 | -0.16306 | 0.026454 | -0.585 | -0.47246 | 0.242764 | -0.44309 | -0.34212 | -0.49046 | -0.18619 | 0.46236 | -0.583 | -0.73668 | 0.880012 | -0.60641 | -0.02676 | -0.30519 | 2.529768 | -0.40792 | -0.38512 | [illegible] |
| 734 | -1.23197 | -0.58984 | -0.36471 | -0.49827 | -0.63578 | 1.042486 | -0.06441 | -0.9796 | -0.15117 | -0.55184 | -0.70437 | -0.09194 | -1.27794 | -0.63299 | -0.26795 | 0.002846 | -0.5273 | 1.896914 | -0.76189 | -0.62469 | -0.67734 | -0.35645 | -0.19594 | -0.67414 | -0.33139 | 0.759293 | -0.12806 | -0.51679 | 0.109704 | 0.131538 | 0.779775 | -0.83304 | -0.6893 | 0.316182 | -0.1215 | 0.267492 | -0.44005 | 3.203402 | -0.87929 | -0.05811 | [illegible] |
| 735 | -1.16387 | -0.16466 | -0.46357 | -0.10935 | -0.15926 | 1.145262 | 0.359772 | -0.63885 | -0.5093 | -0.51392 | 0.287198 | 0.328705 | -0.66526 | -0.5451 | -0.1025 | -0.40368 | -0.196 | 2.13641 | -0.5655 | -0.50167 | -0.57806 | -0.30725 | 0.022457 | -0.61502 | -0.37819 | 0.439148 | -0.33804 | -0.00969 | -0.23923 | -0.26685 | 1.0467 | -0.20978 | -0.25639 | -0.29297 | -0.22283 | -0.08671 | -0.36202 | 2.901004 | -1.10566 | -0.4319 | [illegible] |
| 736 | -0.4458 | -0.18855 | 0.23557 | -0.37478 | -0.38983 | 0.900907 | 0.371699 | -0.57575 | -0.42742 | -0.13739 | 0.374752 | 0.139358 | -0.49294 | -0.4186 | -0.24888 | 0.342595 | -0.27443 | 1.588585 | -0.00435 | -0.11306 | -0.28078 | -0.6861 | 0.537808 | -0.65144 | 0.156795 | 0.136485 | 0.688106 | 0.125906 | -0.17265 | 0.196334 | 0.967805 | -0.45905 | -0.39093 | -0.33491 | -0.13394 | -0.21684 | -0.01244 | 2.84608 | -0.77606 | -0.37777 | [illegible] |
| 737 | -0.49636 | -0.44515 | 0.964909 | -0.83223 | -0.35442 | 0.341373 | 0.65831 | -0.29691 | -0.23658 | -0.11594 | 0.86464 | 0.44177 | -0.40148 | -0.56008 | 0.165772 | 0.928655 | -0.01946 | 0.979696 | 0.08429 | 0.494151 | -0.66456 | -0.94631 | 0.309545 | -0.25178 | 0.063935 | -0.21565 | 1.330834 | 0.342202 | -0.06689 | -0.22253 | 0.945281 | -0.10023 | -0.3495 | -0.18351 | 0.132519 | 0.031745 | -0.22521 | 2.596526 | -0.86816 | -0.20671 | [illegible] |
| 738 | 0.072216 | -0.58229 | 0.874079 | -0.83566 | -0.45535 | -0.05979 | 0.568394 | 0.159488 | -0.04119 | 0.090564 | 0.544829 | 0.104117 | -0.58968 | -0.86462 | 0.216833 | 0.594282 | 0.214898 | -0.47455 | -0.27453 | 0.412294 | -0.72521 | -1.0334 | 0.06391 | -0.54781 | -0.0469 | -0.19134 | 1.627148 | 0.180122 | -0.02844 | 0.072653 | 0.874128 | 0.170821 | -0.51406 | -0.12754 | 0.4766 | 0.093089 | 0.02591 | -0.16538 | -1.12518 | -0.79467 | [illegible] |
| 739 | -0.00058 | -0.274 | 0.735687 | -1.14902 | -0.81713 | -0.81954 | 0.354495 | -0.47806 | 0.382284 | 0.012115 | 0.890338 | -0.04781 | -0.8473 | -1.16657 | 0.02266 | -0.10709 | -0.0523 | -0.34808 | -0.33613 | 0.700406 | -1.17744 | -0.85511 | 0.252742 | -0.67131 | -0.32206 | -0.41694 | 1.336263 | 0.089631 | -0.16344 | -0.16001 | 0.83401 | 0.272769 | -0.54169 | 0.115707 | 0.538817 | -0.45181 | 0.340975 | -0.51851 | -1.06703 | -1.0798 | [illegible] |
| 740 | 0.087852 | -0.55734 | 0.727836 | -1.11696 | -0.88286 | -0.59019 | 0.361249 | -0.78085 | 0.450473 | -0.00255 | 0.126405 | -0.15744 | -0.84782 | -1.11101 | -0.11355 | -0.07252 | -0.37927 | 0.20601 | -0.36994 | 0.913295 | -1.24576 | -1.2832 | 0.354284 | -0.45993 | -0.49736 | -0.19582 | 1.744254 | -0.4837 | -0.28471 | -0.09083 | 0.290135 | 0.255169 | -0.41303 | 0.167506 | 0.621915 | -0.4431 | 0.109947 | -0.72812 | -0.93949 | -0.71218 | [illegible] |
| 741 | -0.05107 | -0.81249 | 0.751403 | -1.06583 | -0.65086 | -0.65927 | 0.022465 | -0.48296 | -0.02966 | -0.26031 | -0.57766 | -0.45863 | -1.00572 | -1.22169 | -0.20144 | -0.44998 | -0.48433 | 0.136402 | -0.62733 | 0.447209 | -1.36863 | -0.69417 | -0.13063 | -0.13752 | -0.21035 | -0.00433 | 0.899852 | -0.64629 | -0.5427 | -0.60837 | -0.37481 | 0.23163 | -0.70908 | -0.09111 | 0.630493 | -0.34609 | 0.359311 | -0.5547 | -0.98215 | -0.60098 | [illegible] |
| 742 | -0.31314 | -0.80524 | 0.474401 | -0.81623 | -0.99278 | -0.99891 | 0.113044 | -0.61875 | -0.31908 | -0.69155 | -1.0378 | -1.08589 | -1.23769 | -0.9191 | -0.24078 | -0.81283 | -0.44455 | -0.15122 | -0.98351 | -0.18955 | -1.64236 | -0.40855 | -0.32237 | -0.18703 | 0.115933 | 0.224799 | 0.638547 | -0.64286 | -0.70041 | -0.39166 | -0.4569 | 0.308658 | -0.57475 | -0.27603 | 0.486423 | -0.3204 | 0.086284 | -0.58064 | -0.56736 | -0.73425 | [illegible] |
| 743 | -0.59744 | -0.51045 | 0.258429 | -0.75575 | -0.79281 | -0.85186 | 0.14659 | -0.5628 | 0.003 | -0.79376 | -1.38168 | -1.31309 | -1.18439 | -0.50845 | -0.33477 | -0.57479 | -0.53062 | 0.338068 | -1.06365 | 0.15702 | -1.43891 | -0.47033 | -0.0845 | 0.054815 | -0.11824 | 0.22559 | 0.337024 | -0.60541 | -0.76685 | -0.47883 | -0.6519 | 0.020157 | -0.32468 | -0.70323 | 0.053748 | -0.23396 | -0.08847 | -0.30334 | -0.23321 | -0.68234 | [illegible] |
| 744 | -0.59812 | -0.7613 | 0.275865 | -0.7153 | -0.59531 | -0.77141 | 0.14659 | -0.04981 | -0.09166 | -0.93108 | -1.36005 | -1.31299 | -1.14377 | -0.49408 | -0.16379 | -0.39093 | -0.32136 | -0.02427 | -0.88194 | -0.2597 | -1.13323 | -0.36194 | -0.08249 | 0.42243 | -0.08555 | -0.07118 | 0.437285 | -0.55988 | -0.88428 | -0.28037 | -0.71762 | -0.27598 | -0.48536 | -0.70309 | -0.13673 | -0.1657 | -0.27908 | -0.59749 | -0.36104 | -0.39721 | [illegible] |
| 745 | -0.41246 | -0.64792 | 0.225555 | -0.77355 | -0.24884 | -0.8819 | -0.14927 | 0.050571 | -0.34283 | -1.13725 | -0.67004 | -1.27764 | -1.24782 | -0.22602 | -0.16351 | -0.1088 | -0.46727 | -0.3125 | -0.74232 | -0.54545 | -0.71313 | 0.008356 | -0.18403 | 0.336194 | 0.136548 | 0.059134 | 0.564623 | -0.69219 | -0.5398 | -0.29779 | -0.47127 | -0.55207 | -0.68696 | -0.84209 | -0.61312 | -0.2495 | -0.19012 | -0.27038 | -0.24677 | -0.51545 | [illegible] |
| 746 | -0.25786 | -0.4495 | -0.63148 | -0.78703 | -0.31725 | -0.69683 | 0.10141 | 0.067442 | -0.22521 | -1.32711 | -0.45368 | -1.29567 | -1.62114 | -0.3749 | -0.15769 | -0.36851 | -0.30616 | -0.12787 | -0.45983 | -0.43763 | -0.32208 | -0.1535 | -0.15685 | 0.050207 | -0.26914 | 0.059134 | 0.554591 | -0.88583 | -0.64769 | -0.40836 | -0.20401 | -0.89877 | -0.49739 | -0.6808 | -0.59324 | -0.22192 | -0.38901 | -0.42282 | -0.08384 | -0.37689 | [illegible] |
| 747 | -0.10574 | -0.28829 | -0.68505 | -0.73937 | -0.06451 | -0.5451 | -0.36377 | 0.148173 | -0.49704 | -0.89931 | -0.37788 | -1.16657 | -1.12486 | -0.57565 | -0.00281 | -0.35467 | -0.60198 | -0.01041 | -0.23441 | -0.61202 | -0.04177 | -0.30863 | 0.28931 | 0.034167 | -0.87946 | -0.43476 | 0.06059 | -0.81938 | -0.63837 | -0.53664 | -0.18485 | -1.41451 | -0.26488 | -0.50049 | -0.58949 | -0.35338 | 0.223855 | -0.39652 | -0.17585 | -0.29196 | [illegible] |
| 748 | -0.08302 | -0.54539 | -0.60463 | -0.81018 | -0.41005 | -0.29444 | -0.36377 | -0.2235 | -0.72138 | -0.77255 | 0.005189 | -0.78409 | -0.94314 | -0.56364 | -0.01504 | -0.18949 | -0.45666 | -0.49665 | 0.399154 | -0.59321 | -0.09578 | -0.11539 | -0.13129 | -0.06332 | -0.89138 | -0.16067 | 0.06059 | -0.83428 | -0.40663 | -0.31957 | 0.250897 | -1.28254 | -0.20315 | -0.02777 | -0.33388 | -0.71818 | 0.346786 | -0.39652 | 0.295286 | -0.14691 | [illegible] |
| 749 | 0.103393 | -0.60409 | -0.67867 | -0.50344 | -0.31132 | 0.351391 | -0.71476 | -0.15657 | -0.63528 | -0.64565 | 0.156004 | -0.60165 | -0.90929 | -0.34676 | 0.160759 | 0.453525 | 0.137824 | -0.48363 | 0.399325 | -0.42378 | 0.007155 | -0.23908 | 0.10946 | -0.24893 | -0.62937 | 0.521829 | -0.26544 | -0.91715 | -0.10693 | -0.2125 | 0.468998 | -0.93038 | -0.12432 | -0.08634 | -0.53749 | -0.36793 | 0.862242 | -0.29813 | 0.505075 | 0.001827 | [illegible] |
| 750 | -0.23204 | -0.70421 | -0.26115 | -0.47228 | -0.1847 | 1.136856 | -0.45915 | -0.00709 | -0.7626 | -0.48398 | -0.82001 | -0.48404 | -0.66327 | -0.47263 | -0.04614 | 0.093634 | 0.560601 | -0.28215 | 1.003531 | -0.72034 | -0.01442 | -0.3316 | 0.384148 | -0.83016 | -0.93593 | 0.377199 | -0.23881 | -0.42822 | -0.36507 | -0.15975 | 0.296234 | -0.56278 | -0.26126 | 0.169842 | -0.59248 | -0.37063 | 0.841228 | 0.15993 | 0.709851 | 0.083619 | [illegible] |
| 751 | -0.25735 | -0.90047 | -0.12343 | -0.53602 | -0.32523 | 1.331392 | -0.51808 | 0.07781 | -0.27127 | -0.27185 | -0.64123 | -0.2115 | -0.468 | -0.35431 | 0.199096 | 0.093634 | 0.721963 | -0.30034 | 0.876878 | -1.04114 | -0.27175 | -0.75792 | 0.307626 | -0.96663 | -0.85969 | 0.470618 | -0.17578 | -0.02929 | -0.20219 | 0.204048 | 0.324275 | 0.022935 | -0.27692 | -0.15416 | -0.54266 | -0.25704 | 0.715695 | -0.21109 | 0.671875 | -0.15419 | [illegible] |
| 752 | -0.03143 | -1.18424 | 0.001599 | -0.97324 | -0.81543 | 1.633302 | -0.48638 | -0.28026 | -0.30609 | -0.38036 | -0.58248 | -0.10691 | -1.15464 | -0.37423 | -0.28633 | 0.285236 | 0.971883 | -0.55149 | 0.36061 | -0.87422 | -0.06125 | -0.77308 | 0.405293 | -1.202 | -0.39556 | 1.089747 | 0.15851 | -0.20303 | -0.34288 | 0.273706 | 0.581623 | -0.1056 | -0.55743 | -0.14357 | -0.41951 | -0.33032 | 0.446869 | -0.07333 | 0.634581 | -0.39233 | [illegible] |
| 753 | 0.012269 | -1.16016 | 0.086283 | -1.08677 | -0.63022 | 1.320298 | -0.84471 | -0.32367 | -0.94645 | -0.18658 | -0.25065 | 0.132069 | -0.6733 | -0.38082 | -0.48843 | 0.207611 | 0.842932 | -0.61021 | -0.01513 | -1.3047 | 0.164759 | -0.87926 | 0.60681 | -1.03043 | -0.04341 | 0.808794 | 0.22317 | 0.243086 | -0.40886 | -0.15719 | 0.338579 | -0.0003 | -0.34172 | -0.59979 | -0.528 | -0.55669 | 0.508661 | -0.35088 | 0.231487 | -0.38301 | [illegible] |
| 754 | -0.42054 | -0.97712 | 0.01991 | -1.3413 | -0.70058 | 0.753096 | -0.66965 | -0.71978 | -0.78166 | 0.081331 | -0.16259 | -0.13152 | -0.68069 | -0.57962 | -0.81514 | -0.33128 | 0.081461 | -0.42941 | -0.01117 | -1.3797 | -0.09583 | -0.94721 | 0.118595 | -1.23908 | -0.36407 | 0.428599 | 0.765043 | 0.517526 | -0.13971 | -0.13479 | 0.080854 | 0.034507 | -0.37679 | -0.51086 | -0.52661 | -0.84471 | 0.029243 | -0.48654 | 0.504194 | -0.6895 | [illegible] |
| 755 | 0.074795 | -0.77583 | -0.40698 | -1.11825 | -0.90553 | 0.293161 | -0.46086 | -0.9821 | -0.92151 | 0.224101 | 0.506942 | -0.31894 | -0.70496 | -0.60426 | -0.47194 | -0.21744 | -0.25125 | -0.79078 | -0.42912 | -1.06941 | -0.07195 | -0.8385 | -0.15791 | -0.70476 | -0.16068 | 0.591679 | 1.059198 | -0.06738 | -0.35225 | -0.04678 | 0.174397 | -0.10569 | -0.13818 | -0.66471 | -0.18495 | -0.88284 | 0.317857 | -1.01474 | 0.337047 | -0.73035 | [illegible] |
| 756 | -0.10135 | -0.69823 | -0.32634 | -0.96795 | -0.62401 | -0.08541 | -0.49813 | -1.17918 | -1.35679 | 0.329277 | 0.256176 | -0.64716 | -0.84317 | -0.44881 | -0.64839 | -0.46573 | -0.56904 | -0.75625 | -0.39917 | -1.06299 | -0.34675 | -0.2958 | -0.17966 | -0.79237 | -0.24566 | 0.64147 | 1.348501 | -0.25707 | -0.55978 | -0.09685 | 0.040787 | -0.58809 | 0.054496 | -0.38118 | -0.5967 | -1.21459 | 0.36267 | -0.56642 | 0.155732 | -0.72996 | [illegible] |
| 757 | -0.4433 | -0.61344 | -0.27619 | -0.87549 | -0.09698 | -0.529 | -0.36772 | -0.93083 | -1.08957 | 0.364169 | -0.01392 | -0.66922 | -0.29394 | -0.33184 | -0.61525 | -0.64059 | -0.67813 | -0.73578 | 0.286092 | -0.78257 | -0.55917 | -0.12551 | -0.57406 | -0.55701 | -0.52243 | -0.03043 | 1.000984 | -0.16775 | -0.49884 | 0.031682 | -0.03069 | -0.38873 | -0.11376 | -0.4232 | -0.94429 | -1.12703 | 0.618842 | -0.7358 | 0.106443 | -0.38768 | [illegible] |
| 758 | -0.56393 | -0.52226 | -0.19855 | -0.49537 | -0.34423 | -0.44143 | -0.29998 | -0.58899 | -0.614 | 0.311022 | -0.22271 | -1.03053 | -0.77843 | -0.65171 | -0.76628 | -0.24436 | -0.30135 | -0.13597 | 0.114003 | -0.69903 | -0.54286 | 0.196109 | -0.48269 | -0.5072 | -0.65467 | -0.02357 | 1.723866 | -0.33941 | -0.47338 | -0.02191 | -0.29428 | -0.25089 | -0.47016 | -0.5088 | -0.77377 | -0.17205 | 0.186271 | -0.52497 | 0.038398 | -0.39983 | [illegible] |
| 759 | -0.17782 | -0.4083 | -0.03285 | -0.28749 | -0.35584 | -0.48999 | 0.05019 | -0.07887 | -0.88031 | -0.03961 | -0.38161 | -1.18246 | -0.42006 | -0.50791 | -0.88338 | -0.09154 | -0.13436 | -0.24481 | -0.15716 | -0.30416 | -0.33733 | 0.296064 | -0.22208 | -0.2955 | -0.1854 | -0.29623 | 1.783514 | -0.53132 | -1.09613 | -0.11219 | -0.52805 | -0.34172 | -0.4957 | -0.46268 | -0.73052 | 0.077865 | 0.200538 | -0.37613 | -0.40498 | -0.29265 | [illegible] |
| 760 | -0.32601 | -0.48872 | -0.0015 | -0.60413 | -0.51406 | -0.948 | -0.44774 | 0.01203 | -0.50352 | -0.13344 | -0.84209 | -0.79591 | -0.27378 | -0.14685 | -0.96585 | 0.194602 | 0.027375 | 0.131076 | -0.5218 | -0.22111 | -0.20876 | 0.061478 | -0.22026 | -0.24464 | -0.15855 | -0.4216 | 1.31096 | -0.45122 | -0.93483 | -0.22151 | -0.73884 | -0.52352 | -0.63049 | -0.57062 | -0.55586 | 0.107566 | -0.00303 | -0.36202 | -0.72663 | -0.51091 | [illegible] |
| 761 | -0.12201 | -0.38144 | -0.37071 | 0.315591 | -0.68931 | -1.07754 | -0.07211 | 0.137024 | -0.47617 | -0.0712 | -0.86288 | -0.75158 | -0.0187 | 0.080249 | -0.80743 | 0.385009 | 0.117035 | 0.15458 | -0.4363 | 0.182522 | 0.159759 | -0.01682 | -0.12199 | 0.593895 | 0.066222 | -0.76772 | 1.219624 | -0.39303 | -0.79452 | -0.08481 | -0.56733 | -0.23884 | -0.71167 | -0.64633 | -0.6029 | 0.349745 | -0.11903 | -0.27516 | -0.77156 | -0.02341 | [illegible] |
| 762 | -0.08151 | -0.39438 | -0.57793 | 0.695532 | -1.22528 | -1.28401 | -0.19525 | 0.418704 | -0.64963 | -0.1433 | -0.76552 | -0.61795 | -0.32302 | -0.53847 | -0.47942 | 0.263425 | 0.030974 | 0.580446 | -1.03999 | -0.07312 | 0.086034 | -0.01682 | -0.16771 | 0.727818 | 0.138758 | -0.56356 | 1.447172 | -0.45976 | -0.79872 | -0.2702 | -0.70528 | 0.03635 | -0.55128 | -0.90812 | -0.2055 | 0.497355 | -0.68634 | -0.12095 | -0.68828 | 0.111761 | [illegible] |
| 763 | -0.30751 | -0.31775 | -0.60336 | 0.793982 | -1.16783 | -1.50611 | -0.60865 | 0.628469 | -0.64456 | -0.23402 | -0.71509 | -0.31825 | -0.55307 | -0.24756 | -0.16643 | -0.12263 | -0.51368 | 0.045529 | -0.64158 | 0.319175 | 0.228061 | -0.48742 | -0.31444 | 0.542888 | -0.06932 | -0.77385 | 0.65963 | -0.06277 | -0.68455 | -0.01132 | -0.19576 | -0.10899 | -0.67613 | -0.58378 | -0.02591 | -0.21071 | -0.62901 | -0.29163 | -0.80864 | 0.14819 | [illegible] |
| 764 | -0.34044 | -0.31775 | -0.66617 | 0.437574 | -1.12568 | -1.58987 | -0.78066 | 0.901939 | -0.92768 | -0.02041 | -1.09737 | 0.220132 | -1.0521 | -0.40633 | -0.14632 | -0.12263 | -0.51368 | 0.238861 | -0.68441 | 0.522208 | 0.180188 | -0.39574 | -0.39815 | 0.633005 | -0.23747 | -0.46112 | 0.470153 | 0.564334 | -0.55579 | -0.01631 | -0.09346 | 0.134228 | -0.69479 | -0.58378 | 0.511073 | -0.30996 | -0.67932 | -0.31624 | -0.57096 | 0.232812 | [illegible] |
| 765 | -0.58493 | -0.31775 | -0.56882 | 0.732655 | -0.5949 | -1.41602 | -0.8371 | 0.923871 | -1.06928 | 0.010757 | -0.84367 | -0.42317 | -1.07672 | -0.61688 | -0.14632 | -0.16272 | -0.70302 | 0.068672 | -0.42677 | 0.187991 | 0.027738 | -0.29816 | -0.39815 | 0.633005 | -0.34619 | -0.38548 | 0.109856 | 0.802072 | -0.46962 | 0.151949 | -0.06022 | 0.364714 | -0.53047 | -0.50984 | 0.123305 | -0.08762 | -0.67932 | -0.18769 | -0.60026 | 0.256375 | [illegible] |
| 766 | -0.86559 | -0.48101 | 0.094688 | -0.02698 | -0.56909 | -1.04006 | -1.07132 | 0.621455 | -0.57573 | -0.05063 | -0.92238 | -0.05167 | -0.95521 | -1.09216 | -0.14509 | -0.10484 | -0.86762 | 0.064743 | -0.31115 | 0.104571 | 0.029964 | -0.38457 | -0.39815 | -0.1792 | -0.35719 | -0.43818 | -0.13675 | 0.556677 | -0.27165 | -0.2354 | 0.019177 | 0.204947 | -0.44315 | -0.66522 | 0.445017 | -0.28666 | -0.57244 | -0.59199 | -0.41061 | 0.291306 | [illegible] |
| 767 | -0.77364 | -0.41856 | 0.22613 | -0.33231 | -0.35879 | -1.02179 | -1.17865 | 0.086618 | -0.40227 | -0.16479 | -0.99368 | -0.1853 | -0.75742 | -0.45547 | -0.20866 | 0.016785 | -1.1117 | -0.36589 | 0.044905 | 0.12439 | 0.207485 | -0.38457 | -0.05833 | -0.31312 | -0.42972 | -0.63622 | -0.40344 | 0.734211 | 0.012148 | -0.19319 | 0.157128 | -0.05303 | -0.43528 | -0.22268 | 0.321556 | -0.32487 | -0.24819 | -0.72396 | -0.66276 | 0.13312 | [illegible] |
| 768 | -0.41097 | -0.52069 | 0.049317 | -0.91127 | -0.11345 | -0.80848 | -0.88711 | -0.11523 | -0.35185 | -0.3112 | -0.88653 | 0.223239 | -0.40625 | -0.78 | -0.20866 | 0.016785 | -0.90233 | 0.268184 | -0.15523 | -0.22664 | -0.02013 | -0.12942 | -0.08927 | -0.40913 | -0.41876 | -0.69504 | -0.40344 | 0.332497 | 0.255449 | -0.19693 | -0.21645 | -0.25458 | -0.23148 | -0.22268 | -0.12667 | -0.25869 | 0 | -0.22016 | -0.79882 | 0.26818 | [illegible] |
| 769 | -0.41097 | -0.54235 | 0.330781 | -0.62458 | 0.148369 | -0.37343 | -0.9774 | -0.56143 | 0.026225 | -0.54953 | -0.42711 | 0.077843 | -0.09233 | -0.42113 | -0.05067 | -0.05049 | -0.96049 | 0.103979 | -0.07947 | -0.5701 | -0.2739 | -0.33607 | -0.08059 | -0.40913 | -0.45456 | -0.69315 | -0.73601 | -0.26183 | 0.220312 | -0.19693 | 0.054296 | -0.47647 | -0.04272 | -0.43797 | -0.4713 | -0.25869 | -0.04421 | -0.199 | -0.78289 | 0.409982 | [illegible] |
| 770 | -0.16909 | -0.6388 | -0.09361 | -0.37435 | -0.097 | -0.65551 | -0.86807 | -0.53797 | 0.407594 | -0.73903 | -0.39602 | 0.643852 | -0.24672 | -0.05433 | -0.05067 | -0.01814 | -0.66677 | 0.255832 | -0.27375 | -0.40312 | 0.068157 | -0.19876 | -0.42159 | 0.027929 | -0.36443 | -0.79948 | -0.66755 | -0.14112 | 0.010934 | -0.20987 | 0.175609 | -0.48591 | 0 | -0.34054 | -0.62971 | -0.25869 | -0.12807 | -0.17895 | -0.48084 | 0.863027 | [illegible] |
| 771 | -0.22436 | -0.60258 | -0.51253 | -0.44202 | 0.239452 | -1.26058 | -0.77535 | -0.56943 | 0.1369 | -1.19155 | -0.26151 | 0.799674 | -0.14169 | 0.216247 | -0.24509 | 0.988931 | -0.61766 | -0.07121 | -0.42837 | -0.12761 | -0.21597 | 0.089366 | -0.42159 | 0.103925 | -0.29453 | -0.54387 | -0.56739 | -0.25833 | 0.072246 | -0.62702 | -0.01538 | -0.22658 | -0.13694 | -0.37023 | -0.88637 | 0.424625 | -0.31577 | -0.12228 | -0.69356 | 1.394444 | [illegible] |
| 772 | -0.35119 | -0.5233 | -0.83697 | -0.24328 | 0.275466 | -0.97259 | -0.65569 | -0.44737 | -0.00693 | -1.16353 | -0.00808 | 1.054199 | -0.34431 | 0.04075 | -0.43972 | 1.464398 | -0.41297 | 0.066158 | -0.54842 | -0.2046 | -0.37702 | 0.124133 | -0.75719 | 0.020269 | -0.44641 | -0.11536 | -0.40524 | -0.51636 | -0.15962 | -0.63937 | -0.19683 | 0.207748 | -0.34302 | -0.70775 | -0.94855 | 1.028769 | -0.28214 | 0.036166 | -0.49786 | 1.306217 | [illegible] |
| 773 | -0.60198 | -0.65675 | -0.97735 | -0.01897 | 0.26466 | -1.034 | -0.31071 | -0.36304 | -0.04006 | -1.44454 | -0.17261 | 0.57337 | -0.45007 | 0.114352 | -0.37067 | 1.544187 | -0.17187 | -0.34908 | -0.52203 | 0.125656 | -0.43157 | 0.046823 | -0.74004 | 0.130481 | -0.36534 | 0.381816 | -0.5543 | -0.37027 | -0.5955 | -0.68054 | -0.2091 | 0.500892 | -0.42457 | -0.91436 | -0.56273 | 1.220535 | -0.22956 | -0.36181 | -0.43797 | 1.151596 | [illegible] |
| 774 | -0.60998 | -1.21005 | -1.34313 | -0.01157 | -0.5095 | -1.15534 | 0.003759 | -0.33889 | -0.22878 | -1.10943 | -0.44573 | 0.815501 | -0.32425 | 0.062872 | -0.40809 | 1.789703 | 0.134567 | -0.29811 | -0.39455 | -0.06499 | -0.36385 | 0.102314 | -0.70912 | 0.037966 | -0.74083 | 0.338462 | -0.12124 | -0.15087 | -0.58942 | -1.03398 | -0.55806 | 0.613953 | -0.26011 | -0.66197 | -0.66083 | 1.205479 | 0.523456 | -0.48726 | -0.60798 | 0.865086 | [illegible] |
| 775 | -0.65407 | -1.54259 | -0.80287 | -0.25437 | -0.53725 | -1.64143 | 0.188927 | -0.36235 | -0.39665 | -0.79865 | -0.64982 | 0.923145 | -0.40824 | -0.76438 | -0.43788 | 1.268469 | -0.43331 | -0.64053 | -0.24948 | 0.122543 | -0.40245 | -0.08238 | -0.69683 | -0.52669 | -0.56216 | -0.18102 | -0.30494 | -0.50574 | -0.25047 | -1.06862 | -0.82688 | 0.6403 | -0.26011 | -0.59062 | -0.49769 | 0.855453 | 0.656114 | -0.73342 | -0.64241 | 0.678296 | [illegible] |
| 776 | -0.54512 | -1.4154 | -0.69773 | -0.31552 | -0.88789 | -1.50517 | -0.12587 | -0.04128 | -0.43307 | -1.04758 | -0.66283 | 0.538302 | -0.51005 | -0.82948 | -0.53846 | 0.352475 | -0.15191 | -0.20217 | -0.84098 | -0.14092 | -0.09936 | -0.24997 | -0.53046 | -0.28261 | -0.60062 | -0.49224 | -0.61709 | -0.48259 | -0.68029 | -0.43836 | -0.86031 | 0.635697 | -0.38785 | -0.38505 | -0.54844 | 0.383229 | 0.980587 | -0.66663 | -0.42969 | -0.23338 | [illegible] |
| 777 | -0.85452 | -1.45135 | -0.65401 | -0.61345 | -0.66445 | -1.95568 | -0.15941 | 0.126891 | -0.31562 | -0.91115 | -0.7657 | 0.08279 | -0.83533 | -0.96157 | -0.27703 | -0.42184 | -0.13279 | -0.19008 | -1.06142 | -0.03129 | -0.03921 | -0.38784 | -0.26203 | -0.5503 | -0.57104 | -0.95875 | -0.45239 | -0.02226 | -0.55804 | -0.12149 | -0.8427 | -0.37794 | -0.36689 | -0.18441 | -0.57606 | -0.16287 | 0.833181 | -0.70362 | -0.52832 | -0.35615 | [illegible] |
| 778 | -0.82149 | -1.07686 | -0.45208 | -0.70491 | -0.38341 | -1.98316 | -0.0249 | -0.19622 | -0.18313 | -0.53602 | -0.39955 | -0.07705 | -0.55122 | -0.77716 | -0.34269 | -0.55922 | -0.78174 | 0.131527 | -1.29294 | -0.13546 | -0.255 | -0.48092 | 0.155473 | -0.50032 | -0.28334 | -1.26593 | -0.33361 | -0.17889 | -0.35595 | -0.02246 | -0.83136 | -0.50924 | -0.26527 | 0.129839 | -1.07011 | -0.21109 | 0.74146 | -0.60046 | -0.53724 | 0.121191 | [illegible] |
| 779 | -0.91632 | -0.73602 | -0.16696 | -1.07612 | -0.11715 | -2.08826 | -0.16889 | -0.10847 | -0.45363 | -0.79507 | -0.29301 | -0.33671 | -0.72861 | -0.8688 | -0.61427 | -0.81493 | -0.86693 | -0.02779 | -1.33954 | -0.11493 | 0.020686 | -0.4298 | -0.10158 | -0.57293 | -0.25082 | -1.3261 | -0.22008 | -0.17396 | -0.6979 | 0.791816 | -0.93318 | -0.65149 | -0.74971 | 0.263291 | -1.26779 | -0.15056 | -0.27643 | -0.40216 | -0.61815 | 0.120125 | [illegible] |
| 780 | -1.02745 | -0.08773 | -0.33398 | -1.14149 | 0.445617 | -1.52601 | -0.38974 | -0.48516 | -0.56502 | -0.66573 | 0.106245 | -0.24567 | -0.76655 | -0.79373 | -0.55041 | -0.19188 | -0.63029 | -0.06454 | -1.64899 | -0.54393 | -0.10333 | -0.78091 | 0.223235 | -0.57685 | 0.093631 | -0.94689 | 0.351485 | -0.08792 | -0.91034 | 1.280109 | -0.96875 | -0.75393 | -0.91498 | -0.11555 | -0.92078 | 0.404887 | -0.33936 | -0.53751 | -0.74289 | -0.28611 | [illegible] |
| 781 | -1.05735 | 0.055621 | -0.52769 | -1.09846 | 0.70201 | -1.25975 | -0.29843 | -0.57391 | -0.73446 | 0.093604 | 0.17759 | -0.49437 | -0.85526 | -0.89739 | -0.21775 | -0.50826 | -1.06206 | -0.26143 | -1.29051 | -0.47772 | -0.08929 | -0.71241 | -0.0667 | -0.69917 | -0.097 | -0.67287 | 0.593586 | -0.12116 | -0.76165 | 1.201948 | -0.97561 | -0.98131 | -0.70988 | -0.33573 | -0.54566 | 0.627438 | -0.51495 | -0.49854 | -0.87795 | -0.06189 | [illegible] |
| 782 | -0.9644 | -0.11468 | -0.18526 | -1.05426 | 0.045604 | -1.46346 | -0.36827 | -0.59231 | -0.75033 | -0.06645 | 0.174131 | -0.27461 | -0.32092 | -0.76614 | -0.32095 | -0.1814 | -1.04419 | 0.033268 | -1.38158 | -0.75383 | -0.02762 | -0.31237 | -0.39556 | -0.67518 | -0.22846 | -0.39505 | 0.412754 | -0.72572 | -0.97003 | 0.946194 | -0.86657 | -0.13461 | -0.62828 | -0.63725 | -0.52571 | 0.558976 | -0.76655 | -0.3045 | -1.30686 | 0.067983 | [illegible] |
| 783 | -0.82392 | -0.14689 | -0.09052 | -0.87306 | -0.28898 | -1.84062 | -0.39166 | -0.6395 | -1.02584 | 0.010785 | -0.23571 | 0.099165 | -0.79781 | -0.96814 | -0.4381 | -0.04213 | -0.64652 | -0.31835 | -1.35312 | -0.73687 | 0.426146 | -0.10926 | -1.06449 | -0.92947 | -0.45851 | -0.99706 | 0.534577 | -1.02944 | -0.74603 | 1.120627 | -0.56697 | -0.14192 | -0.71884 | -0.86372 | -0.34322 | 0.858632 | -0.59802 | -0.11935 | -1.60899 | -0.38067 | [illegible] |
| 784 | -0.52784 | -0.16437 | 0.156477 | -0.53123 | -0.64908 | -2.11054 | -0.8614 | -0.81144 | -0.73001 | 0.404208 | -0.08232 | 1.143283 | -0.48924 | -1.19221 | -0.30594 | -0.10284 | -0.65648 | -0.3039 | -1.42385 | -0.49166 | 0.329607 | 0.102426 | -0.92722 | -0.90469 | -0.14296 | -0.95571 | 0.221251 | -1.31238 | -0.12532 | 0.967172 | -0.2368 | -0.04294 | -0.10928 | -0.73423 | -0.22695 | 0.867329 | -0.2099 | -0.51846 | -1.16156 | -0.12561 | [illegible] |
| 785 | -0.57216 | -0.64843 | 0.190013 | -0.41825 | -1.51569 | -1.80126 | -0.63232 | 0.022783 | -0.32444 | 0.254447 | -0.43315 | 1.331935 | -0.41641 | -0.82252 | -0.62456 | -0.3063 | -0.75554 | 0.076284 | -0.90151 | -0.08684 | -0.39069 | 0.572002 | -0.98006 | -0.80929 | -0.85053 | -0.74222 | -0.03118 | -0.60428 | 0.256085 | 0.018051 | -0.07924 | 0.050378 | 2.850653 | -0.53105 | -0.17909 | 0.270868 | -0.1113 | -0.19744 | -1.2164 | 0.000614 | [illegible] |
| 786 | -0.37257 | -0.5365 | 0.355426 | -0.57414 | -1.36521 | -1.26533 | -0.68072 | 0.034765 | -0.10101 | 0.211398 | -0.50456 | 1.504777 | -0.1933 | -0.7583 | -0.72006 | -0.20697 | -0.20276 | 0.260148 | -0.67135 | -0.18301 | -0.59732 | 0.495033 | -0.86471 | -0.31674 | -0.5204 | -1.1674 | 0.019518 | -0.05274 | 0.297902 | 0.049307 | 0.035979 | 0.333794 | 5.30746 | -0.40221 | -0.18409 | -0.60663 | -0.36123 | -0.15826 | -1.05652 | -0.19218 | [illegible] |
| 787 | -0.16303 | -0.22506 | 0.253188 | -0.06057 | -0.46795 | -0.6153 | -0.28959 | -0.36232 | -0.12068 | 0.386124 | -0.3694 | 1.152427 | -0.28502 | -0.99592 | -0.65326 | -0.56364 | -0.01685 | -0.2334 | 0.37287 | 0.287316 | -0.55808 | 0.0702 | -0.79915 | 0.315185 | -0.50305 | -0.99044 | -0.25942 | -0.15735 | 0.553974 | 0.095616 | -0.23568 | -0.19528 | 5.455096 | -0.08195 | -0.03633 | -0.55971 | -0.17127 | -0.64144 | 0.126077 | -0.05941 | [illegible] |
| 788 | -0.45061 | -0.23569 | 0.025668 | 0.08464 | -0.51392 | -0.48883 | -0.6624 | -0.20478 | 0.065946 | 0.571751 | -0.08881 | 1.018563 | 0.143157 | -1.11981 | -0.3872 | -0.66825 | -0.48416 | -0.25199 | 1.132869 | 0.334414 | -0.70329 | -0.0494 | -0.74454 | 0.889556 | -0.88568 | -0.33354 | -0.87797 | 0.13128 | 0.325473 | 0.060949 | -0.44923 | 0.125152 | 5.722286 | -0.05128 | 0.147327 | -0.93241 | -0.12274 | -0.68268 | 1.391229 | 0.022784 | [illegible] |
| 789 | -1.00912 | 0.045104 | -0.19932 | 0.09587 | -0.24608 | -0.20744 | -0.08976 | -0.06176 | 0.466801 | 0.333535 | -0.46268 | -0.10992 | -0.57073 | -1.07104 | -0.35313 | -0.96819 | -0.92876 | -0.30109 | 1.278342 | -0.07897 | -0.65176 | -0.17894 | -0.73299 | 1.030526 | -0.95415 | -0.38746 | -0.63311 | 0.089123 | 0.351129 | -0.1112 | -0.55633 | -0.0655 | 5.216821 | -0.13524 | -0.03813 | -1.08725 | -0.37929 | -0.69408 | 1.069941 | -0.29397 | [illegible] |
| 790 | -1.0175 | 0.313633 | -0.43314 | -0.36889 | -0.38069 | -0.29467 | -0.0069 | -0.79059 | 0.427691 | 0.358295 | -0.82748 | -0.55933 | -0.29565 | -0.99868 | -0.09452 | -1.15372 | -0.49523 | -0.23676 | 0.802397 | -0.44183 | -0.20575 | -0.01521 | -0.766 | 0.888991 | -0.50588 | -0.44472 | -0.5336 | -0.67617 | 0.415442 | 0.441585 | -0.53181 | 0.000283 | 2.600974 | -0.2154 | -0.17505 | -0.90918 | -0.38981 | -0.78284 | 1.028613 | -0.10349 | [illegible] |
| 791 | -1.31828 | -0.13665 | -0.19834 | 0.065238 | -0.5778 | -0.76235 | -0.15326 | -0.75634 | 0.136739 | 0.22861 | -0.53336 | -0.32506 | -0.60217 | -1.23973 | -0.2069 | -0.85484 | -0.69201 | -0.47151 | 0.701212 | -0.61848 | -0.0929 | -0.21135 | -0.8328 | 0.690451 | -0.38664 | -0.27917 | -0.71476 | -1.02802 | 0.326622 | 0.101662 | -0.6993 | -0.38056 | 0.221041 | 0.146429 | -0.4016 | -0.19027 | -0.28074 | -0.74849 | 0.65661 | -0.22295 | [illegible] |
| 792 | -1.08432 | -0.49601 | -0.36721 | 0.011892 | -0.94305 | -0.55103 | -0.42166 | -0.3482 | 0.279687 | 0.033277 | -0.38966 | -0.17226 | -0.87109 | -0.41247 | -0.4608 | -1.02846 | -0.42888 | -0.1018 | -0.04252 | -0.75271 | -0.04508 | -0.1873 | -0.5165 | 0.268333 | -0.18532 | -0.16318 | -0.64303 | -0.84795 | 0.064069 | -0.07272 | -0.49174 | -0.28692 | 0.469569 | 0.132972 | -0.71458 | -0.46038 | 0.066606 | -0.85698 | -0.42782 | -0.48923 | [illegible] |
| 793 | -0.73901 | -0.33111 | -0.29703 | -0.24963 | -0.85843 | -0.01067 | 0.022601 | 0.030281 | 0.51764 | -0.0442 | -0.69993 | -0.05178 | -0.84401 | 0.063596 | -0.81225 | -1.10483 | 0.054221 | -0.18616 | -0.93834 | -1.0082 | -0.03917 | -0.04085 | -0.27478 | -0.1351 | 0.152006 | 3.89E-05 | -0.18251 | -0.72631 | 0.054006 | -0.42208 | -0.88968 | -0.83722 | 0.457463 | 0.248262 | -1.34461 | -0.21644 | 0.027957 | -1.38016 | -1.11026 | -0.62072 | [illegible] |
| 794 | -0.50434 | -0.4219 | -0.45269 | -0.08684 | -0.93357 | 0.009758 | -0.43182 | -0.02721 | 1.242814 | 0.06442 | -0.42726 | 0.075279 | -0.27093 | 0.257983 | -0.62154 | -0.74241 | 0.183183 | -0.12495 | -0.879 | -0.80263 | -0.15061 | -0.23481 | -0.26437 | -0.35465 | 0.579284 | -0.0668 | -0.54448 | -0.66567 | 0.488152 | -0.60476 | -0.87526 | -0.75236 | 0.64172 | 0.00419 | -0.98802 | -0.28842 | -0.01414 | -1.18056 | -1.43358 | -0.4473 | [illegible] |
| 795 | 0.017467 | -0.5596 | -0.58091 | 0.271642 | -0.53058 | -0.22246 | 0.013154 | 0.244086 | 0.709262 | -0.19323 | 0.0621 | 0.290732 | -0.32485 | -0.05454 | -0.58251 | -0.32523 | 0.19009 | -0.44866 | -0.57072 | -0.34461 | -0.17479 | -0.46599 | -0.48241 | -0.51123 | 0.412629 | 0.46605 | -0.25189 | -0.63466 | 0.125302 | -0.82309 | -0.81312 | -0.6247 | 0.579221 | 0.318656 | -1.15258 | -0.14167 | -0.1963 | -1.27771 | -1.11114 | -0.74717 | [illegible] |
| 796 | 0.007204 | -0.57385 | -0.73728 | -0.07815 | -0.69612 | -0.20327 | -0.24352 | 0.912129 | 1.158792 | -0.37278 | -0.5968 | 1.251767 | -0.13821 | 0.465805 | -0.66212 | -0.49812 | 0.116374 | -0.13316 | -0.56253 | -0.26117 | -0.58359 | -0.19206 | -0.63483 | -0.74201 | -0.0869 | 1.266934 | -0.25977 | -0.97117 | 0.252126 | -0.85714 | -0.98665 | 0.075033 | 0.312417 | 0.07082 | -1.18051 | -0.30525 | 0.239597 | -1.11502 | -0.59948 | -0.49941 | [illegible] |
| 797 | -0.03489 | -0.19322 | -0.38051 | -0.12331 | -0.94736 | -0.52996 | 0.239377 | 1.216569 | 1.513096 | -0.1551 | -1.37226 | 1.394469 | -0.15989 | 0.167761 | -0.37339 | -0.21566 | 0.078986 | -0.10991 | -0.80635 | -0.51218 | -0.53124 | -0.14848 | -0.68212 | -0.50304 | -0.25151 | 1.072862 | 0.35142 | -0.84987 | 0.920557 | -0.85445 | -1.31196 | 0.484191 | -0.29047 | -0.00925 | -1.18649 | -0.46929 | 0.233733 | -0.9643 | -0.41911 | -0.36591 | [illegible] |
| 798 | -0.3349 | -0.24093 | -0.14605 | 0.74894 | -1.22941 | -0.96027 | 0.288424 | 0.369017 | 1.048117 | -0.0734 | -1.19917 | 0.986641 | -0.42247 | -0.50228 | -0.19038 | -0.02667 | 0.123741 | 0.128222 | -0.65457 | -0.60188 | -0.60799 | -0.15935 | -0.75571 | 0.206815 | -0.42322 | 0.668113 | 0.530164 | -0.97342 | 0.903648 | -0.40165 | -0.80892 | 0.832224 | -0.41299 | -0.14457 | -0.62132 | -0.88075 | 0.150962 | -0.50313 | -0.71877 | -0.56479 | [illegible] |
| 799 | -0.05508 | -0.11309 | -0.31397 | 1.410231 | -0.77927 | -0.98995 | 0.419622 | 0.410633 | -0.19027 | -0.11242 | -1.47181 | 1.215605 | -0.6141 | -0.68527 | -0.17448 | -0.00161 | 0.272647 | 0.044989 | -0.87913 | -0.90244 | -0.46597 | -0.10761 | -0.63922 | 1.30854 | -0.71381 | 0.609451 | 0.562469 | -0.69593 | 0.121647 | -0.16336 | -0.70474 | 0.742382 | -0.42486 | -0.21033 | -0.57625 | -0.74109 | 0.267107 | -0.66525 | -0.44956 | -0.82216 | [illegible] |
| 800 | -0.60248 | -0.15606 | 0.178842 | 2.210193 | -0.53835 | -0.87919 | 0.119851 | 0.378107 | -0.08299 | -0.25577 | -1.69628 | 0.889876 | -0.90321 | -0.63455 | -0.54121 | -0.19656 | 0.059348 | 0.347784 | -0.64465 | -1.06829 | -0.5385 | -0.03329 | -0.20477 | 2.617163 | -0.36153 | 0.082961 | 0.40007 | -0.77048 | 0.394317 | -0.08448 | -0.18516 | 0.310154 | -0.68646 | -0.33191 | -0.5562 | -0.46062 | 0.381648 | -0.44237 | -0.4567 | -0.72206 | [illegible] |
| 801 | -0.47755 | -0.26468 | 0.125167 | 2.180203 | -0.49639 | -0.83174 | 0.392055 | -0.29681 | -0.29747 | -0.29605 | -1.72587 | -0.4055 | -1.21454 | -1.30734 | -0.44615 | -0.34513 | 0.011885 | 0.170797 | -0.6745 | -1.00995 | -0.32582 | -0.42662 | 0.044 | 1.986566 | -0.38935 | -0.22454 | 0.464946 | -0.25157 | 0.345278 | -0.07587 | 0.090141 | -0.23801 | -0.59587 | -0.69064 | -0.46551 | -0.44638 | -0.28255 | -0.50599 | -0.33601 | -0.68574 | [illegible] |
| 802 | -0.68598 | -0.44091 | -0.03051 | 1.779756 | -0.13825 | -0.8943 | -0.08462 | -0.28596 | -0.4663 | -0.73484 | -1.27179 | -0.80507 | -0.7735 | -1.69813 | -0.31635 | -0.08154 | -0.30423 | -0.02634 | -0.07906 | -1.2425 | -0.37474 | -0.55501 | -0.12803 | 1.72964 | -0.18426 | -0.49699 | -0.06795 | -0.02637 | -0.06619 | -0.07535 | 0.38926 | -0.77986 | -0.49751 | -0.75831 | -0.30924 | -0.08544 | -0.55778 | -0.34975 | 0.232799 | -0.68564 | [illegible] |
| 803 | -0.55291 | -0.42472 | 0.26455 | 0.525481 | 0.09624 | -0.81609 | -0.43505 | 0.577403 | -0.05129 | -0.22223 | -1.42103 | -1.05619 | -1.04612 | -1.5307 | -0.67579 | -0.29553 | -0.41286 | -0.00115 | -0.20434 | -1.09531 | -0.15221 | -0.95829 | 0.02562 | 1.671198 | 0.092102 | -0.41783 | -0.12774 | 0.523988 | -0.16333 | -0.44437 | -0.13642 | -1.0962 | -0.97706 | -1.1523 | -0.51534 | 0.483316 | -0.94153 | -0.59642 | 0.56551 | -0.30168 | [illegible] |
| 804 | -0.64902 | -0.95166 | 0.762412 | -0.5879 | 0.012845 | -0.89181 | -0.55275 | -0.10708 | 0.287216 | -0.14345 | -1.51743 | -1.79068 | -1.04176 | -1.61605 | -0.47793 | -0.33784 | -0.69945 | 0.299531 | -0.13118 | -0.87563 | -0.2811 | -1.00233 | 0.086695 | 1.063948 | -0.07148 | -0.71114 | 0.110762 | -0.05642 | -0.11367 | -0.70427 | -0.15262 | -1.01431 | -0.86547 | -1.45088 | -0.709 | 0.411226 | -1.31119 | -0.24722 | 0.653306 | -0.1835 | [illegible] |
| 805 | -0.66533 | -0.9294 | 0.448717 | -1.47082 | 0.817575 | -0.96428 | -1.23393 | -0.07303 | 0.104612 | 0.290847 | -1.27377 | -1.73277 | -0.89045 | -1.39719 | -0.20543 | -0.46652 | -0.76551 | -0.23901 | 0.149608 | -1.18054 | -0.2898 | -1.16278 | -0.20898 | -0.0092 | 0.607246 | -0.75274 | 0.037893 | 0.144338 | -0.5352 | -0.71412 | -0.72132 | -0.9373 | -0.80854 | -1.45247 | -0.59773 | 0.086042 | -1.60566 | -0.21251 | 0.705187 | -0.25559 | [illegible] |
| 806 | -0.65385 | -0.58652 | 0.392314 | -1.34804 | 1.158679 | -1.04556 | -0.97084 | -0.47159 | 0.081561 | 0.526286 | -0.54589 | -1.61118 | -0.77864 | -1.04067 | -0.20072 | -0.30265 | -0.73887 | -0.34425 | -0.08056 | -0.85541 | -0.12616 | -1.12817 | -0.47705 | 0.648345 | 1.165789 | -1.02911 | 0.134987 | -0.2818 | -0.36798 | -0.57918 | -0.59108 | -0.84572 | -0.80744 | -1.14569 | -0.53841 | 0.312952 | -1.33272 | -0.49074 | 0.129806 | -0.35805 | [illegible] |
| 807 | -0.94092 | -0.6211 | 0.441647 | -1.06274 | 2.304441 | -0.68047 | -1.02985 | -0.87867 | -0.36678 | 1.182891 | -0.49949 | -1.31699 | -1.10818 | -0.97977 | -0.32843 | -0.79303 | -0.86317 | -0.69109 | -0.73904 | -0.59591 | -0.22044 | -1.10855 | -0.3232 | 0.716442 | 2.281308 | -0.89561 | 0.203972 | -0.50083 | -0.34093 | -0.34529 | -0.76414 | -0.62605 | -0.73981 | -1.03014 | -0.67172 | 0.30787 | -1.01789 | -0.7573 | -0.24016 | -0.55457 | [illegible] |
| 808 | -0.60712 | -0.79397 | 0.084014 | -0.69783 | 2.35148 | -0.55513 | -0.95808 | -1.45925 | -0.60401 | 0.21631 | -0.39697 | -1.34942 | -0.77699 | -0.70156 | -0.24875 | -0.78116 | -0.84164 | -0.79526 | -0.62335 | -0.3727 | -0.82628 | -0.83948 | -0.36739 | 0.244573 | 2.760048 | -0.61962 | 0.114056 | -1.08191 | -0.31255 | -0.46447 | -0.54937 | -0.72503 | -0.44031 | -0.55898 | -0.41806 | -0.17858 | -0.78159 | -0.67127 | -0.48481 | -0.69274 | [illegible] |
| 809 | -0.34903 | 0.215659 | -0.06729 | -0.49773 | 2.428321 | -0.17513 | -0.84554 | -0.98264 | -1.21609 | -0.3879 | -0.04748 | -0.7696 | -1.06182 | -0.7872 | -0.7546 | -0.89419 | -0.82572 | -0.91151 | -0.57099 | -0.36389 | -0.66847 | -0.33273 | -0.56056 | -0.2622 | 3.053925 | -0.05883 | -0.18212 | -0.57878 | -0.46851 | -0.47431 | -0.65738 | -0.55197 | -0.65661 | -0.01539 | 0.013607 | -0.30519 | -0.21475 | -1.10551 | -0.92963 | -0.85647 | [illegible] |
| 810 | -0.01479 | 0.38275 | -0.03456 | -0.23266 | 1.293544 | -0.13111 | -0.50555 | -0.86761 | -0.76015 | -0.72606 | -0.14845 | -0.96023 | -1.01955 | -0.92078 | -0.66598 | -0.78169 | -0.8187 | -0.59824 | -1.19521 | 0.048705 | -1.09385 | 0.127368 | -0.15558 | -0.43733 | 1.615984 | -0.01421 | -0.56737 | -0.60387 | -0.11162 | -0.71831 | -0.37782 | -0.25916 | -1.00403 | -0.12902 | 0.016061 | -0.44859 | -0.15746 | -1.19948 | -1.07895 | -0.2321 | [illegible] |
| 811 | 0.065661 | 0.076937 | -0.12943 | -0.06298 | 0.881954 | -0.1312 | -0.80368 | -0.66802 | -1.07935 | -1.34249 | -0.19315 | -0.73404 | -1.19467 | -0.8047 | -0.55992 | -1.05753 | -1.36994 | -0.39413 | -1.0276 | -0.10354 | -0.95641 | 0.295308 | 0.091148 | -0.24672 | 1.172056 | 0.118729 | -0.55099 | -0.32899 | -0.49343 | -0.86096 | -0.58412 | -0.41392 | -1.11025 | -0.24786 | -0.13309 | -0.51497 | -0.23984 | -1.24545 | -0.82311 | -0.29877 | [illegible] |
| 812 | 0.361148 | 0.619327 | -0.17882 | -0.09878 | -0.36667 | -0.14112 | -1.21653 | -0.79404 | -0.89392 | -1.40832 | -0.06204 | -0.50204 | -0.99937 | -0.90355 | -0.63554 | -0.75857 | -0.95046 | -0.04049 | -0.47141 | -0.25345 | -1.08558 | 0.698084 | 0.386447 | -0.106 | 0.083076 | 0.058758 | -0.43136 | -0.26781 | -0.42616 | -0.62764 | 0.005917 | -0.19052 | -0.93562 | 0.020881 | 0.026585 | -0.56043 | -0.78176 | -0.98777 | -0.7752 | -0.31703 | [illegible] |
| 813 | 0.096945 | 1.592669 | 0.223817 | -0.21882 | -0.65885 | -0.00641 | -1.23852 | -0.91477 | -1.2225 | -1.2815 | -0.07561 | -0.07802 | -1.10776 | -1.50835 | -0.66487 | -0.84118 | -1.02749 | -0.15032 | -0.71156 | -0.49214 | -0.96008 | 1.600032 | 0.605424 | -0.34589 | -0.67667 | 0.098035 | -0.4011 | -0.20956 | -0.68413 | -0.44585 | -0.02193 | -0.09331 | -1.06999 | -0.0326 | -0.3196 | -0.38948 | -0.80918 | -0.63018 | -0.61589 | -0.52833 | [illegible] |
| 814 | -0.54198 | 1.517978 | 0.121177 | 0.205529 | -0.66813 | -0.48175 | -1.13554 | -0.76059 | -0.64563 | -1.12975 | -0.20452 | -0.00653 | -0.87665 | -1.14082 | -0.30045 | -0.80878 | -0.69382 | -0.16972 | -0.58541 | -0.68114 | -1.37692 | 2.052454 | 0.908715 | -0.1945 | -1.12824 | 0.02446 | -0.56671 | -0.17639 | -0.05714 | -0.6446 | 0.251608 | -0.41213 | -1.15139 | -0.18388 | -0.64562 | -0.1578 | -1.04142 | -0.43232 | -0.41498 | -0.4993 | [illegible] |
| 815 | -0.80576 | 2.084139 | 0.215912 | -0.18009 | -0.27732 | -0.51931 | -0.85598 | -1.01598 | -0.4591 | -0.76686 | -0.22033 | 0.053701 | -0.96472 | -1.16377 | -0.60371 | -0.80507 | -0.5246 | -0.07847 | -0.53799 | -0.64308 | -0.90273 | 2.463892 | 0.385899 | 0.025447 | -0.76652 | -0.22414 | -0.3252 | -0.19221 | -0.09789 | -0.34697 | 0.251023 | -0.51918 | -0.77328 | 0.135363 | -0.38159 | -0.09913 | -0.76898 | -0.28585 | -0.38613 | -0.70658 | [illegible] |
| 816 | -1.0438 | 2.361404 | 0.300198 | -0.17408 | -0.14857 | -0.33138 | -0.6661 | -0.71479 | -0.19735 | -0.39023 | -0.52846 | 0.100999 | -0.66655 | -1.1582 | -0.20691 | -0.48202 | 0.263964 | 0.012842 | -0.45399 | -0.77892 | -1.25619 | 2.4858 | 0.276653 | -0.28919 | -0.93942 | -0.39513 | -0.58836 | -0.07302 | -0.04269 | -0.1442 | 0.138781 | -0.13946 | -0.04326 | 0.135733 | 0.148846 | -0.39011 | -0.55101 | -0.33761 | -0.13197 | -0.67208 | [illegible] |
| 817 | -1.07225 | 2.037382 | 0.445012 | -0.10512 | -0.07361 | -0.36286 | -0.2144 | -0.71639 | -0.47191 | -0.71467 | -0.61666 | -0.07826 | -0.80868 | -0.74664 | -0.24996 | -0.28496 | -0.09636 | -0.33507 | -0.43955 | -0.71367 | -1.41826 | 2.318683 | -0.21456 | 0.369439 | -1.08145 | -0.1107 | -0.72063 | -0.28222 | 0.001044 | -0.47939 | -0.28779 | -0.36239 | 0.134964 | 0.267134 | 0.021302 | -0.35212 | -0.2553 | -0.48771 | 0.05947 | -0.66776 | [illegible] |
| 818 | -1.01428 | 0.890699 | -0.16648 | -0.05457 | 0.394574 | 1.420058 | -0.26377 | -0.15883 | -0.45353 | -0.63078 | -0.77149 | -0.09719 | -0.5762 | -0.09326 | 0.133209 | -0.16754 | -0.03335 | -0.21224 | -0.15631 | -0.90268 | -1.17266 | 1.241523 | -0.59544 | 0.704572 | -0.9735 | 1.521545 | -0.64641 | -0.32109 | 0.026808 | -0.80351 | -0.2628 | -0.28671 | 0.667738 | 0.395653 | 0.573016 | -0.34325 | -0.19632 | -0.95747 | -0.06591 | -0.51145 | [illegible] |
| 819 | -0.98902 | 0.064533 | -0.18144 | -0.41339 | 0.448278 | 2.68196 | -0.06642 | 0.012422 | -0.96901 | -0.38059 | -0.74644 | -0.38006 | -0.69244 | -0.38285 | -0.03916 | -0.2143 | -0.05523 | -0.13935 | -0.17379 | -0.73149 | -1.2011 | 0.46055 | -0.88044 | 0.782104 | -0.60533 | 2.119862 | -0.05439 | 0.144072 | -0.55647 | -0.63706 | -0.38422 | -0.1135 | 0.755645 | 0.090847 | 0.721218 | -0.08799 | -0.03515 | -0.64156 | 0.205177 | -0.42174 | [illegible] |
| 820 | -1.19025 | -0.85622 | -0.34145 | -0.31772 | -0.12813 | 2.93315 | -0.16879 | 0.344188 | -1.32537 | -0.74599 | -0.58897 | -0.52389 | -0.39159 | -0.13952 | 0.539587 | -0.37444 | 0.058262 | -0.02143 | -0.36607 | -1.05873 | -1.29787 | -0.36336 | -0.72425 | 0.847925 | -0.67485 | 2.439596 | -0.03706 | 0.53301 | -0.94328 | -0.84608 | 0.178953 | -0.3539 | 0.933485 | -0.21171 | 0.126516 | 0.630935 | -0.47122 | -0.48197 | 0.054363 | -0.26364 | [illegible] |
| 821 | -0.85078 | -1.07546 | -0.28935 | 0.789495 | 0.358691 | 2.816997 | -0.12417 | 0.167472 | -0.5523 | -0.28402 | 0.313912 | -1.03176 | -0.18557 | -0.56954 | -0.18098 | -0.84452 | -0.3076 | -0.00781 | -0.33717 | -1.39319 | -1.27971 | -0.55747 | -0.59543 | 1.188671 | -0.9083 | 2.789203 | 0.249865 | 0.298856 | -0.0029 | -0.48738 | 0.558764 | -0.70601 | 0.529315 | -0.33325 | -0.32472 | 1.625144 | -0.65841 | 0.006115 | -0.29211 | -0.54934 | [illegible] |
| 822 | -0.9064 | -1.13869 | -0.56299 | 1.234655 | 0.495773 | 2.41174 | -0.60452 | 0.399947 | -0.00793 | -0.05272 | 0.301462 | -1.02224 | -0.25061 | -0.47852 | -0.25828 | -0.97638 | -0.29335 | 0.579089 | -0.99539 | -1.52733 | -1.07379 | -0.787 | -0.58576 | 1.299099 | -1.07937 | 2.435122 | 0.535916 | 0.430695 | 0.476853 | -0.47088 | 0.986179 | -0.91426 | -0.10129 | -0.50266 | -0.22371 | 1.757481 | -1.34574 | 0.59581 | -0.43366 | -0.34666 | [illegible] |
| 823 | -1.077 | -0.98732 | -0.48384 | 2.476643 | 0.493062 | 0.503177 | -0.20877 | 0.1541 | 0.358764 | -0.23858 | 0.691192 | -1.08251 | -0.26505 | -0.46865 | -0.3859 | -0.77041 | -0.29745 | 0.516433 | -1.01362 | -1.221 | -1.12041 | -0.88897 | -0.28201 | 1.332973 | -0.75362 | 0.735908 | 0.386329 | 0.546656 | 1.10175 | -0.11747 | 1.23779 | -0.88755 | -0.38337 | -0.61023 | -0.25257 | 1.979671 | -1.2336 | 0.984287 | -0.5428 | -0.48675 | [illegible] |
| 824 | -0.94346 | -0.63047 | -0.16034 | 2.709251 | 0.506807 | -0.06947 | -0.39973 | 0.113248 | 0.964155 | 0.284898 | 0.756049 | -1.11017 | 0.110704 | 0.174494 | -0.28314 | -0.62996 | -0.66764 | 0.595814 | -1.06169 | -1.31601 | -0.74023 | -0.96804 | -0.54386 | 1.583848 | -0.52922 | 0.304847 | -0.03232 | 0.194935 | 1.100751 | 0.421831 | 1.512831 | -0.81103 | -0.27687 | 0.022589 | -0.48078 | 1.739162 | -1.3722 | 1.048076 | -0.51216 | -0.47149 | [illegible] |
| 825 | -0.86412 | -0.28832 | -0.18124 | 2.582938 | 0.5152 | -0.0419 | -0.56516 | -0.07969 | 1.066611 | 0.240488 | 0.802147 | -0.98593 | -0.32613 | 0.073974 | -0.5661 | -0.42477 | -1.05312 | 0.415627 | -1.12049 | -1.08532 | -0.54868 | -0.9333 | -0.5918 | 1.720692 | -0.41676 | 0.400527 | -0.00388 | -0.31336 | 1.341866 | 0.682142 | 0.662738 | -0.65737 | -0.53199 | 0.177192 | -0.1999 | 0.975408 | -0.97772 | 0.542505 | -0.67535 | -0.75709 | [illegible] |
| 826 | -0.87308 | -0.05431 | -0.15853 | 1.316553 | 0.108512 | 0.035339 | -0.5854 | -0.23191 | 0.131532 | 0.025868 | 0.105312 | -0.54868 | -0.58818 | 0.273673 | -0.40332 | -0.07894 | -0.85339 | 0.209782 | -1.31707 | -0.95548 | -0.25757 | -0.76018 | -0.61137 | 1.413806 | -0.11373 | -0.08765 | -0.37379 | -0.3422 | 0.698665 | 0.263741 | 0.536952 | -0.71159 | -0.74887 | 0.446502 | -0.30741 | -0.03755 | -0.79107 | 0.335485 | -0.36021 | -0.66354 | [illegible] |
| 827 | -0.64713 | -0.21075 | -0.12076 | 0.609408 | -0.04534 | 0.445033 | 0.057993 | -0.25603 | -0.14186 | 0.08313 | 0.213186 | -0.4675 | -0.62328 | 0.066597 | -0.2747 | -0.2826 | -0.86494 | -0.19938 | -1.08364 | -0.94792 | -0.12923 | -0.59728 | -0.328 | 0.411826 | -0.05707 | -0.20716 | -0.49217 | -0.2375 | 0.148367 | 0.522526 | 0.087748 | -0.71463 | -0.32686 | 0.264464 | -0.22246 | -0.23002 | -0.06321 | 0.099247 | -0.64717 | -0.65586 | [illegible] |
| 828 | [illegible] | | | | | | | | | | | | | | | | | | | | | | | | | | | | | | | | | | | | | | | | |
| 829 | [illegible] | | | | | | | | | | | | | | | | | | | | | | | | | | | | | | | | | | | | | | | | |
| 830 | [illegible] | | | | | | | | | | | | | | | | | | | | | | | | | | | | | | | | | | | | | | | | |

| | |
|---|---|
| 831–997 | [illegible] |

| | | | | | | | | | | | | | | | | | | | | | |
|---|---|---|---|---|---|---|---|---|---|---|---|---|---|---|---|---|---|---|---|---|---|
| 998 | -0.72398 | -0.52715 | -0.41246 | -0.27716 | 0.532244 | -0.39914 | 0.540996 | -0.38086 | -0.56582 | -0.64754 | -0.14535 | -0.35549 | -0.13265 | -0.68277 | 1.342207 | -0.18589 | -0.01768 | 0.434131 | -0.332 | 0.094216 | [illegible] |
| 999 | -0.77465 | -0.53634 | -0.18036 | -0.00105 | 0.451925 | -0.26637 | 0.281036 | -0.57238 | -0.81829 | -0.58084 | 0.026348 | -0.41519 | -0.35197 | -0.52596 | 1.326116 | -0.47533 | -0.07351 | 0.923956 | -0.49972 | 0.104832 | [illegible] |
| 1000 | -0.71686 | -0.26063 | 0.089023 | -0.0815 | 0.646023 | 0.015941 | 0.697691 | -0.71093 | -0.91368 | -0.26101 | -0.41778 | -0.10343 | 0.066827 | -0.62817 | 1.408339 | 0.017756 | -0.03176 | 0.789382 | -0.17917 | 0.086805 | [illegible] |
| 1001 | -0.89267 | 0.034542 | -0.11822 | -0.52386 | 0.752658 | 0.198193 | 0.23741 | -0.36109 | -0.51832 | -0.2983 | -0.39049 | 0.071174 | -0.54539 | -0.68796 | 1.173265 | -0.03646 | 0.215267 | 0.263044 | 0.023451 | 0.223094 | [illegible] |
| 1002 | -1.1713 | -0.30781 | -0.44231 | -0.89222 | 0.239539 | 0.142221 | -0.10842 | -0.02094 | -0.58354 | 0.477794 | -0.60337 | 0.342641 | -0.51487 | -0.95813 | 0.924669 | -0.62456 | -0.08426 | 0.222754 | -0.3688 | -0.15681 | [illegible] |
| 1003 | -1.19223 | -0.45331 | -0.92218 | -1.0985 | 0.275268 | -0.71431 | -0.34685 | 0.161517 | -0.31758 | 1.357683 | -0.54019 | 0.469274 | -0.86661 | -0.91909 | -0.24226 | -0.59919 | 0.079883 | -0.29947 | -0.81443 | -0.36491 | [illegible] |
| 1004 | -1.37925 | -0.17257 | -0.95835 | -1.5587 | -0.33657 | -0.43655 | -0.25068 | 0.158086 | 0.026693 | 1.086235 | -0.61286 | 0.424553 | -1.18878 | -0.37106 | -0.24728 | -0.05577 | -0.349 | -0.56717 | -0.98244 | -0.49984 | [illegible] |
| 1005 | -1.42088 | -0.39776 | -0.99653 | -1.50487 | -0.47073 | -0.56268 | -0.70788 | -0.15752 | 0.010479 | 0.846272 | -0.51619 | -0.02123 | -1.16273 | -0.30777 | 0.298762 | -0.90987 | -0.82538 | -0.29512 | -0.84618 | -0.46972 | [illegible] |
| 1006 | -1.41598 | -0.74947 | -0.76775 | -1.70351 | -0.96893 | -0.83144 | -0.45261 | -0.22748 | -0.27718 | 0.844609 | -0.28436 | -0.15126 | -1.51619 | -0.31236 | 0.077214 | -0.39916 | -1.364 | -0.43718 | -1.05925 | -0.48418 | [illegible] |
| 1007 | -0.88047 | -0.15886 | -0.57584 | -1.37668 | -0.39233 | -0.73377 | -0.46381 | -0.23593 | -0.5025 | 0.387532 | 0.03579 | -0.76881 | -1.67535 | 0.487218 | 0.046416 | -0.35609 | -1.69076 | -0.49718 | -0.91151 | -0.33984 | [illegible] |
| 1008 | -0.94523 | -0.15992 | -0.10697 | -1.26935 | -0.32435 | -0.74118 | -0.10731 | -0.11306 | -0.5606 | -0.40125 | -0.24076 | -0.77742 | -1.74818 | 0.759235 | 0.500839 | -0.71094 | -1.61598 | -0.48663 | -0.3903 | -0.43827 | [illegible] |
| 1009 | -0.41538 | -0.4291 | -0.38772 | -0.85211 | -0.1072 | -1.06847 | -0.30639 | -0.06271 | -0.27479 | 0.00355 | -0.12273 | -0.84954 | -1.80263 | 0.606859 | 0.625533 | -0.9404 | -1.4905 | -0.00367 | -0.18905 | -0.57061 | [illegible] |
| 1010 | -0.24945 | -0.36254 | -0.62405 | -0.60665 | -0.26713 | -1.18632 | -0.28813 | 0.603916 | -0.11794 | -0.04117 | -0.24354 | -0.79226 | -1.64095 | 0.489755 | 0.27455 | -0.25865 | -1.15795 | -0.2026 | -0.09251 | -0.40518 | [illegible] |
| 1011 | -0.20428 | -0.21216 | -0.5354 | 0.196893 | -0.07734 | -1.25338 | -0.40641 | 0.471678 | -0.18287 | -0.0431 | -0.03855 | -0.72969 | -0.57363 | 0.852604 | 0.568113 | -0.73949 | -0.70461 | -0.01804 | 0.098058 | -0.38862 | [illegible] |
| 1012 | -0.1808 | -0.46042 | -0.69983 | 0.251764 | -0.26955 | -1.51116 | -0.51641 | 0.534988 | -0.31846 | -0.36211 | -0.24933 | -0.24095 | -0.33119 | 0.296635 | 0.51076 | -0.36438 | -0.23527 | 0.157864 | 0.197432 | -0.60751 | [illegible] |
| 1013 | -0.32514 | -0.44159 | -0.90693 | 0.677352 | -0.39758 | -1.51012 | -0.78669 | 0.009455 | -0.50357 | -0.39931 | -0.1759 | -0.36835 | -0.10322 | 0.18928 | 0.271404 | -0.33812 | -0.17134 | 0.303082 | -0.11011 | -0.30363 | [illegible] |
| 1014 | -0.71181 | -0.29888 | -1.00765 | 0.993834 | -0.22899 | -1.14815 | -0.60842 | -0.07891 | -0.71985 | -0.6691 | -0.5606 | -0.11585 | -0.02338 | 0.106869 | -0.01648 | -0.34753 | 0.280488 | -0.02358 | -0.26446 | -0.13063 | [illegible] |
| 1015 | -0.61687 | -0.37028 | -0.68167 | 0.777169 | -0.12899 | -0.69813 | -0.30252 | -0.63243 | -0.63114 | -0.61511 | -0.51913 | -0.33334 | 0.088896 | 0.223973 | -0.17631 | -0.6272 | 0.47336 | -0.10632 | -0.48994 | -0.29607 | [illegible] |
| 1016 | -0.74193 | -0.2064 | -0.5728 | 0.634938 | -0.00751 | -0.33913 | 0.366869 | -0.6917 | -0.43709 | -0.48012 | -0.84882 | -0.29372 | -0.51586 | 0.042125 | -0.35013 | -0.3318 | 0.593872 | 0.015169 | -0.74494 | -0.37593 | [illegible] |
| 1017 | -0.82127 | -0.07887 | -0.29264 | 0.368755 | -0.19599 | -0.17411 | 0.690257 | -1.04298 | -0.37288 | -0.75727 | -0.61527 | -0.58986 | -0.64297 | 0.115903 | -0.45362 | -0.33254 | 0.379186 | -0.39116 | -0.84111 | -0.0978 | [illegible] |
| 1018 | -0.57325 | -0.07887 | -0.49157 | -0.06036 | 0.191746 | -0.39337 | 0.596833 | -1.06289 | -0.15782 | -0.48628 | -0.82953 | -0.58986 | -0.56415 | 0.114218 | -0.45362 | 0.035052 | 0.191107 | -0.3718 | -0.86658 | -0.52407 | [illegible] |
| 1019 | -0.33014 | -0.07887 | 0.0106 | -0.28696 | 0.191746 | -0.46448 | 0.228259 | -0.8486 | -0.15782 | -0.24406 | -0.55349 | -0.86591 | -0.31059 | 0.078988 | -0.2549 | 0.007624 | -0.43405 | -0.44492 | -0.6225 | -0.52407 | [illegible] |
| 1020 | -0.33014 | 0.127538 | 0.002469 | -0.613 | 0.191746 | -0.79087 | -0.23903 | -0.72217 | -0.15782 | -0.25332 | -0.57065 | -0.59203 | -0.44839 | 0.078988 | 0.214764 | 0.169499 | -0.43405 | -0.23769 | -0.39705 | -0.52407 | [illegible] |
| 1021 | -0.07934 | 0.127538 | -0.06823 | -0.94308 | 0.448092 | -0.79087 | -0.81381 | -0.50303 | -0.24916 | -0.25332 | -0.57065 | -0.59203 | -0.18609 | -0.18435 | 0.338682 | -0.14971 | -0.43405 | -0.30009 | -0.10625 | -0.26605 | [illegible] |
| 1022 | -0.30295 | 0 | -0.15586 | -0.67689 | 0.454763 | -0.64504 | -0.81381 | -0.54431 | 0.012733 | -0.14499 | -0.89158 | -0.26758 | -0.05898 | -0.48856 | 0.593581 | -0.25711 | -0.56892 | -0.04304 | -0.01008 | -0.62128 | [illegible] |
| 1023 | -0.63355 | 0 | 0.043081 | -1.04045 | 0.256346 | -0.32639 | -1.14413 | -0.3046 | 0 | -0.16882 | -0.61252 | -0.26758 | -0.1378 | -0.48687 | 0.593581 | -0.40984 | -0.38084 | -0.0624 | -0.00677 | -0.74706 | [illegible] |
| 1024 | -0.95932 | -0.0996 | 0.043081 | -1.04045 | 0.256346 | -0.32639 | -1.13465 | -0.3046 | 0.037436 | -0.16882 | -0.61252 | -0.32696 | -0.1378 | -0.49377 | 0.593581 | -0.70326 | -0.1685 | -0.27967 | -0.00677 | -0.74706 | [illegible] |
| 1025 | -1.17377 | -0.0996 | -0.13 | -0.71441 | 0.256346 | 0 | -1.1146 | -0.3046 | 0.066927 | -0.16882 | -0.29894 | -0.64274 | -0.33165 | -0.83046 | -0.13789 | -0.97907 | -0.1685 | -0.50557 | -0.23222 | -1.07598 | [illegible] |
| 1026 | -0.93543 | -0.32614 | -0.13 | -0.42651 | -0.17209 | -0.01039 | -1.15493 | -0.3046 | -0.24418 | -0.46141 | -0.20907 | -0.68384 | -0.53754 | -0.36998 | -0.2422 | -0.37209 | -0.14024 | -0.54215 | -0.1433 | -1.24079 | [illegible] |
| 1027 | -0.80537 | -0.17512 | 0.208105 | -0.45448 | -0.28628 | -0.31561 | -1.51232 | 0.138396 | -0.58723 | -0.50054 | -0.25584 | -0.56751 | -0.54074 | -0.15402 | 0.088167 | -0.14201 | 0.238637 | -0.27732 | 0.454119 | -0.88674 | [illegible] |
| 1028 | -0.74771 | 8.42E-05 | 0.623278 | -0.31339 | -0.35247 | -0.60884 | -1.31033 | -0.09932 | -0.68576 | -0.73293 | -0.2684 | -0.73655 | -0.73707 | -0.15402 | 0.207386 | -0.29416 | 0.198162 | -0.33142 | 0.650412 | -0.40616 | [illegible] |
| 1029 | -0.41314 | 0.261503 | 0.758942 | -0.50165 | 0.175884 | -0.81696 | -1.34799 | -0.57773 | -0.556 | -0.81385 | -0.59762 | -0.82163 | -0.86539 | -0.1409 | -0.06993 | 0.127401 | 0.255951 | 0.216149 | 0.66888 | -0.41422 | [illegible] |
| 1030 | -0.37126 | 0.380947 | 0.887795 | -0.09162 | 0.014096 | -0.91412 | -0.84547 | -0.35647 | -0.5608 | -0.93117 | -0.38997 | -0.36733 | -0.56871 | 0.423876 | -0.18105 | 0.322223 | -0.13554 | 0.203039 | 1.045866 | -0.08406 | [illegible] |
| 1031 | -0.70212 | 0.704398 | 0.887795 | -0.37254 | 0.174076 | -0.90374 | -0.75926 | -0.35647 | -0.58562 | -0.95031 | -0.21535 | -0.44443 | -0.34869 | 0.163176 | -0.02142 | 0.267071 | -0.15319 | 0.278254 | 1.200645 | 0.036946 | [illegible] |
| 1032 | -0.39577 | 0.127586 | 0.13995 | -0.604 | 0.003689 | -0.71365 | -0.44732 | -0.67943 | -0.43939 | -0.41259 | 0.324043 | -0.84745 | -0.07068 | 0.507692 | -0.38693 | 0.238279 | -0.40563 | -0.18858 | 0.620991 | -0.02614 | [illegible] |
| 1033 | -0.21441 | -0.09692 | -0.36084 | -1.02856 | -0.28414 | -1.06964 | -0.41003 | -0.58664 | -0.82752 | -0.27744 | 0.047779 | -0.63813 | 0.090544 | -0.04273 | -0.41183 | 0.512755 | -0.46019 | 0.562451 | 0.944324 | -0.53885 | [illegible] |
| 1034 | -0.26153 | -0.3737 | -0.34741 | -0.95423 | -0.57676 | -1.35287 | -0.10058 | 0.091022 | -0.80418 | 0.000748 | 0.657575 | -0.15251 | 0.124175 | -0.11593 | -0.40474 | 0.188977 | -0.6554 | 1.009399 | 0.813912 | -0.39906 | [illegible] |
| 1035 | -0.21121 | -0.11823 | -0.47652 | -1.40559 | -0.40658 | -1.25415 | -0.61117 | -0.16439 | -1.0255 | 0.091991 | 0.537332 | -0.40299 | 0.173216 | -0.37296 | -0.21029 | 0.087348 | -0.41113 | 1.870426 | 0.159653 | -0.81669 | [illegible] |
| 1036 | -0.32251 | 0.677759 | -0.33612 | -1.25006 | -0.47765 | -1.42865 | -0.69765 | -0.56378 | -0.88347 | 0.220605 | 0.279403 | -0.62988 | 0.428311 | -0.48678 | -0.43068 | 0.041037 | -0.42573 | 1.928781 | -0.31036 | -0.51106 | [illegible] |
| 1037 | -0.78965 | 1.511275 | 0.303191 | -1.16569 | -0.16349 | -1.1819 | -0.52719 | -0.35781 | -1.00016 | -0.35018 | -0.12692 | -0.5463 | 0.372359 | -1.22849 | -0.55766 | -0.52745 | -0.52566 | 1.904446 | -0.60111 | -0.35202 | [illegible] |
| 1038 | -0.83628 | 2.48654 | 0.936074 | -0.7713 | -0.07048 | -0.30496 | -0.48454 | -0.21886 | -0.60562 | -0.20353 | -0.05203 | -0.50906 | 0.567256 | -0.81859 | -0.85509 | -0.66061 | -0.31191 | 0.965852 | -0.84801 | 0.022166 | [illegible] |
| 1039 | -0.84566 | 2.717306 | 0.630255 | -0.85639 | -0.40285 | 0.273562 | -0.58879 | -0.78872 | -0.98595 | -0.46522 | -0.55682 | -0.62957 | 0.597636 | -1.07199 | -0.51352 | -0.47695 | -0.37446 | 0.046202 | -0.58277 | -0.03872 | [illegible] |
| 1040 | -0.93346 | 2.683449 | 0.402582 | -0.40763 | -0.61633 | 0.176623 | -0.37707 | -0.88796 | -1.08441 | -0.58167 | -0.60368 | -0.61539 | 0.932606 | -0.99261 | -0.51534 | -0.37483 | 0.103372 | -0.73009 | -0.47274 | 0.293564 | [illegible] |
| 1041 | -0.72964 | 1.93251 | 0.026374 | -0.24441 | -0.57681 | 0.245579 | -0.47983 | -0.2998 | -0.96869 | -0.45219 | -0.6435 | -0.47177 | 0.442597 | -1.03015 | -0.49239 | -0.73144 | 0.157132 | -0.88355 | -0.47572 | -0.26387 | [illegible] |
| 1042 | -0.48112 | 1.993972 | -0.38773 | 0.068002 | -0.59392 | 0.033652 | -0.69324 | -0.61845 | -0.95885 | -0.46453 | -0.30635 | -0.29698 | 0.466943 | -0.69993 | -0.48782 | -0.47839 | 0.183711 | -0.65839 | -0.49 | -0.35865 | [illegible] |
| 1043 | -0.60423 | 1.127822 | -1.19355 | 0.233316 | -0.52151 | -0.35676 | -0.59963 | -0.5782 | -0.81178 | -0.83441 | 0.044348 | -0.36454 | 0.359944 | -0.87626 | 0.015709 | -0.57739 | 0.067711 | -0.40859 | -0.83948 | -0.5222 | [illegible] |
| 1044 | -0.55655 | 0.754991 | -1.03683 | 0.371035 | -0.49591 | -0.44726 | -0.46357 | -0.19001 | -0.622 | -1.03344 | 0.181991 | -0.8613 | 0.281366 | -0.3327 | -0.01583 | -0.70639 | -0.00921 | -0.36098 | -1.04131 | -0.49704 | [illegible] |
| 1045 | -0.2279 | 0.413932 | -0.61865 | -0.14407 | -0.22238 | -0.37643 | -0.5613 | -0.17034 | -0.45223 | -0.6864 | 0.192102 | -0.83133 | -0.16132 | -0.19494 | -0.38176 | -0.37915 | -0.33168 | -0.35234 | -1.17222 | -0.54704 | [illegible] |
| 1046 | -0.5424 | 0.006104 | -0.48534 | -0.18633 | -0.48146 | -0.51893 | -0.96811 | -0.742 | -0.89471 | -0.94129 | 0.496351 | -1.03297 | 0.044328 | 0.086345 | -0.51213 | -0.0717 | -0.25108 | -0.30107 | -1.29666 | -0.25197 | [illegible] |
| 1047 | -0.52603 | -0.5723 | -0.14703 | -0.3769 | -0.53267 | -0.34777 | -0.77058 | -0.29599 | -0.46414 | -0.79875 | 0.436256 | -1.14558 | -0.28182 | 0.38458 | -0.25634 | -0.26927 | -0.35715 | -0.37806 | -1.0496 | -0.1266 | [illegible] |
| 1048 | -0.19116 | -0.76874 | 0.061147 | -0.11107 | 0.088594 | -0.23202 | -0.84557 | -0.46094 | -0.64776 | -0.62029 | 0.107523 | -1.33463 | -0.54311 | 0.513929 | -0.95987 | -0.56087 | -0.65394 | -0.42143 | -0.71541 | -0.17377 | [illegible] |
| 1049 | -0.41756 | -0.45683 | 0.192671 | -0.59235 | 0.01559 | -0.4954 | -1.00429 | -0.62947 | -0.64938 | 0.121436 | 0.006408 | -0.72857 | -0.81929 | -0.05842 | -1.22179 | -0.44845 | -0.64427 | -0.48191 | -1.01421 | -0.35117 | [illegible] |
| 1050 | -0.00451 | -0.4953 | 0.042238 | -0.07954 | 0.147269 | -0.62419 | -0.92288 | -0.58186 | -0.67772 | -0.16791 | -0.25368 | -0.2559 | -1.14763 | -0.46333 | -0.69 | -0.90597 | -0.93067 | -0.72385 | -0.33517 | -0.25323 | [illegible] |
| 1051 | 0.389843 | -0.30129 | 0.165378 | -0.19148 | 0.450589 | -0.50891 | -0.31338 | -0.15118 | -0.33148 | -0.18934 | -0.80263 | 0.048415 | -1.22324 | -0.60657 | -0.40957 | -0.93896 | -1.11666 | -0.87462 | -0.11564 | -0.18785 | [illegible] |
| 1052 | 0.335821 | -0.27746 | 0.033986 | -0.00281 | 0.77006 | -0.81196 | -0.7812 | -0.15877 | -0.56079 | 0.123151 | -1.09058 | 0.10142 | -1.01189 | -1.03305 | -0.75539 | -0.37472 | -0.77273 | -0.98581 | -0.11116 | -0.34894 | [illegible] |
| 1053 | 0.244561 | -0.11799 | 0.145958 | -0.42344 | 0.037382 | -1.04938 | -0.79492 | -0.29334 | -0.71817 | 0.042605 | -0.63826 | 0.149437 | -1.06802 | -0.70447 | -0.35228 | 0.127494 | -0.42471 | -0.9995 | -0.11609 | -0.22451 | [illegible] |
| 1054 | 0.407611 | -0.47984 | -0.15067 | 0.3704 | 0.342718 | -1.08757 | -0.68745 | -0.10345 | -0.68597 | -0.33891 | -0.45056 | -0.0985 | -0.77014 | -0.6566 | -0.28564 | -0.09371 | 0.098548 | -0.99608 | 0.201691 | -0.33825 | [illegible] |
| 1055 | -0.25001 | -0.44137 | -0.13803 | -0.00055 | -0.11923 | -0.96568 | -0.84333 | 0.020744 | -0.90443 | -0.29638 | -0.5666 | -0.4318 | -0.26779 | -0.43988 | -0.53895 | 0.31178 | 0.53948 | -0.69876 | -0.47008 | -0.46564 | [illegible] |
| 1056 | -0.03204 | -0.53547 | -0.16125 | 0.157994 | -0.40706 | -1.15287 | -1.33391 | -0.02705 | -0.61683 | -0.22637 | -0.30756 | -0.42681 | -0.23896 | -0.36303 | -0.18705 | 0.292062 | 0.370371 | -0.78538 | -0.4316 | -0.71575 | [illegible] |
| 1057 | -0.04577 | -0.59459 | -0.30459 | -0.18997 | -0.82923 | -0.80856 | -1.22299 | -0.29145 | -0.67396 | -0.60053 | -0.08876 | -0.66634 | 0.141909 | -0.12374 | 0.090582 | -0.23745 | 0.356043 | -0.41011 | -0.54582 | -0.50744 | [illegible] |
| 1058 | -0.24431 | -0.73965 | -0.34455 | -0.13657 | -0.29964 | -0.57635 | -1.13737 | -0.32873 | -0.31352 | -0.62753 | -0.3391 | -0.29771 | 0.58085 | -0.31808 | -0.29137 | -0.42877 | 0.093727 | -0.39488 | -0.64339 | -0.12456 | [illegible] |
| 1059 | -0.01246 | -0.49869 | -0.01372 | -0.71313 | -0.59915 | -0.59083 | -1.24003 | -0.04553 | -0.57164 | -0.42632 | -0.49603 | -0.14156 | 0.469403 | -0.21552 | -0.32005 | -0.23445 | 0.164109 | -0.36265 | -1.00033 | -0.31856 | [illegible] |
| 1060 | 0.101567 | -0.55302 | 0.074725 | -0.62534 | -0.27985 | -0.88976 | -0.87678 | -0.15355 | -0.38294 | -0.26253 | -0.13141 | -0.39972 | 0.389039 | -0.36491 | -0.18683 | -0.35401 | -0.34313 | -0.27239 | -0.69931 | -0.30201 | [illegible] |
| 1061 | -0.10207 | 0.065544 | 0.052858 | -0.93247 | 0.024174 | -0.5698 | -0.76133 | -0.38203 | -0.38052 | -0.17856 | -0.65873 | -0.18984 | 0.409044 | -0.25354 | -0.60661 | -0.54405 | -0.25689 | -0.17473 | -0.87157 | 0.50445 | [illegible] |
| 1062 | -0.24088 | 0.072646 | 0.208063 | -0.71994 | -0.09083 | -0.72714 | -0.81464 | -0.03275 | -0.26951 | 0.184112 | -0.80704 | 0.081164 | -0.28129 | -0.28313 | -0.47293 | -0.36261 | -0.49981 | -0.4552 | -1.02313 | -0.31358 | [illegible] |
| 1063 | 0.075145 | -0.10538 | 0.176988 | -0.51044 | -0.42899 | -0.98183 | -0.8406 | 0.200683 | -0.52275 | 0.794866 | -0.87268 | -0.23865 | -0.66139 | -0.50403 | -0.35201 | -0.42526 | -0.46843 | -0.4215 | -1.08315 | -0.36423 | [illegible] |
| 1064 | 0.111957 | -0.10123 | 0.243457 | -0.43334 | -0.41879 | -0.92838 | -0.39355 | -0.56083 | -0.38679 | 0.747144 | -0.7057 | -0.39673 | -0.58576 | -0.09443 | -0.17124 | -0.60244 | -0.62495 | -0.66465 | -0.95926 | -0.34728 | [illegible] |
| 1065 | -0.12363 | 0.104037 | 0.064803 | -0.94807 | -0.45571 | -0.58008 | -0.46414 | -0.57394 | -0.52537 | 0.4438 | -0.59939 | -0.3386 | -0.51978 | -0.1346 | -0.25611 | -0.6075 | -0.28877 | -0.93981 | -1.18216 | -0.23354 | [illegible] |
| 1066 | -0.5509 | -0.19792 | 0.253771 | -0.61731 | -0.78545 | -0.82829 | -0.28626 | -0.26461 | -0.86474 | 0.68279 | 0.324181 | -0.66962 | -0.83821 | 0.081633 | -0.21817 | -0.72342 | -0.18056 | -0.7572 | -1.35342 | -0.91645 | [illegible] |
| 1067 | -0.19907 | -0.20308 | 0.996663 | -0.78619 | -0.74447 | -0.76077 | -0.14081 | -0.35202 | -0.74067 | 0.464344 | 0.384064 | -0.74156 | -0.49775 | -0.06491 | -0.19885 | -0.73331 | -0.22396 | -0.56225 | -1.03665 | -0.18174 | [illegible] |
| 1068 | -0.5345 | 0.231882 | 1.298139 | -1.09348 | -0.77192 | -0.59982 | 0.001314 | -0.4496 | -0.53938 | -0.02678 | 0.209189 | -0.66674 | -0.48276 | -0.03294 | 0.065491 | -0.57965 | -0.19677 | -0.65685 | -0.98748 | -0.77271 | [illegible] |
| 1069 | -0.9484 | 0.482827 | 1.197253 | -0.36339 | -1.42945 | -0.58895 | -0.60946 | -0.11159 | -0.55182 | 0.073379 | 0.056386 | -0.70178 | -0.35255 | -0.22453 | 0.068891 | -0.42013 | -0.54127 | -0.46486 | -0.77191 | -0.1577 | [illegible] |
| 1070 | -0.66747 | 0.331889 | 1.529946 | -0.31209 | -1.34211 | -0.84133 | -0.65425 | -0.0644 | -0.83302 | 0.548131 | -0.03115 | -0.72886 | -0.24461 | -0.48489 | -0.00399 | -0.12492 | -0.53286 | -0.62411 | -0.31312 | -0.25355 | [illegible] |
| 1071 | -0.45213 | 0.217051 | 1.181011 | -0.30627 | -1.20301 | -1.23731 | -0.66962 | 0.031464 | -0.7244 | 0.466079 | -0.91923 | -0.56275 | -0.49019 | -0.96194 | -0.09214 | -0.02109 | -0.62779 | -0.5854 | -0.26107 | -0.25401 | [illegible] |
| 1072 | -0.72952 | 0.35962 | 0.134694 | -0.49175 | -0.75586 | -1.01457 | -0.36823 | -0.10716 | -0.96994 | 0.183157 | -0.52292 | -1.26209 | -0.25299 | -1.01718 | -0.28626 | -0.07107 | -0.44957 | -1.00456 | -0.24451 | -0.35486 | [illegible] |
| 1073 | -0.38568 | 0.006693 | -0.11376 | -0.18445 | -1.00277 | -0.96576 | -0.59288 | -0.11236 | -1.14558 | 0.265163 | -0.65425 | -1.59235 | 0.242625 | -1.15659 | -0.37276 | -0.41824 | -0.42195 | -0.90996 | -0.11744 | 0.039956 | [illegible] |
| 1074 | 0.098612 | -0.42938 | -0.48887 | -1.07819 | -0.2172 | -0.90802 | -0.21976 | -0.42225 | -1.15754 | -0.33888 | -0.51149 | -1.39418 | 0.239303 | -1.24746 | -0.64205 | -0.31768 | -0.24736 | -0.73918 | -0.07041 | -0.59771 | [illegible] |
| 1075 | 0.117329 | -0.70712 | -0.38039 | -1.02705 | -0.61186 | -0.87915 | -0.15617 | -0.75326 | -0.70385 | -0.6493 | -0.73187 | -1.34024 | -0.09703 | -0.89851 | -0.43038 | -0.89693 | -0.32409 | -0.5822 | -0.4453 | -0.81375 | [illegible] |
| 1076 | -0.07629 | -0.65086 | -0.04072 | -1.05651 | -0.438 | -0.23497 | -0.00288 | -0.92998 | -0.58871 | -0.83486 | -0.22112 | -1.29142 | 0.508874 | -0.74906 | -0.44417 | -0.78623 | -0.4735 | -0.59619 | -0.09704 | -0.75158 | [illegible] |
| 1077 | 0.099032 | -0.65086 | 0.22845 | -0.70215 | -0.64308 | -0.87532 | -0.23167 | -1.22418 | -0.67901 | -0.44606 | -0.50229 | -0.49199 | 0.057143 | -0.5168 | -0.4331 | -0.63081 | -0.71606 | -0.27743 | -0.12886 | -0.74001 | [illegible] |
| 1078 | 0.11186 | -0.16623 | 0.184867 | -0.70215 | -0.26015 | -0.84309 | 0.085161 | -1.05233 | -0.52581 | -0.30135 | -0.33628 | -0.38452 | -0.45484 | -0.31426 | -0.3499 | -0.28413 | -0.18779 | -0.27743 | -0.16364 | -0.97463 | [illegible] |
| 1079 | -0.05446 | 0.193825 | 0.661767 | -0.48554 | -0.43109 | -0.87799 | 0.085161 | -0.45829 | -0.4442 | -0.0477 | -0.40586 | -0.27527 | -0.42474 | 0.219067 | 0.064058 | -0.48111 | -0.2479 | -0.06669 | -0.16364 | -0.8156 | [illegible] |
| 1080 | -0.01477 | 0.662809 | 0.611448 | 0.224136 | -0.14433 | -0.91699 | -0.14281 | -0.11141 | -0.29466 | -0.03017 | -0.29058 | -0.39373 | 0.021535 | 0.61058 | 0.144673 | -0.20398 | -0.19206 | 0.072101 | -0.27769 | -0.50295 | [illegible] |
| 1081 | 0.328323 | 0.690999 | 0.341703 | 0.196384 | -0.08025 | -0.86822 | -0.2885 | -0.45594 | -0.12443 | 0.038097 | -0.27299 | -0.59565 | 0.006633 | 0.435147 | -0.16962 | -0.05467 | 0.489384 | -0.21398 | -0.27769 | -0.83288 | [illegible] |
| 1082 | 0.087302 | 1.050126 | 0.319012 | 0.081701 | -0.34256 | -0.64473 | -0.30101 | -0.41393 | 0.227763 | -0.26585 | -0.01038 | -0.5675 | 0.082629 | 0.912699 | -0.16962 | -0.31597 | 0.62045 | -0.21398 | -0.42043 | -0.94557 | [illegible] |
| 1083 | -0.03369 | 0.806893 | 0.319012 | 0.081701 | 0.157987 | -0.5799 | -0.54943 | -0.36511 | 0.227763 | -0.40127 | -0.12709 | -0.47312 | 0.239459 | 1.681091 | -0.42479 | 0.157921 | 0.004428 | -0.42235 | -0.66027 | -0.71095 | [illegible] |
| 1084 | -0.14306 | 0.872077 | 0.085909 | 0.361751 | 1.058578 | -0.66872 | -0.8446 | -0.3623 | 0.262119 | -0.65839 | -0.18456 | -0.14831 | 0.239459 | 1.367693 | -0.84545 | 0.577787 | 0.070687 | -0.85791 | -0.60435 | -0.57742 | [illegible] |
| 1085 | 0.186708 | 0.691243 | -0.3329 | -0.34963 | 1.058578 | -0.41005 | -0.62777 | -0.3623 | -0.24214 | -0.89798 | -0.28808 | 0.111305 | -0.39178 | 0.527057 | -0.77582 | 0.577787 | 0.091582 | -0.92517 | -0.36145 | -0.54492 | [illegible] |
| 1086 | 0.055771 | 0.45007 | -0.37252 | -0.44483 | 0.67922 | -0.6547 | -0.54332 | -0.07017 | -0.55029 | -0.76164 | -0.52395 | 0.39163 | -0.40542 | 0.606699 | -0.46651 | 0.621893 | -0.31525 | -0.66565 | -0.47381 | -0.60829 | [illegible] |
| 1087 | 0.049122 | 0.225193 | -0.62716 | -0.59157 | 1.183867 | -0.76103 | -0.32233 | 0.146578 | -0.56092 | -0.79176 | -1.09951 | 0.069346 | -0.24858 | -0.17809 | -0.74627 | 0.883193 | 0.164026 | -0.89423 | -0.49694 | -0.20529 | [illegible] |
| 1088 | -0.15759 | -0.02648 | -0.79822 | -0.88047 | 0.347109 | -0.6833 | -0.14397 | 0.000264 | -0.21646 | -0.89301 | -0.96933 | -0.1229 | -0.23562 | -0.99475 | -0.35849 | 0.148956 | 0.396649 | -0.59853 | -0.54612 | -0.30468 | [illegible] |
| 1089 | -0.51644 | -0.41523 | -0.64425 | 0.165669 | -0.32779 | -0.28761 | -0.12857 | -0.37653 | -0.23638 | -0.53073 | -1.0903 | -0.46153 | -0.36997 | -1.26053 | -0.14148 | -0.03036 | 0.613663 | -0.42691 | -0.77625 | -0.39379 | [illegible] |
| 1090 | -1.12219 | -0.71131 | -0.71005 | 0.202835 | -0.22554 | -0.5572 | -0.09627 | -0.65829 | 0.593756 | -0.29115 | -0.79719 | -0.62907 | 0.281133 | -1.07885 | -0.25414 | -0.17009 | 0.710448 | -0.48355 | -0.83136 | -0.6024 | [illegible] |
| 1091 | -0.98219 | -0.755 | -0.92125 | 0.054591 | 0.211212 | -0.17149 | -0.42129 | -0.46128 | 0.731743 | -0.39131 | -0.82618 | -0.56631 | 0.015431 | -0.80327 | -0.52644 | -0.28217 | 0.72697 | -0.31717 | -0.97964 | -0.2268 | [illegible] |
| 1092 | -0.84426 | -1.21572 | -0.99844 | 0.229943 | -0.11209 | -0.31419 | -0.58455 | -0.42768 | 0.49827 | -0.39939 | -0.77599 | -0.0802 | -0.46605 | -0.31555 | -0.23516 | -0.12669 | 0.167701 | 0.139809 | -0.81653 | -0.35838 | [illegible] |
| 1093 | -0.57694 | -0.95457 | -1.01041 | 0.391142 | 0.027494 | -0.66316 | -0.52668 | -0.76042 | -0.20117 | -0.33844 | -0.2984 | 0.286683 | -0.549 | -0.13867 | -0.60744 | -0.03149 | 0.05175 | 0.032044 | -0.4505 | 0.126304 | [illegible] |
| 1094 | -0.31381 | -0.91114 | -1.51026 | -0.97374 | -0.30637 | -0.79246 | -0.55572 | -0.66985 | -0.31614 | -0.49487 | 0.090908 | 0.202433 | 0.011258 | -0.07205 | -0.58202 | -0.1234 | -0.2578 | 0.007998 | -0.22585 | -0.26496 | [illegible] |
| 1095 | 0.646523 | -0.61618 | -1.38892 | -0.88533 | -0.55157 | -0.88041 | -0.8542 | -0.59661 | -1.00607 | -0.70903 | -0.11174 | 0.510886 | -0.34115 | 0.082483 | -0.55641 | -0.03483 | -0.21582 | 0.050125 | -0.21837 | -0.23908 | [illegible] |
| 1096 | 0.839661 | -0.22283 | -1.24693 | -0.89299 | -0.91981 | -1.61508 | -0.73354 | -1.24362 | -1.18708 | -0.60886 | 0.135529 | 0.413091 | 0.339671 | -0.35772 | -0.29397 | -0.28714 | -0.46667 | -0.2718 | -0.19403 | -0.47882 | [illegible] |
| 1097 | 0.749977 | 0.103649 | -0.89241 | -0.9407 | -0.71988 | -1.2651 | -0.9281 | -1.21702 | -1.03488 | -0.52109 | 0.398298 | 0.406314 | 0.385714 | -0.68651 | -0.3055 | -0.65915 | -0.49181 | -0.5002 | -0.07874 | -0.75362 | [illegible] |
| 1098 | 0.689377 | -0.12853 | -0.70938 | -1.06572 | -0.52325 | -0.99386 | -1.07296 | -0.83677 | -0.82114 | -0.44217 | 0.08106 | 0.299136 | 0.365084 | -0.63606 | 0.08991 | -0.57394 | -0.4927 | -0.67304 | -0.31922 | -1.36222 | [illegible] |
| 1099 | 0.808037 | -0.08791 | -0.45837 | -0.82261 | -0.2297 | -0.9392 | -0.76415 | -0.8225 | -0.78771 | -0.3909 | 0.07366 | 0.19819 | 0.005313 | -0.53635 | -0.00473 | -0.77874 | -0.67816 | -0.88947 | -0.65433 | -0.62844 | [illegible] |
| 1100 | 0.096333 | -0.25355 | -0.07505 | -0.68671 | -0.22386 | -0.6328 | -0.40249 | -0.614 | -0.31336 | -0.04607 | 0.111713 | -0.10093 | 0.02964 | -0.48363 | -0.13095 | -0.9877 | -0.81692 | -0.44737 | -0.7803 | -0.41399 | [illegible] |
| 1101 | 0.123238 | -0.5474 | 0.12467 | -0.53247 | -0.19627 | -0.3932 | 0.034712 | -0.01938 | -0.46028 | -0.11878 | -0.25223 | -0.1443 | -0.6284 | 0.840711 | -0.35924 | -0.92119 | -0.91123 | -0.38543 | -0.96219 | -0.30492 | [illegible] |
| 1102 | 0.219469 | -0.71184 | -0.09616 | -0.37373 | -0.824 | -0.65201 | 0.148482 | -0.15722 | -0.51205 | -0.1088 | -0.27398 | -0.52293 | -0.83491 | 1.597277 | -0.03684 | -0.89997 | -0.85908 | -0.36495 | -0.86494 | -0.28413 | [illegible] |
| 1103 | 0.450017 | -0.52485 | -0.2287 | -0.30018 | -0.50358 | -1.19567 | 0.060925 | -0.14021 | -0.48742 | -0.14795 | -0.53856 | -0.60816 | -0.78193 | 1.4381 | 1.718827 | -1.1832 | -0.7504 | -0.11475 | -0.77238 | -0.31354 | [illegible] |
| 1104 | 0.08397 | -0.39841 | -0.16705 | 0.530312 | -0.66544 | -1.22406 | -0.32244 | -0.01145 | -0.47462 | -0.47516 | -0.55495 | -0.61019 | -1.03087 | 1.51304 | 3.432489 | -1.25228 | -0.72565 | 0.317449 | -0.46044 | -0.85899 | [illegible] |
| 1105 | 0.114329 | -0.34633 | -0.40843 | 0.464706 | -0.72601 | -1.03803 | -0.54108 | -0.05846 | -0.64908 | -0.56307 | -0.63406 | -0.53836 | -1.12377 | 1.636089 | 3.704352 | -0.66497 | -0.62224 | -0.08684 | -0.28684 | -0.82948 | [illegible] |
| 1106 | -0.06623 | -0.40252 | -0.60086 | 0.789792 | -0.76839 | -0.96193 | -0.17286 | -0.42976 | -0.41025 | -0.74309 | -0.38285 | -0.8624 | -1.12 | 0.501644 | 4.38785 | 0.264092 | -0.48043 | -0.30017 | -0.20777 | -0.72904 | [illegible] |
| 1107 | -0.00435 | -0.49123 | -0.15077 | 0.519248 | -0.14143 | -0.63144 | -0.09747 | -0.32853 | -0.28177 | -0.95186 | -0.45732 | -0.57761 | -0.27173 | 0.105239 | 4.478244 | 0.543825 | -0.21985 | -0.66938 | -0.40849 | -1.0942 | [illegible] |
| 1108 | -0.41335 | -0.91662 | 0.035944 | 0.549326 | -0.57322 | 0.101224 | 0.023716 | -0.40849 | -0.03804 | -0.43611 | -0.56653 | -0.82712 | -0.53948 | 0.27581 | 2.217377 | 0.563899 | -0.40292 | -0.89625 | -0.40469 | -1.10085 | [illegible] |
| 1109 | -0.12425 | -1.15123 | -0.00906 | -0.38253 | -0.22329 | -0.05989 | 0.519701 | -0.79429 | 0.081178 | -0.09743 | -0.57339 | -0.62928 | -0.36674 | 0.080563 | 0.543106 | 0.899758 | -0.32607 | -1.01402 | -0.59292 | -0.47239 | [illegible] |
| 1110 | -0.43498 | -1.40141 | -0.05509 | -0.19632 | -0.08392 | -0.09653 | 0.70794 | -0.91287 | 0.067765 | -0.19185 | -0.45107 | -0.80947 | -0.17257 | -0.0779 | 0.37514 | 0.426513 | -0.32848 | -0.92407 | -0.85717 | -0.46269 | [illegible] |
| 1111 | -0.32644 | -1.56815 | -0.11248 | -0.30272 | 0.078677 | -0.53517 | 0.039075 | -0.24339 | -0.07096 | -0.08191 | -0.50458 | -0.161 | 0.183653 | -0.06213 | -0.03901 | -0.2509 | -0.48745 | -0.63372 | -0.84498 | -0.55918 | [illegible] |
| 1112 | -0.35878 | -1.35676 | -0.60565 | -0.31213 | -0.05283 | -1.48562 | -0.11125 | -0.28247 | -0.07975 | 0.337561 | -0.32367 | -0.23866 | -0.26947 | -0.61821 | -0.15832 | -0.53183 | -0.78172 | -0.41313 | -0.82355 | -0.22597 | [illegible] |
| 1113 | -0.5066 | -1.07339 | -0.95594 | -0.03136 | -0.27121 | -1.6715 | -0.48314 | 0.155386 | -0.30547 | -0.36645 | 0.05012 | 0.093249 | 0.114578 | -0.96768 | -0.24158 | -0.11887 | -0.63053 | -0.30909 | -0.94551 | 0.333576 | [illegible] |
| 1114 | -0.45261 | -0.50668 | -0.93319 | 0.053807 | -0.74463 | -1.85909 | -0.61136 | 0.183543 | -0.62872 | -0.22237 | -0.31733 | -0.28956 | 0.074413 | -0.87642 | -0.2435 | -0.41203 | -0.35217 | -0.63901 | -0.81786 | 0.099461 | [illegible] |
| 1115 | -0.23668 | -0.40334 | -0.88894 | -0.03625 | -0.68632 | -1.9714 | -1.01484 | 0.178505 | -0.55116 | -0.45221 | -0.07919 | -0.04268 | -0.0025 | -1.08734 | -0.46794 | -0.26598 | -0.77127 | -0.57411 | -0.64473 | 0.028521 | [illegible] |
| 1116 | -0.55014 | -0.09462 | -0.90016 | -0.54792 | -0.96084 | -1.30364 | -0.88578 | -0.41524 | -0.72056 | -0.30942 | -0.2765 | -0.36711 | 0.102863 | -1.1572 | -0.29798 | -0.77224 | -0.42549 | -0.53095 | -0.31781 | 0.172948 | [illegible] |
| 1117 | -0.76568 | -0.12102 | -0.54962 | -0.46995 | -1.31145 | -0.71995 | -0.69226 | -0.71615 | -0.95367 | -0.35337 | -0.69303 | -0.3808 | -0.41647 | -0.84344 | -0.59146 | -0.40944 | -0.22732 | -0.41759 | -0.2657 | 0.300192 | [illegible] |
| 1118 | -0.63409 | -0.24997 | -0.29777 | -0.73152 | -0.78288 | -0.91212 | -0.55084 | -1.34859 | -0.67982 | -0.27941 | -1.02647 | -0.60848 | -0.69847 | -0.50537 | -0.03787 | -0.55728 | -0.50378 | -0.70632 | -0.04183 | -0.19368 | [illegible] |
| 1119 | -0.77151 | -0.75752 | -0.2423 | -0.82135 | -0.83272 | -0.77328 | -0.94536 | -1.1926 | -0.54632 | 0.508601 | -0.70468 | -0.29889 | -0.95092 | -0.87233 | 0.227348 | -0.23834 | -0.73739 | -0.48292 | -0.09467 | -0.31889 | [illegible] |
| 1120 | -0.65888 | -0.67346 | -0.51142 | -0.91092 | -0.97542 | -0.75179 | -0.32809 | -1.03679 | -0.54632 | 1.173618 | -0.96387 | -0.58601 | -0.99831 | -0.79183 | 0.444317 | -0.23834 | -0.58336 | -0.45517 | -0.07877 | -0.71494 | [illegible] |
| 1121 | -0.75601 | -0.69141 | -0.12822 | -0.79144 | -0.51286 | -0.78563 | -0.48562 | -0.9068 | -0.087 | 1.071335 | -0.54615 | -0.25598 | -1.44397 | -0.887 | 0.049294 | -0.09673 | -0.78659 | -0.50008 | 0.112739 | -0.67092 | [illegible] |
| 1122 | -0.64692 | -0.72109 | -0.31883 | -0.74901 | -0.19129 | -0.43965 | -0.48562 | -0.66408 | 0.078185 | 1.237422 | -0.39282 | -0.18078 | -0.93709 | -0.70289 | 0.189197 | -0.20731 | -0.89594 | -0.57664 | 0.230149 | -0.73326 | [illegible] |
| 1123 | -0.3039 | -0.66361 | -0.05534 | -0.53951 | -0.6198 | -0.2506 | -0.54837 | -0.43658 | -0.35679 | 1.149657 | -0.04672 | 0.382456 | -0.97891 | -0.77674 | 0.581441 | -0.39917 | -0.25255 | -0.27223 | 0.171385 | -0.83006 | [illegible] |
| 1124 | -0.08282 | -0.40608 | -0.21102 | -0.42804 | -0.32759 | -0.13709 | -0.12874 | -0.26249 | -0.22408 | 0.694604 | -0.24288 | 0.704837 | -0.55692 | -0.36062 | 0.542443 | -0.37224 | -0.48204 | -0.14422 | 0.203515 | -0.70489 | [illegible] |
| 1125 | -0.15915 | -0.1479 | -0.09466 | -0.59041 | -0.24171 | -0.30657 | -0.43825 | -0.35894 | -0.20152 | 0.188418 | -0.37654 | 1.214237 | -0.51754 | -0.1488 | 0.734046 | -0.52873 | -0.3294 | 0.049674 | 0.268401 | -0.42082 | [illegible] |
| 1126 | 0.234336 | -0.30679 | -0.42667 | 0.011471 | -0.35517 | 0.070928 | -0.018 | -0.06679 | -0.37082 | 0.597527 | -0.4492 | 0.777292 | -0.23548 | 0.165973 | 0.436784 | -0.2452 | -0.22625 | 0.222249 | 0.63735 | -0.43628 | [illegible] |
| 1127 | 0.314098 | -0.17078 | -0.4693 | 0.097741 | -0.48354 | 0.532742 | -0.00201 | 0.082141 | -0.44897 | 0.517118 | -0.29936 | 0.945605 | -0.18985 | 0.497344 | 0.940324 | -0.47575 | -0.32693 | 0.21547 | 0.551119 | -0.42238 | [illegible] |
| 1128 | 0.07398 | -0.06049 | -0.56827 | -0.07809 | -0.25426 | 0.478248 | 0.173102 | 0.162168 | -0.25375 | 0.62262 | -0.40114 | 0.674584 | -0.00086 | 0.799461 | 0.563265 | -0.36977 | -0.62689 | 0.265696 | 0.611431 | 0.15325 | [illegible] |
| 1129 | 0.144093 | -0.31263 | -0.16655 | -0.33442 | -0.39707 | 0.029409 | 0.261904 | -0.07798 | -0.34659 | -0.1788 | -0.6091 | 0.113815 | -0.1256 | 0.249472 | 0.106431 | -0.46392 | -0.20904 | 0.131649 | 0.871021 | 0.229111 | [illegible] |
| 1130 | 0.10227 | -0.11804 | -0.07833 | 0.205413 | -0.3244 | 0.231285 | 0.483111 | -0.16899 | -0.23958 | -0.17024 | 0.02707 | -0.08338 | -0.10838 | 0.350927 | -0.20528 | -0.59794 | -0.26294 | -0.29735 | 0.948431 | 0.18645 | [illegible] |
| 1131 | -0.2906 | 0.006864 | 0.099268 | 0.077288 | -0.36647 | -0.20591 | 0.390262 | -0.36768 | -0.3441 | 0.299619 | 0.016933 | 0.096186 | -0.62154 | -0.03835 | 0.01914 | -0.78095 | -0.28443 | -0.56004 | 0.387967 | 0.350536 | [illegible] |
| 1132 | -0.32393 | -0.30623 | 0.165999 | 0.086335 | -0.32919 | -0.74797 | -0.14302 | -0.55852 | -0.39982 | 0.401121 | 0.0175 | -0.18937 | -0.56555 | -0.65595 | -0.77194 | -0.65846 | -0.23569 | -0.34358 | 0.156183 | 0.279278 | [illegible] |
| 1133 | -0.16722 | -0.14123 | -0.14867 | 0.152691 | -0.55681 | -0.94948 | -0.63117 | -0.62961 | -0.06821 | 0.439411 | 0.106619 | -0.06356 | -0.44435 | -0.94979 | -1.16021 | -0.22879 | -0.32017 | -0.59841 | -0.0286 | 0.239476 | [illegible] |
| 1134 | -0.67341 | -0.31101 | -0.40811 | 0.039038 | 0.092703 | -0.36064 | -1.00886 | -0.97633 | 0.130089 | 0.456689 | 0.247341 | -0.04446 | -1.13308 | -0.67128 | -0.89629 | -0.37644 | -0.63362 | -0.64709 | -0.27532 | -0.06902 | [illegible] |
| 1135 | -0.5655 | -0.68569 | -0.43725 | -0.34791 | 0.102548 | -0.68406 | -0.79659 | -1.17773 | -0.08433 | 0.596394 | -0.44774 | 0.054041 | -0.71073 | -0.69318 | -0.39577 | -0.29726 | -1.06615 | -0.53959 | -0.29789 | -0.39968 | [illegible] |
| 1136 | -0.50581 | -0.5729 | -0.6987 | -0.56026 | 0.291961 | -0.72472 | -0.95358 | -1.11486 | 0.032662 | -0.31448 | -0.09247 | -0.03786 | -0.45019 | -0.52889 | 0.010577 | -0.30164 | -0.71593 | -0.47091 | -0.35569 | -0.72149 | [illegible] |
| 1137 | -0.50152 | -0.35097 | -0.9469 | -0.53103 | 0.544167 | -0.77168 | -0.61954 | -0.60693 | 0.203392 | -0.81862 | -0.46591 | 0.018012 | -0.11193 | -0.66852 | 0.093071 | 0.027235 | -0.75052 | -0.28974 | -0.56045 | -0.58565 | [illegible] |
| 1138 | -0.65503 | -0.46892 | -0.83492 | -0.70428 | 0.591796 | -0.57115 | -0.2675 | -0.38562 | -0.05332 | -0.75407 | -0.5886 | -0.34939 | 0.286577 | -0.96855 | 0.424453 | 0.303048 | -0.50555 | -0.04195 | -0.82056 | -1.05082 | [illegible] |
| 1139 | -0.37029 | -0.66433 | -0.76305 | -0.47094 | -0.10308 | -0.47525 | -0.1161 | 0.227354 | -0.31852 | -0.68902 | -0.40801 | 0.213198 | 0.911 | -0.72991 | 0.948801 | 0.453012 | -0.44368 | 0.01547 | -0.92541 | -0.64463 | [illegible] |
| 1140 | -0.69561 | -0.63953 | -0.70656 | -0.5222 | -0.2236 | -0.17827 | -0.57914 | 0.636889 | -0.13158 | -0.70402 | -0.23229 | 0.557675 | 0.465274 | -0.4702 | 0.607684 | 0.504469 | -0.05905 | 0.221745 | -1.29923 | -0.17225 | [illegible] |
| 1141 | -0.41542 | -0.83872 | -0.7141 | -0.30086 | -0.32332 | -0.28495 | -0.7581 | 0.517815 | 0.153873 | -0.80357 | -0.58567 | 0.374661 | 0.678415 | -0.62294 | 0.350939 | 0.594461 | -0.43998 | 0.537668 | -1.67149 | -0.3611 | [illegible] |
| 1142 | -0.34131 | -1.2679 | -1.05725 | -0.05969 | -0.70222 | -0.02034 | -0.60258 | -0.09797 | 0.188126 | -0.43931 | -0.64394 | 0.080068 | 0.286639 | -0.90653 | 0.727892 | 0.316156 | -0.1844 | 0.04312 | -1.44967 | -0.18886 | [illegible] |
| 1143 | -0.08417 | -1.33879 | -1.54172 | 0.023856 | -0.45273 | 0.198791 | -0.19006 | -0.27696 | -0.05417 | -0.68322 | -0.38654 | 0.448244 | -0.1354 | -0.94017 | 0.035131 | -0.21384 | -0.44617 | -0.03741 | -1.03096 | 0.160572 | [illegible] |
| 1144 | -0.32481 | -1.32068 | -1.85351 | -0.08149 | -0.38558 | -0.34521 | -0.10447 | -0.49122 | 0.283089 | -0.47411 | -0.37618 | -0.21217 | -0.13519 | -1.48649 | -0.53439 | -0.17878 | -0.00996 | 0.154848 | -0.8346 | 0.145594 | [illegible] |
| 1145 | -0.28433 | -1.18859 | -2.14583 | 0.108155 | -0.16907 | -0.65358 | -0.29599 | -0.68515 | 0.208112 | -0.58139 | -0.34634 | -0.36931 | -0.21057 | -1.81794 | -0.88507 | -0.39956 | 0.366542 | 0.070587 | -0.52917 | 0.240141 | [illegible] |
| 1146 | -0.34526 | -0.77809 | -1.20555 | -0.33569 | 0.049342 | -0.5526 | -0.25639 | -0.9221 | 0.016158 | -0.55061 | -0.45689 | -0.17544 | -0.34015 | -1.91127 | -0.8067 | -0.35738 | 0.833776 | -0.44498 | -0.65559 | 0.439626 | [illegible] |
| 1147 | -0.80515 | -0.13239 | -0.37112 | -0.51676 | -0.0475 | -1.00222 | 0.042179 | -0.60034 | -0.18438 | -0.65943 | -0.37203 | -0.15668 | -0.65166 | -1.65462 | -1.36392 | -0.56915 | 0.454714 | -0.54541 | -0.98893 | 0.155188 | [illegible] |
| 1148 | -0.87425 | -0.08555 | 0.105864 | -0.19468 | -0.19829 | -1.19924 | -0.28389 | -0.45552 | -0.17134 | -0.73911 | -1.09115 | -0.18247 | -0.66141 | -1.41467 | -1.10466 | -0.78513 | 0.210936 | -0.35918 | -1.2187 | 0.23567 | [illegible] |
| 1149 | -0.5998 | 0.192363 | 0.111423 | 0.039685 | -0.04871 | -0.76641 | -0.24151 | -0.01361 | -0.42188 | -0.71763 | -1.4582 | 0.11599 | -0.84467 | -1.18337 | -1.1356 | -0.79468 | -0.12023 | -0.58336 | -1.4671 | 0.076384 | [illegible] |
| 1150 | -0.54522 | 0.124459 | 0.138935 | 0.055966 | -0.65308 | -0.62676 | -0.12429 | 0.159933 | -0.76456 | -0.85332 | -1.57357 | -0.15057 | -1.06122 | -1.39777 | -0.88738 | -0.26625 | -0.45382 | -0.77168 | -1.4138 | -0.10618 | [illegible] |
| 1151 | -1.04791 | -0.58156 | -0.67087 | 0.110334 | -1.13978 | -0.70574 | -0.29096 | 0.104325 | -1.21742 | -0.99309 | -1.863 | -0.47275 | -1.01046 | -1.11885 | -0.72992 | -0.07704 | -0.76877 | -0.83071 | -0.97665 | -0.17276 | [illegible] |
| 1152 | -0.96301 | -0.99712 | -1.00878 | -0.10746 | -0.96161 | -0.68327 | -0.71091 | 0.024949 | -0.97784 | -1.28266 | -1.42909 | -0.35121 | -1.07384 | -1.04469 | -0.19072 | 0.020864 | -0.83722 | -0.38031 | -0.58268 | 1.252304 | [illegible] |
| 1153 | -1.34951 | -0.90038 | -1.19231 | -0.78167 | -0.99703 | -0.59918 | -0.69848 | -0.08265 | -0.89366 | -1.00261 | -0.13447 | -0.9454 | -0.94754 | -1.44977 | 0.242589 | 0.313021 | -0.41736 | -0.50721 | -0.33686 | 1.21499 | [illegible] |
| 1154 | -1.49232 | -1.00696 | -0.69387 | -0.81098 | -1.24889 | -0.60552 | -0.47536 | -0.46694 | -0.57403 | -1.07984 | 0.264488 | -1.31849 | -0.75865 | -1.09418 | 0.24917 | 0.069701 | -0.09781 | -0.14453 | -0.14362 | 1.423922 | [illegible] |
| 1155 | -1.46899 | -0.61843 | -0.05995 | -1.12768 | -0.77781 | -0.37537 | -0.25272 | -0.62018 | -0.28216 | -0.8603 | 0.37281 | -1.31753 | -1.15503 | -0.71625 | 0.380459 | 0.041591 | -0.01357 | -0.52655 | -0.20296 | 1.276421 | [illegible] |
| 1156 | -1.08341 | -0.47712 | 0.083005 | -0.99904 | -0.38245 | -0.70013 | 0.200394 | -0.22716 | -0.16691 | -0.44499 | 0.468901 | -1.13374 | -0.73036 | -0.61908 | 0.150057 | 0.083714 | -0.01419 | -0.29615 | -0.07554 | 1.129562 | [illegible] |
| 1157 | -0.52499 | -0.2251 | -0.0055 | -0.82919 | -0.31845 | -0.63666 | 0.360377 | -0.40112 | -0.21634 | -0.35987 | 0.385675 | -1.15915 | -0.61911 | -0.50775 | -0.21288 | 0.25142 | 0.044223 | -0.45109 | -0.03522 | -0.23851 | [illegible] |
| 1158 | -0.43685 | -0.07565 | 0.30819 | -0.56593 | -0.18196 | -0.54368 | 0.61825 | -0.39797 | -0.21607 | -0.28147 | -0.35887 | -0.85603 | -0.91542 | 0.334929 | -0.22446 | -0.55399 | -0.03763 | -0.62054 | -0.01848 | -0.73482 | [illegible] |
| 1159 | -0.48833 | -0.07758 | -0.0634 | -0.8716 | -0.25771 | -0.70422 | -0.06797 | -0.21786 | -0.43619 | -0.29959 | -0.74399 | -0.63872 | -1.23122 | 0.540824 | -0.38884 | -0.34624 | -0.15708 | -0.90999 | -0.19211 | -0.97916 | [illegible] |
| 1160 | -0.53168 | -0.43875 | -0.1658 | -0.43911 | -0.64341 | -0.96615 | -0.05132 | -0.28197 | -0.60169 | -0.42136 | -0.91047 | -0.67626 | -0.63166 | 0.435119 | -0.28755 | -0.52245 | -0.11923 | -0.48832 | -0.52203 | -0.66435 | [illegible] |
| 1161 | -0.45753 | -0.18814 | -0.23228 | -0.36689 | -0.62836 | -0.54859 | -0.23165 | -0.25122 | -0.5634 | -0.55686 | -0.8676 | -0.39479 | -1.04582 | 0.13827 | 1.114774 | -0.91923 | -0.16229 | -0.59502 | -0.59842 | -0.31399 | [illegible] |
| 1162 | -0.62953 | 0.054633 | 0.19424 | -0.36102 | -0.35631 | -0.32857 | -0.58373 | -0.16544 | -0.97263 | -0.22779 | -0.95292 | -0.26276 | -0.72643 | 0.107853 | 1.233658 | -0.9659 | 0.003395 | -0.5947 | -0.69559 | -0.50983 | [illegible] |
| 1163 | -0.5867 | -0.25019 | 0.169182 | 0.068358 | -0.48259 | -0.35801 | -0.82767 | -0.00551 | -0.53294 | -0.38821 | -1.50899 | -0.15755 | -0.1915 | -0.49707 | 1.011735 | -0.16058 | -0.3476 | -0.73863 | -0.94939 | -0.44413 | [illegible] |
| 1164 | -0.12787 | 0.076315 | 0.032674 | 0.347968 | -0.18247 | -0.38678 | -0.48731 | -0.28308 | -0.58061 | -0.09487 | -1.25588 | -0.28041 | 0.759718 | -0.82294 | 1.50886 | -0.16369 | -0.69153 | -0.84917 | -1.03852 | -0.42165 | [illegible] |

| | | | | | | | | | | | | | | | | | | | | |
|---|---|---|---|---|---|---|---|---|---|---|---|---|---|---|---|---|---|---|---|---|
| 1165 | 0.259625 | 0.1631 | -0.23211 | 3.13E-05 | 0.118709 | -0.3899 | -1.03733 | 0.019502 | -0.56 | 0.421525 | -1.13715 | -0.43379 | 0.692055 | -0.43463 | 1.052546 | -0.34442 | -0.94896 | -1.04323 | -1.01899 | -0.55979 |
| 1166 | 0.441815 | 0.13567 | -0.11073 | -0.26466 | -0.02146 | -0.28053 | -1.08565 | -0.03117 | -0.38205 | 0.714111 | -1.14995 | -0.71306 | 1.099208 | -0.25192 | -0.12726 | 0.364042 | -1.19856 | -0.9786 | -1.21939 | -0.77922 |
| 1167 | 0.523092 | -0.1071 | -0.2758 | -0.28102 | 0.030523 | -0.26332 | -0.53423 | 0.39927 | -0.32734 | 0.716152 | -1.11415 | -0.00125 | 0.840284 | -0.3355 | -0.09184 | 0.861593 | -1.2029 | -1.13749 | -1.2172 | -0.69718 |
| 1168 | 0.603343 | -0.02668 | -0.33401 | -0.83116 | 0.274133 | -0.67139 | -0.39584 | 0.605182 | -0.64522 | 0.895757 | -0.75416 | 1.328413 | 0.463152 | 0.03092 | -0.20492 | 1.299769 | -0.76536 | -0.75551 | -1.04655 | -0.51415 |
| 1169 | 0.16699 | -0.15664 | -0.18759 | -0.65772 | 0.2955 | -0.34624 | -0.28617 | 0.883189 | -0.29541 | 0.540629 | -0.68555 | 2.103049 | -0.09651 | 0.272682 | -0.23713 | 1.41404 | -0.66995 | -0.38097 | -0.06197 | -0.69362 |
| 1170 | -0.00333 | -0.20653 | -0.44477 | -0.44736 | 0.222424 | -0.17209 | 0.09866 | 0.691225 | -0.45955 | -0.12601 | -0.98432 | 2.609606 | 0.041297 | -0.09274 | -0.19793 | 1.321717 | -0.45966 | -0.1032 | 0.618545 | -0.95091 |
| 1171 | -0.01231 | -0.28159 | -0.31709 | -0.24913 | -0.1164 | -0.03449 | 0.190249 | 0.532589 | -0.55538 | -0.35569 | -0.73498 | 2.444281 | -0.30487 | 0.011721 | -0.1122 | 0.430447 | -0.1664 | 0.249517 | 0.730637 | -0.93229 |
| 1172 | -0.36998 | -0.3202 | -0.62394 | -0.33079 | -0.66552 | 0.103304 | 0.072962 | -0.01875 | -0.37489 | -0.58515 | -1.00045 | 1.981035 | -0.09349 | 0.110319 | -0.36262 | -0.21434 | -0.21682 | 0.044956 | 0.825612 | -0.81421 |
| 1173 | -0.48927 | -0.36174 | -0.70319 | 0.120503 | -0.8022 | 0.612091 | -0.00291 | -0.49721 | -0.53466 | -0.60632 | -0.86465 | 0.454823 | -0.74377 | 0.007517 | -0.34324 | -0.85556 | -0.43869 | 0.0934 | 1.012607 | -0.73255 |
| 1174 | -0.23939 | -0.38741 | -0.89371 | -0.14702 | -0.9716 | 0.090864 | -0.28833 | -0.30012 | -1.04196 | -0.1411 | -0.83911 | -0.22619 | -1.05746 | -0.52689 | -0.81637 | -0.59464 | 0.665006 | -0.39392 | -0.02267 | -0.52139 |
| 1175 | -0.45202 | -0.52055 | -0.77778 | -0.53274 | -0.65244 | 0.182224 | 0.110309 | 0.663475 | -0.61536 | 0.115455 | -0.58414 | -0.7623 | -1.20784 | -0.75127 | -0.81116 | -0.41297 | 2.899445 | -0.75539 | -0.69566 | -0.34946 |
| 1176 | -0.6106 | -0.21888 | -0.84797 | -0.61856 | -0.24167 | -0.22921 | -0.17614 | 0.92354 | -0.41267 | -0.113 | -0.62815 | -0.12155 | -1.29028 | -0.79978 | -1.14896 | 0.066686 | 4.611257 | -1.10746 | -0.70829 | -0.24381 |
| 1177 | -0.66481 | -0.19472 | -0.47076 | -0.51818 | -0.11461 | -0.32817 | -0.37616 | 1.02519 | -0.21622 | 0.045417 | -0.27159 | -0.34163 | -1.38237 | -1.01041 | -0.96413 | 0.174985 | 6.94084 | -0.73271 | -0.55326 | -0.22477 |
| 1178 | -0.56566 | 0.103413 | -0.30963 | -1.19204 | -0.38791 | -0.71733 | -0.60673 | 0.836839 | -0.00027 | -0.20691 | -0.24499 | 0.056121 | -0.81357 | -1.33799 | -0.96339 | -0.05282 | 8.587987 | -1.01803 | -0.59062 | 0.130963 |
| 1179 | -0.8185 | 0.066288 | -0.21239 | -0.92492 | -0.33262 | -0.25931 | -0.20741 | 0.873394 | -0.1137 | -0.6581 | -0.35414 | 0.231696 | -0.67642 | -1.36011 | -0.98075 | -0.41171 | 7.891527 | -0.84543 | -0.07513 | 0.290161 |
| 1180 | -0.5619 | 0.116007 | 0.111016 | -0.58261 | -0.65524 | -0.87957 | -0.44756 | -0.05941 | 0.042548 | -0.89876 | -0.3489 | 0.400526 | -0.35963 | -1.08028 | -0.39537 | -0.19364 | 5.832047 | -0.45237 | 0.139384 | -0.05823 |
| 1181 | -0.681 | 0.458405 | 0.232709 | -0.57092 | -1.09907 | -1.2139 | -0.08512 | -0.33349 | -0.27125 | -0.65425 | -0.50063 | -0.11724 | -0.41274 | -1.11257 | -0.03154 | -0.5552 | 4.007271 | -0.42565 | 0.330725 | -0.02935 |
| 1182 | -0.47855 | 0.28839 | 0.032364 | -0.58733 | -1.24801 | -0.85295 | 0.000323 | -0.22199 | -0.53733 | -0.70855 | -0.76467 | -0.06493 | -0.34217 | -0.62622 | -0.12627 | -0.66023 | 1.850073 | -0.44681 | -0.00921 | 0.015739 |
| 1183 | -0.19711 | 0.031585 | -0.32964 | -0.05176 | -1.24877 | -0.84624 | -0.12836 | 0.069748 | -0.55306 | -0.53769 | -0.93744 | 0.286242 | -0.09546 | -0.17059 | 0.154963 | -0.24101 | 0.184253 | -0.44291 | -0.06061 | -0.22618 |
| 1184 | -0.20155 | 0.235032 | -0.21717 | -0.43138 | -1.08698 | -0.98889 | 0.335641 | -0.05169 | -0.33615 | -0.41305 | -0.88912 | 0.287751 | -0.41713 | 0.985162 | 0.402203 | -0.45264 | -0.29095 | -0.53309 | -0.53105 | -0.73665 |
| 1185 | -0.26631 | 0.388066 | -0.55144 | -0.50438 | -0.67236 | -0.78984 | -0.22946 | 0.233167 | -0.92278 | -0.31427 | -0.87234 | 0.17063 | -0.64491 | 0.989374 | 0.111449 | -0.95197 | -0.52056 | -0.62079 | -0.44429 | -0.4012 |
| 1186 | -0.45432 | 0.089803 | -0.7341 | -0.50304 | 0.534966 | -0.4456 | -0.386 | 0.35809 | -0.86271 | -0.29084 | -0.77916 | 0.070809 | -0.57965 | 0.921115 | -0.33141 | -1.00805 | -0.39114 | -0.56434 | -0.53357 | -0.10176 |
| 1187 | -0.58081 | 0.274261 | -0.8055 | -0.96471 | 1.149335 | -1.18217 | -0.36434 | 0.696664 | -0.93448 | -0.23063 | -0.62868 | -0.2786 | -0.55105 | 0.452966 | -0.29247 | -1.17626 | -1.02776 | -1.22615 | -0.34867 | -0.13112 |
| 1188 | -1.10282 | 0.400475 | -0.69354 | -0.80085 | 1.423392 | -1.1737 | -0.28173 | 0.621213 | -1.00828 | -0.42162 | -0.68847 | -0.70723 | -1.02633 | 0.387931 | -0.57016 | -1.10279 | -0.8288 | -1.33975 | -0.3719 | -0.3038 |
| 1189 | -0.74466 | 0.232916 | -0.60171 | -0.88088 | 1.290426 | -1.4253 | -1.00562 | 0.444168 | -0.95647 | -0.32935 | -0.67325 | -0.77916 | -0.56158 | -0.4167 | -0.50733 | -0.43416 | -0.90761 | -1.46136 | -0.18449 | -0.05673 |
| 1190 | -0.68712 | -0.04085 | 0.428737 | -1.16547 | 0.741547 | -1.39457 | -0.92275 | 0.072662 | -0.69834 | -0.39728 | -0.66754 | -0.46107 | -0.66916 | -0.44974 | -0.51222 | -0.00496 | -0.85758 | -1.65963 | -0.40249 | -0.06503 |
| 1191 | -0.39091 | -0.26823 | 0.733833 | -1.19276 | -0.34576 | -1.42337 | -1.02006 | -0.11787 | -0.51477 | -0.40783 | -0.0754 | -0.27299 | -1.12659 | -0.39703 | -0.48663 | 0.301608 | -0.72452 | -1.5912 | -0.46891 | -0.5086 |
| 1192 | -0.21686 | -0.38217 | 0.595288 | -0.86078 | -0.81503 | -1.2148 | -1.75289 | -0.46241 | -0.19729 | -0.50704 | 0.08814 | 0.231693 | -1.0779 | 0.234049 | -0.11015 | 0.680443 | -0.09724 | -0.9942 | -0.69914 | -0.64824 |
| 1193 | -0.1779 | -0.49482 | 0.634135 | -1.26909 | -0.93763 | -0.93812 | -1.59615 | -0.40091 | -0.23753 | -0.10378 | -0.08025 | 0.158898 | -0.73385 | 0.622196 | 1.132483 | 0.536602 | 0.200408 | -0.83925 | -0.69505 | -0.52129 |
| 1194 | -0.44769 | -0.23114 | 0.628474 | -1.02552 | -0.90817 | -0.45244 | -1.66608 | -0.39778 | -0.18767 | -0.03995 | -0.17419 | 0.190379 | -0.69806 | 1.241843 | 2.492708 | -0.02604 | 0.283465 | -0.76612 | -0.78678 | -0.43089 |
| 1195 | -0.53291 | -0.37446 | -0.38118 | -0.66407 | -0.54168 | -0.20565 | -1.4195 | -0.52752 | 0.337567 | 0.151556 | -0.53867 | -0.00744 | -0.67766 | 1.333556 | 2.563065 | -0.25363 | 0.56647 | -0.43409 | -0.76041 | -0.06617 |
| 1196 | -0.52077 | -0.60881 | -1.03261 | -0.44472 | -0.33154 | 0.040527 | -1.44421 | -0.4171 | 1.352822 | -0.03738 | -0.8252 | 0.045448 | -0.24301 | 1.288805 | 2.474317 | -0.61376 | 0.475744 | 0.153298 | -0.01299 | -0.28112 |
| 1197 | -0.43122 | -0.27413 | -0.78421 | -0.11552 | 0.576769 | 0.123472 | -0.92481 | -0.59404 | 1.420843 | 0.430172 | -1.03674 | -0.55446 | -0.25472 | 1.106404 | 2.345598 | -0.9183 | 0.744689 | 0.297821 | -0.15586 | -0.0334 |
| 1198 | -0.06798 | 0.14928 | -0.36611 | 0.036343 | 1.250675 | -0.27657 | -1.13815 | -0.50881 | 1.569972 | -0.0263 | -0.79847 | -0.61021 | -0.3265 | 0.529211 | 1.260786 | -1.14939 | 0.294241 | 0.316878 | -0.15071 | -0.1484 |
| 1199 | 0.325544 | 0.684506 | -0.29207 | 0.098835 | 1.426369 | -0.44488 | -1.01893 | -0.52496 | 1.516048 | -0.3612 | -0.84289 | -0.59063 | -0.71212 | -0.23987 | 0.015379 | -1.24037 | 0.528991 | 0.63634 | -0.07024 | -0.4935 |
| 1200 | 0.650218 | 0.837863 | -0.46929 | 0.314354 | 1.556897 | -0.5324 | -0.79945 | -0.42514 | 1.202974 | -0.17138 | -0.43996 | -0.88435 | -0.40561 | -0.28606 | -0.01482 | -0.83545 | 0.251639 | 0.678948 | 0.10392 | -0.73847 |
| 1201 | 2.082877 | 1.090082 | -0.34892 | 0.214093 | 1.117756 | -0.65282 | -0.15571 | -0.3593 | 0.154042 | 0.100288 | -0.93995 | -1.0328 | -0.46268 | -0.17205 | 0.019513 | -0.34433 | -0.008 | -0.15339 | -0.69242 | -0.27764 |
| 1202 | 2.637851 | 0.979633 | 0.043295 | 0.356341 | 0.230101 | -0.8097 | 0.08913 | 0.077232 | -0.25856 | -0.25122 | -0.73298 | -0.72085 | 0.00515 | -0.34144 | -0.15444 | -0.18749 | -0.43419 | -0.78205 | -0.59014 | -0.2962 |
| 1203 | 2.820138 | 0.521686 | -0.09942 | 0.433236 | -0.57379 | -0.35395 | 0.268707 | 0.199703 | -0.60123 | -0.2077 | -0.62015 | -1.15421 | 0.271464 | -0.26986 | -0.21099 | -0.31997 | -0.27762 | -0.52153 | -0.45866 | -0.18742 |
| 1204 | 2.588507 | -0.29238 | -0.13123 | 0.192187 | -0.77528 | -0.42023 | 0.537062 | 0.232427 | -0.71477 | 0.10316 | -0.04387 | -1.30746 | 0.241929 | -0.2997 | -0.19898 | -0.14322 | 0.393042 | -0.70873 | -0.31933 | -0.26109 |
| 1205 | 2.102592 | -0.34794 | 0.373351 | 0.077203 | -0.99942 | -0.6088 | 0.288251 | 0.275795 | -0.86897 | -0.28858 | 1.1975 | -1.25445 | -0.08033 | -0.38343 | -0.5148 | -0.55919 | -0.09646 | -0.61163 | -0.67101 | -0.15884 |
| 1206 | 0.700968 | -0.81062 | 0.25948 | 0.090341 | -0.55118 | -0.64875 | -0.50866 | 0.284792 | -1.01512 | -0.37775 | 1.401345 | -0.93737 | -0.14356 | -0.64728 | -0.58025 | -0.62531 | -0.02001 | -0.42036 | -0.39215 | -0.19226 |
| 1207 | 0.166746 | -1.21145 | -0.31802 | -0.5005 | -0.23202 | -0.34139 | -0.29206 | -0.23921 | -0.03339 | -0.9327 | 1.284589 | -0.88712 | -0.86293 | -0.27981 | -0.70908 | -0.6702 | -0.09324 | 0.428131 | -0.20787 | -0.48412 |
| 1208 | -0.32479 | -1.12525 | -0.39486 | -0.85964 | 0.613613 | -0.77625 | -0.20773 | -0.35195 | 0.604905 | -1.01314 | 1.178093 | -0.70237 | -1.267 | 0.487377 | -0.68317 | -0.36204 | 1.670971 | 0.412617 | -0.59746 | -0.88707 |
| 1209 | -0.51924 | -0.90793 | -0.51553 | -0.42121 | 0.841076 | -0.71467 | -0.2237 | -0.70452 | 1.09267 | -0.95961 | 0.785226 | -0.66963 | -0.92446 | 1.097495 | -0.73171 | -0.26672 | 2.175152 | 0.612951 | -0.73654 | -0.51315 |
| 1210 | -0.43968 | -0.86611 | -0.89051 | -0.72344 | 0.856467 | -0.53029 | -0.22419 | -0.35699 | 0.912538 | -0.70946 | -0.42406 | -0.6082 | -0.80007 | 1.058725 | -0.29066 | -0.4341 | 2.615368 | 0.398081 | -0.56822 | -0.61412 |
| 1211 | -0.57022 | -0.48858 | -0.12361 | -0.72307 | 0.682665 | -0.48403 | 0.154967 | -0.40496 | 1.074578 | -0.8083 | -0.27195 | -0.69737 | -0.53832 | 1.474024 | 0.027541 | -0.5764 | 2.501664 | 0.18676 | -0.88152 | -0.5627 |
| 1212 | -0.42148 | -0.45262 | 0.561951 | -0.44487 | 0.469735 | -0.58676 | -0.09129 | -0.3522 | 0.28907 | -0.2607 | -0.30095 | -0.69559 | -0.29742 | 1.146179 | 0.004302 | -0.52115 | 2.844584 | -0.32406 | -0.88299 | -0.34372 |
| 1213 | -0.14645 | -0.5195 | 1.119495 | -0.14449 | -0.15691 | -0.52147 | 0.384124 | -0.5665 | -0.02231 | 0.054062 | -0.06687 | -0.22852 | -0.3044 | 0.293229 | -0.01653 | -0.70443 | 0.869943 | -0.61524 | -0.75903 | -0.27584 |
| 1214 | 0.010804 | -0.54359 | 1.096011 | -0.31296 | -0.29663 | -0.49113 | 0.241509 | -0.38089 | -0.2831 | -0.13329 | -0.2936 | 0.136414 | -0.24599 | -0.13761 | -0.08799 | -0.75751 | -0.34384 | -0.71673 | -0.79271 | -0.34459 |
| 1215 | 0.071709 | -0.28848 | 1.267646 | -0.14878 | -0.45075 | -0.34935 | 0.358077 | -0.92067 | -0.2716 | -0.44798 | -0.65799 | 0.729981 | -0.23951 | -0.1701 | -0.21983 | -0.20826 | -0.27364 | -0.6848 | -0.7131 | -0.37358 |
| 1216 | 0.011509 | 0.11448 | 0.840588 | -0.00121 | -0.26952 | -0.23503 | 0.546023 | -0.90414 | -0.44896 | -0.37164 | -0.56162 | 0.722313 | -0.38289 | -0.39684 | -0.33549 | -0.2374 | 0.123771 | -0.521 | -0.49616 | -0.57178 |
| 1217 | -0.04953 | 0.395532 | 0.313546 | -0.33809 | -0.18532 | -0.3387 | 0.680902 | -0.9162 | -0.54401 | -0.37504 | -0.66605 | 0.634112 | -0.02377 | -0.42071 | -0.28737 | -0.29239 | -0.13001 | -0.49888 | -0.42336 | -0.73536 |
| 1218 | 0.017197 | 0.468352 | -0.27023 | -0.24744 | -0.15173 | -0.14253 | 0.041355 | -1.36 | -0.59116 | -0.32721 | -0.67578 | 0.5122 | -0.06719 | -0.73613 | -0.39722 | 0.06682 | -0.05487 | -0.32048 | -0.36843 | -0.43332 |
| 1219 | 0.055342 | 0.240815 | -0.16264 | -0.11371 | -0.15173 | -0.6814 | -0.04102 | -1.18947 | -0.79213 | -0.307 | -0.62132 | 0.187589 | -0.32711 | -0.68159 | -0.13676 | 0.012846 | -0.06838 | -0.25159 | -0.48831 | -0.32394 |
| 1220 | 0.055342 | 0.232184 | -0.01415 | -0.60965 | -0.17881 | -0.781 | 0.009783 | -1.09856 | -0.87693 | 0.00608 | -0.26959 | -0.33177 | -0.51199 | -0.22622 | -0.23479 | -0.13038 | 0.034674 | -0.28954 | -0.42145 | -0.35656 |
| 1221 | -0.07206 | -0.08097 | -0.2335 | -0.9949 | -0.14833 | -1.02299 | -0.19055 | -1.12119 | -0.59594 | 0.00608 | -0.37378 | -0.63791 | -0.40047 | -0.35068 | -0.34679 | -0.09367 | 0.240857 | -0.27571 | -0.51207 | -0.23021 |
| 1222 | -0.33634 | -0.22027 | -0.11342 | -0.60849 | -0.30774 | -1.25718 | -0.59745 | -1.10389 | -0.63186 | 0 | -0.18626 | -0.78051 | -0.76777 | -0.35471 | -0.28042 | 0.117604 | 0.307479 | -0.07703 | -0.42426 | -0.08326 |
| 1223 | -0.42014 | -0.22027 | -0.11342 | -0.60849 | -0.30774 | -1.53219 | -0.57465 | -0.51883 | -0.63186 | 0 | -0.18626 | -0.78051 | -0.51099 | 0.16394 | 0.028908 | -0.21565 | 0.35146 | -0.07703 | -0.06506 | -0.14411 |
| 1224 | -0.45829 | -0.14876 | -0.11342 | -0.36604 | -0.30774 | -1.00836 | -0.75811 | -0.54211 | -0.49039 | -0.17371 | 0 | -0.88592 | -0.25885 | 0.109399 | -0.23155 | -0.37563 | 0.766027 | -0.39331 | 0.185291 | -0.24969 |
| 1225 | -0.68793 | -0.25163 | -0.16751 | -0.06786 | -0.27869 | -0.90447 | -0.75811 | -0.27711 | -0.19075 | -0.15844 | -0.04183 | -0.77768 | -0.26323 | -0.23265 | -0.36939 | -0.41628 | 0.636413 | -0.31922 | 0.025162 | 0.144228 |
| 1226 | -0.36814 | -0.26886 | 0.068113 | 0.261478 | -0.51051 | -0.318 | -0.41306 | -0.46253 | -0.408 | -0.0199 | -0.04183 | -0.49757 | -0.27662 | -0.22137 | -0.14042 | -0.37059 | 0.444632 | -0.2474 | -0.1393 | -0.09382 |
| 1227 | -0.09827 | -0.03007 | 0.068113 | -0.12642 | -0.49314 | -0.41394 | -0.2108 | -0.31235 | -0.48532 | -0.30104 | 0.016203 | -0.4144 | -0.89425 | -0.37593 | -0.14076 | -0.52592 | 0.247804 | -0.74359 | -0.40746 | -0.13131 |
| 1228 | -0.08038 | -0.03652 | -0.02226 | -0.12642 | -0.60884 | 0.126821 | -0.23214 | -0.23439 | -0.74749 | -0.42379 | 0.052234 | -0.58122 | -0.76784 | -0.63963 | -0.20264 | -0.14546 | 0.247804 | -0.59399 | -0.69666 | -0.25471 |
| 1229 | -0.40889 | 0.017521 | -0.07644 | -0.49093 | -0.78376 | 0.772001 | -0.1956 | -0.38111 | -1.0735 | -0.57076 | -0.28702 | -0.69136 | -1.0372 | -0.86702 | -0.52592 | -0.16778 | 0.059129 | -0.47593 | -0.69666 | -0.03442 |
| 1230 | -0.17925 | -0.25625 | -0.21052 | -0.28195 | -0.61299 | 0.56492 | -0.36452 | -0.68794 | -1.38715 | -0.85295 | -0.38488 | -0.78076 | -0.86553 | -0.98984 | -0.58761 | -0.42572 | -0.24786 | -0.62241 | -0.63195 | -0.61307 |
| 1231 | -0.50216 | -0.09451 | -0.03817 | -0.33569 | -0.59019 | 0.133731 | -1.00121 | -0.43985 | -1.06673 | -1.00523 | -0.16535 | -0.64948 | -0.81555 | -1.0196 | -0.58761 | -0.42572 | -0.39107 | -0.6277 | -0.6741 | -0.18548 |
| 1232 | -0.75703 | 0.157466 | 0.153103 | 0.017574 | -0.81676 | 0.56249 | -1.04783 | -0.41766 | -0.53661 | -1.14417 | -0.25329 | -0.32971 | 0.094789 | -0.56115 | -0.70727 | -0.63782 | -0.07801 | -0.17574 | -0.71262 | 0.158068 |
| 1233 | -1.23761 | 0.806058 | 0.156944 | 0.414026 | -0.37125 | 0.845342 | -0.71041 | -0.13672 | 0.255272 | -1.277 | -0.22781 | -0.37685 | 0.065541 | -0.05904 | -0.21521 | -1.25012 | -0.10846 | -0.264 | -0.75315 | 0.499949 |
| 1234 | -1.23735 | 1.010639 | 0.376208 | 2.18998 | -0.03174 | 0.564812 | -0.42105 | 0.168431 | 0.814453 | -0.87594 | 0.248702 | 0.399695 | 0.60321 | 0.922475 | 0.310788 | -0.99085 | -0.31842 | 0.315069 | -0.8417 | 0.388447 |
| 1235 | -1.52158 | 1.516309 | 0.826268 | 3.463062 | -0.36377 | 0.314858 | 0.084835 | 0.581077 | 1.475053 | -0.96892 | 0.558091 | 0.965222 | 1.193873 | 1.086426 | 0.731394 | -0.28973 | -0.10151 | 0.329045 | -0.72204 | 1.246331 |
| 1236 | -1.27966 | 1.737004 | 0.63764 | 3.505288 | -0.27461 | 0.604727 | 0.473957 | 0.741385 | 1.426375 | -0.93951 | 0.274536 | 1.06846 | 2.044188 | 1.346685 | 0.843735 | -0.23446 | -0.01027 | 0.810301 | -0.43825 | 1.119041 |
| 1237 | -1.30881 | 1.529894 | 0.386828 | 3.949845 | 0.576944 | 1.562245 | 0.459063 | 0.568421 | 1.133933 | -0.0767 | 0.242452 | 0.7476 | 1.816917 | 1.779281 | 1.066435 | 0.215604 | 0.093268 | 0.917569 | -0.0792 | 0.959256 |
| 1238 | -0.88921 | 0.845605 | 0.572137 | 4.071752 | -0.01872 | 2.307326 | 0.403604 | 0.279032 | 0.268889 | 0.078569 | 0.116936 | 1.022836 | 1.360047 | 1.767844 | 0.45006 | 1.002461 | 0.239147 | 0.76897 | 0.223629 | 0.993652 |
| 1239 | -0.52393 | 0.496902 | 0.394577 | 2.574448 | 0.241781 | 1.935643 | 0.687662 | 0.337722 | 0.119549 | 0.307144 | 0.158165 | 0.575629 | 1.030814 | 1.275391 | 0.507282 | 1.085864 | 0.18707 | 0.061592 | 0.609082 | 0.852398 |
| 1240 | -0.2862 | 0.857667 | 0.262496 | 1.248483 | 0.424323 | 2.083783 | 0.640468 | 0.245409 | -0.18762 | 1.053485 | 0.288831 | 0.015622 | 0.429386 | 1.5866 | 0.486771 | 1.009739 | 0.216042 | 0.447982 | 0.506258 | 0.385626 |
| 1241 | -0.60134 | 0.577753 | 0.112486 | 0.938104 | 0.503902 | 2.022384 | 0.602811 | -0.1827 | -0.30127 | 1.166804 | 0.107086 | -0.31322 | -0.39856 | 1.594216 | 0.321323 | 0.863824 | 0.094297 | 0.022972 | 0.667913 | 0.260713 |
| 1242 | -0.35862 | 0.4035 | 0.172024 | 0.924924 | -0.28203 | 0.711802 | 0.582756 | -0.26902 | -0.17405 | 0.724071 | 0.169077 | -0.45186 | -0.61654 | 0.967214 | 0.212365 | 0.380053 | 0.180083 | -0.29777 | 0.548438 | 0.098434 |
| 1243 | -0.6685 | 0.789393 | 0.122238 | 0.265982 | -0.07479 | -0.66328 | 0.315135 | -0.08117 | 0.60991 | 0.895469 | 0.540678 | -0.34015 | -0.52046 | 1.442043 | 0.060312 | 0.158172 | -0.09968 | -0.40361 | 0.479939 | 0.666879 |
| 1244 | -0.60831 | 1.910566 | 0.134709 | 0.04088 | -0.13803 | -0.71947 | 0.153734 | 0.038531 | 0.544226 | 0.586512 | 0.598226 | -0.48952 | -0.0752 | 1.43456 | -0.31253 | -0.07761 | -0.05003 | -0.35531 | -0.17162 | 1.142101 |
| 1245 | -0.644 | 1.799534 | 0.179811 | 0.541359 | 0.108401 | -0.23013 | 0.072973 | 0.198635 | 0.609577 | 0.311291 | 0.627648 | 0.055618 | 0.166243 | 1.127824 | -0.23125 | -0.46961 | -0.00824 | -0.86461 | -0.0322 | 1.131554 |
| 1246 | -0.39054 | 2.320528 | 0.990717 | 0.710835 | 0.365521 | -0.35586 | -0.00207 | 0.581279 | 0.821666 | 0.164334 | 0.873424 | 0.321931 | 0.602753 | 1.124883 | 0.052657 | -0.29301 | 0.135685 | -0.51257 | 0.047487 | 1.529286 |
| 1247 | -0.7855 | 2.114491 | 1.226983 | 0.111622 | 1.215048 | -0.22029 | -0.09329 | 1.067666 | 0.421403 | 0.408017 | 0.982777 | 0.77418 | 1.013186 | 0.850609 | 0.354377 | 0.297325 | -0.13518 | -0.22147 | 0.011756 | 1.446364 |
| 1248 | -0.70404 | 1.550647 | 0.955436 | -0.05164 | 1.233096 | -0.1389 | 0.220974 | 0.619047 | -0.08563 | 0.888989 | 1.002712 | 0.601191 | 1.223056 | -0.1724 | 0.512373 | 0.031058 | 0.029142 | -0.02837 | -0.17065 | 0.517422 |
| 1249 | -0.73426 | 0.720575 | 0.850642 | -0.22621 | 1.115132 | -0.47429 | 0.439672 | 0.62286 | -0.14264 | 1.17067 | 0.996495 | 0.84246 | 1.040323 | -0.39318 | 0.559594 | 0.683674 | -0.11769 | 0.286893 | 0.056405 | 0.097543 |
| 1250 | -0.51221 | 0.421633 | 0.594943 | -0.32627 | 1.408806 | -0.90357 | 0.500439 | 0.599332 | 0.207709 | 1.806208 | 0.946481 | 0.208399 | 1.308918 | -0.50022 | 0.006129 | 1.446234 | 0.167981 | 0.730918 | -0.12563 | 0.633587 |
| 1251 | -0.72065 | 0.237994 | -0.30273 | -0.33638 | 1.121897 | -0.82369 | 0.736996 | 0.259232 | 0.413752 | 1.796912 | 0.735682 | 0.105933 | 0.739616 | -0.24674 | -0.04149 | 1.595291 | 0.192682 | 0.327922 | -0.13404 | 0.521092 |
| 1252 | -0.7341 | 0.733802 | -0.43142 | -0.33194 | 0.9003 | -0.621 | 1.198162 | 0.034813 | 0.732085 | 1.400776 | 0.296207 | 0.093758 | 0.27883 | 0.173431 | -0.55065 | 1.318012 | 0.036435 | 0.542716 | 0.19542 | 0.995064 |
| 1253 | -0.72035 | 1.256904 | -0.42602 | -0.08086 | 1.334052 | -0.66374 | 1.35776 | 0.447264 | 0.935794 | 0.906327 | -0.17603 | 0.176464 | 0.631075 | 0.746472 | -0.21979 | 1.679875 | -0.04425 | 0.664682 | 0.301073 | 1.390382 |
| 1254 | -1.00927 | 0.940927 | -0.32122 | 0.249129 | 0.951291 | -0.34194 | 1.0416 | 0.039192 | 0.817959 | 0.308346 | -0.7385 | -0.00644 | 0.396372 | 0.599647 | -0.1542 | 1.124382 | 0.102405 | 0.886682 | 0.358671 | 1.473089 |
| 1255 | -1.32362 | 0.492219 | -0.46136 | -0.19675 | 0.554593 | 0.319804 | 0.63951 | -0.31752 | 0.384906 | -0.72411 | -1.0626 | 0.174473 | 0.273289 | 0.43271 | -0.04075 | 0.303245 | -0.245 | 0.724609 | 0.59825 | 0.78247 |
| 1256 | -1.23782 | -0.33698 | -0.2276 | -0.02084 | 0.337635 | 0.544406 | 0.322028 | -0.09205 | 0.069555 | -1.25178 | -0.96632 | 0.302841 | 0.259914 | -0.04947 | -0.27705 | 0.146019 | -0.47532 | 0.712394 | 0.459742 | 0.856088 |
| 1257 | -0.87801 | -0.50697 | -0.33517 | 0.092147 | -0.22658 | 0.687409 | 0.061851 | 0.051356 | -0.04859 | -1.02694 | -0.76424 | 0.292815 | 0.292178 | -0.56808 | -0.2164 | 0.007635 | -0.05783 | 0.280908 | 0.037436 | 0.481041 |
| 1258 | -0.72778 | -0.80997 | -0.38643 | -0.07786 | -0.61977 | 0.877235 | -0.39594 | -0.27891 | -0.14003 | -1.22908 | -0.62432 | 0.153107 | -0.13641 | -0.81992 | -0.6838 | -0.21409 | 0.000394 | 0.158942 | -0.29878 | 0.268945 |
| 1259 | -0.74128 | -0.72477 | -0.56263 | -0.29849 | -0.4809 | 0.938043 | 0.085216 | -0.39485 | -0.15207 | -0.91278 | -0.2234 | 0.172004 | -0.2826 | -0.81225 | -0.8615 | -0.41085 | -0.22569 | -0.42492 | -0.32395 | 0.21839 |
| 1260 | -0.68292 | -0.39648 | -0.5046 | -0.25057 | -0.30107 | 0.619733 | 0.250718 | -0.30997 | -0.37127 | -0.34461 | 0.109544 | 0.046853 | -0.73953 | -0.65502 | -0.62508 | -0.35283 | -0.10987 | -0.60731 | -0.15805 | 0.353921 |
| 1261 | -0.46308 | 0.264293 | -0.38767 | -0.44042 | 0.083253 | 0.35998 | 0.247594 | -0.23362 | -0.62269 | 0.367209 | 0.521371 | 0.250243 | -0.55116 | -0.31656 | -0.66069 | -0.60625 | -0.29001 | -0.69349 | -0.44106 | -0.33621 |
| 1262 | -0.43722 | 0.36065 | -0.57298 | -0.44848 | 0.082582 | 0.097659 | 0.287791 | -0.42628 | -0.35838 | 0.567449 | 0.669133 | 0.031119 | -0.29742 | 0.101609 | -0.26715 | -0.59024 | 0.076242 | -0.48751 | -0.65986 | -0.31423 |
| 1263 | -0.54845 | 0.425301 | -0.14218 | -0.48581 | -0.24559 | -0.3769 | 0.616833 | -0.53144 | -0.60796 | 0.653595 | 0.523028 | 0.466396 | -0.03852 | 0.426938 | 0.036658 | -0.77683 | 0.451091 | -0.36811 | -0.62389 | -0.28401 |
| 1264 | -0.41118 | 0.918143 | 0.633918 | -0.1474 | -0.22677 | -0.30571 | -0.04293 | -0.65148 | -0.73484 | 0.272873 | 1.070486 | 0.904285 | 0.051656 | 0.528119 | 0.000556 | -0.36311 | 0.67506 | -0.17003 | -0.97382 | -0.0886 |
| 1265 | -0.41951 | 1.325545 | 1.12378 | -0.09201 | -0.51515 | -0.56497 | -0.17713 | -0.33429 | -0.20438 | -0.3263 | 1.284687 | 1.307617 | -0.07709 | 0.565151 | 0.090691 | -0.23641 | 0.588235 | 0.074556 | -1.30893 | -0.2406 |
| 1266 | -0.55652 | 1.396639 | 1.190157 | -0.30354 | -1.08558 | -0.71537 | -0.21132 | -0.397 | -0.04591 | 0.021677 | 1.700716 | 0.862277 | -0.26427 | 0.484733 | 0.016928 | -0.09036 | 0.851703 | 0.299931 | -1.31049 | 0.174013 |
| 1267 | -0.48137 | 1.380703 | 1.27859 | -0.26817 | -0.72432 | -0.50962 | -0.21675 | -0.48361 | -0.44538 | -0.21581 | 1.573113 | 0.756129 | -0.20654 | -0.04801 | -0.29239 | 0.065237 | 0.0587 | 0.659156 | -1.35619 | 0.390231 |
| 1268 | -0.40417 | 1.43403 | 1.080195 | -0.05476 | -0.05839 | -0.35821 | -0.39939 | -0.33772 | -0.29157 | -0.08877 | 1.864053 | 0.57167 | -0.53465 | -0.23917 | -0.56617 | -0.01281 | -0.63019 | 0.331424 | -1.278 | 0.182336 |
| 1269 | 0 | 0 | 0 | 0 | 0 | 0 | 0 | 0 | 0 | 0 | 0 | 0 | 0 | 0 | 0 | 0 | 0 | 0 | 0 | 0 |
| 1270 | 0 | 0 | 0 | 0 | 0 | 0 | 0 | 0 | 0 | 0 | 0 | 0 | 0 | 0 | 0 | 0 | 0 | 0 | 0 | 0 |
| 1271 | 0 | 0 | 0 | 0 | 0 | 0 | 0 | 0 | 0 | 0 | 0 | 0 | 0 | 0 | 0 | 0 | 0 | 0 | 0 | 0 |
| 1272 | 0 | 0 | 0 | 0 | 0 | 0 | 0 | 0 | 0 | 0 | 0 | 0 | 0 | 0 | 0 | 0 | 0 | 0 | 0 | 0 |
| 1273 | 0 | 0 | 0 | 0 | 0 | 0 | 0 | 0 | 0 | 0 | 0 | 0 | 0 | 0 | 0 | 0 | 0 | 0 | 0 | 0 |
| 1274 | 0 | 0 | 0 | 0 | 0 | 0 | 0 | 0 | 0 | 0 | 0 | 0 | 0 | 0 | 0 | 0 | 0 | 0 | 0 | 0 |

| | | | | | | | | | | | | | | | | | | | | |
|---|---|---|---|---|---|---|---|---|---|---|---|---|---|---|---|---|---|---|---|---|
| 1165 | 0.780247 | 0.329038 | -0.15185 | 0.500197 | 0.112814 | -0.2187 | -0.69217 | 0.005893 | 0.549464 | 0.712568 | -0.22192 | 0.240222 | 1.618072 | 0.299833 | 0.587394 | -0.80577 | -0.22155 | 1.125517 | 0.159073 | 0.144793 |
| 1166 | 0.695376 | 0.28899 | -0.18506 | 0.695996 | 0.227689 | -0.33247 | -0.63624 | 0.396119 | 0.590384 | 0.900153 | -0.46608 | -0.16859 | 2.11484 | 0.528988 | -0.63914 | -0.62891 | -0.35926 | 1.008838 | -0.04347 | -0.65297 |
| 1167 | 2.001048 | 0.104469 | 2.296179 | 2.787408 | 3.43878 | -0.49167 | -0.24028 | 1.239329 | 0.764069 | 1.004394 | -0.46257 | 1.251991 | 4.703292 | 3.154005 | 0.927763 | 2.424959 | -0.39463 | 2.567943 | -0.00984 | -0.24615 |
| 1168 | 3.561908 | -0.37167 | 4.451809 | 4.742099 | 6.044185 | -0.58474 | 0.355644 | 1.758394 | 1.418585 | 1.87952 | 0.082749 | 2.965113 | 5.660845 | 5.367027 | 2.504539 | 5.171249 | 0.229515 | 4.734311 | 0.837434 | -0.10313 |
| 1169 | 3.48951 | -0.53425 | 4.645539 | 4.490954 | 6.410687 | -0.23454 | 0.127869 | 1.729261 | 1.639385 | 1.446654 | 0.270476 | 3.580012 | 4.775801 | 5.575084 | 2.788774 | 6.103264 | 0.148772 | 4.544398 | 1.46699 | -0.13419 |
| 1170 | 3.30068 | -0.47871 | 4.927284 | 4.512874 | 6.689711 | 0.228059 | 0.297955 | 1.540384 | 1.365952 | 1.197707 | -0.11922 | 4.027594 | 4.351119 | 5.657739 | 2.847662 | 6.581252 | 0.339264 | 3.483507 | 1.446223 | -0.07837 |
| 1171 | 2.773656 | -0.65018 | 4.810788 | 4.832054 | 6.449613 | 0.353087 | 0.560156 | 1.281019 | 0.988698 | 1.004643 | 0.198164 | 4.118551 | 3.921951 | 5.550893 | 3.424108 | 5.921335 | 0.051938 | 4.034585 | 1.669373 | 0.287474 |
| 1172 | 1.464888 | -0.49468 | 2.437306 | 2.376973 | 3.325235 | 0.587552 | -0.08336 | -0.02504 | 0.580415 | 0.583993 | 0.322684 | 2.813894 | 1.690904 | 2.689996 | 1.687274 | 3.122511 | 0.162353 | 2.710142 | 1.486734 | -0.21857 |
| 1173 | -0.24973 | -0.00114 | 0.055874 | 0.267794 | 0.714769 | 0.903038 | -0.43067 | -0.66985 | -0.17754 | -0.43028 | -0.20203 | 1.020026 | 0.257338 | 0.234476 | -0.01881 | 0.516433 | -0.06905 | 0.702265 | 1.224513 | -0.39855 |
| 1174 | -0.77059 | 0.25061 | -0.04927 | 0.129486 | 0.407196 | 0.985834 | 0.501076 | -0.64722 | -0.60539 | -0.65769 | -0.42565 | 0.289113 | -0.10285 | -0.32572 | -0.40785 | -0.23174 | 0.204208 | 0.07567 | 0.396031 | -0.37395 |
| 1175 | -1.14532 | 0.161996 | -0.49297 | -0.19427 | 0.056501 | 0.885227 | 0.84274 | -0.13214 | -0.4303 | -0.55182 | -0.17559 | -0.26364 | -0.15317 | -0.87433 | -0.50496 | -0.59751 | 2.202397 | -0.58967 | -0.1656 | -0.0682 |
| 1176 | -0.77189 | 0.084112 | -0.38005 | -0.6889 | 0.171885 | 0.780198 | 0.866381 | 0.211286 | -0.27537 | -0.47243 | -0.19935 | 0.155839 | -0.21042 | -0.8835 | -0.79196 | -0.2706 | 4.376011 | -0.92495 | -0.09377 | -0.10381 |
| 1177 | -0.71172 | -0.05836 | -0.50797 | -0.49579 | -0.07531 | 0.527321 | 1.138834 | 0.310887 | -0.26521 | -0.19803 | -0.16523 | -0.17545 | -0.28441 | -0.8828 | -0.64395 | -0.26826 | 6.700671 | -1.11071 | 0.156763 | 0.02596 |
| 1178 | -0.47482 | -0.28073 | -0.38118 | -1.01946 | -0.40359 | 0.005529 | 0.954543 | 0.521034 | -0.30196 | -0.14097 | 0.147858 | -0.23853 | -0.2696 | -0.82283 | -0.30645 | -0.33688 | 7.873656 | -0.9391 | 0.422825 | 0.193446 |
| 1179 | -0.18934 | -0.33197 | -0.63508 | -0.54579 | -0.57907 | 0.027837 | 0.264236 | 1.028888 | -0.02543 | -0.18047 | 0.418358 | -0.0272 | -0.05778 | -0.78523 | -0.0836 | -0.25379 | 7.57526 | -0.66363 | 0.760491 | 0.338445 |
| 1180 | -0.65868 | -0.58031 | -0.56738 | -0.29072 | -0.82943 | -0.16459 | -0.53374 | 0.571156 | -0.18644 | -0.22924 | 0.62953 | -0.07456 | 0.035914 | -0.78173 | 0.2886 | -0.53047 | 5.465898 | 0.247258 | 0.773363 | 0.222399 |
| 1181 | -0.5377 | 0.12293 | -0.26274 | 0.513868 | -0.82162 | -0.29362 | -0.27665 | 0.40519 | -0.16945 | -0.28195 | 0.766405 | -0.16364 | 0.072535 | -0.5906 | 0.643117 | -0.12389 | 3.456489 | 1.039684 | 0.627758 | 0.120469 |
| 1182 | -0.82404 | 0.079168 | 0.194251 | 0.645521 | -0.17927 | -0.57311 | -0.07651 | 0.580782 | -0.23683 | -0.55132 | 0.603515 | -0.02671 | 0.176724 | -0.02363 | 0.68129 | -0.21518 | 0.978716 | 1.143652 | 0.41518 | -0.11269 |
| 1183 | -1.13994 | 0.08993 | -0.22996 | 1.245425 | 0.472362 | -0.53209 | -0.3254 | 0.567823 | -0.3234 | -0.38264 | 0.08925 | 0.384188 | 0.215169 | 0.562221 | 0.782027 | 0.002273 | -0.24984 | 1.318024 | -0.09565 | -0.28604 |
| 1184 | -1.46042 | 0.027527 | -0.42448 | 1.560353 | 0.599301 | -0.23114 | 0.061571 | 0.079784 | -0.41552 | -0.26284 | -0.2693 | 0.470457 | 0.282572 | 1.764614 | 0.985265 | 0.81483 | -0.12117 | 1.316476 | -0.36244 | -0.13965 |
| 1185 | -0.98641 | 0.140425 | -0.50731 | 1.345563 | 1.464535 | -0.15976 | 0.412664 | 0.172825 | -0.59311 | -0.28512 | -0.7531 | 0.509567 | 0.248634 | 2.092688 | 0.44996 | 1.272599 | -0.20049 | 0.622375 | -0.21382 | -0.13221 |
| 1186 | -1.0139 | -0.43548 | -1.01102 | 0.743191 | 1.033731 | 0.074308 | 0.090504 | 0.284538 | -0.45846 | -0.23373 | -0.94236 | 0.044297 | 0.192719 | 2.012616 | 0.243419 | 0.701558 | -0.18051 | -0.66589 | -0.57321 | 0.115959 |
| 1187 | -1.07576 | -0.42339 | -1.13095 | 0.689678 | 0.660565 | 0.31854 | -0.16507 | 0.737451 | -0.35004 | -0.19517 | -0.79667 | 0.146306 | -0.03556 | 1.589344 | 0.086515 | 0.822152 | -0.01108 | -0.56138 | -0.42775 | 0.155247 |
| 1188 | -1.15118 | -0.21179 | -0.70688 | 0.3108 | 0.39321 | 0.409737 | 0.1635 | 0.333582 | -0.2886 | -0.58238 | -0.81722 | -0.31469 | 0.000927 | 1.157642 | -0.79564 | 0.646227 | -0.32799 | -0.60955 | -0.48829 | -0.15593 |
| 1189 | -0.77946 | -0.1595 | -0.58673 | -0.5055 | 0.215834 | 0.007188 | -0.30173 | 0.320273 | 0.007367 | -0.83588 | -0.59304 | -0.3148 | 0.106471 | -0.0235 | -1.023 | -0.23152 | -0.73064 | -0.77138 | -0.67864 | -0.55962 |
| 1190 | -0.85326 | -0.02018 | -0.43063 | -0.80604 | -0.35795 | -0.02924 | -0.42921 | 0.173819 | 0.313189 | -0.56868 | -0.28096 | -0.15148 | 0.057275 | -0.17482 | -1.11159 | -0.30217 | -0.98701 | -1.33625 | -1.03837 | -0.75491 |
| 1191 | -0.69587 | -0.06963 | -0.39412 | -0.85635 | -0.04943 | -0.2841 | -0.62001 | 0.173819 | 0.261262 | -0.40492 | 0.174062 | 0.34918 | -2.4E-06 | -0.04428 | -1.22965 | -0.36577 | -1.06533 | -1.01833 | -0.94427 | -0.84244 |
| 1192 | -0.3669 | -0.19356 | -0.20306 | -1.42504 | -0.01998 | -0.19753 | -0.71045 | -0.26183 | -0.12631 | -0.47572 | 0.264148 | -0.03285 | 0.036457 | -0.12894 | -0.45202 | -0.05281 | -1.19185 | -0.85834 | -1.12015 | -0.94028 |
| 1193 | -0.40695 | 0.044734 | 0.15606 | -0.79789 | -0.24257 | -0.14564 | -0.75117 | 0.140696 | -0.43865 | -0.15277 | 0.263748 | -0.10145 | 0.154071 | 0.263792 | 1.407712 | 0.000492 | -0.42629 | -0.96334 | -0.97092 | -0.75823 |
| 1194 | -0.6886 | 0.156769 | 0.429841 | -0.68182 | -0.5276 | -0.1858 | -0.7095 | 0.078911 | -0.94596 | 0.119522 | 0.177883 | -0.42565 | 0.228549 | 0.869884 | 2.242772 | -0.30521 | -0.33701 | -0.8743 | -0.87395 | -0.52851 |
| 1195 | -0.23189 | 0.021037 | 0.508741 | -0.10311 | -0.5372 | -0.30892 | -0.55801 | 0.071397 | -0.54972 | 0.005158 | 0.100798 | -0.47714 | 0.281018 | 1.124448 | 2.601404 | -0.41204 | 0.072169 | -0.25007 | -0.66209 | -0.3708 |
| 1196 | -0.27956 | -0.13859 | 0.435759 | 0.088743 | -0.30361 | -0.04473 | -0.3466 | -0.14346 | 2.088781 | -0.12683 | -0.34036 | -0.72042 | 0.122161 | 0.69794 | 2.675105 | -0.34432 | 0.384359 | 0.268415 | -0.49981 | -0.60932 |
| 1197 | -0.10103 | 0.790246 | -0.24748 | 0.545492 | -0.4492 | -0.23165 | -0.45873 | -0.35182 | 2.966884 | -0.13211 | -0.65157 | -0.91109 | 0.121802 | 1.354293 | 2.223506 | -0.8769 | 0.915069 | -0.30787 | -0.71919 | -0.7065 |
| 1198 | 0.193049 | 0.716704 | -0.633 | 0.651455 | 0.001667 | 0.027039 | -0.75767 | -0.3648 | 2.857465 | -0.38829 | -0.27051 | -1.00056 | -0.27008 | 2.521431 | 0.790603 | -1.02355 | 0.432231 | -0.14631 | -0.92748 | -0.58767 |
| 1199 | 1.154411 | 0.967516 | -0.49065 | 0.956445 | 0.408298 | -0.2192 | -1.02688 | -0.05084 | 2.950663 | -0.23671 | -0.51962 | -0.89935 | -0.59085 | 3.489603 | -0.04325 | -0.93269 | 0.388415 | -0.15632 | -0.63333 | -0.93366 |
| 1200 | 1.197101 | 0.954443 | -0.87614 | 0.918632 | 0.455708 | -0.06595 | -1.09589 | 0.147641 | 2.201599 | -0.16541 | -0.39314 | -1.2872 | -0.35847 | 3.441928 | -0.40368 | -0.37456 | 0.387395 | -0.20905 | -0.57802 | -0.57077 |
| 1201 | 3.075294 | 1.196388 | -0.6585 | 0.816257 | -0.15145 | -0.43968 | -0.66376 | 0.503158 | -0.41063 | -0.24285 | -0.46397 | -1.37499 | -0.23857 | 3.45876 | -0.72831 | -0.07943 | 0.034828 | -0.37923 | -1.23614 | -0.60174 |
| 1202 | 3.358585 | 0.75114 | -0.42513 | 0.787955 | 0.153821 | -0.05633 | -0.31856 | 0.831057 | -0.93882 | 0.049605 | -0.61929 | -0.81938 | 0.002636 | 2.524425 | -0.75016 | -0.14808 | -0.27672 | 0.240996 | -1.04374 | -0.15803 |
| 1203 | 3.745214 | 0.859987 | -0.40524 | 0.166716 | -0.50004 | -0.48691 | 0.016841 | 0.576486 | -0.47026 | 0.232413 | -0.76078 | -0.54192 | 0.606846 | 0.847286 | -0.74547 | -0.05122 | -0.16127 | -0.17558 | -0.8455 | -0.06128 |
| 1204 | 2.939696 | 0.497735 | -0.82187 | -0.26486 | -0.33481 | -0.23383 | 0.510165 | 0.414451 | -0.37419 | -0.16371 | -0.0447 | -0.64111 | 0.748977 | -0.52733 | -0.6312 | 0.206214 | 0.296875 | -0.16672 | -0.97943 | 0.265039 |
| 1205 | 2.230247 | 0.241314 | -0.63637 | -0.51225 | -0.28718 | -0.12649 | 0.19695 | 0.297402 | 0.082324 | -0.27377 | 0.819855 | -0.53573 | 0.512329 | -0.70932 | -0.56566 | -0.4042 | -0.06727 | -0.00163 | -1.08132 | 0.068205 |
| 1206 | 0.101471 | -0.1348 | -1.2901 | -0.5114 | 0.166584 | 0.039563 | -0.40544 | 0.258376 | 0.319237 | -0.07978 | 0.9623 | -0.8912 | 0.156568 | -0.73325 | -0.58014 | -0.61437 | 0.185891 | -0.35324 | -0.59074 | 0.156424 |
| 1207 | -0.22063 | -0.32906 | -1.11401 | -0.70995 | -0.29056 | -0.08094 | -0.34241 | 0.131948 | 1.150756 | -0.00527 | 1.075237 | -1.14739 | -0.09439 | -0.04779 | -1.02641 | -0.43855 | 0.058259 | -0.46422 | -0.51092 | 0.100886 |
| 1208 | -0.5714 | -0.55087 | -1.26495 | -0.49352 | 0.201296 | -0.1343 | -0.16202 | -0.00014 | 1.218081 | -0.0084 | 0.733317 | -1.48455 | -0.56077 | 0.73969 | -0.88385 | -0.78034 | 1.938865 | -0.28954 | -0.27093 | -0.14661 |
| 1209 | -0.5724 | -0.77891 | -0.99117 | -0.4554 | -0.02285 | -0.26524 | -0.28389 | -0.24425 | 1.61372 | 0.26544 | 0.317861 | -1.45986 | -0.58294 | 1.099785 | -1.26385 | -0.74268 | 3.125045 | -0.44061 | -0.43416 | -0.18405 |
| 1210 | -0.35346 | -0.5691 | -0.74056 | -0.615 | -0.27684 | -0.39057 | -0.11147 | -0.3455 | 1.27607 | 0.649272 | -0.90794 | -1.5719 | -0.60068 | 1.464083 | -1.23191 | -0.9513 | 3.438324 | -0.66651 | -0.37788 | 0.179292 |
| 1211 | 0.009731 | -0.54913 | 0.107455 | -0.57303 | -0.36915 | -0.49962 | 0.044127 | -0.52333 | 0.866104 | 0.432685 | -0.92283 | -0.91776 | -0.04421 | 1.527106 | -0.81543 | -0.87775 | 3.269667 | -0.52558 | -0.402 | 0.350313 |
| 1212 | -0.16683 | -0.78575 | 0.67247 | -0.11218 | -0.10831 | -0.67428 | -0.03637 | -0.11372 | 0.012152 | 0.453148 | -0.73955 | -0.78411 | 0.491044 | 1.289855 | -0.3418 | -0.66922 | 3.553728 | -0.40332 | -0.52052 | 0.210784 |
| 1213 | 0.119375 | -0.96248 | 1.573497 | 0.248001 | -0.44022 | -0.54062 | 0.226122 | 0.113258 | -0.08244 | 0.388815 | -0.47993 | -0.68365 | 0.481624 | 0.427401 | -0.6271 | -0.48683 | 1.451549 | -0.20089 | -0.77012 | 0.06239 |
| 1214 | 0.397771 | -0.97954 | 1.400836 | 0.309648 | -0.72504 | -0.21002 | -0.0207 | 0.26722 | -0.79772 | 0.208559 | -0.40679 | -0.61494 | 0.358906 | -0.09249 | -0.35335 | -0.62397 | 0.031681 | -0.14933 | -0.86024 | 0.16877 |
| 1215 | 0.525225 | -1.1117 | 1.322696 | 0.539677 | -0.38167 | -0.52538 | -0.00632 | 0.006612 | -0.41163 | -0.15025 | -0.31002 | -0.56937 | 0.23626 | -0.78848 | -0.35973 | -0.138 | 0.131103 | -0.27145 | -1.01558 | -0.2653 |
| 1216 | 1.000998 | -0.66784 | 0.94299 | 0.330155 | -0.28728 | -0.21799 | -0.21513 | -0.14584 | -0.36231 | -0.08198 | -0.42581 | -0.73058 | 0.170914 | -0.63557 | -0.53774 | -0.13783 | 0.379301 | -0.22952 | -0.8347 | -0.32842 |
| 1217 | 1.156471 | -0.40104 | 0.332655 | -0.04966 | -0.6124 | -0.20583 | -0.64123 | -0.54391 | -0.54471 | -0.11033 | -0.43359 | -0.43615 | -0.0194 | -0.74124 | -0.76085 | -0.42132 | 0.040843 | -0.34596 | -0.70494 | -0.20398 |
| 1218 | 0.727329 | -0.11369 | -0.46227 | -0.47069 | -0.51202 | -0.34183 | -1.03311 | -0.543 | -0.56424 | -0.06758 | -0.60832 | -0.37899 | -0.02561 | -0.45288 | -0.74259 | -0.23834 | -0.0344 | -0.54695 | -0.65066 | 0.005293 |
| 1219 | 0.661487 | 0.130817 | -0.3232 | -0.70011 | -0.53049 | -0.74301 | -1.05096 | -0.76 | -0.25727 | -0.0046 | -1.14613 | -0.36973 | 0.048831 | -0.12085 | -0.17661 | -0.12951 | -0.21553 | -0.67748 | -0.44076 | -0.28512 |
| 1220 | 0.632198 | 0.136424 | -0.33061 | -0.94938 | -0.48486 | -0.44896 | -0.83254 | -0.45483 | -0.40635 | 0.086186 | -1.16694 | -0.12122 | 0.29539 | 0.532826 | -0.27776 | -0.25551 | -0.02081 | -0.476 | 0.139272 | -0.30108 |
| 1221 | 0.187006 | -0.21883 | -0.43878 | -0.93621 | -0.61909 | -0.44896 | -0.86172 | -0.22618 | -0.47273 | -0.06966 | -1.04557 | -0.1777 | 0.29539 | 0.532826 | -0.13276 | -0.39023 | 0.169727 | -0.51517 | 0.139272 | -0.30108 |
| 1222 | -0.11936 | -0.23933 | -0.28901 | -0.87773 | 0.080158 | -0.63594 | -0.78347 | -0.33582 | -0.23023 | -0.13628 | -0.97449 | -0.60517 | -0.0759 | 0.552931 | 0.031136 | -0.34032 | 0.332066 | -0.69016 | 0.031399 | -0.27608 |
| 1223 | -0.12976 | -0.06577 | -0.23765 | -0.84696 | 0.080158 | -0.61068 | -0.72377 | -0.47394 | -0.23023 | -0.13628 | -0.7349 | -0.51962 | 0.02184 | 0.342278 | 0.533715 | -0.49638 | 0.350433 | -0.50627 | 0.106276 | -0.43433 |
| 1224 | -0.1433 | -0.21193 | -0.23765 | 0.084275 | 0.320767 | -0.37583 | -0.29362 | -0.39373 | 0.152044 | -0.13628 | -0.23589 | -0.63038 | 0.417686 | 0.081542 | -0.00863 | -0.5053 | 0.531561 | -0.48096 | -0.06346 | -0.33113 |
| 1225 | -0.3505 | -0.17579 | -0.439 | 0.121623 | 0.011537 | -0.55541 | -0.29362 | -0.54724 | 0.166517 | -0.15585 | -0.14829 | -0.84434 | 0.18781 | -0.19549 | 0.244026 | -0.49755 | 0.136493 | -0.57584 | -0.50523 | -0.43707 |
| 1226 | -0.72112 | 0.009448 | -0.20135 | 0.128088 | 0.000753 | -0.77267 | -0.10167 | -0.37061 | 0.382278 | 0.066305 | -0.33102 | -0.83269 | 0.023661 | -0.19549 | 0.244026 | -0.4879 | -0.10073 | -0.5104 | -0.75318 | -0.38809 |
| 1227 | -0.5633 | -0.2603 | -0.46546 | 0.270427 | -0.40272 | -0.49064 | 0.388563 | -0.23128 | 0.282117 | -0.16296 | -0.40498 | -0.96557 | 0.312245 | -0.29703 | 0.109529 | -0.05244 | -0.28246 | -0.38256 | -0.84224 | -0.63077 |
| 1228 | -0.30326 | -0.70241 | -0.46546 | 0.270427 | -0.6528 | -0.006 | 0.38674 | -0.08115 | 0.503003 | -0.2885 | -0.62885 | -1.14071 | 0.22326 | -0.2533 | -0.3884 | 0.429299 | -0.11755 | -0.78115 | -1.00574 | -0.69131 |
| 1229 | -0.55367 | -0.30208 | -0.6447 | -0.60886 | -0.71497 | 0.553792 | 0.270176 | 0.004096 | 0.120726 | -0.41654 | -0.65028 | -1.15535 | -0.21488 | -0.19044 | -0.7122 | 0.508478 | -0.22688 | -0.87811 | -1.19948 | -0.70865 |
| 1230 | -0.60609 | -0.20566 | -0.7118 | -0.68213 | -0.30285 | 1.132095 | 0.384145 | 0.166792 | 0.120726 | -0.6275 | 0.009964 | -0.94139 | -0.33149 | -0.50753 | -0.98005 | 0.260382 | -0.20636 | -0.96714 | -1.18902 | -0.80573 |
| 1231 | -0.44792 | 0.230714 | -0.70771 | -0.69305 | -0.05966 | 1.349349 | 0.229183 | 0.283233 | 0.128882 | -0.6938 | 0.192698 | -0.60374 | -0.30902 | -0.50753 | -1.22094 | 0.234963 | -0.43943 | -1.21011 | -0.89915 | -0.6928 |
| 1232 | -0.45751 | 0.260479 | -0.6793 | -0.5048 | -0.10196 | 0.991494 | -0.25186 | 0.639285 | 0.001103 | 0.044823 | -0.11063 | -0.31031 | -0.93047 | -0.19725 | -1.06125 | 0.004779 | -0.04791 | -1.27025 | -0.91409 | -0.07581 |
| 1233 | -0.27951 | 0.532766 | -0.61189 | -0.49488 | 0.374393 | 1.640695 | -0.22517 | 0.643911 | -0.04391 | -0.15544 | 0.045486 | -0.19437 | -0.96244 | 0.141284 | -0.78581 | -0.18509 | 0.02895 | -0.77967 | -0.91978 | 0.177015 |
| 1234 | -0.43179 | 0.229832 | -0.11415 | 1.463852 | 0.399147 | 1.084584 | -0.17043 | 0.946906 | -0.11545 | 0.16502 | -0.15852 | 0.033833 | -0.78813 | 0.478532 | -0.55601 | -0.08624 | -0.04763 | -0.71045 | -0.51004 | 0.748325 |
| 1235 | -0.08918 | 0.573907 | -0.15635 | 2.908536 | 0.456531 | 0.700409 | -0.2435 | 1.173238 | -0.02883 | 0.589409 | -0.65495 | 0.049364 | -0.88077 | 0.941787 | -0.18359 | 0.534077 | -0.03378 | -0.59364 | -0.17926 | 1.070917 |
| 1236 | -0.25227 | -0.14644 | -0.21741 | 2.990794 | 0.468597 | 0.643988 | -0.29596 | 0.719741 | -0.3581 | 1.225391 | -0.46178 | -0.23401 | -0.64192 | 0.595804 | 0.119549 | 0.414732 | 0.130292 | -0.67398 | -0.08861 | 0.772736 |
| 1237 | -0.59751 | 0.477055 | 0.018291 | 3.217273 | 0.403777 | 2.031366 | -0.14766 | 0.012976 | -0.25344 | 1.238498 | -0.36144 | -0.52579 | -0.30905 | 1.030124 | -0.07542 | 0.02812 | -0.00122 | -0.03775 | 0.165096 | 0.268994 |
| 1238 | -1.06267 | 0.453373 | -0.07503 | 3.877377 | 0.221589 | 2.161583 | -0.17434 | -0.13168 | -0.41819 | 1.720075 | -0.23496 | -0.41527 | -0.43846 | 1.170652 | 0.273485 | -0.00425 | -0.26904 | 0.076352 | 0.163248 | -0.04859 |
| 1239 | -0.79828 | 0.498196 | 0.137628 | 2.001699 | -0.02339 | 2.1579 | -0.11483 | -0.5307 | -0.46667 | 2.1045 | -0.08094 | -0.41797 | -0.3092 | 0.892962 | 0.312513 | -0.08089 | -0.34176 | 0.843092 | 0.365843 | -0.30109 |
| 1240 | -1.07946 | 0.453487 | 0.252964 | 1.086171 | 0.151229 | 1.882448 | -0.09015 | -0.55379 | -0.00896 | 2.319481 | -0.29647 | -0.13858 | 0.00603 | 0.609193 | 0.631883 | -0.41977 | -0.72166 | 0.641064 | 0.221364 | -0.1449 |
| 1241 | -0.6967 | 0.737459 | 0.529208 | 1.2303 | 0.059127 | 1.690033 | 0.488676 | -0.28047 | 0.289337 | 1.683499 | -0.48964 | 0.224836 | 0.482436 | 0.876217 | 0.593508 | 0.033975 | -0.71685 | 0.733464 | 0.301786 | -0.2305 |
| 1242 | -0.39547 | 0.675783 | 0.755215 | 0.819618 | 0.215228 | 0.434079 | 0.731926 | -0.22487 | 0.734781 | 1.757053 | -0.2127 | 0.630133 | 0.524398 | 0.693671 | 0.727296 | 0.201412 | -0.92089 | 1.009092 | 0.403901 | 0.516373 |
| 1243 | -0.02668 | 0.354982 | 0.78716 | 0.177392 | 0.20824 | -0.78386 | 0.70287 | -0.3657 | 0.988328 | 1.76753 | -0.12247 | 0.253761 | 0.888095 | 0.327465 | 0.198922 | -0.05284 | -0.67509 | 1.159107 | 0.738542 | 1.566965 |
| 1244 | -0.22524 | 0.574742 | 0.258825 | 0.541495 | 0.928759 | -0.66483 | 0.641352 | -0.17038 | 0.820462 | 1.195559 | -0.06185 | 0.230525 | 0.589592 | 0.556545 | -0.0822 | 0.185018 | -0.47074 | 0.859632 | 0.223617 | 1.115343 |
| 1245 | 0.011043 | 0.532025 | 0.22141 | 0.858738 | 0.62427 | -0.35842 | 0.958423 | -0.27646 | 0.465995 | 0.989171 | 0.497393 | -0.03353 | 0.385504 | 0.553407 | -0.22311 | -0.04018 | -0.1265 | 1.104099 | 0.073983 | 0.894656 |
| 1246 | 0.010841 | 0.532025 | 0.082894 | 0.813743 | 0.676725 | -0.06418 | 0.511709 | -0.13661 | 0.82304 | 1.672628 | 0.265358 | -0.04104 | -0.33083 | 0.844778 | 0.002632 | -0.25871 | -0.0717 | 1.498347 | 0.04587 | 1.068173 |
| 1247 | 0.138829 | 0.859335 | 0.080044 | 1.01589 | 0.894346 | 0.073823 | 0.157209 | 0.396771 | 0.599186 | 1.679919 | 0.387784 | 0.213287 | -0.37279 | 0.533483 | 0.018777 | 0.025285 | -0.16263 | 1.187334 | 0.157345 | 0.166128 |
| 1248 | -0.00414 | 0.838674 | 0.256102 | 0.718157 | 0.611037 | 0.219795 | 0.170767 | 0.74064 | 0.055343 | 2.109166 | -0.0244 | 0.512937 | -0.68787 | 0.514515 | 0.264671 | 0.186884 | -0.2279 | 1.38044 | 0.311917 | -0.60092 |
| 1249 | 0.286547 | 0.355574 | 0.303731 | 0.426014 | 0.171549 | 0.383835 | 0.097536 | 1.003244 | 0.561379 | 2.192716 | 0.17753 | 0.699842 | -0.62354 | 0.315294 | 0.862072 | 0.128568 | -0.09296 | 1.363489 | 1.009033 | 0.205741 |
| 1250 | 0.424873 | 0.95929 | 0.885782 | -0.26322 | -0.04838 | 0.173755 | -0.04618 | 0.914074 | 0.527805 | 2.335337 | 0.05294 | 0.647853 | -0.41945 | 0.93999 | 1.019594 | 0.627815 | 0.155152 | 1.451494 | 1.405924 | 0.748211 |
| 1251 | 0.027421 | 1.418586 | 0.805024 | -0.18257 | -0.23938 | 0.221116 | -0.17951 | 0.335068 | 0.872011 | 1.721189 | 0.173215 | 0.704826 | -0.2767 | 1.015835 | 0.644902 | 0.731221 | -0.02312 | 1.376372 | 1.3773 | 0.983524 |
| 1252 | -0.5104 | 0.730606 | 0.902798 | -0.49056 | -0.23802 | 0.253069 | -0.08756 | -0.10107 | 0.973665 | 1.154419 | -0.19247 | 0.691053 | -0.63408 | 1.156783 | 0.277548 | 0.584515 | 0.380536 | 1.100114 | 0.866686 | 1.747179 |
| 1253 | -0.90506 | 1.021748 | 1.773069 | -0.11551 | -0.24354 | 0.204864 | 0.34827 | -0.07303 | 1.404005 | 0.102594 | 0.213607 | 0.313585 | 0.12042 | 1.251249 | 0.198428 | 0.708356 | 0.200734 | 0.647035 | 0.698601 | 1.942416 |
| 1254 | -0.91576 | 1.280353 | 2.021388 | -0.23583 | -0.23813 | 0.479904 | 0.153871 | -0.53526 | 1.014049 | -0.29053 | -0.13219 | 0.014586 | 0.170277 | 1.036678 | -0.27624 | 0.124983 | -0.17213 | 0.145445 | 0.489431 | 1.660219 |
| 1255 | -1.40826 | 0.306466 | 1.695963 | -0.25541 | -0.23866 | 1.002463 | -0.29282 | -0.95468 | 0.857777 | -0.87431 | -0.50916 | -0.10358 | 0.534124 | 0.205703 | -0.82606 | -0.18186 | -0.50963 | -0.04201 | -0.07926 | 1.083136 |
| 1256 | -1.37298 | -0.33572 | 1.538831 | -0.22521 | -0.26139 | 1.129777 | -0.19822 | -0.60168 | 0.201902 | -1.07056 | -0.78909 | 0.238128 | 0.282632 | -0.36213 | -0.80918 | -0.15063 | -0.3855 | -0.02791 | -0.30928 | 1.317532 |
| 1257 | -1.04581 | -0.27458 | 1.244498 | -0.23269 | -0.31689 | 0.450713 | 0.072183 | -0.56889 | 0.001939 | -1.09717 | -0.46079 | 0.161459 | 0.640013 | -0.89938 | -0.52251 | -0.24573 | -0.41028 | -0.34138 | -0.21906 | 0.579826 |
| 1258 | -0.77132 | -0.49263 | 0.192132 | -0.54857 | -0.1485 | 0.620582 | -0.4584 | -0.89698 | -0.38548 | -0.64085 | -0.87848 | 0.344785 | 0.086302 | -1.07801 | -0.53135 | -0.38036 | -0.4314 | -0.48691 | -0.63488 | 0.143384 |
| 1259 | -1.02371 | -0.63806 | -0.08162 | -0.42131 | -0.15254 | -0.1692 | -0.10395 | -0.90164 | -0.36657 | -0.49881 | -0.7798 | 0.427262 | -0.0995 | -0.96706 | -0.45 | -0.12511 | -0.4173 | -0.30481 | -0.86679 | 0.326392 |
| 1260 | -0.98114 | -0.27844 | -0.28212 | -0.32981 | -0.13958 | -0.60571 | 0.111644 | -0.49199 | -0.40461 | -0.07464 | -0.29297 | 1.066721 | -0.49419 | -1.01957 | -0.57585 | -0.13979 | -0.10448 | -0.18571 | -0.53842 | 0.598123 |
| 1261 | -1.13691 | -0.10385 | -0.08191 | -0.55242 | 0.118605 | -0.64617 | 0.21347 | -0.26515 | -0.50779 | -0.18773 | 0.223658 | 0.774574 | -0.6814 | -0.95077 | -0.5191 | -0.26735 | -0.1338 | -0.29334 | -0.99233 | -0.11355 |
| 1262 | -1.30932 | 0.129111 | 0.127727 | -0.4212 | -0.47939 | 0.074184 | -0.16926 | -0.09826 | -0.63988 | -0.09464 | 0.168629 | 0.449208 | -1.02708 | -0.70506 | -0.26592 | 0.178567 | -0.34555 | -0.36899 | -0.81025 | 0.259506 |
| 1263 | -1.76489 | 0.562911 | -0.27238 | -0.35684 | -0.38158 | -0.03433 | -0.2192 | 0.312841 | -0.72115 | -0.16335 | 0.510449 | 0.808444 | -0.84882 | -0.45223 | -0.18023 | 0.220611 | -0.36684 | -0.51987 | -1.07007 | 0.599416 |
| 1264 | -1.53315 | 0.964559 | -0.19951 | -0.43635 | -0.51696 | 0.406477 | -0.4291 | 0.695206 | -0.84147 | -0.00789 | 1.390471 | 1.048546 | -0.70859 | -0.47506 | -0.10102 | 0.36004 | -0.31517 | -0.34254 | -1.02564 | 0.307163 |
| 1265 | -1.1241 | 0.82965 | 0.480651 | -0.38415 | -0.49822 | 0.2013 | -0.43695 | 0.818524 | -0.93476 | -0.38073 | 1.116005 | 0.703682 | -0.18474 | -0.21009 | 0.044035 | 0.467745 | -0.57185 | -0.44207 | -1.30273 | 0.085056 |
| 1266 | -0.71431 | 0.660757 | 0.56071 | -0.30903 | -0.58807 | -0.23806 | -0.4311 | 0.730184 | -0.87998 | 0.167219 | 0.835951 | 0.252712 | 0.241994 | -0.0685 | -0.0491 | 0.627555 | -0.58719 | -0.59941 | -0.75862 | 0.475791 |
| 1267 | -0.65285 | 0.399543 | 0.211024 | -0.03657 | -0.49997 | -0.20017 | -0.60248 | 0.698689 | -0.87341 | 0.391459 | 0.956887 | 0.162021 | 0.802726 | -0.17432 | -0.28019 | 0.303834 | -0.58925 | -0.37154 | -0.72015 | 0.583348 |
| 1268 | -0.17973 | 0.363899 | 0.737274 | -0.01312 | -0.46441 | -0.4856 | -0.56742 | 0.129178 | -0.59116 | 0.398561 | 0.635226 | 0.238089 | 0.52303 | -0.52038 | -0.28526 | 0.103618 | -0.3634 | -0.12059 | -0.2208 | 0.589321 |
| 1269 | 0 | 0 | 0 | 0 | 0 | 0 | 0 | 0 | 0 | 0 | 0 | 0 | 0 | 0 | 0 | 0 | 0 | 0 | 0 | 0 |
| 1270 | 0 | 0 | 0 | 0 | 0 | 0 | 0 | 0 | 0 | 0 | 0 | 0 | 0 | 0 | 0 | 0 | 0 | 0 | 0 | 0 |
| 1271 | 0 | 0 | 0 | 0 | 0 | 0 | 0 | 0 | 0 | 0 | 0 | 0 | 0 | 0 | 0 | 0 | 0 | 0 | 0 | 0 |
| 1272 | 0 | 0 | 0 | 0 | 0 | 0 | 0 | 0 | 0 | 0 | 0 | 0 | 0 | 0 | 0 | 0 | 0 | 0 | 0 | 0 |
| 1273 | 0 | 0 | 0 | 0 | 0 | 0 | 0 | 0 | 0 | 0 | 0 | 0 | 0 | 0 | 0 | 0 | 0 | 0 | 0 | 0 |
| 1274 | 0 | 0 | 0 | 0 | 0 | 0 | 0 | 0 | 0 | 0 | 0 | 0 | 0 | 0 | 0 | 0 | 0 | 0 | 0 | 0 |

| | | | | | | | | | | | | | | |
|---|---|---|---|---|---|---|---|---|---|---|---|---|---|---|
| 1165 | 0.029699 | 0.497313 | 0.081592 | 0.445686 | 0.473789 | -0.02593 | -0.38959 | 1.969955 | -0.52831 | 3.781863 | 0.269646 | -0.34428 | -0.73517 | 1.548946 |
| 1166 | -0.3938 | 0.672461 | -0.27304 | 0.238148 | 1.515803 | 0.279738 | 0.018055 | 3.078989 | -0.74845 | 4.452965 | 0.374169 | -0.20685 | -0.38687 | 1.498919 |
| 1167 | -0.50263 | 0.751201 | 0.579614 | 0.077012 | 4.248561 | 0.80811 | 0.112254 | 5.59411 | 2.898725 | 4.186971 | -0.02114 | 1.736794 | -0.6394 | 2.2164 |
| 1168 | -1.00749 | 0.596404 | 1.9757 | 0.481447 | 6.163587 | 0.239644 | 0.569225 | 7.166694 | 6.274623 | 3.592092 | 0.157931 | 3.148476 | -0.4301 | 3.760732 |
| 1169 | -0.85143 | -0.05404 | 1.703566 | 0.374122 | 6.436996 | -0.01031 | 0.401308 | 6.255391 | 6.70573 | 1.764499 | 0.256206 | 3.341861 | -0.37528 | 3.222904 |
| 1170 | -0.86033 | -0.07904 | 1.48238 | 0.271235 | 5.584212 | 0.013107 | 0.550465 | 5.157388 | 6.973175 | 0.048777 | 0.264606 | 3.124021 | -0.59994 | 2.87096 |
| 1171 | -0.54293 | -0.54463 | 1.832333 | 0.30141 | 4.422425 | 0.127431 | 0.525114 | 4.811262 | 7.391385 | -0.53557 | 0.358669 | 3.262203 | -0.8174 | 2.816263 |
| 1172 | -0.53284 | -0.23627 | 0.753462 | 0.314491 | 1.461318 | 0.36356 | 0.201071 | 2.384339 | 4.024896 | -0.43255 | 0.786754 | 1.013523 | 0.163793 | 1.81759 |
| 1173 | -0.29518 | -0.02318 | -0.31173 | -0.44699 | -0.24113 | 0.790546 | -0.22945 | 0.531922 | 0.52239 | -0.63436 | 0.349121 | -0.36189 | -0.19527 | 1.146534 |
| 1174 | -0.65437 | -0.06938 | -0.64598 | -0.49586 | -0.58424 | 0.603389 | -0.29105 | 0.37127 | -0.00781 | -0.46309 | 0.293472 | -0.41269 | -0.11934 | 0.921007 |
| 1175 | -0.01623 | -0.4569 | -0.87984 | -0.82015 | -0.78692 | 0.848468 | -0.68907 | 0.32657 | -0.46803 | -0.28788 | 0.325877 | -0.26357 | -0.08708 | 1.029174 |
| 1176 | 0.08791 | -0.08441 | -0.47566 | -0.80932 | -0.56819 | 0.660189 | -0.67249 | -0.0753 | -0.48478 | -0.31587 | -0.05691 | -0.32109 | -0.0238 | 1.079172 |
| 1177 | 0.120384 | -0.35544 | -0.24457 | -0.47529 | -0.672 | 0.338293 | -0.53928 | -0.32609 | -0.58267 | -0.13808 | -0.54977 | -0.47283 | -0.66337 | 1.109356 |
| 1178 | -0.02127 | 0.20172 | -0.22503 | -0.44489 | -0.66308 | 0.314336 | -0.77598 | -0.19886 | -0.64803 | -0.3222 | -0.68799 | -0.76624 | -0.33819 | 1.030535 |
| 1179 | 0.501481 | 0.480968 | -0.17826 | -0.31469 | -0.66305 | 0.633726 | -0.78644 | -0.10879 | -0.7411 | -0.25877 | -0.84172 | -0.83898 | -0.61662 | 1.132514 |
| 1180 | 0.158793 | 0.511865 | 0.229693 | 0.112859 | -0.33969 | 0.48035 | -0.46292 | -0.34777 | -0.63865 | -0.01884 | -0.80498 | -0.95103 | -0.41579 | 1.232239 |
| 1181 | 0.010482 | 0.247393 | 0.332078 | 0.04884 | -0.26863 | 0.29474 | -0.47847 | -0.4246 | -0.86015 | 0.025415 | -0.60598 | -0.64033 | -0.37258 | 1.344327 |
| 1182 | -0.04776 | 0.400217 | 0.115098 | -0.4291 | -0.09534 | 0.279929 | -0.88753 | 0.076617 | -1.19339 | -0.24609 | -0.19507 | -0.11034 | -0.67239 | 1.333076 |
| 1183 | -0.02112 | -0.29953 | 0.293238 | -0.45188 | -0.03809 | -0.30245 | -0.96656 | 0.338077 | -0.94519 | -0.08006 | 0.932604 | 0.463837 | -0.32513 | 1.6175 |
| 1184 | -0.33158 | -0.22016 | 0.604667 | -0.58952 | 0.089623 | -0.42524 | -0.06354 | -0.00743 | -1.38284 | -0.47431 | 2.00154 | 0.408529 | 0.450045 | 1.659119 |
| 1185 | -0.50944 | -0.1012 | 0.705781 | -0.76689 | -0.45364 | -0.48526 | 0.128841 | -0.26018 | -1.09174 | -0.83189 | 2.015965 | 0.526582 | 0.517431 | 1.605604 |
| 1186 | -0.51575 | -0.16056 | 0.507598 | -0.33112 | -0.67473 | -0.67528 | -0.07286 | -0.42033 | -0.90307 | -1.03209 | 2.422356 | 0.012684 | 0.541844 | 1.617194 |
| 1187 | -0.68864 | -0.44369 | 0.706026 | 0.012766 | -0.67809 | -0.90167 | -0.06691 | -0.94577 | -0.97173 | -1.0788 | 2.247932 | -0.07626 | 0.830392 | 1.464041 |
| 1188 | -0.67412 | -0.29642 | 0.592599 | -0.11332 | -0.95967 | -0.46533 | 0.494921 | -0.67965 | -0.90981 | -0.9144 | 1.113871 | -0.28516 | 0.139356 | 1.15869 |
| 1189 | -0.8004 | -0.6317 | 0.743397 | -0.1099 | -0.99979 | -0.48738 | -0.16647 | -0.91625 | -0.33244 | -0.7037 | 0.167984 | -0.01549 | -0.45508 | 0.843249 |
| 1190 | -0.65841 | -0.84066 | 0.296361 | -0.23665 | -1.08758 | -0.74602 | -0.09599 | -0.84428 | -0.42122 | -0.8807 | 0.015303 | -0.09963 | -0.71456 | 0.79023 |
| 1191 | -0.60793 | -0.57054 | 0.030979 | -0.55012 | -1.04126 | -0.46251 | 0.001986 | -0.66083 | -0.1277 | -0.60619 | -0.24407 | -0.3228 | -0.64517 | 0.749285 |
| 1192 | -0.34655 | -0.55407 | -0.28669 | -0.78167 | -1.29537 | 0.006877 | 0.617025 | -0.47177 | -0.03944 | -0.67094 | -0.32617 | -0.45252 | -0.26996 | 0.866209 |
| 1193 | -0.33729 | -0.59282 | -0.46469 | -0.47379 | -1.23564 | -0.01703 | 0.490124 | -0.83806 | -0.17008 | -0.57248 | 0.03349 | -0.49095 | 0.050918 | 0.92341 |
| 1194 | -0.19041 | -0.77224 | -0.78109 | -0.31837 | -1.07344 | -0.19578 | 0.136607 | -0.70045 | -0.07223 | -0.91402 | -0.05117 | -1.06766 | -0.28246 | 0.936376 |
| 1195 | 0.194181 | -0.79538 | -0.78109 | -0.31623 | -0.51274 | 0.176038 | 0.109345 | 0.640995 | -0.24403 | -0.6816 | -0.12926 | -1.1789 | 0.081033 | 0.928204 |
| 1196 | 0.21907 | -0.78901 | -0.95859 | -0.81675 | -0.25772 | 0.283063 | -0.13593 | 1.392905 | -0.61342 | -0.85984 | -0.32755 | -0.48716 | 0.082996 | 1.169098 |
| 1197 | 0.267906 | -0.48751 | -0.85067 | -0.92217 | -0.42129 | -0.01614 | -0.44575 | 1.568717 | -0.18984 | -0.64667 | -0.16113 | -0.15666 | -0.02717 | 1.406721 |
| 1198 | 0.646513 | -0.66259 | -1.0222 | -0.87257 | -0.35393 | -0.00683 | -0.39063 | 1.085894 | -0.01514 | -0.72747 | -0.35281 | 0.015489 | -0.01741 | 1.162942 |
| 1199 | 0.921546 | -0.79871 | -0.90161 | -0.85735 | -0.59315 | 0.089072 | -0.0995 | 1.324162 | -0.11143 | -0.3736 | -0.3228 | 0.156869 | 0.278403 | 1.248173 |
| 1200 | 0.638972 | -0.53338 | -0.83569 | -0.29378 | -0.77735 | -0.07046 | -0.51668 | 0.140199 | 0.203281 | -0.47346 | 0.00442 | 0.28349 | 0.316741 | 1.107756 |
| 1201 | 0.657364 | -0.54477 | -0.70266 | 0.114696 | -0.81828 | -0.30464 | -0.45458 | -0.75401 | 0.693217 | -0.16346 | 0.250517 | -0.3064 | 0.288939 | 1.070264 |
| 1202 | 0.500051 | -0.63167 | -0.37437 | 0.160766 | -0.31574 | -0.24137 | -0.62756 | -0.73126 | 0.715036 | 0.313789 | 0.506644 | -0.27965 | 0.156648 | 1.043588 |
| 1203 | -0.02342 | -0.60129 | -0.32441 | -0.10552 | -0.24061 | -0.57107 | -0.89118 | -0.24375 | 0.569722 | -0.0065 | 1.064791 | -0.9955 | -0.06892 | 1.013527 |
| 1204 | -0.44048 | -0.674 | -0.32652 | -0.20105 | -0.19888 | -0.8665 | -1.00859 | -0.56434 | 0.543798 | -0.21558 | 1.37407 | -1.07139 | 0.052866 | 0.979572 |
| 1205 | -0.51201 | -0.48764 | -0.66335 | -0.47055 | -0.21118 | -0.89355 | -1.04029 | -0.47797 | -0.06297 | -0.16483 | 1.007338 | -1.27405 | -0.18629 | 1.142334 |
| 1206 | -0.3493 | -0.408 | -0.73578 | -0.76005 | -0.33789 | -0.92769 | -0.8536 | -0.68265 | -0.51412 | -0.15263 | 1.083664 | -0.9424 | -0.57117 | 0.921113 |
| 1207 | -0.32842 | -0.42487 | -0.63062 | -0.32558 | -0.76207 | -0.64162 | -0.78129 | -0.53932 | -0.79803 | -0.65454 | 0.6076 | -1.72777 | -0.90666 | 1.039684 |
| 1208 | -0.26126 | -0.38163 | -0.26738 | 0.01591 | -1.01722 | -0.14644 | -0.88243 | -0.87347 | -1.00938 | -0.78237 | -0.21878 | -1.46875 | -1.1254 | 1.104352 |
| 1209 | 0.123686 | -0.13424 | -0.25459 | -0.11912 | -0.9338 | 0.283917 | -0.26012 | -0.78319 | -1.22401 | -0.27838 | -0.60829 | -1.2239 | -0.94482 | 1.044518 |
| 1210 | 0.301409 | -0.19772 | -0.08373 | -0.33634 | -0.81484 | 0.272497 | -0.11099 | -0.76268 | -0.7625 | -0.4104 | -0.83078 | -1.14779 | -0.8825 | 1.071159 |
| 1211 | 0.178193 | -0.25081 | 0.143073 | 0.099441 | -0.75356 | 0.288717 | -0.04611 | 0.927778 | -0.66928 | -0.69915 | -1.03321 | -1.34179 | -0.70004 | 1.066116 |
| 1212 | 0.179035 | -0.28534 | -0.15607 | 0.216877 | -0.47864 | -0.33459 | 0.170893 | 2.711849 | -0.65652 | -0.71602 | -0.88939 | -1.26654 | -0.05752 | 1.350259 |
| 1213 | 0.457208 | 0.119567 | -0.17066 | 0.287404 | -0.47169 | -0.89246 | 0.2351 | 3.698345 | -0.60052 | -0.08462 | -0.60466 | -1.33376 | 0.388886 | 1.467085 |
| 1214 | 0.2702 | 0.256748 | -0.13947 | 0.269487 | -0.48823 | -1.20995 | -0.40599 | 3.814074 | -0.56355 | -0.33307 | -0.46978 | -1.51138 | -0.09699 | 1.42077 |
| 1215 | -0.01157 | -0.04566 | -0.27734 | 1.046454 | -0.86357 | -1.25801 | -0.43469 | 3.529308 | -0.9637 | -0.25411 | -0.457 | -1.5236 | -0.14563 | 1.230407 |
| 1216 | 0.063952 | -0.13366 | -0.34017 | 0.481925 | -1.02657 | -1.1498 | -0.00761 | 2.282945 | -0.97966 | -0.10932 | -0.69645 | -1.15864 | -0.12264 | 1.161238 |
| 1217 | -0.002 | 0.114995 | -0.58224 | 0.247632 | -0.95862 | -0.6006 | -0.1112 | 0.305465 | -0.96849 | -0.03481 | -0.92986 | -1.05579 | -0.77148 | 1.021137 |
| 1218 | -0.21811 | -0.33704 | -0.5963 | 0.01235 | -0.65142 | -0.24927 | 0.079576 | -0.38526 | -0.83305 | -0.31647 | -1.09848 | -0.92488 | -0.9088 | 0.782041 |
| 1219 | -0.21811 | -0.35542 | -0.30161 | -0.11955 | -0.79911 | -0.2261 | 0.367359 | -0.32721 | -0.65222 | -0.31647 | -1.635 | -0.60064 | -0.99225 | 0.84762 |
| 1220 | -0.16391 | -0.37786 | 0.047933 | -0.94559 | -0.56598 | -0.07371 | 0.139035 | 0.004127 | -0.55966 | -0.12078 | -1.26731 | -0.31273 | -0.85758 | 0.88117 |
| 1221 | 0.075403 | -0.18673 | 0.172217 | -0.50441 | -0.31437 | -0.07763 | -0.25271 | 0.01579 | -0.69973 | -0.37163 | -0.66557 | -0.43329 | -0.72169 | 0.771466 |
| 1222 | 0.039504 | -0.53651 | 0.489733 | -0.53113 | -0.31792 | -0.31068 | -0.28336 | -0.02451 | -0.87574 | -0.5092 | -0.69411 | -0.05356 | -0.41236 | 0.788246 |
| 1223 | 0.252366 | -0.26494 | 0.211728 | -0.53113 | -0.24512 | -0.25501 | -0.34525 | 0.192497 | -0.7829 | -0.42849 | -0.40662 | 0.205874 | -0.30452 | 0.687591 |
| 1224 | 0.343936 | -0.40077 | -0.10243 | -0.26169 | 0.035255 | -0.01277 | -0.36099 | 0.192497 | -0.80718 | -0.6355 | -0.05883 | 0.022599 | 0 | 0.640865 |
| 1225 | 0.169231 | -0.17763 | -0.08169 | 0 | 0.035255 | 0.002091 | -0.14841 | 0.192497 | -0.53676 | -0.88453 | -0.09922 | -0.14192 | 0 | 0.711714 |
| 1226 | -0.26496 | -0.50973 | -0.32965 | 0.872528 | -0.03676 | -0.2088 | -0.40841 | 0.244438 | -0.39204 | -0.63367 | -0.54165 | 0.022974 | -0.07738 | 0.729665 |
| 1227 | -0.26268 | -0.60262 | -0.62679 | 0.749222 | -0.20228 | -0.3384 | -0.40841 | 0.069473 | -0.11504 | -0.5405 | -0.45269 | -0.2599 | 0.141139 | 0.831559 |
| 1228 | 0.005057 | -0.88567 | -0.42567 | 0.749222 | -0.17477 | -0.46136 | -0.34348 | -0.28443 | 0.09787 | -0.3499 | -0.41174 | -0.51275 | -0.13521 | 0.93826 |
| 1229 | -0.04027 | -1.00927 | -0.39739 | 0.562878 | -0.41745 | -0.78225 | -0.67986 | -0.50303 | -0.03841 | -0.12495 | -0.20211 | -0.23137 | 0.058499 | 0.924289 |
| 1230 | -0.10456 | -1.07836 | -0.52432 | 0.455328 | -0.55704 | -0.90528 | -0.77761 | -0.26524 | -0.03841 | 0.134699 | -0.48887 | -0.31823 | -0.09162 | 0.823359 |
| 1231 | 0.3097 | -0.637 | -0.36482 | -0.48954 | -0.70384 | -0.68163 | -0.79229 | -0.31718 | 0.344464 | -0.09456 | -0.38969 | -0.46459 | -0.07343 | 0.812118 |
| 1232 | -0.03614 | -0.60782 | 0.508736 | -0.35887 | -0.42563 | -0.70884 | -0.3428 | -0.3461 | 0.592739 | 0.294909 | -0.2284 | 0.228045 | -0.4047 | 1.153685 |
| 1233 | 0.425467 | -0.57022 | 0.646401 | 0.064465 | -0.64304 | -0.54788 | -0.09972 | -0.2734 | 0.301701 | 0.26029 | -0.80715 | 0.639593 | -0.08243 | 1.253411 |
| 1234 | 0.590471 | -0.37125 | 0.545164 | 0.591397 | -0.64843 | 0.158747 | 0.13324 | 0.215965 | 0.207494 | 0.529424 | -0.84464 | 0.987836 | -0.38596 | 1.434175 |
| 1235 | 0.873897 | 0.036062 | 0.280527 | 1.239034 | 0.368976 | 0.896368 | 0.6856 | -0.21364 | 0.257995 | 0.778162 | -0.69772 | 1.013488 | -0.02677 | 1.459683 |
| 1236 | 0.49622 | -0.28077 | 0.441654 | 1.237629 | 1.310689 | 1.356419 | 1.24266 | 0.089947 | -0.15213 | 1.358053 | -0.63689 | 1.552531 | 0.000172 | 1.408395 |
| 1237 | 1.231556 | -0.02371 | -0.25 | 1.427887 | 1.376734 | 1.488293 | 0.470959 | 0.575108 | -0.43158 | 1.346053 | -0.72391 | 1.123387 | 0.280373 | 1.392319 |
| 1238 | 0.489653 | -0.00732 | -0.27406 | 1.186312 | 1.710743 | 1.480191 | 0.238039 | 0.788472 | -0.36313 | 0.943579 | -0.42755 | 1.28652 | 0.461432 | 1.451675 |
| 1239 | 0.648044 | 0.296495 | -0.47964 | 1.128167 | 1.69204 | 1.202455 | 0.586549 | 0.387806 | -0.26643 | 0.901363 | -0.62154 | 0.801196 | 1.085347 | 1.43217 |
| 1240 | 0.622964 | -0.01812 | 0.03772 | 0.436601 | 0.655548 | 0.473927 | 0.358437 | 0.810461 | -0.07321 | 0.58574 | -0.14802 | 0.835648 | 0.984797 | 1.371526 |
| 1241 | 0.535696 | 0.073533 | -0.18004 | 0.532279 | -0.28391 | -0.21485 | 0.025525 | 0.53859 | 0.052527 | 0.232553 | -0.3816 | 0.240395 | 1.056663 | 1.274267 |
| 1242 | 0.501329 | -0.25471 | -0.15972 | 0.334653 | -0.75818 | -0.2703 | 0.044993 | 0.257312 | -0.05939 | -0.10966 | -0.56579 | 0.298147 | 0.561523 | 1.329679 |
| 1243 | 0.370767 | -0.06202 | -0.04015 | 0.240796 | -0.94745 | -0.55229 | 0.496694 | 0.199685 | -0.04972 | 0.209095 | -0.54287 | -0.08398 | 0.186338 | 1.341201 |
| 1244 | -0.01874 | 0.055335 | 0.4097 | -0.06489 | -0.16362 | -0.45427 | 0.491031 | 0.493192 | -0.1925 | 0.442 | 0.05463 | 0.000919 | -0.65725 | 1.421389 |
| 1245 | -0.20022 | -0.18777 | 0.349056 | 0.197916 | -0.29052 | -0.46552 | 0.685715 | 0.447175 | -0.01817 | 0.880594 | 0.630066 | 0.001294 | -0.75298 | 1.515644 |
| 1246 | 0.387616 | -0.41424 | 0.993488 | -0.14598 | -0.22572 | -0.30713 | 1.251911 | 0.919563 | 0.254187 | 1.303454 | 1.09416 | 0.287454 | -0.27712 | 1.390015 |
| 1247 | -0.0323 | -0.23304 | 1.102971 | -0.49126 | 0.322425 | -0.05024 | 1.510312 | 0.618262 | 0.294963 | 1.131431 | 1.596979 | 0.325355 | 0.104875 | 1.481628 |
| 1248 | 0.093711 | -0.06994 | 1.055081 | -0.09792 | 0.355148 | 0.20186 | 1.085182 | 0.269465 | 0.162698 | 1.059163 | 2.334751 | 0.46673 | 0.529697 | 1.327539 |
| 1249 | 0.286826 | -0.54879 | 0.573626 | -0.01314 | 0.198717 | -0.18295 | 1.122735 | 0.400803 | 0.357167 | 0.594651 | 2.091762 | 0.376599 | 1.040928 | 1.208669 |
| 1250 | 0.497487 | -0.49026 | 0.809134 | -0.07481 | 0.524113 | 0.079337 | 1.128433 | 0.362415 | -0.18286 | 0.176142 | 0.958871 | 0.697601 | 0.893157 | 1.367904 |
| 1251 | -0.20734 | -0.4048 | 0.146136 | 0.242949 | 1.218946 | 0.014542 | 0.572531 | -0.50825 | -0.58514 | -0.22932 | 0.799121 | 0.460899 | 0.266132 | 1.252887 |
| 1252 | -0.02572 | -0.74465 | -0.2355 | 0.605738 | 1.34202 | -0.45459 | 0.979026 | -0.30329 | -0.27564 | 0.100349 | 0.739607 | 0.443935 | 0.077042 | 1.285098 |
| 1253 | -0.18148 | -0.17758 | -0.01807 | 0.103142 | 1.616395 | 0.046371 | 1.294369 | -0.35479 | 0.240474 | -0.14361 | 0.071083 | 0.250407 | -0.0823 | 1.359499 |
| 1254 | -0.41436 | 0.26247 | 0.493853 | -0.14854 | 1.099791 | 0.412767 | 1.320369 | -0.79675 | 0.687769 | -0.47524 | -0.24979 | 0.055425 | -0.54006 | 1.490103 |
| 1255 | -0.94298 | 0.825716 | 0.488991 | -0.27649 | 1.041101 | 0.216998 | 0.615828 | -1.20403 | 0.662354 | -0.70892 | -0.04771 | -0.3122 | -0.46912 | 1.631533 |
| 1256 | -0.89093 | 0.736372 | 0.630659 | -0.41783 | 0.392246 | 0.209198 | 0.565023 | -0.83746 | 0.64953 | -0.98742 | -0.13472 | -0.56512 | -0.73216 | 1.991783 |
| 1257 | -1.14058 | 1.475197 | 1.105168 | -0.37672 | -0.22505 | 0.483693 | 0.105929 | -0.55757 | 0.144368 | -1.34464 | -0.4466 | -0.5058 | -0.74629 | 2.420819 |
| 1258 | -0.92201 | 0.621094 | 0.486457 | -0.35536 | -0.49942 | 0.087992 | -0.41249 | -0.41316 | -0.5212 | -1.27 | -0.29733 | -0.68192 | -0.86356 | 3.045575 |
| 1259 | -1.17112 | 0.328906 | 0.454318 | -0.30313 | -0.60563 | -0.48958 | -0.7177 | -0.15737 | -1.0063 | -1.14328 | -0.37367 | -0.48408 | -0.75556 | 4.356056 |
| 1260 | -0.83455 | -0.21531 | 0.020494 | -0.13952 | -0.75219 | -0.70699 | -0.75768 | 0.020067 | -0.9233 | -1.32183 | -0.41498 | -0.61999 | -0.83504 | 4.019228 |
| 1261 | -0.47816 | 0.062575 | 0.05185 | -0.09414 | -0.96727 | -0.80979 | -0.57626 | -0.01675 | -0.8448 | -0.73277 | -0.38142 | -0.2279 | -0.72952 | 3.057691 |
| 1262 | -0.35547 | 0.022682 | -0.01973 | -0.45683 | -0.74568 | -0.86408 | -0.62938 | -0.29046 | -0.24396 | -0.48993 | -0.4654 | -0.57339 | -0.93368 | 2.35365 |
| 1263 | -0.80467 | 0.007585 | 0.621314 | -0.5469 | -0.95347 | -1.05391 | -0.78645 | -0.09286 | -0.21201 | -0.2216 | -0.97419 | -0.1904 | -1.30492 | 1.926857 |
| 1264 | -0.44698 | -0.07125 | 0.455225 | -0.84634 | -0.80166 | -0.70616 | -0.91603 | -0.65308 | -0.3251 | -0.20184 | -0.60634 | 0.100618 | -1.61018 | 1.228545 |
| 1265 | -0.54963 | -0.05186 | 0.261805 | -0.69402 | -0.95387 | -0.37923 | -0.38109 | -0.3675 | -0.04508 | 0.085276 | -0.84667 | 0.30954 | -1.20271 | 1.176177 |
| 1266 | -0.87098 | 0.281129 | 0.15268 | -0.84216 | -0.746 | 0.086771 | -0.65104 | -0.8058 | 0.050986 | -0.48499 | -1.19328 | 0.158581 | -1.2597 | 1.182542 |
| 1267 | -0.64223 | 0.070063 | -0.10114 | -0.52922 | -0.92255 | 0.047462 | -0.55369 | -0.80721 | -0.26636 | -0.55603 | -0.63031 | 0.069912 | -0.86834 | 1.295777 |
| 1268 | -0.07597 | 0.479835 | -0.54661 | -0.4664 | -0.85293 | 0.121323 | -0.30411 | -0.48117 | -0.4356 | -0.78891 | 0.225977 | -0.28695 | -0.68341 | 1.287929 |
| 1269 | 0 | 0 | 0 | 0 | 0 | 0 | 0 | 0 | 0 | 0 | 0 | 0 | 0 | 0 |
| 1270 | 0 | 0 | 0 | 0 | 0 | 0 | 0 | 0 | 0 | 0 | 0 | 0 | 0 | 0 |
| 1271 | 0 | 0 | 0 | 0 | 0 | 0 | 0 | 0 | 0 | 0 | 0 | 0 | 0 | 0 |
| 1272 | 0 | 0 | 0 | 0 | 0 | 0 | 0 | 0 | 0 | 0 | 0 | 0 | 0 | 0 |
| 1273 | 0 | 0 | 0 | 0 | 0 | 0 | 0 | 0 | 0 | 0 | 0 | 0 | 0 | 0 |
| 1274 | 0 | 0 | 0 | 0 | 0 | 0 | 0 | 0 | 0 | 0 | 0 | 0 | 0 | 0 |
